# Heart-brain connections: phenotypic and genetic insights from 40,000 cardiac and brain magnetic resonance images

**DOI:** 10.1101/2021.11.01.21265779

**Authors:** Bingxin Zhao, Tengfei Li, Zirui Fan, Yue Yang, Xifeng Wang, Tianyou Luo, Jiarui Tang, Di Xiong, Zhenyi Wu, Jie Chen, Yue Shan, Chalmer Tomlinson, Ziliang Zhu, Yun Li, Jason L. Stein, Hongtu Zhu

## Abstract

Cardiovascular health interacts with cognitive and psychological health in complex ways. Yet, little is known about the phenotypic and genetic links of heart-brain systems. Using cardiac and brain magnetic resonance imaging (CMR and brain MRI) data from over 40,000 UK Biobank subjects, we developed detailed analyses of the structural and functional connections between the heart and the brain. CMR measures of the cardiovascular system were strongly correlated with brain basic morphometry, structural connectivity, and functional connectivity after controlling for body size and body mass index. The effects of cardiovascular risk factors on the brain were partially mediated by cardiac structures and functions. Using 82 CMR traits, genome-wide association study identified 80 CMR-associated genomic loci (*P* < 6.09 × 10^-10^), which were colocalized with a wide spectrum of heart and brain diseases. Genetic correlations were observed between CMR traits and brain-related complex traits and disorders, including schizophrenia, bipolar disorder, anorexia nervosa, stroke, cognitive function, and neuroticism. Our results reveal a strong heart-brain connection and the shared genetic influences at play, advancing a multi-organ perspective on human health and clinical outcomes.

---

A growing amount of evidence suggests close interplays between heart heath and brain health (Fig. 1A). Cardiovascular diseases may provide a pathophysiological background for several brain diseases, such as stroke^1^, dementia^2^, and cognitive impairment^3–5^. For example, atrial fibrillation has been linked to increased incidence of dementia^6^ and silent cerebral damage^7^, even in stroke-free cohorts^8^. It is consistently observed that heart failure is associated with cognitive impairment and eventually dementia^9^, likely due to the reduced cerebral perfusion caused by the failing heart^10^. Mental disorders and negative psychological factors, on the other hand, contribute substantially to the initiation and progression of cardiovascular diseases^11,12^. Patients with mental illnesses (such as schizophrenia, bipolar disorder, or depression) show an increased incidence of cardiovascular diseases^13–16^. Acute mental stress may cause a higher incidence of atherosclerosis due to stress-induced vascular inflammation and leucocytes migration^17^. In part due to the lack of data, almost all prior studies on heart-brain interactions focused on one (or a few) specific diseases or used small samples. The overall picture of structural and functional links between the heart and the brain remains unclear.

**Fig. 1.**
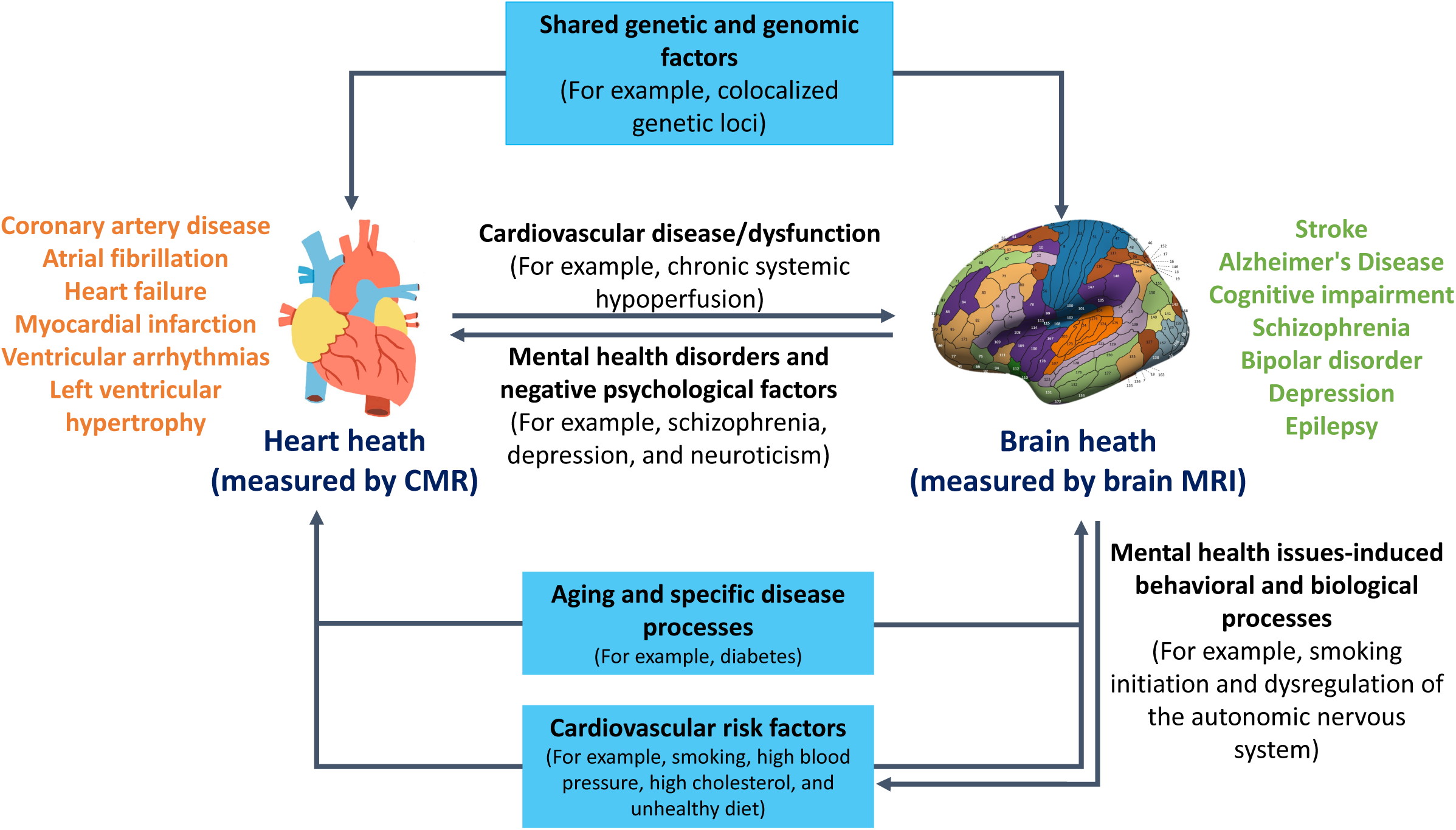
Illustration of potential heart-brain connections. We illustrated examples of the potential heart-brain connections. For example, cardiovascular disease/dysfunction may result in cognitive impairment and Alzheimer’s Disease, while mental health disorders and negative psychological health factors may contribute to higher risk of cardiovascular disease. Multi-system aging and disease processes as well as cardiovascular risk factors may influence both brain and heart in a negative way. There may also exist the shared genetic influences between heart and brain heath.

In heart and brain diseases, magnetic resonance imaging (MRI) traits are well-established endophenotypes. Cardiovascular magnetic resonance imaging (CMR) is used to assess cardiac structure and function, yielding insights into the risk and pathological status of cardiovascular diseases^18–20^. Brain MRI provides detailed information about brain structure and function^21^. Clinical applications of brain MRI have revealed the associated abnormalities that accompany multiple neurological and neuropsychiatric disorders^22–24^. Moreover, twin and family studies have shown that CMR and brain MRI traits are moderately to highly heritable^25–28^. For example, the left ventricular mass (LVM) has a heritability greater than 0.8^27^. The majority of brain structural MRI traits are highly heritable (heritability ranges from 0.6 to 0.8)^29^ and the heritability of brain functional connectivity is largely between 0.2 and 0.6^30^. A few recent genome-wide association study (GWAS) have been separately conducted on CMR^31–36^ and brain MRI traits^37–42^. Although MRI is widely used in clinical research and genetic mapping, few studies have used multi-organ MRI to examine heart-brain connections and identify the heart’s and brain’s shared genetic signatures.

In this paper, we investigated heart-brain connections using multi-organ imaging data obtained from over 40,000 subjects in the UK Biobank (UKB) study. By using a newly developed heart segmentation and feature extraction pipeline^43^, we generated 82 CMR traits from the raw short-axis, long-axis, and aortic cine images. These CMR traits included global measures of 4 cardiac chambers (the left ventricle (LV), right ventricle (RV), left atrium (LA), and right atrium (RA)) and 2 aortic sections (the ascending aorta (AAo) and descending aorta (DAo)), as well as regional phenotypes of the LV myocardial-wall thickness and strain (**Table S1** and **Supplementary Note**). Then we identified the relationships between the 82 CMR traits and a large number of the brain MRI traits discovered from multi-modality images, including T1 structural MRI (sMRI), diffusion brain MRI (dMRI), resting functional MRI (resting fMRI), and task functional MRI (task fMRI). These brain MRI traits provided fine details of basic brain morphometry^38^ (regional brain volumes and cortical thickness traits), brain structural connectivity^44^ (diffusion tensor imaging (DTI) parameters of white matter tracts), and brain intrinsic and extrinsic functional organizations^42^ (functional activity and connectivity at rest and during a task) (**Table S2**). To evaluate the genetic determinates underlying heart-brain connections, we performed GWAS for the 82 CMR traits to uncover the genetic architecture of heart and aorta. In comparison to existing GWAS of CMR traits^31–36^, our study used a much broader group of cardiac and aortic traits, which allowed us to identify the shared genetic components with a wide variety of brain-related complex traits and disorders. For example, Pirruccello, et al. ^35^ mainly focused on 9 measures of the right heart, Aung, et al. ^31^ analyzed 6 LV traits, and Thanaj, et al. ^36^ studied 3 traits of diastolic function. Our study design is summarized in **Fig. S1**. The GWAS results of 82 CMR traits can be explored and are freely available through the heart imaging genetics knowledge portal (Heart-KP) http://heartkp.org/.

## RESULTS

### Phenotypic heart-brain connections

To verify that the 82 CMR traits are well-defined and biologically meaningful, we first examined their reproducibility using the repeat scans obtained from the UKB repeat imaging visit (*n* = 2,903, average time between visits = 2 years). For each trait, we calculated the correlation between two observations from all revisited individuals. The average reproducibility was 0.652 (range = (0.369, 0.970), **Table S1**). A few volumetric traits had very high reproducibility (> 0.9), including the LV end-diastolic volume (LVEDV), LV myocardial mass (LVM), RV end-diastolic volume (RVEDV), RV end-systolic volume (RVESV), AAo maximum area, AAo minimum area, DAo maximum area, and DAo minimum area. The ejection fraction (such as the LV ejection fraction (LVEF)) and distensibility traits (such as the descending aorta distensibility (DAo aorta distensibility)) had the lowest reproducibility among all volumetric traits (mean = 0.573 and 0.537, respectively). In addition, the average reproducibility was 0.758 for the 17 wall thickness traits, 0.530 for the 7 longitudinal peak strains, 0.568 for the 17 circumferential strains, and 0.514 for the 17 radial strains. Overall, these results suggest that the extracted CMR traits have moderate to high within subject reliability and can consistently annotate the cardiac and aortic structure and function.

With the control of a large number of covariates, we examined the associations between the CMR traits and brain MRI traits in UKB individuals of white British ancestry (*n* = 31,875, Methods). Particularly, we adjusted for body size (height and weight) and body mass index in all our analyses. At the Bonferroni significance level (*P* < 1.33 × 10^-6^), CMR traits were associated with a wide variety of brain MRI traits, including regional brain volumes, cortical thickness, DTI parameters, and resting and task fMRI traits (Fig. 2A and **Fig. S2**). For example, the total brain volume had strong positive associations with volumetric measures of right and left heart as well as aortic sections, with the top ranked traits being the AAo maximum area, RA stroke volume (RASV), DAo minimum area, RVESV, RVEDV, and right atrium maximum volume (RAV max) (*β* > 0.112, *P* < 1.60 × 10^-61^, **Fig. S3**). These volumetric traits were also widely associated with regional brain volumes, after adjusting for the total brain volume. A large proportion of these associations were positive, and the negative ones were specifically observed in the cerebrospinal fluid (CSF) and ventricle volumes (e.g., the lateral ventricle and third ventricle). The wall thickness traits were positively associated with subcortical structures (e.g., the putamen, caudate, and hippocampus) and negatively associated with the lateral occipital and rostral middle frontal. In addition, cortical thickness measures were associated with multiple CMR traits, and the strongest associations were observed in the left ventricular cardiac output (LVCO) and DAo aorta distensibility (**Fig. S4**). These results uncover that heart structure and function have close relationships with basic brain morphometry.

**Fig. 2.**
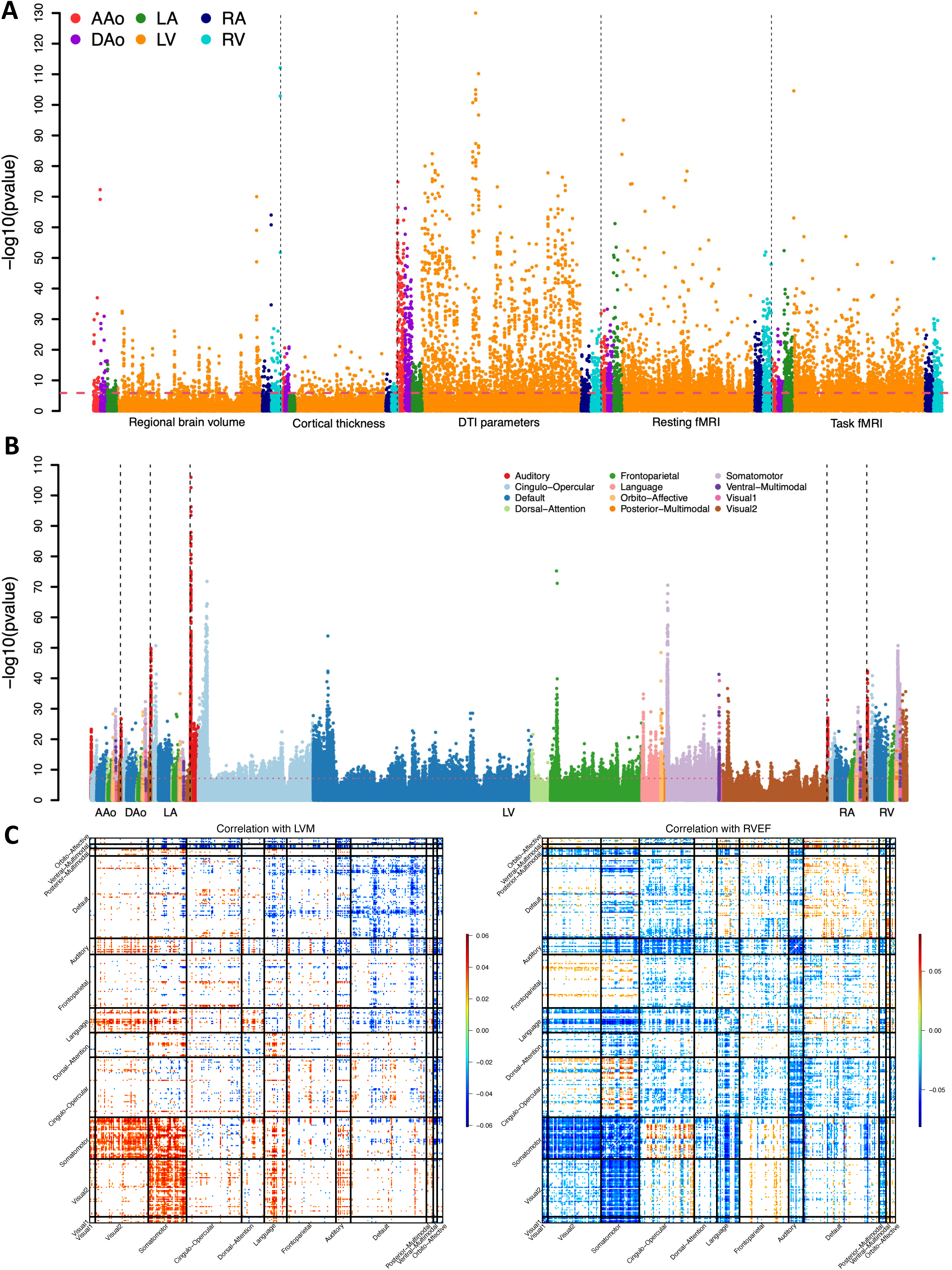
Phenotypic heart-brain associations. **(A)** The -log10(pvalue) of phenotypic correlations between 82 CMR traits and 5 groups of brain MRI traits, including 101 regional brain volumes, 63 cortical thickness traits, 110 DTI parameters, 92 resting fMRI traits, and 92 task fMRI traits. The dashed line indicates the Bonferroni-significance level (*P* < 1.33 × 10^-6^). Each CMR trait category is labeled with a different color. **(B)** The -log10(pvalue) of phenotype correlations between 82 CMR traits and 8,531 resting fMRI functional connectivity traits across 12 functional networks. The dashed line indicates the Bonferroni-significance level (*P* < 7.15 × 10^-8^). Functional networks are labeled with different colors. **(C)** Significant correlations between resting functional connectivity traits and left ventricular myocardial mass (LVM, left) and right ventricular ejection fraction (RVEF, right). AAo, ascending aorta; DAo, descending aorta; LA, left atrium; LV, left ventricle; RA, right atrium; RV, right ventricle; Visual1, the primary visual network; and Visual2, the secondary visual network.

CMR traits were also strongly correlated with brain structural and functional connectivity. For example, fractional anisotropy (FA) was a robust measure of brain structural connectivity and white matter microstructure. The average FA was positively associated with the LV ejection fraction (LVEF), DAo aorta distensibility, and AAo aorta distensibility (*β* > 0.035, *P* < 1.69 × 10^-8^, **Fig. S5**), and had negative correlations with many other CMR traits, including the LVM, LVEDV, left ventricular end-systolic volume (LVESV), global wall thickness, AAo maximum area, and DAo maximum area. For resting fMRI, both mean functional connectivity and mean amplitude (functional activity) were negatively associated with volumetric measures of 4 cardiac chambers, such as the LVCO, RV ejection fraction (RVEF), LA stroke volume (LASV), and RA ejection fraction (RAEF) (*β* < -0.062, *P* < 4.87 × 10^-24^, **Fig. S6**). Positive correlations were mainly located in the AAo aorta distensibility, DAo aorta distensibility, longitudinal strain, and peak circumferential strain. Similar patterns were observed for task fMRI traits (**Fig. S7**).

Brain functional traits could be more directly related to human behavioral and cognitive differences than brain structural traits^45^. To further discover fine details of CMR-connections with brain functions, we examined pairwise associations between 82 CMR traits and 64,620 high-resolution functional connectivity traits^42^ in resting fMRI. Significant associations (*P* < 7.15 × 10^-8^) were observed across the functional connectivity of the whole brain, with enrichment in specific brain functional areas and networks (Fig. 2B and **Fig. S8**). For example, the somatomotor network and its connectivity with the secondary visual network had strong associations with multiple CMR traits. Specifically, positive somatomotor-associations were observed in the LVM, LVESV, RVEDV, RVESV, RAV min, AAo aorta distensibility, DAo aorta distensibility, global peak circumferential strain, and global longitudinal peak strain (Fig. 2C and **Figs. S9-S16**), and negative correlations were observed in all the 4 ejection fraction traits (RVEF, LAEF, RAEF, and LVEF), LVCO, and global radial strain (Fig. 2D and **Figs. S17-S22**). Furthermore, the auditory network had strong negative associations with volumetric measures of heart, including all the 4 stroke volumes (LVSV, RVSV, LASV, and RASV), LVCO, LVEDV, RVEF, left atrium maximum volume (LAV max), and RAEF (**Figs. S17, S19, S21,** and **S23-S28**). On the other hand, the AAo aorta distensibility and DAo aorta distensibility were positivity correlated with the auditory network (**Figs. S13-S14**). There were also significant correlations with cognitive networks, including the default mode, cingulo-opercular, dorsal attention, and frontoparietal networks. For example, the default mode network was associated with the 4 ejection fraction traits, most of the associations were positive (**Fig. S29**). The default mode network was also negatively associated with other volumetric heart measures (such as the LVESV, LVSV, LVM, LASV, RVEDV, RVESV, RVSV, and RASV), whose associations were enriched in the hippocampal and visual areas of the default model network (**Figs. S30-S31**). For aortic measures, the associations were mainly located in the hippocampal areas of the default model network (**Fig. S32**). In summary, MRI-based endophenotype analysis reveals substantial neuro-cardiac interactions. Our results show the specific pattern of heart-brain connections and highlight important brain areas and networks that might be strongly related to heart health.

### Mediation analysis with cardiovascular risk factors and biomarkers

Environmental factors and biomarkers may play an important role in the underlying mechanisms of heart-brain interactions. Clear evidence from clinical and epidemiological studies exists that cardiovascular risk factors (such as hypertension, high cholesterol, high fasting glucose, metabolic syndrome, and chronic kidney disease) negatively influence brain health and neurocognitive performance^1,3,46,47^. However, it remains challenging to understand how these cardiac risk factors cause brain damages. To bridge this gap, we performed mediation analysis by using the 82 CMR traits as intermediate variables^48^. Specifically, we investigated whether cardiovascular risk factors could influence brain structure and function indirectly through heart conditions captured by the 82 CMR traits (Methods). We examined cardiovascular risk factors, including diastolic blood pressure, systolic blood pressure, diabetes mellitus, smoking, drinking, and basal metabolic rate, as well as 34 biomarkers collected in the UKB study.

Significant CMR-mediation effects were observed for 1,148 pairs between 34 cardiovascular risk factors/ biomarkers and 235 brain MRI traits (*P* < 2.21 × 10^-6^, **Table S3**). These results suggest that associations between cardiovascular risk factors and brain health were partially mediated by cardiac structure and function. For example, the diastolic blood pressure was widely associated with cortical thickness, regional brain volumes, DTI parameters, and functional connectivity (Fig. 3A). On average, 40.3% of diastolic blood pressure’s effects on DTI parameters were indirect and mediated through CMR traits (Fig. 3B). The other diastolic blood pressure’ effects (59.7%) could be direct effects on DTI parameters or indirect effects through non-cardiac mechanisms. The average proportion of mediated effects was 56.2% for cortical thickness, 38.6% for regional brain volumes, 37.5% for resting fMRI traits, and 33.6% for task fMRI traits. Similar meditation patterns were also observed on systolic blood pressure and hypertension (**Fig. S33**). Hypertension has been reported to be associated with cognitive dysfunction^49^ and brain white matter hyperintensities, which might be a consequence of chronic ischaemia caused by cerebral small vessel disease^50^. Mediation effects were also observed on multiple cardiovascular biomarkers, such as low-density lipoprotein (LDL) cholesterol, high-density lipoprotein (HDL) cholesterol, and triglyceride (**Fig. S34**).

**Fig. 3.**
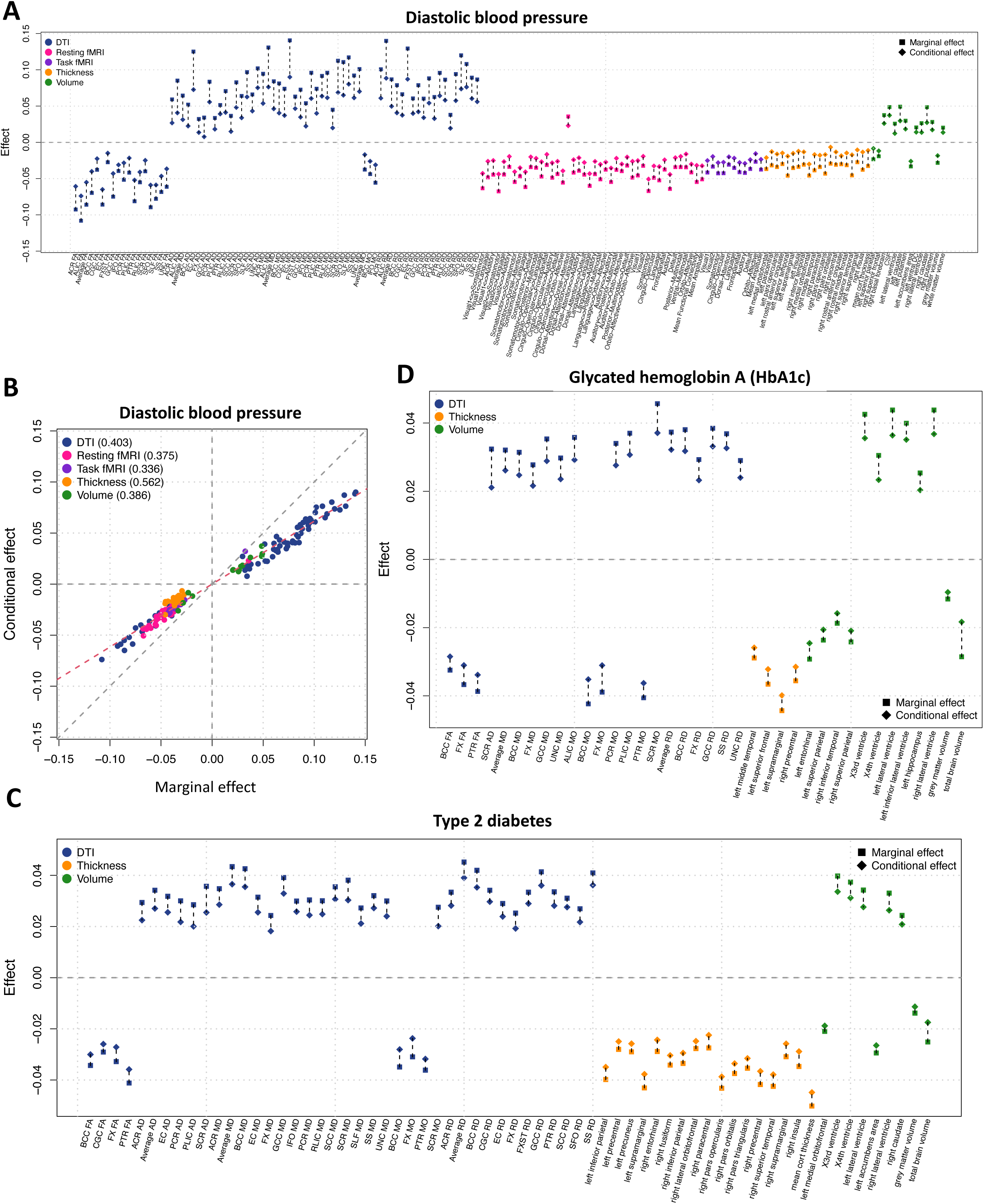
Effects of cardiovascular risk factors on brain MRI mediated through CMR traits. **(A)** We illustrate marginal effects (after adjusting for covariates) of diastolic blood pressure on brain MRI traits and the corresponding conditional effects after further adjusting for CMR traits. The difference between marginal and condition effects indicates the indirect effect mediated through CMR traits. As illustrated in **(B)**, the average proportion of CMR-mediated diastolic blood pressure’s effects was 40.3% for DTI parameters, 37.5% for resting fMRI, 33.6% for task fMRI, 56.2% for cortical thickness, and 38.6% for regional brain volumes. **(C)** Marginal effects (after adjusting for covariates) of type 2 diabetes (ICD 10 code E11.9) on brain MRI traits and the corresponding conditional effects after further adjusting for CMR traits. **(D)** Marginal effects (after adjusting for covariates) of glycated hemoglobin A (HbA1c) on brain MRI traits and the corresponding conditional effects after further adjusting for CMR traits.

CMR-mediated effects of type 2 diabetes (T2D) on the brain were observed (Fig. 3C). Insulin signaling plays a critical role in brain health and patients with diabetes are at higher risk of cognitive impairment and dementia^51^. The average proportion of CMR-mediated T2D effects was 16.8% for DTI parameters, 10.3% for cortical thickness, and 13.1% for regional brain volumes. For example, T2D was negatively related to total brain volume (*β* = -0.025, *P* = 9.64 × 10^-9^). After adjusting for the CMR traits, the effect became smaller (*β* = -0.017, *P* = 5.80 × 10^-5^), suggesting that 32% (0.008/0.025) of the T2D effects were mediated through CMR traits. In addition, mediation effects were observed on glycated hemoglobin A (HbA1c) and glucose, which were biomarkers of diabetes. High HbA1c level (hyperglycaemia) is associated with worse brain health and cognitive functions^52^, such as memory loss, poorer processing speed, attention, concentration, and executive functions^53,54^. As the primary source of energy for the brain, glucose metabolism plays an important role in the physiological and pathological functioning of the brain^55^. About 15.5% of HbA1c’s effects on DTI parameters and 14.1% of glucose’s effects on cortical thickness were CMR-mediated (Fig. 3D and **Fig. S35**). Other biomarkers whose effects on brain MRI were mediated by CMR included creatinine, urate, total protein, and gamma-glutamyl transpeptidase (GGT) (**Figs. S36-S37**). Creatinine, total protein, and urate are kidney biomarkers of renal function, which have been consistently associated with white matter brain deficits^56^ and neurodegenerative diseases^57^. GGT is a liver biomarker of potential hepatic or biliary diseases and has been reported to be associated with cognitive decline and increased risk of dementia^58^. In summary, multiple categories of factors and biomarkers strongly influence brain health, and their influences are partly mediated by cardiovascular conditions. Understanding these mediation mechanisms underlying heart-brain connections might assist in preventing and detecting brain diseases.

### Heritability and the associated genetic loci of 82 CMR traits

We estimated the single-nucleotide polymorphism (SNP) heritability for the 82 CMR traits using UKB individuals of white British ancestry^59^ (*n* = 31,875, Methods). The mean heritability (*h^2^*) was 23.0% for the 82 traits (range = (7.06%, 70.3%), Fig. 4A), all of which remained significant adjusting for multiple testing using the Benjamini-Hochberg procedure to control the false discovery rate (FDR) at 0.05 level (**Table S4**). The *h^2^* of the AAo maximum area, AAo minimum area, DAo maximum area, and DAo minimum area was larger than 50%. Among cardiac traits, the global wall thickness, RVESV, RVEDV, LVESV, LVEDV, and LVM had the highest heritability (*h^2^* > 37.7%). As expected, more reproducible CMR traits show higher heritability (correlation = 0.88, *P* < 2.2 × 10^-16^, **Fig. S38**). To identify reliable genetic signals for these CMR traits, large-scale samples from a homogeneous cohort are needed, especially for CMR traits with relatively low heritability.

**Fig. 4.**
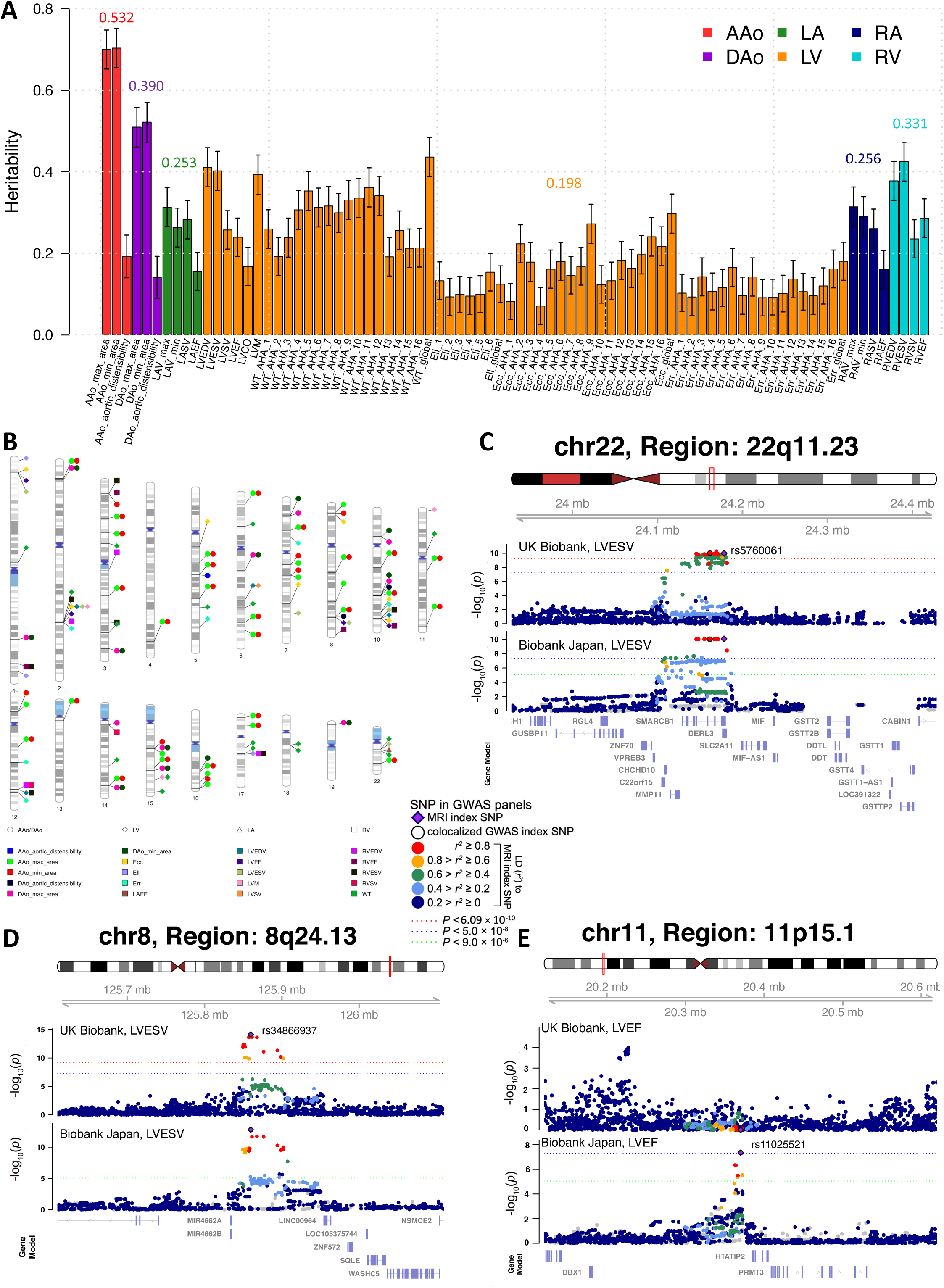
Genetics of CMR traits in the UK Biobank. **(A)** SNP heritability of 82 CMR traits across the 6 categories. The *x* axis displays the ID of CMR traits, see Table S1 for full names of these traits. AAo, ascending aorta; DAo, descending aorta; LA, left atrium; LV, left ventricle; RA, right atrium; and RV, right ventricle. The average heritability of each category is labeled. **(B)** Ideogram of 80 genomic regions associated with CMR traits (*P* < 6.09 × 10^-10^). **(C)** Left ventricular end-systolic volume (LVESV) was associated with the 22q11.23 region in both UK Biobank and Biobank Japan studies (shared index variant rs5760061). **(D)** Left ventricular end-systolic volume (LVESV) was associated with the 8q24.13 region in both UK Biobank and Biobank Japan studies (shared index variant rs34866937). **(E)** Left ventricular ejection fraction (LVEF) was associated with the 8q24.13 region in the Biobank Japan study (index variant rs11025521), but not in the UK Biobank study.

We next performed GWAS for the 82 CMR traits using this white British cohort (*n* = 31,875, Methods). All Manhattan and QQ plots can be browsed through the server on Heart-KP (http://67.205.180.40:443/). The intercepts of linkage disequilibrium (LD) score regression (LDSC)^60^ were all close to one, suggesting no genomic inflation of test statistics due to confounding factors (mean intercept = 1.0002, range = (0.987, 1.019)). At the significance level 6.09 × 10^-10^ (5 × 10^-8^/82, that is, the standard GWAS significance threshold, additionally Bonferroni-adjusted for the 82 traits), we identified independent (LD *r^2^* < 0.1) significant associations in 80 genomic regions (cytogenetic bands) for 49 CMR traits, including 35 for left ventricular, 35 for ascending aorta, 14 for descending aorta, 10 for right ventricular, and 1 for left atrium (Fig. 4B and **Table S5**, Methods). Detailed interpretations of these identified regions can be found in next Section. When relaxing the significance level to 5 × 10^-8^, there were 156 significant genomic regions (76 more) for 72 CMR traits.

On average, the 80 (covering 12.3% of genetic variants) and 76 (10.9% variants) CMR-associated genetic regions separately explained 20.5% (that is, 4.7%/23%) and 12.6% (2.9%/23%) of the heritability of CMR traits, indicating that about 66.9% (15.4%/23%) of the heritability remained unexplained by these significant regions (**Fig. S39**). These results illustrate a highly polygenic genetic architecture for CMR traits^61^. In addition, the 80 CMR-associated regions explained 14.3% (6.8%/47.6%) of the SNP heritability of DTI parameters, suggesting that the genetic effects on white matter microstructure were enriched in these CMR-associated regions. We next used the previously identified DTI-associated genes as an annotation^40^ to perform partitioned heritability enrichment analysis^62^ for the 82 CMR traits. At FDR 5% level, heritability of 26 CMR traits was significantly (*P* range = (6.89 × 10^-5^, 1.15 × 10^-2^)) enriched in genomic regions influencing DTI parameters (**Fig. S40**). In addition to DTI parameters, other brain MRI traits showed overlapped genetic influences with CMR traits, but with less evidence for enrichment. Specifically, the 80 CMR-associated regions can explain 13.7% (1.7%/12.8%) of the heritability of task fMRI traits, 13.4% (2.8%/20.9%) of cortical thickness, 12.5% (4.6%/36.8%) of regional brain volumes, and 12.2% (1.8%/14.8%) of resting fMRI traits. Overall, these results suggest that the CMR traits had shared heritability with brain MRI traits, especially with DTI parameters measuring white matter microstructure.

To replicate the identified loci, we performed separate GWAS using hold-out datasets in the UKB study that were independent from our discovery dataset. First, we repeated GWAS on a European dataset with 8,252 subjects (Methods). For the 248 independent (LD *r^2^* < 0.1) CMR-variant associations in the 80 genomic regions, 57 (23%, in 25 regions) passed the Bonferroni significance level (2.02 × 10^-4^, 0.05/248) in this European validation GWAS, and 184 (74.19%, in 61 regions) were significant at nominal significance level (0.05). All the 184 associations had concordant directions in the two independent GWAS and the correlation of their genetic effects was 0.93 (**Fig. S41** and **Table S6**). These results show a high degree of generalizability of our GWAS findings among European cohorts. We also performed GWAS on two non-European UKB validation datasets: the UKB Asian (UKBA, *n* = 500) and UKB Black (UKBBL, *n* = 271). One association between 8q24.3 and the RVEF passed the Bonferroni significance level (*P* = 6.20 × 10^-5^) in UKBA, and 13 more regions passed nominal significance level. For UKBB, 11 regions passed the nominal significance level and none of them survived the Bonferroni significance level, which may partially be due to the small sample size of this non-European GWAS.

Additionally, we evaluated the ancestry specific effects using Asian GWAS summary statistics of three CMR traits (analogous to the LVEDV, LVESV, and LVEF^34^), which were generated from 19,000 subjects in the BioBank Japan (BBJ) study^63^. At the 5 × 10^-8^ threshold, the BBJ identified independent (LD *r^2^* < 0.1) significant associations in 6 regions (2p14, 11p15.1, 22q11.23, 8q24.13, 10q22.2, and 18q12.1), 4 of which (22q11.23, 8q24.13, 10q22.2, and 18q12.1) were among the 156 regions we discovered based on data from UKB white British. Specifically, the 22q11.23 and 8q24.13 regions were significantly associated with the LVSEV in both UKB and BBJ studies (Figs. 4C-4D). The 10q22.2 and 18q12.1 regions were separately associated with the LVEDV and LVESV in BBJ but not in UKB (**Figs. S42-S43**). Instead, the two regions were significantly associated with the wall thickness traits in UKB (*P* < 6.77 × 10^-10^). Additionally, only CMR traits in BBJ were associated with the 11p15.1 and 2q14 regions (Figs. 2E and **S44**), likely representing population specific genetic influences in Asian population.

Finally, we constructed polygenic risk scores (PRS) via lassosum^64^ to evaluate the out of sample prediction power of the discovery GWAS results. We used 2,000 subjects from our European validation GWAS as validation data and evaluated the performance on the left independent subjects (*n* = 5,551). Among the 82 CMR traits, 73 had significant PRS at FDR 5% level (*P* range = (7.38 × 10^-136^, 3.49 × 10^-2^), **Table S7**). The highest incremental R-squared (after adjusting the effects of age, sex, and ten genetic principal components (PCs)) was observed on the AAo minimum area and AAo maximum area (8.52% and 8.36%, respectively). Other traits whose incremental R-squared larger than 1% included the DAo minimum area, DAo maximum area, RVEDV, RVESV, LAV max, and wall thickness traits (R-squared range = (1.01%, 3.32%), *P* < 4.33 × 10^-15^). To evaluate the cross-population performance, PRS was also constructed on UKB white British discovery GWAS data using BBJ GWAS summary statistics of the LVEDV, LVESV, and LVEF. We found that the PRS of these three traits were all significant in the UKB (*P* range = (1.58 × 10^-11^, 8.13 × 10^-7^), R-squared range= (3.90 × 10^-4^, 1.35 × 10^-3^)). The prediction accuracy was lower than that in the above within European prediction analysis (R-squared range= (7.72 × 10^-3^, 9.67 × 10^-3^)), which may be explained by the smaller training GWAS sample size in BBJ^65^ and the differences of genetic effects and LD among European and Asian populations^66^. More efforts are needed to identify causal variants associated with CMR traits in global diverse populations and quantify population-specific heterogeneity of genetic effects.

### Pleiotropy of genetic variants across body systems

To identify the shared genetic effects between CMR traits and complex traits, we carried out association lookups for independent (LD *r^2^* < 0.1) significant variants (and variants in their LD, *r*^2^ ≥ 0.6, *P* < 6.09 × 10^-10^) detected in our UKB white British GWAS. In the NHGRI-EBI GWAS catalog^67^, our results tagged variants that have been linked to a wide range of traits and diseases, including heart diseases, heart structure and function, blood pressure, lipid traits, blood traits, diabetes, stroke, neurological and neuropsychiatric disorders, psychological traits, cognitive traits, lung function, parental longevity, smoking, drinking, and sleep. To evaluate whether two associated genetic signals were consistent with the shared causal variant, we applied the Bayesian colocalization analysis^68^ for CMR traits and selected complex traits whose GWAS summary statistics were publicly available. Evidence of pairwise colocalization was defined as having a posterior probability of the shared causal variant hypothesis (PPH4) > 0.8^68,69^.

First, our results replicated 25 genomic regions that have been linked to cardiac and aortic traits in previous GWAS, including 5 regions with left ventricular traits (such as 22q11.23 with fractional shortening^70^, ejection fraction^63^, and left ventricular internal dimension^63^, **Fig. S45**); 17 regions with heart rate and electrocardiographic traits (such as 1p36.32, 5q33.2, 10q25.2, 12q24.21, and 14q24.2 with PR interval^71^ and QRS duration^72^) (**Figs. S46-S50**); and 6 regions with aortic measures (such as in 15q21.1 with thoracic aortic aneurysms and dissections^73^ and in 15q24.1 with aortic root size^70^, **Figs. S51-S52**). In addition, 24 regions had shared associations (LD *r*^2^ ≥ 0.6) with cardiovascular diseases, including 9 regions with coronary artery disease^74,75^ (such as 17p13.3 and 12q24.12, **Figs. S53-S54**); 9 regions with atrial fibrillation^76,77^ (such as 6p21.2, 1p13.1, 2q31.2, 8q24.13, 15q26.3, and 22q12.1, Figs. 5A and **S55-S59**); 6q22.33, 10q23.33, and 11p15.5 with hypertension^78,79^ (**Figs. S60-S62**); 15q21.1 and 13q12.11 with abdominal aortic aneurysm^80,81^ (**Figs. S51** and **S63**); 14q24.2 with mitral valve prolapse^82^ (**Fig. S50**); and 10q26.11 and 1p36.13 with idiopathic dilated cardiomyopathy^83^ (**Figs. S64-S65**). There was widespread evidence of colocalization on many on many loci (PPH4 > 0.849). Additionally, 41 of the 80 genomic regions were associated with blood pressure traits, such as systolic blood pressure, diastolic blood pressure, pulse pressure^84^, and mean arterial pressure^85^ (Figs. 5B and **S66-S85**). CMR traits were in LD (*r*^2^ ≥ 0.6) with various cardiovascular biomarkers in 32 genomic regions, such as 12q24.12, 6q22.33, 5q31.1, and 17q12 with blood metabolite levels^86^ and lipid traits^87,88^ (including HDL, LDL, and triglycerides, **Figs. S54, S60, S71**, and **S86**); and 3p13, 3p25.1, 5q15, 12q14.1, and 22q13.1 with red blood cell count^89^, blood protein levels^90^, red cell distribution width^91^, and plateletcrit^89^ (**Figs. S87-S91**).

**Fig. 5.**
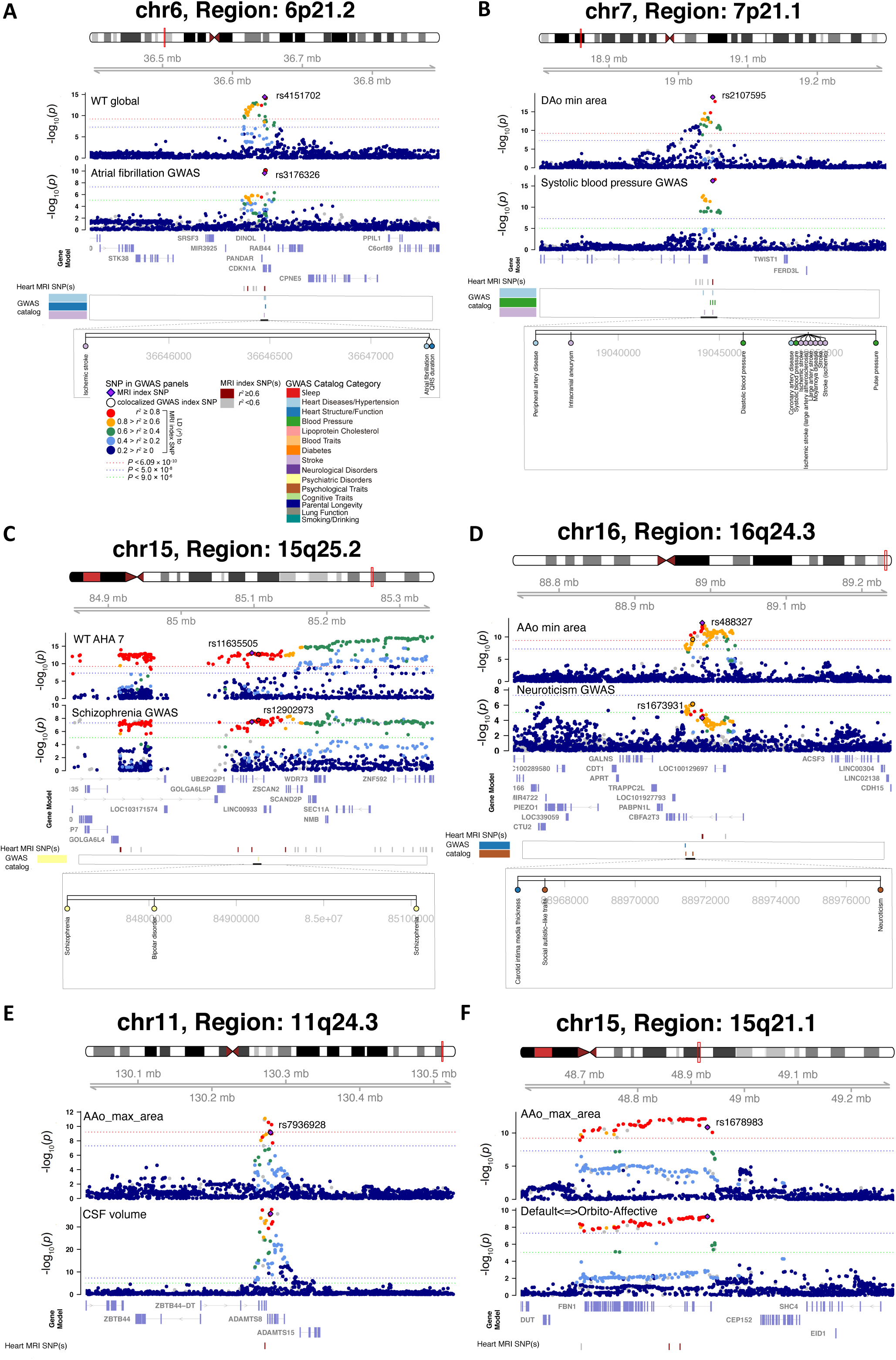
Selected genetic loci that were associated with both CMR trait and other complex traits and diseases. **(A)** In 6p21.2, we observed colocalization between the global myocardial-wall thickness at end-diastole (WT global, index variant rs4151702) and atrial fibrillation (index variant rs3176326). The posterior probability of Bayesian colocalization analysis for the shared causal variant hypothesis (PPH4) is 0.904. In this region, the WT global was also in LD (*r*^2^ ≥ 0.6) with ischemic stroke. **(B)** In 7p21.1, we observed colocalization between the ascending aorta minimum area (DAo min area) and systolic blood pressure (shared index variant rs2107595, PPH4 = 0.960). In this region, the DAo min area was also in LD (*r*^2^ ≥ 0.6) with stroke, intracranial aneurysm, coronary artery disease, and moyamoya disease. **(C)** In 15q25.2, we observed colocalization between the regional myocardial-wall thickness at end-diastole (WT AHA 7, index variant rs11635505) and schizophrenia (index variant rs12902973, PPH4 = 0.890). In this region, the WT AHA 7 was also in LD (*r*^2^ ≥ 0.6) with bipolar disorder. **(D)** In 16q24.3, we observed colocalization between the ascending aorta minimum area (AAo min area, index variant rs488327) and neuroticism (index variant rs1673931, PPH4 = 0.991). **(E)** We illustrated the colocalization between the ascending aorta maximum area (AAo max area) and cerebrospinal fluid (CSF) volume (shared index variant rs7936928) in 11q24.3 (PPH4 = 0.902). **(F)** We illustrated the colocalization between the AAo max area and functional connectivity between the default mode and orbito-affective networks (Default <-> Orbito-Affective, shared index variant rs1678983) in 15q21.1 (PPH4 = 0.964).

We found genetic pleiotropy between CMR traits and multiple brain-related complex traits and disorders. In 6p21.2, 7p21.1, and 12q24.12 regions, CMR traits were in LD (*r*^2^ ≥ 0.6) with stroke^92^ (such as ischemic stroke, large artery stroke, and small-vessel ischemic stroke), intracranial aneurysm^93^, and Moyamoya disease^94^ (Figs. 5A-5B and **S54**). The index variants of 7p21.1 (rs2107595) and 12q24.12 (rs597808) were expression quantitative trait loci (eQTLs) of *TWIST1, ALDH2,* and *NAA25* in human brain tissues^95^, suggesting that these CMR-associated variants were known to affect gene expression levels in human brain. When the GWAS significance threshold was relaxed to 5 × 10^-8^, the shared associations (LD *r*^2^ ≥ 0.6) with stroke were found in four more CMR-significant loci (12q13.3, 16q23.1, 4q25, 7p15.1). CMR traits were also in LD (*r*^2^ ≥ 0.6) with neurodegenerative and neuropsychiatric disorders, such as in 17q21.31 (rs62062271, brain eQTL of multiple genes such as *CRHR1* and *WNT3*) with Parkinson’s disease^96^, corticobasal degeneration^97^, multiple system atrophy^98^, Alzheimer’s disease^99^ and associated cognitive impairment^100^, autism spectrum disorder^101^, and progressive supranuclear palsy^102^ (**Fig. S92**); in 12p12.1 (rs4148674, brain eQTL of *ABCC9*) with hippocampal sclerosis of aging^103^ (**Fig. S78**); in 15q25.2 (rs11635505, brain eQTL of *UBE2Q2L, WDR73,* and *GOLGA6L4*) and 17p13.3 (rs2281727, brain eQTL of *SRR*) with schizophrenia^104^ (Figs. 5C and **S53**); in 17q12 (rs903503, brain eQTL of multiple genes such as *PGAP3* and *MED24*) and 6q22.31 (rs1334489, brain eQTL of *CEP85L*) with bipolar disorder^105,106^ (**Figs. S86** and **S93**); and in 3p14.3 (rs2686630, brain eQTL of *ABHD6*) with eating disorder^107^ (**Fig. S94**). In addition, CMR traits were in LD (*r*^2^ ≥ 0.6) with mental health traits, such as in 16q24.3, 17q21.31, 8p23.1 (rs903503, brain eQTL of multiple genes such as *FAM167A* and *DEFB134*), and 11p11.2 (rs11039348, brain eQTL of multiple genes such as *FAM180B* and *SLC39A13*) with neuroticism^108^, depressive symptoms^109^, subjective well-being^110^, and risk-taking tendency^111^ (Figs. 5D**, S92** and **S95-S96**). For cognitive traits and education, we tagged 17q21.31, 11p11.2, and 11q13.3 with cognitive function^112^ and educational attainment^113^ (**Figs. S92, S96**, and **S97**); 7q32.1 (rs2307036, brain eQTL of *ATP6V1FNB*) with reading disability^114^ (**Fig. S98**); and in 12q24.12 (rs597808, brain eQTL of *ALDH2* and *NAA25*) with reaction time^112^ (**Fig. S54**). We also found shared associations (LD *r*^2^ ≥ 0.6) in 11q24.3 (rs11222084, brain eQTL of *ADAMTS8*), 12q24.12 (rs7310615, brain eQTL of *ALDH2* and *NAA25*), 17p13.3 (rs10852923, brain eQTL of *SRR* and *SMG6*), 17q12 (rs10852923, brain eQTL of *SRR* and *SMG6*), and 17q21.31 with DTI parameters^40^ (**Figs. S99-S103**); in 11q24.3 (rs7936928, brain eQTL of *ADAMTS8*), 3p13, 11p11.2 (rs7107356, brain eQTL of multiple genes such as *FAM180B* and *SLC39A13*), 17q21.31 (rs118087478, brain eQTL of multiple genes such as *ARL17A* and *ARL17B*) with regional brain volumes^38^ (Fig. 5E and **Figs. S104-S106**); and in 15q21.1, 8p23.1 (rs10093774, brain eQTL of multiple genes such as *RP1L1* and *FAM167A*), 10q23.33 (rs10093774, brain eQTL of *HELLS*), 11q13.3 (rs10093774, brain eQTL of *HELLS*), and 17q21.31 (rs62062271, brain eQTL of multiple genes such as *ARL17A* and *LRRC37A2*) with fMRI traits^42^ (Fig. 5F and **Figs. S107-S110**). Furthermore, CMR traits were colocalized with these brain-related complex traits in many regions, such as in 15q25.2 with schizophrenia, in 16q24.3 with neuroticism, in 11q24.3 with cerebrospinal fluid volume, and in 15q21.1 with functional connectivity (Figs. 5C-5F, PPH4 > 0.890). There is substantial evidence to support the interaction between cardiovascular health status and brain health. For example, people with better heart healthy have better cognitive abilities^115^ and lower risk for brain disorders, such as stroke and Alzheimer’s disease^116^. In addition, mental health disorders may result in biological processes and behaviors that related to cardiovascular risk factors, such as smoking initiation and physical inactivity^11,117^. Our results reveal that cardiovascular conditions have substantial genetic overlaps with both mental health and cognitive functions, which may partially explain the heart-brain connections. The majority of shared genetic variants were found to be eQTLs in brain tissues. Future studies are needed to explore the potential genomic pathways and mechanisms underlying the shared genetic influences between heart and brain.

Genetic overlaps with other diseases and complex traits were also observed. For example, RVEDV is in LD (*r*^2^ ≥ 0.6) with type 1 diabetes^118^, T2D^119^, coronary artery disease^75^, and parental longevity^120^ in the 12q24.12 region (**Fig. S54**). CMR traits were in LD (*r*^2^ ≥ 0.6) with lung function and diseases in 7 regions, such as in 17q12 with asthma^121^ (**Fig. S86**); in 17q21.31 with idiopathic pulmonary fibrosis^122^ and interstitial lung disease^123^ (**Fig. S92**); and in 10q26.11, 3p22.1, 5q31.1, and 15q23 with lung function (FEV1/FVC)^124^ (**Figs. S64, S68, S71**, and **S81**). We also found shared genetic associations (LD *r*^2^ ≥ 0.6) in 11p11.2, 12q24.12, and 17q12 with smoking^111^ (**Figs. S96, S54**, and **S86**); and in 17p13.3, 11p11.2, and 17q21.31 with alcohol consumption and alcohol use disorder^125^ (**Figs. S53, S96**, and **S92**). All the above results can be found in **Table S8**.

### Genetic correlations with brain disorders and complex traits

First, we examined the genetic correlations (GC)^126^ among the 82 CMR traits. Strong genetic correlations were observed within and between categories of CMR traits (**Fig. S111**). For example, the RVEDV was genetically correlated with other RV traits, including RVSV and RVESV (GC > 0.85, *P* < 1.01 × 10^-137^), as well as RVEF (GC = -0.47, *P* = 4.93 × 10^-10^). The RVEDV was also significantly correlated with CMR traits from other categories, such as AAo maximum area and DAo maximum area (GC > 0.38, *P* < 1.36 × 10^-13^), LASV and RASV (GC > 0.37, *P* < 3.02 × 10^-7^), LVEDV, LVESV, and LVM (GC > 0.594, *P* < 2.74 × 10^-35^), and LVEF (GC = -0.55, *P* = 5.25 × 10^-20^). In addition, we found a strong relationship between phenotypic and genetic correlations among all CMR traits (*β* = 0.751, *P* < 2 × 10^-16^).

Next, we examined the genetic correlations between 82 CMR traits and 33 complex traits, many of which were colocalized traits in the above section. At the FDR 5% level (82 × 33 tests), the CMR traits were significantly associated with heart diseases, lung function, cardiovascular risk factors, and brain-related complex traits and diseases (**Table S9**). For example, hypertension had strong genetic correlations with aortic traits and LV traits (Fig. 6A). The strongest correlation between LV traits and hypertension was found in wall thickness traits (GC range = (0.237, 0.406), *P* < 6.18 × 10^-9^), which were also significantly associated with coronary artery disease, T2D, and stroke. In addition, atrial fibrillation was significantly associated with aortic, LA, and RA traits (|GC| range = (0.175, 0.252), *P* < 7.59 × 10^-4^), suggesting that atrial fibrillation might have a higher genetic similarity with LA/RA traits than LV/RV traits.

**Fig. 6.**
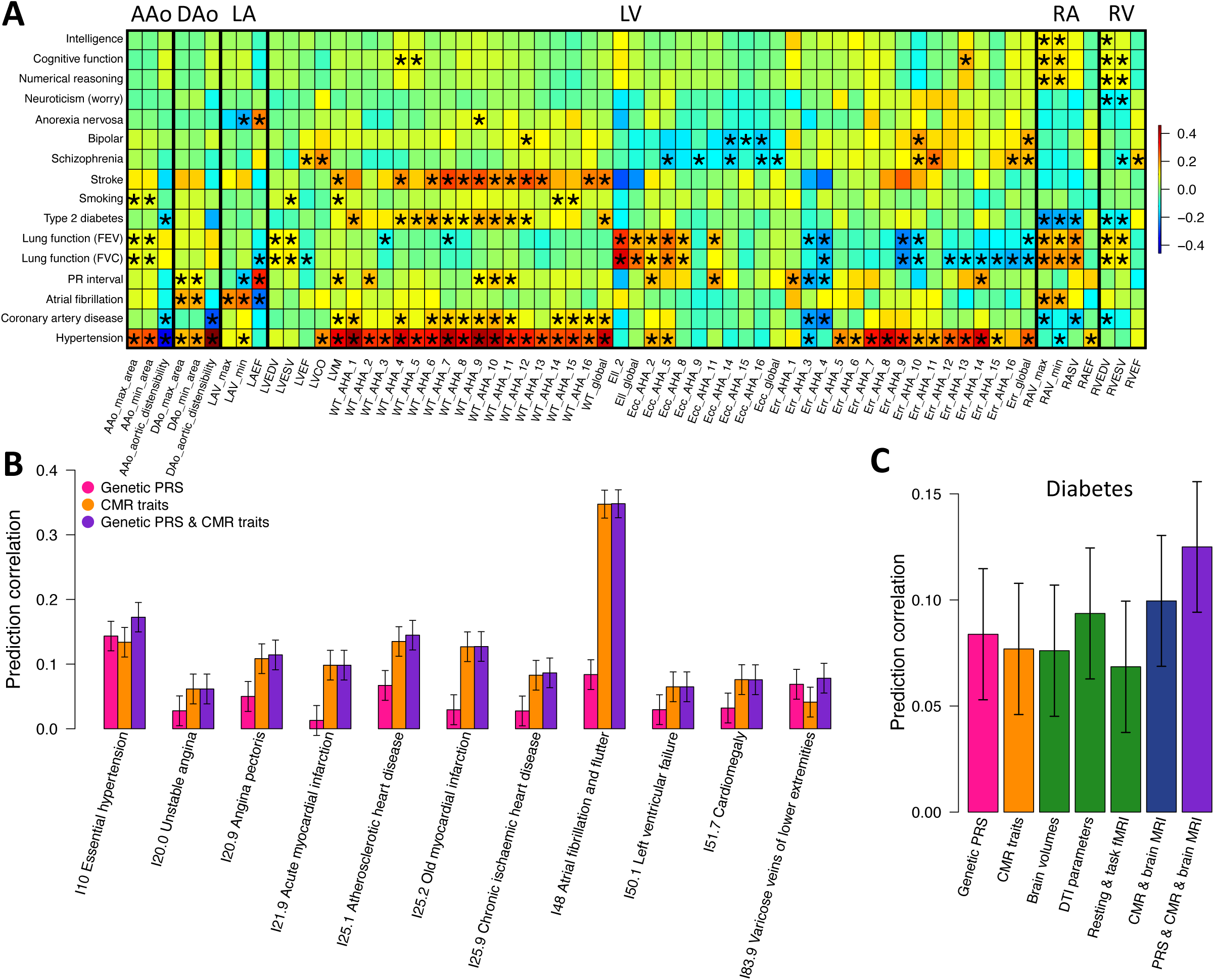
Genetic correlations and integrative prediction. **(A)** We illustrated selected genetic correlations between CMR traits (*x* axis) and complex traits and diseases (*y* axis). The asterisks highlight significant genetic correlations after at the FDR 5% level. **(B)** Predicting heart diseases using genetic variants and CMR traits. Genetic PRS, polygenic risk scores of genetic variants. **(C)** The accuracy of diabetes prediction analysis using different types of data. Brain volume, region brain volumes; and DTI parameters, diffusion tensor imaging parameters.

In both schizophrenia and bipolar disorder, multiple LV traits showed significant genetic correlations. Specifically, on LVCO, LVEF, radial strains, and wall thickness, positive genetic correlations were observed with schizophrenia and bipolar disorder (GC range = (0.122, 0.236), *P* < 9.37 × 10^-3^). Negative genetic correlations with the two brain disorders were observed on peak circumferential strains (GC range = (−0.168, -0.102), *P* < 1.26 × 10^-3^). Additionally, anorexia nervosa (eating disorder) was significant associated with LAV min and LAEF (|GC| range = (0.189, 0.220), *P* < 8.11 × 10^-3^) and cognitive traits and neuroticism were mainly associated with right heart traits (RA and RV traits). For example, intelligence was genetically correlated with the RAV max, RAV min, and RVEDV (GC range = (0.08, 0.12), *P* < 7.10 × 10^-3^). Lung functions (FEV and FVC) had genetic correlations with multiple CMR traits, with longitudinal strains showing the strongest correlations (GC range = (0.296, 0.357), *P* < 7.29 × 10^-6^). There were more associations with other complex traits analyzed in previous GWAS, such as smoking, PR interval, blood pressure, education, risky behaviors, lipid traits, mean corpuscular hemoglobin, and mean corpuscular volume (**Fig. S112A**). We also found very high genetic correlations with previously reported four LV traits^34^ (GC > 0.865, *P* < 6.28 × 10^-99^) (**Fig. S112B**). In summary, genome-wide genetic similarities have been found between CMR traits and a wide range of complex traits and diseases. Discovering such genetic co-variations may improve our understanding of genetic pathways of clinical outcomes from a multi-organ perspective.

### Biological and gene level analyses

We performed gene level association test using GWAS summary statistics of the 82 CMR traits with MAGMA^127^. We identified 160 significant genes for 49 CMR traits (*P* < 3.24 × 10^-8^, Bonferroni-adjusted for 82 traits) (**Table S10**). Next, we mapped significant variants (*P* < 6.09 × 10^-10^) to genes by combining evidence of physical position, eQTL (expression quantitative trait loci) association, and 3D chromatin (Hi-C) interaction via FUMA^128^. We found 576 mapped genes, 434 of which were not identified in MAGMA (**Table S11**). Moreover, 90 MAGMA or FUMA-significant genes had a high probability of being loss-of-function (LoF) intolerant^129^ (pLI > 0.98), indicating significant enrichment of intolerant of LoF variation among these CMR-associated genes (*P* = 1.58 × 10^-4^).

Ten genes (*CACNA1I, REN, ATP1A1, CACNB2, KCNJ8, PDE11A, AHR, ESR1, CYP2C9,* and *ABCC9*) were targets for 32 cardiovascular system drugs^130^, such as 15 calcium channel blockers (anatomical therapeutic chemical (ATC) code: C08) to lower blood pressure, 5 cardiac glycosides (C01A) to treat heart failure and irregular heartbeats, and 3 antiarrhythmics (C01B) to treat heart rhythm disorders (**Table S12**). Three (*CACNA1I, ESR1,* and *CYP2C9*) of these 10 genes and 4 more CMR-associated genes (*ALDH2, HDAC9, NPSR1,* and *TRPA1*) were targets for 11 nervous system drugs, including 4 antiepileptic drugs (N03A) used in the treatment of epileptic seizures and 2 drugs for addictive disorders (N07B). Some drug target genes have known biological functions both in the heart and brain. For example, *ALDH2* plays an important role in clearance of toxic aldehydes, which is an important mechanism related to myocardial and cerebral ischemia/reperfusion injury^131^. Therefore, *ALDH2* has been proposed to be a protective target for heart and brain diseases/dysfunctions triggered by ischemic injury and related risk factors^132,133^. Our results may help identify new drug targets or identify drugs that could be repurposed.

MAGMA gene-set analysis was performed to prioritize the enriched biological pathways, which yielded 36 significantly enriched gene sets at FDR 5% level (*P* < 3.0 × 10^-6^, **Table S13**). Multiple pathways related to cardiac development and heart disease were detected, including “go cardiac septum morphogenesis” (Gene Ontology [GO]: 0060411), “go cardiac muscle tissue development” (GO: 0048738), “go cardiac muscle tissue regeneration” (GO: 0061026), “go cardiac chamber development” (GO: 0003205), “go cardiac chamber development” (GO: 0003205), “go cardiac muscle contraction” (GO: 0060048), “go cardiac ventricle development” (GO: 0003231), “go vasculogenesis involved in coronary vascular morphogenesis” (GO:0060979), “go adult heart development” (GO: 0007512), “go heart development” (GO: 0007507), and “bruneau septation atrial” (M5223). Finally, we performed partitioned heritability analyses^62^ to identify the tissues and cell types^134^ where genetic variation leaded to differences in CMR traits. At FDR 5% level, the most significant heritability enrichments were found in active gene regulation regions of heart and muscle tissues, supporting the biological validity of the identified GWAS signals for CMR traits (**Fig. S113**).

### Complex traits and diseases prediction using genetic and multi-organ MRI data

In this section, we examined the prediction performance of CMR traits for 95 complex traits and diseases. We evaluated the prediction performance in a training, validation, and testing design and removed the effects of age (at imaging), age-squared, sex, age-sex interaction, age-squared-sex interaction, imaging site, height, weight, body mass index, and the top 40 genetic PCs (Methods). At FDR 5% level, the CMR traits had significant prediction power on 44 traits and diseases, including diseases of the circulatory system (ICD-10 group code: “I”); endocrine, nutritional and metabolic diseases (ICD-10: “E”); endocrine, nutritional and metabolic diseases (ICD-10: “F”); mental health and cognitive traits; cardiovascular diseases (self-reported), biomarkers, and risk factors; and disease family history (prediction correlation range = (0.028, 0.5), *P* range = (1.60 × 10^-2^, 5.05 × 10^-234^), **Fig. S114**). For example, CMR traits significantly predicted 14 circulatory system diseases, including atrial fibrillation, essential (primary) hypertension, angina pectoris, atherosclerotic heart disease, old myocardial infarction, chronic ischaemic heart disease, and acute myocardial infarction (*β* > 0.097, *P* < 4.99 × 10^-17^). They also had high prediction power for cardiovascular risk factors, such as systolic blood pressure (*β* = 0.367, *P* = 3.96 × 10^-219^) and diabetes (self-reported) (*β* = 0.114, *P* = 9.50 × 10^-23^). Moreover, CMR traits can predict brain-related psychological factors and cognitive traits, such as risk-taking (*β* = 0.068, *P* = 4.81 × 10^-9^), depression (*β* = 0.046, *P* = 7.48 × 10^-5^), and fluid intelligence (*β* = 0.091, *P* = 5.08 × 10^-15^).

Next, we performed joint prediction with genetic PRS (Methods). In comparison to genetic PRS, CMR traits may be more accurate and provide additional insights when predicting heart diseases (Fig. 6B). For example, the correlation between genetic PRS and atrial fibrillation was 0.084 (*P* = 1.14 × 10^-7^), suggesting that about 0.7% disease variation can be predicted by genetic profile. The prediction correlation of CMR traits was 0.265 (*P* = 2.78 × 10^-65^), which was comparable to the performance when using both CMR traits and genetic PRS (*β* = 0.276, *P* = 1.20 × 10^-70^). These results illustrate that CMR traits had much higher prediction accuracy and suggest that most of the genetic prediction power on atrial fibrillation might be medicated through cardiac conditions captured by CMR traits.

We also found that combining genetic PRS, CMR traits, and brain MRI traits can improve the prediction of multi-system diseases, such as diabetes (Fig. 6C). The prediction correlation of diabetes was 0.084 (*P* = 1.11 × 10^-7^) for genetic PRS and 0.077 (*P* = 1.13 × 10^-6^) for CMR traits. Multiple categories of brain MRI traits also had significant prediction power on diabetes, including DTI parameters (*β* = 0.094, *P* = 3.01 × 10^-9^), regional brain volumes (*β* = 0.076, *P* = 1.47 × 10^-6^), resting fMRI (*β* = 0.055, *P* = 4.85 × 10^-4^), and task fMRI (*β* = 0.045, *P* = 4.46 × 10^-3^). The prediction performance was improved to 0.10 (*P* = 3.84 × 10^-10^) by using all CMR and brain MRI traits and further moved up to 0.125 (*P* = 2.17 × 10^-15^) when adding genetic PRS. In summary, imaging traits could make a unique contribution to the prediction of complex traits and diseases. Multi-organ imaging and genetic PRS can be integrated to improve risk prediction and patient care.

## DISCUSSION

The intertwined connections between heart and brain health are gaining increasing attention. This study quantified the heart-brain associations using CMR and brain MRI data from over 40,000 individuals in one homogeneous study cohort (the UK Biobank). Based on this unique dataset, we identified phenotypic heart-brain connections and discovered patterns of enrichment in specific brain regions and functional networks. We further examined the effects of cardiovascular risk factors on the brain mediated by cardiac conditions. GWAS identified 80 genomic loci for CMR traits, many of which were colocalized with neuropsychiatric and neurological disorders, mental health factors, and brain MRI traits. Integrating traditional genetic PRS with imaging traits can also improve the risk prediction of human diseases. In summary, multiple lines of evidence point to the close phenotypic and genetic relationship between heart health and brain health.

Using multi-organ imaging data as endophenotypes, we identified genetic variations that can affect both heart and brain functions. Interpreting the genetic pleiotropy and understanding how human organs interact in a directional and even bidirectional manner is challenging^11^. We have adjusted for height, weight, and body mass index in our analysis to avoid confounding effects of body size. However, unobserved biological interactions and environmental factors may also confound heart-brain connections. Pleiotropy analysis across organ systems is a relatively new concept, so future research using additional data resources (for example, long-term longitudinal data and large-scale genomics data from multiple organs) may better reveal the shared biology between the brain and heart.

In this study, imaging data were mainly from European ancestry. Comparing UKB GWAS results with those of BBJ, both similarities and differences were found for genetic influences on CMR traits. For example, participants in UKB and BBJ had similar genetic effects on cardiac conditions at 22q11.23 and 8q24.13, but only the BBJ cohort had significant genetic effects at 11p15.1. It can be expected that some of the genetic components that underlie heart-brain connections may also be population-specific. More heart and brain imaging data collected from global populations may enable the development of a better picture of neuro-cardiac interactions.

This paper specifically focused on heart-brain connections. Because of the large amount of data collected in the UKB study, it is also possible to study the relationships between brain and other human organs and systems. For example, increasing evidence supports the gut-brain axis, which involves complex interactions between the central nervous system and the enteric nervous system^135^. Patients with inflammatory bowel disease (such as Crohn’s disease) show altered brain structure and function^136^, impaired cognitive functions^137^, and a higher risk of depression and anxiety^138^. Multi-system analysis using biobank-scale data may provide insights for inter-organ pathophysiological mechanisms and prevention and early detection of brain diseases.

## METHODS

Methods are available in the ***Methods*** section.

*Note: One supplementary information pdf file, one supplementary figure pdf file, and one supplementary table zip file are available*.

## Data Availability

Our GWAS summary statistics of 82 CMR traits have been shared on Zenodo and at Heart-KP https://heartkp.org/. The GWAS summary statistics of brain MRI traits can be freely downloaded at BIG-KP https://bigkp.org/.

## ACKNOWLEDGEMENTS

This research was partially supported by U.S. NIH grants MH086633 (H.Z.) and MH116527 (TF.L.). We thank the individuals represented in the UK Biobank for their participation and the research teams for their work in collecting, processing and disseminating these datasets for analysis. We would like to thank the University of North Carolina at Chapel Hill and Purdue University and their Research Computing groups for providing computational resources and support that have contributed to these research results. We gratefully acknowledge all the studies and databases that made GWAS and eQTL summary-level data publicly available. This research has been conducted using the UK Biobank resource (application number 22783), subject to a data transfer agreement.

## AUTHOR CONTRIBUTIONS

B.Z., H.Z., J.S., and Y.L. designed the study. B.Z., TF.L., Z.F., Y.Y., X.W. TY. L, J.T., D.X., and Z.W. analyzed the data. TF.L., Y.Y., J.T., X.W., D.X., TY. L, J.C., Y.S., C.T., and Z.Z. processed heart imaging data and undertook quantity controls. B.Z. wrote the manuscript with feedback from all authors.

**CORRESPINDENCE AND REQUESTS FOR MATERIALS** should be addressed to H.Z.

## COMPETETING FINANCIAL INTERESTS

The authors declare no competing financial interests.

## METHODS

### Heart and brain imaging data

We extracted imaging traits from the raw brain and cardiac MRI images under the UKB Data-Field 100003. Specifically, following the pipelines developed in Bai, et al. ^43^, we generated 82 CMR traits from the short-axis, long-axis, and aortic cine images. Detailed pipeline implementation and quality controls steps can be found in **Supplementary Note**. The 82 CMR traits can be divided into 6 categories, including 64 left ventricle traits, 4 left atrium traits, 4 right ventricle traits, 4 right atrium traits, 3 ascending aorta traits, and 3 descending aorta traits. Among 64 left ventricle traits, we had well-established volumetric traits such as LVEDV, LVESV, LVSV, LVEF, LVCO, and LVM. In addition, there were global and regional measures for myocardial-wall thickness at end-diastole, peak circumferential strain, radial strain, as well as longitudinal strain. For right ventricle, we had RVEDV, RVESV, RVSV, and RVEF. Four traits were generated for left and right atrium, respectively, including maximum volume (LAV max and RAV max), minimum volume (LAV min and RAV min), stroke volume (LVSV and RASV), and ejection fraction (LAEF and RAEF). For ascending aorta and descending aorta, we had measures for maximum area (AAo max area and DAo max area), minimum area (AAo min area and DAo min area), and distensibility (AAo distensibility and DAo distensibility).

The procedures for deriving brain MRI traits were described in detail in previous papers from our group, including regional brain volumes from T1-weighted MRI image^38^, DTI parameters from diffusion MRI image^40^, functional activity and connectivity traits from resting-state and task-based fMRI image^42^. Briefly, we used the advanced normalization tools^139^ (ANTs) to generate 98 regional brain volumes for cortical and subcortical regions, as well as 3 global brain volume measures, including total gray matter volume, total white matter volume, and total brain volume. In addition, using a similar procedure to that of brain volumetric traits, we used ANTs to extract 63 global and regional cortical thickness traits in this study (**Supplementary Note**). Using the ENIGMA-DTI pipeline^140,141^, we generated 110 tract-averaged parameters for fractional anisotropy, mean diffusivity, axial diffusivity, radial diffusivity, and mode of anisotropy in 21 major white matter tracts and across the whole brain (5 × 22). For fMRI, we used a parcellation-based approach based on the Glasser360 atlas^142^, which divided the cerebral cortex into 360 regions in 12 functional networks^143^. We mainly considered the mean amplitude (that is, functional activity) of each network, the mean functional connectivity for each pair of networks, and the mean amplitude and mean functional connectivity of the whole cortex (92 traits for resting and task fMRI, respectively). To further provide details on functional organizations of cerebral cortex, we also considered 64,620 area-level high-resolution resting functional connectivity in phenotypic analysis with CMR traits.

The UKB study recruited approximately half a million participants aged between 40 and 69 years between 2006 and 2010. The UKB study had obtained ethics approval from the North West Multicentre Research Ethics Committee (approval number: 11/NW/0382). In 2014, UKB started a project to re-invite 100,000 participants to undergo a multi-modal imaging study, including both brain and cardiac MRI^144^. We used the white British individuals in UKB phases 1 to 3 data (released up through 2020, *n* = 31,875 for CMR traits) as main discovery sample in our analysis. The UKB white but non-British subjects in phases 1 to 3 data and the white individuals in newly released UKB phase 4 data (*n* = 8,252, removed relatives of the discovery sample) were treated as European validation sample. In addition, we considered two non-European UKB validation datasets, including UKB Asian (UKBA, *n* = 500) and UKB Black (UKBBL, *n* = 271). We also used the UKB first revisit data (*n* = 2,903) to evaluate the reproducibility of CMR traits. The age range of imaging subjects was 45 to 82 (mean age = 64.16, standard error = 7.67) and the proportion of female was 51.6%.

### Phenotypic and mediation analysis

We performed pairwise linear regression for each pair of CMR and brain MRI traits. For all pairs, we adjusted for the effects of age (at imaging), age-squared, sex, age-sex interaction, age-squared-sex interaction, imaging site, height, weight, body mass index, the top 40 genetic PCs, and total brain volume (for traits other than total brain volume itself). For each continuous variable, the values greater than five times the median absolute deviation from the median were removed. The number of subjects that had both CMR and brain MRI data (after all quality controls) was 31,875 for regional brain volumes and cortical thickness, 30,212 for DTI parameters, 30,792 for resting fMRI traits, and 26,849 for task fMRI traits. For regional cortical thickness, we additionally adjusted for global mean thickness (for traits other than global mean thickness itself). For resting and task fMRI traits, we additional adjusted for head motion, head position, and volumetric scaling variables. We reported the *P* values from two-sided t test and adjusted for multiple testing using Bonferroni correction (considering all pairs of CMR and brain MRI traits).

Mediation analysis was performed for 41 traits, including diastolic blood pressure, systolic blood pressure, high blood pressure, diabetes mellitus, smoking, drinking, basal metabolic rate, and 34 biomarkers. We used the 82 CMR traits as cardiac mediators and performed the analysis separately for all brain MRI traits, including 101 regional brain volumes, 63 cortical thickness, 110 DTI parameters, 92 resting fMRI traits, and 92 task fMRI traits. For each brain MRI trait *y* (such as total brain volume), we fitted two models: *y* = *xβ*_1_ + *zη*_1_ + *w*α + 𝜖_1_ (Model 1) and *y* = *xβ*_2_ + *zη*_2_ + 𝜖_2_ (Model 2), where *x* is the trait of interest (such as diastolic blood pressure), *z* is the set of covariates to be adjusted, *w* is the set of 82 CMR traits (cardiac mediators), β_1_ is the conditional effect of *x* on *y*, β_2_ is the marginal effect of *x* on *y*, η_1_ and η_2_ are effects of covariates, α is the effect of *w* on *y*, and 𝜖_1_ and 𝜖_2_ are random errors. We adjusted the same set of covariates as we used in the above phenotypic analysis. A mediation relationship was built if all the three conditions were satisfied: 1) β_2_ was significant after Bonferroni correction, suggesting the significant effects of *x* on *y* ; 2) α was significant after Bonferroni correction, suggesting significant relationship between *w* and *y*; 3) |β_1_| < |β_2_| and the two coefficients had the directions, suggesting the reduced effects of *x* after adjusting for heart mediators. The proportion of medicated effect was defined by (|β_1_|−|β_2_|)/|β_1_|. A mediation relationship suggests a mechanism that the trait *x* may have implicit influence on the brain MRI trait through the intermediate CMR traits.

### Genetic analysis on 82 CMR traits

We used the UKB imputed genotyping data and performed the following quality controls on the subset of subjects with both CMR traits and genetics data^42^: 1) excluded subjects with more than 10% missing genotypes; 2) excluded variants with minor allele frequency less than 0.01; 3) excluded variants with missing genotype rate larger than 10%; 4) excluded variants that failed the Hardy-Weinberg test at 1 × 10^-7^ level; and 5) removed variants with imputation INFO score less than 0.8. We estimated the SNP heritability using GCTA^59^ with autosomal SNPs in the white British discovery dataset. The effects of age (at imaging), age-squared, sex, age-sex interaction, age-squared-sex interaction, imaging site, height, weight, body mass index, and the top 40 genetic PCs were adjusted. We performed GWAS using linear mixed effect models implemented in fastGWA^145^, with the same set of covariates as in heritability analysis being adjusted. We used Plink^146^ (v2.0) for validation GWAS, in which we adjusted for top ten genetic PCs instead of top 40. The significant (*P* < 6.09 × 10^-10^, that is, 5 × 10^-8^/82) genomic loci were defined using FUMA^128^ (version v1.3.6a). Briefly, to define the LD boundaries, FUMA first identified independent significant variants, which were defined as variants whose *P* values were smaller than 6.09 × 10^-10^ and were independent of other significant variants (LD *r*^2^ < 0.6). Next, FUMA constructed LD blocks by tagging all variants (MAF ≥ 0.0005, including variants from the 1000 Genomes reference panel) in LD (*r*^2^ ≥ 0.6) with at least one independent significant variant. Within these independent significant variants, FUMA defined independent lead variants as those that were independent from each other (LD *r*^2^ < 0.1). For independent significant variants that were close with each other (<250 kb based on their LD boundaries), FUMA merged their LD blocks into one single genomic locus. Independent significant variants and all variants in LD (*r*^2^ ≥ 0.6) were searched on the NHGRI-EBI GWAS catalog (version 2019-09-24) to look for previously reported GWAS findings (*P* < 9 × 10^-6^) with any traits.

Using GWAS summary statistics, we performed genetic correlation analysis via LDSC^126^ (version 1.0.1). The LD scores were from the 1000 Genomes European data and were provided by LDSC. This analysis was focused on HapMap3^147^ variants and the variants in the major histocompatibility complex region were removed. Gene level association was tested for 18,796 protein-coding genes using MAGMA^127^ (version 1.08). Default MAGMA settings were used with zero window size around each gene. FUMA performed functional annotation and mapping analysis, in which genetic variants were annotated and linked to 35,808 candidate genes by a combination of positional, eQTL, and 3D chromatin interaction mappings. We selected heart-tissues/cells related options and default values were used for all other options in FUMA. MAGMA gene-set analysis was used to explore the implicated biological pathway using 10,678 pre-constructed gene sets. Heritability enrichment analysis was performed using partitioned LDSC^62^. We tested for tissue type and cell type specific regulatory elements across multiple tissues and cell types in the Roadmap Epigenomics Project^134^. The baseline models were adjusted when estimating and testing the enrichment scores in partitioned LDSC.

### Prediction models with genetics and multi-organ image data

We examined the prediction performance of CMR traits on 95 complex traits and diseases, most of which were ICD10 diseases (62 diseases from Chapter 3, 4, 5, 6, and 9 in Data-Field 41270), mental health and cognitive traits, family history, self-reported cardiovascular diseases, and cardiovascular risk factors. We focused on the 36,949 unrelated white British subjects and randomly divided the data into three independent parts: training (*n* = 22,169), validation (*n* = 7,390), and testing (*n* = 7,390). The effect sizes of CMR traits were estimated by ridge regression via glmnet^148^ (R version 3.6.0, *n* = 22,169). We removed the effects of age (at imaging), age-squared, sex, age-sex interaction, age-squared-sex interaction, imaging site, height, weight, body mass index, and the top 40 genetic PCs. Model tuning parameters were all estimated in the validation data and the prediction performance was examined on the testing data by calculation the correlation between the predicted values and the observed ones. Next, we examined the performance of genetic PRS for selected heart diseases. We used all UKB white British subjects except for the ones in our validation and testing data (and their relatives) as training data. The genetic PRS was developed by lassosum^64^, with using the validation dataset to tune the parameters. Finally, similar to CMR traits, we also used multiple brain MRI traits as predictors in the prediction model for diabetes. The performance was tested and compared on the subjects in the testing dataset that had all these data types.

### Code availability

We made use of publicly available software and tools. The codes used to generate CMR traits will be shared on Zenodo.

### Data availability

Our GWAS summary statistics of 82 CMR traits have been shared on Zenodo and at Heart-KP https://heartkp.org/. The GWAS summary statistics of brain MRI traits can be freely downloaded at BIG-KP https://bigkp.org/. The individual level UK Biobank data used in this study can be obtained from https://www.ukbiobank.ac.uk/.

