## Supplementary material for "Heart-brain connections: phenotypic and genetic insights from 40,000 cardiac and brain magnetic resonance images": supp_figures

**This PDF file includes:**

Supplementary Figures: Figs. S1 to S114

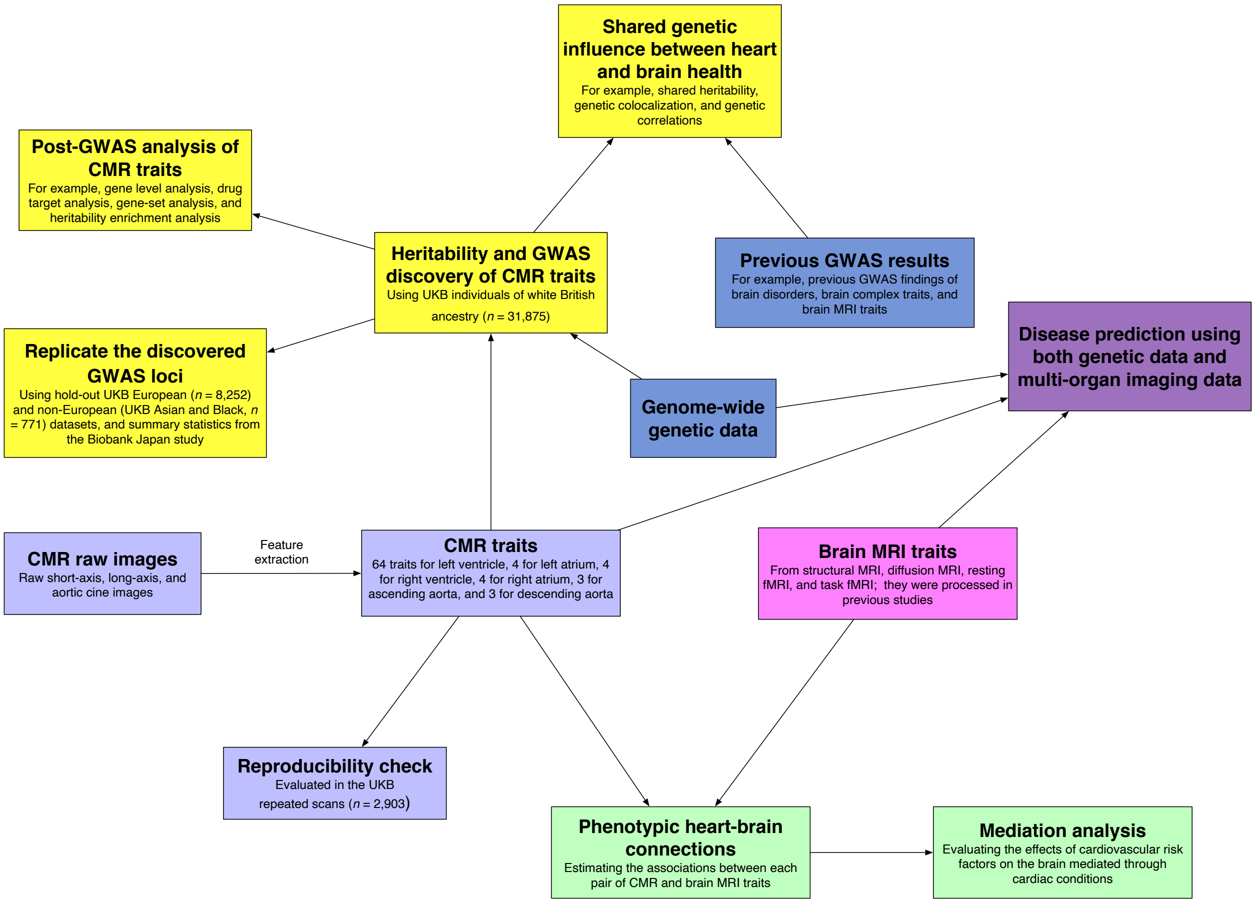


**Fig. S1 Illustration of the study design.** Our study includes the CMR traits generation and reproducibility check, phenotypic and mediation analyses, GWAS of CMR traits and evaluation of their shared genetic influences with brain-related complex traits and diseases, and joint prediction using genetic and imaging data.

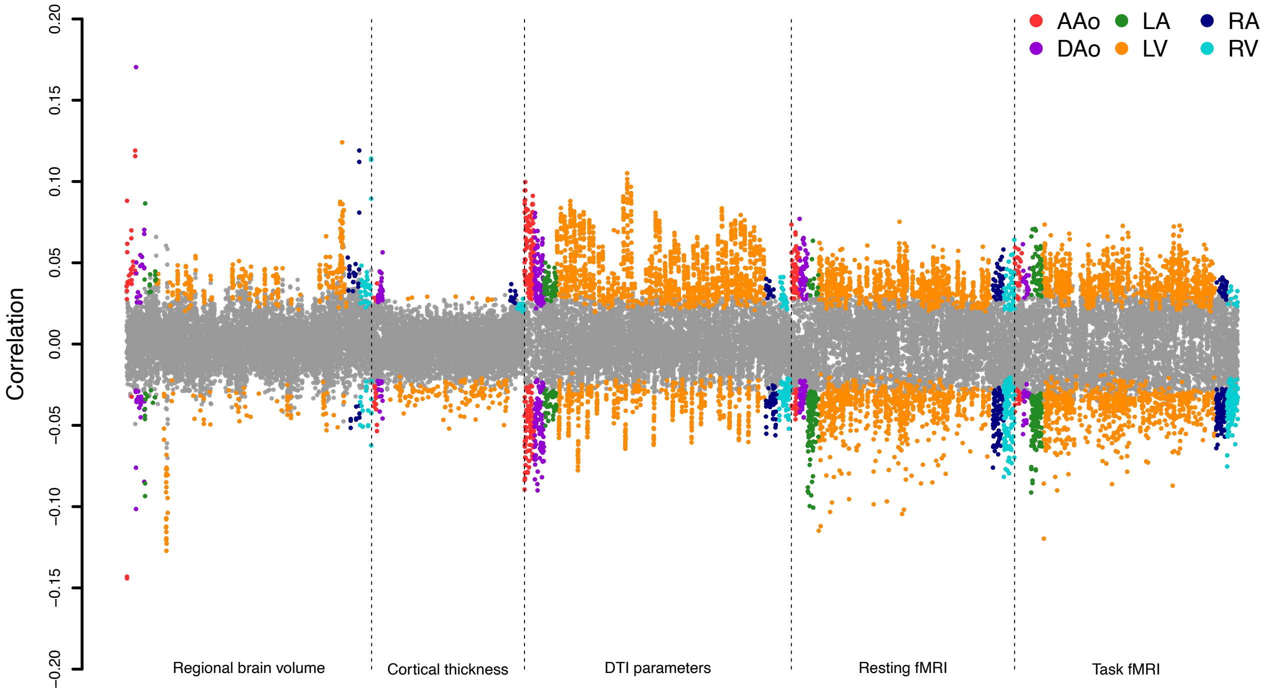


**Fig. S2 Phenotypic heart-brain associations.** The correlation coefficients between 82 CMR traits and 5 groups of brain MRI traits, including 101 regional brain volumes, 63 cortical thickness traits, 110 DTI parameters, 92 resting fMRI traits, and 92 task fMRI traits. The coefficients whose *P* values passing the Bonferroni-significance level ($P < 1.33 \times 10^{-6}$) are highlighted in colors. We label the categories of CMR traits with different colors. AAo, ascending aorta; DAo, descending aorta; LA, left atrium; LV, left ventricle; RA, right atrium; and RV, right ventricle.

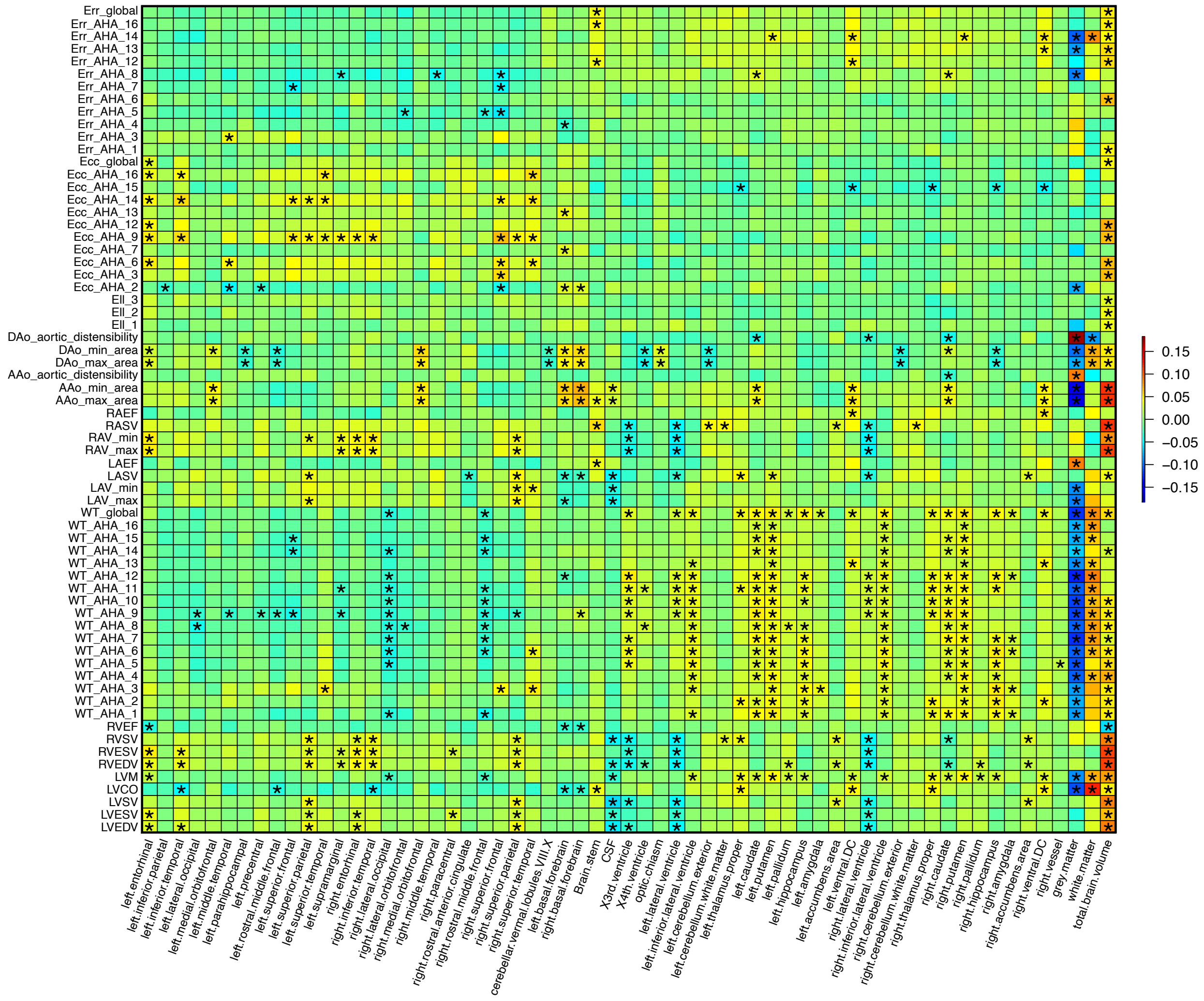


**Fig. S3 Phenotypic associations between CMR traits and regional brain volumes.** We illustrate correlations between CMR traits (*y* axis) and regional brain volumes (*x* axis). The color represents correlation estimates. The asterisks highlight significant correlations at the Bonferroni-significance level ($P < 1.33 \times 10^{-6}$).

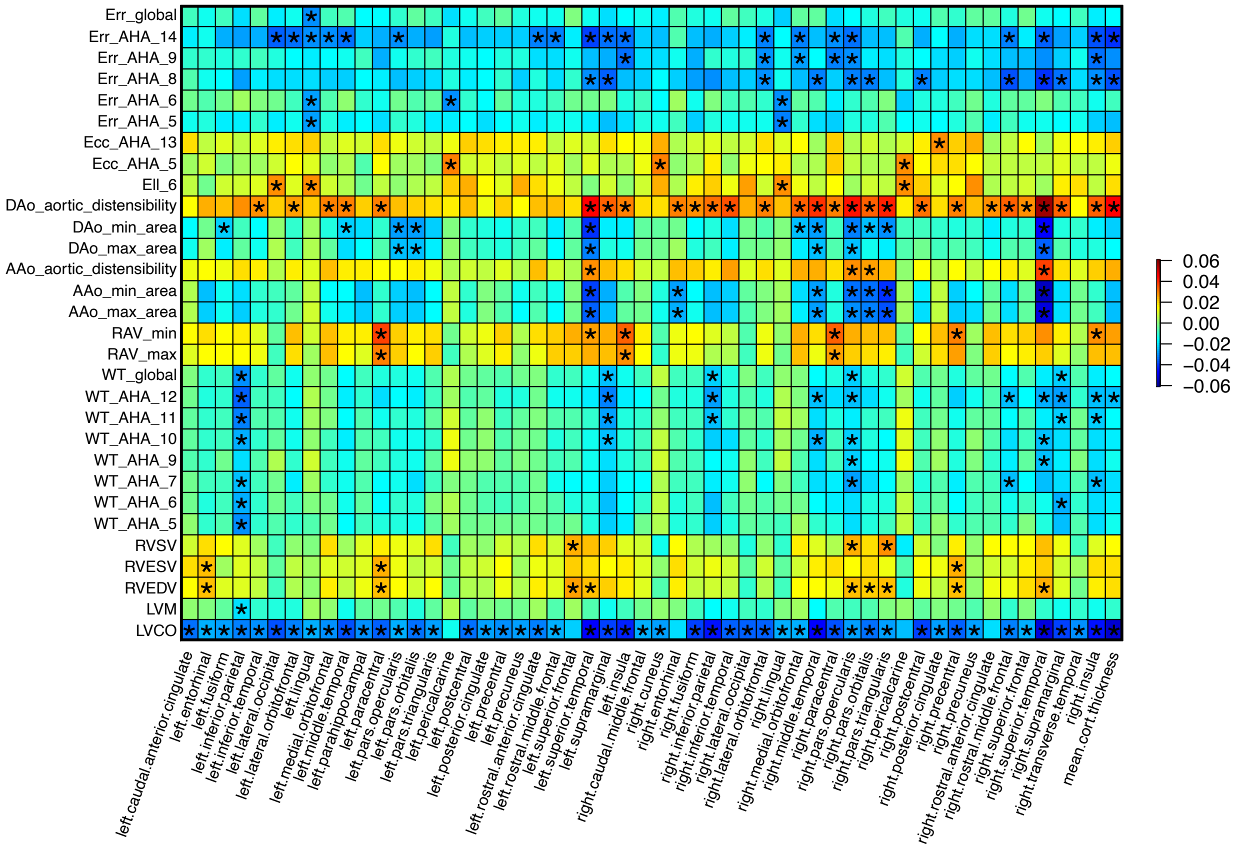


**Fig. S4 Phenotypic associations between CMR traits and cortical thickness traits.** We illustrate correlations between CMR traits (*y* axis) and cortical thickness traits (*x* axis). The color represents correlation estimates. The asterisks highlight significant correlations at the Bonferroni-significance level ($P < 1.33 \times 10^{-6}$).

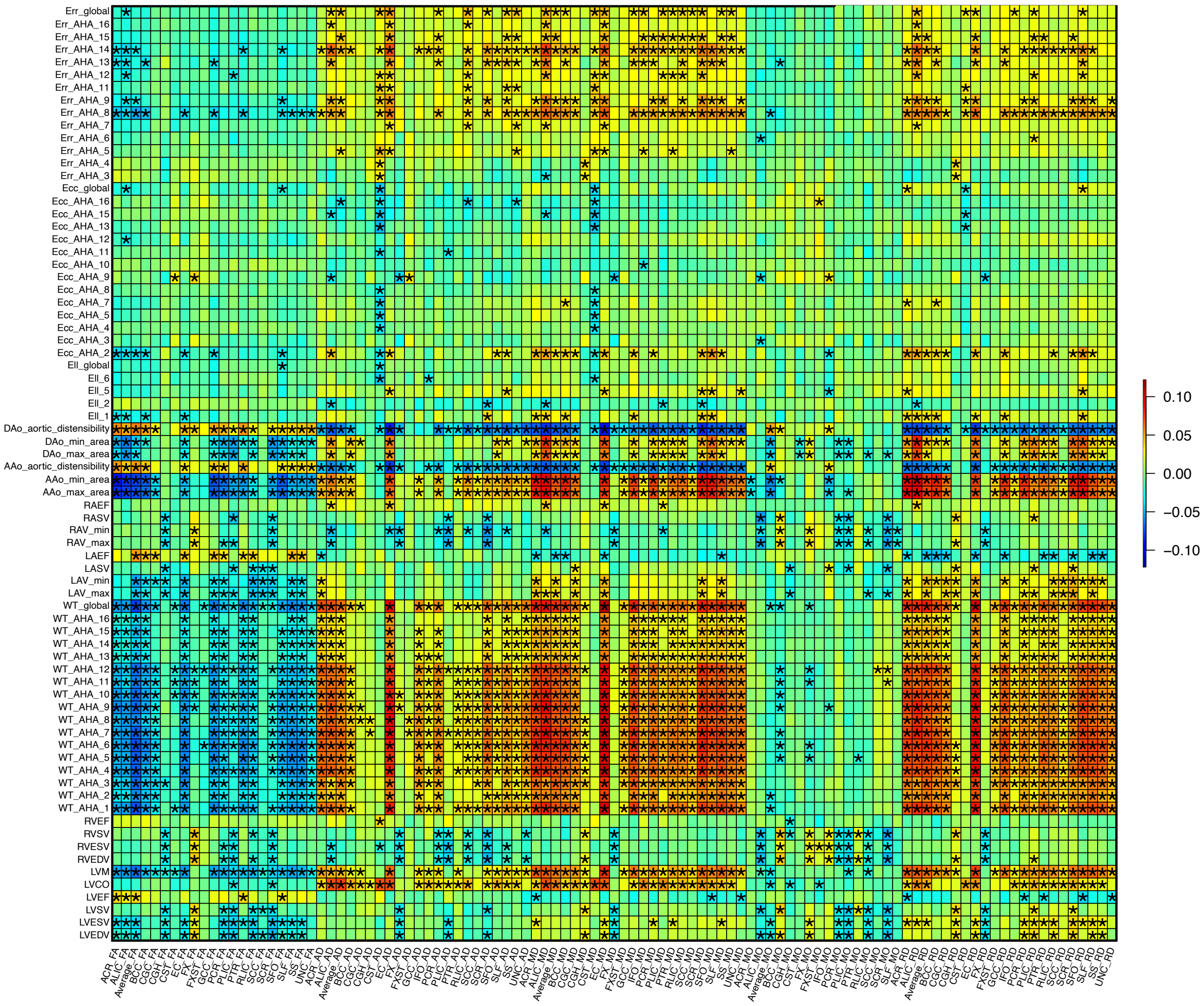


**Fig. S5 Phenotypic associations between CMR traits and DTI parameters.** We illustrate correlations between CMR traits (*y* axis) and DTI parameters (*x* axis). The color represents correlation estimates. The asterisks highlight significant correlations at the Bonferroni-significance level ($P < 1.33 \times 10^{-6}$).

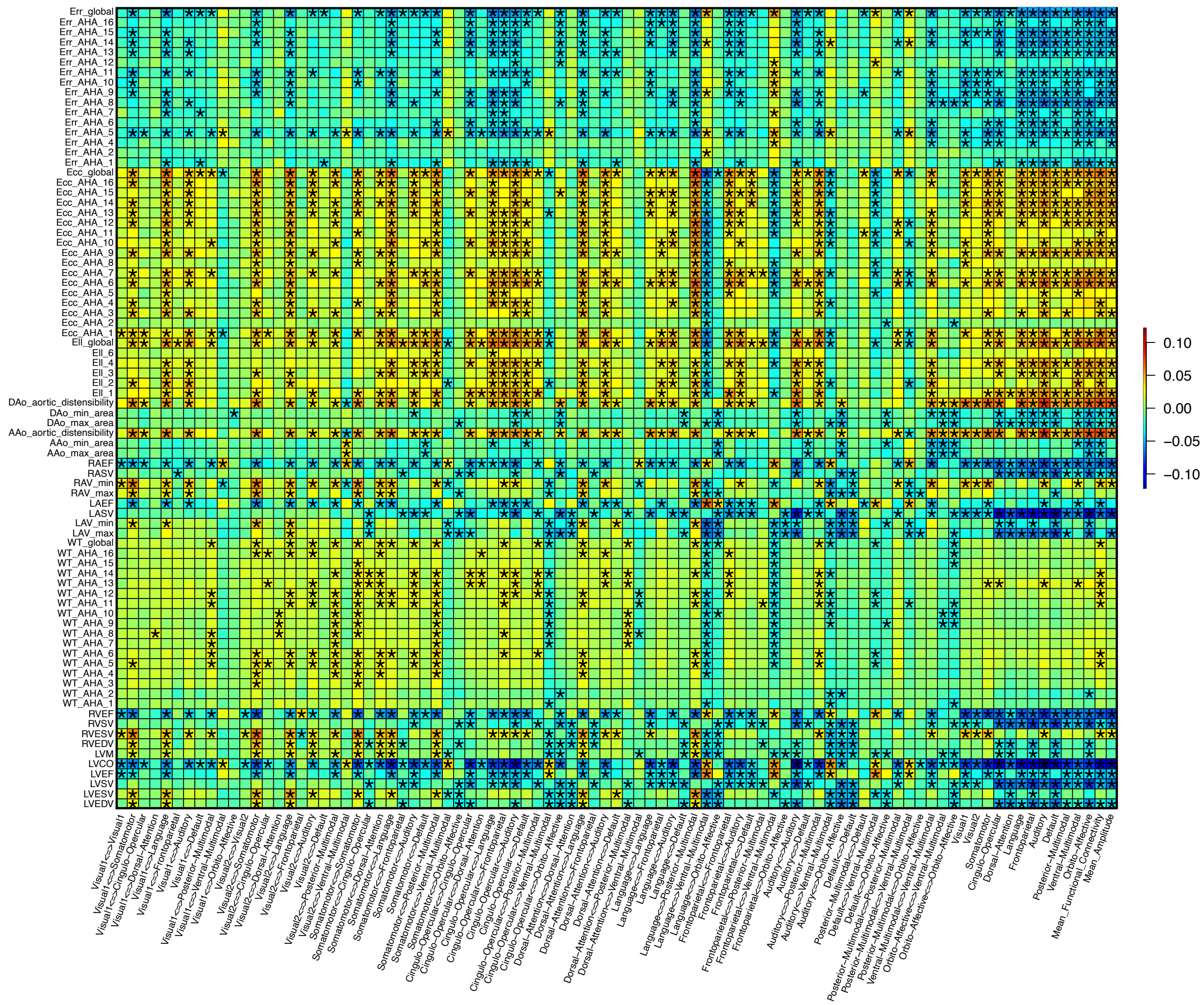


**Fig. S6 Phenotypic associations between CMR traits and resting fMRI traits.** We illustrate correlations between CMR traits (*y* axis) and resting fMRI traits (*x* axis). The color represents correlation estimates. The asterisks highlight significant correlations at the Bonferroni-significance level ($P < 1.33 \times 10^{-6}$).

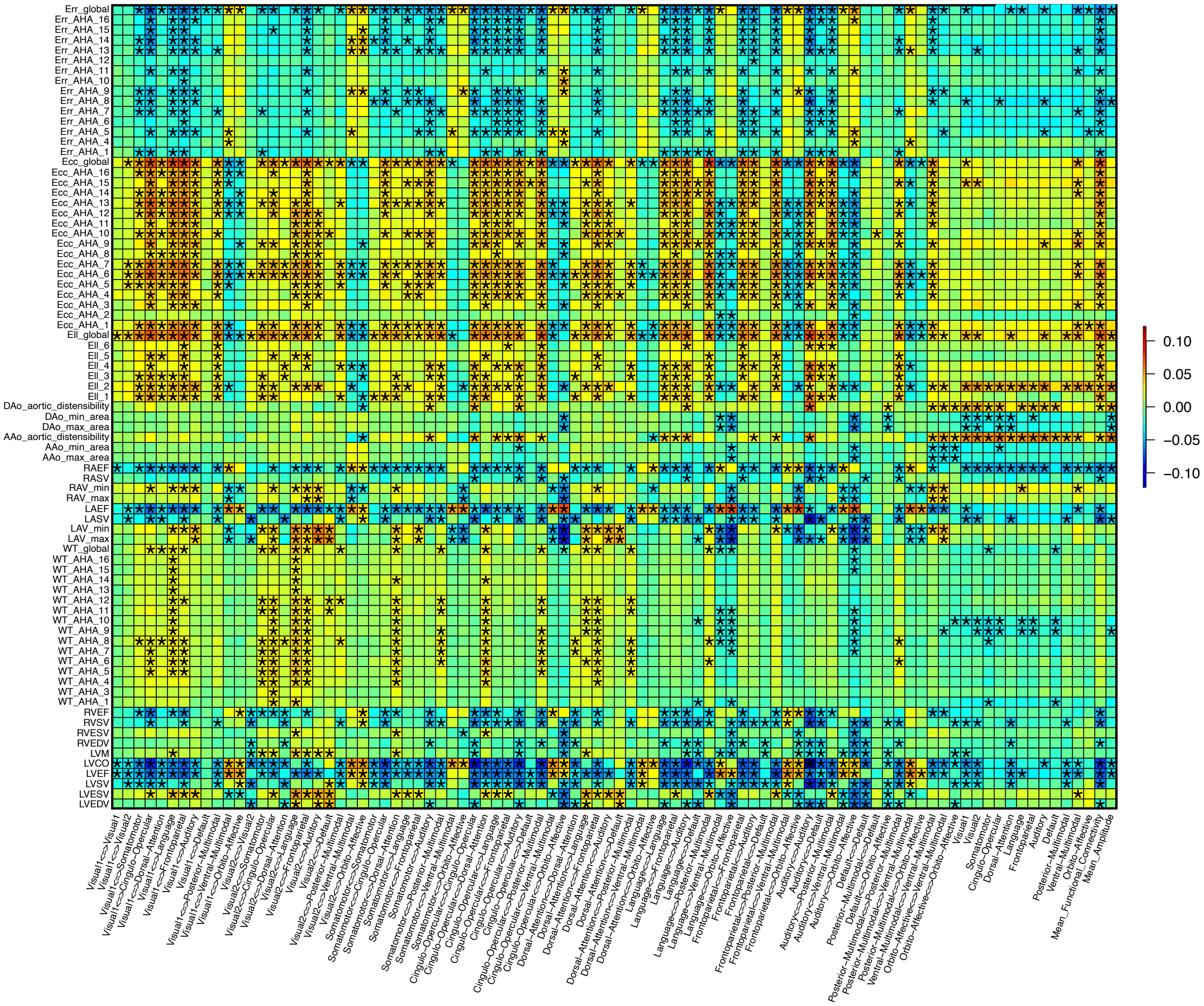


5 **Fig. S7 Phenotypic associations between CMR traits and task fMRI traits.** We illustrate correlations between CMR traits (*y* axis) and task fMRI traits (*x* axis). The color represents correlation estimates. The asterisks highlight significant correlations at the Bonferroni-significance level ($P < 1.33 \times 10^{-6}$).

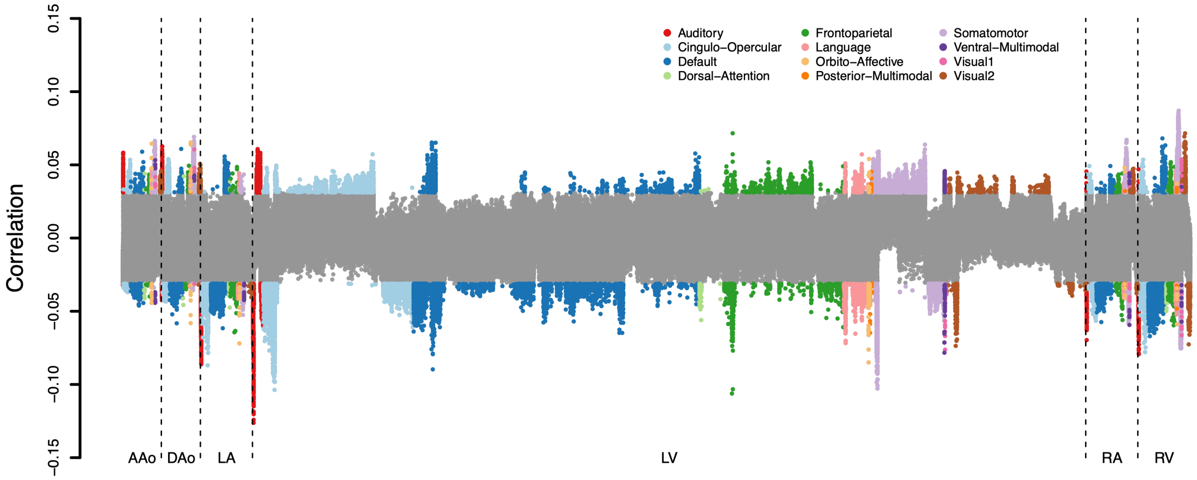


**Fig. S8 Associations with CMR traits across different functional networks in resting fMRI.** We illustrate correlation coefficients between 82 CMR traits and 8,531 within-network resting fMRI traits. The coefficients whose *P* values passing Bonferroni-significance level ($P < 7.15 \times 10^{-8}$) are highlighted in colors. We label the functional networks with different colors. Visual1, the primary visual network; Visual2, the secondary visual network.

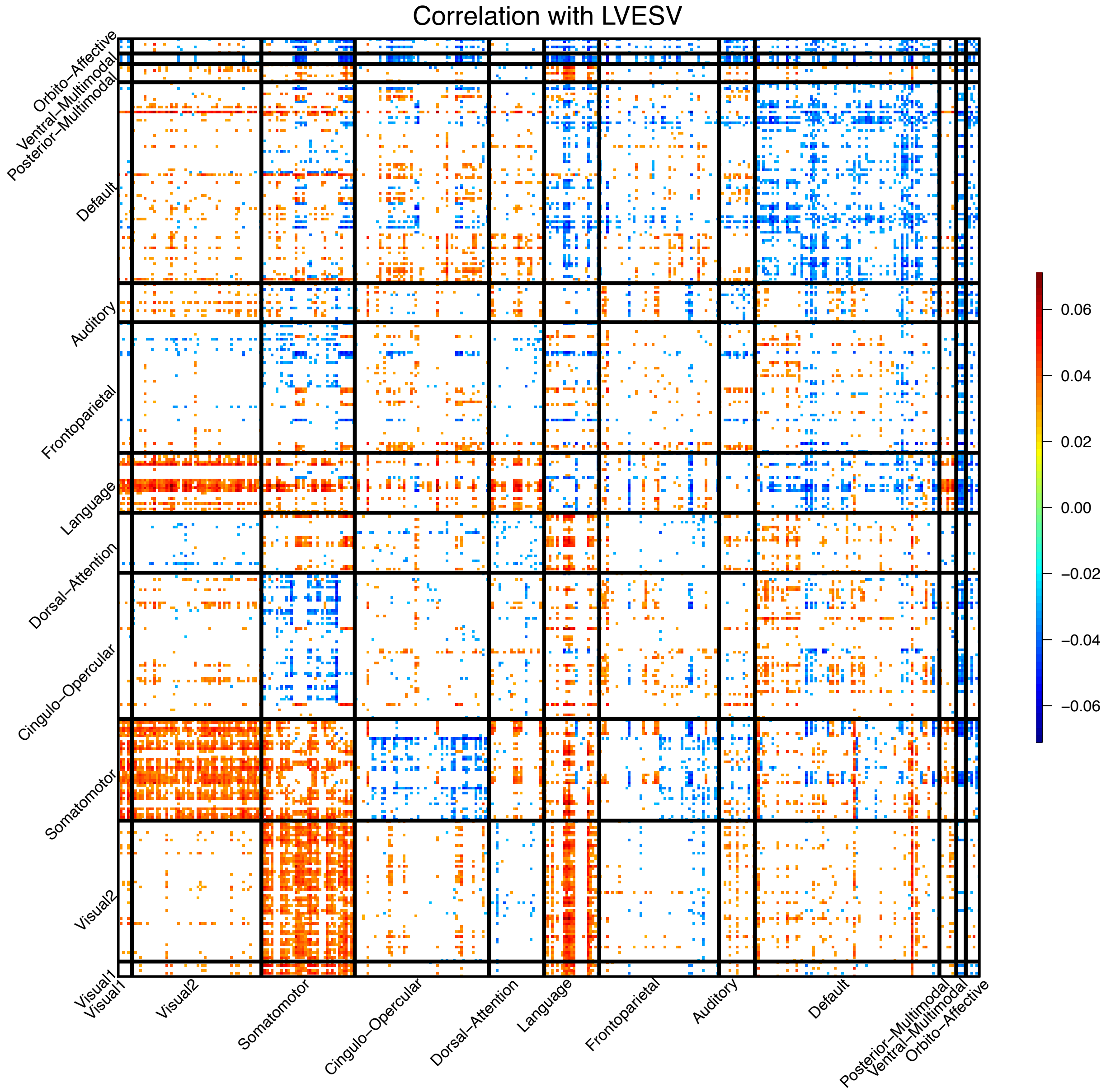


**Fig. S9 Significant associations between LVESV and resting fMRI traits across different networks.** We illustrate significant correlations (at Bonferroni significance level) between LVESV and 64,620 area level resting fMRI traits (both within-network and cross-network). The color represents correlation estimates. Visual1, the primary visual network; Visual2, the secondary visual network.

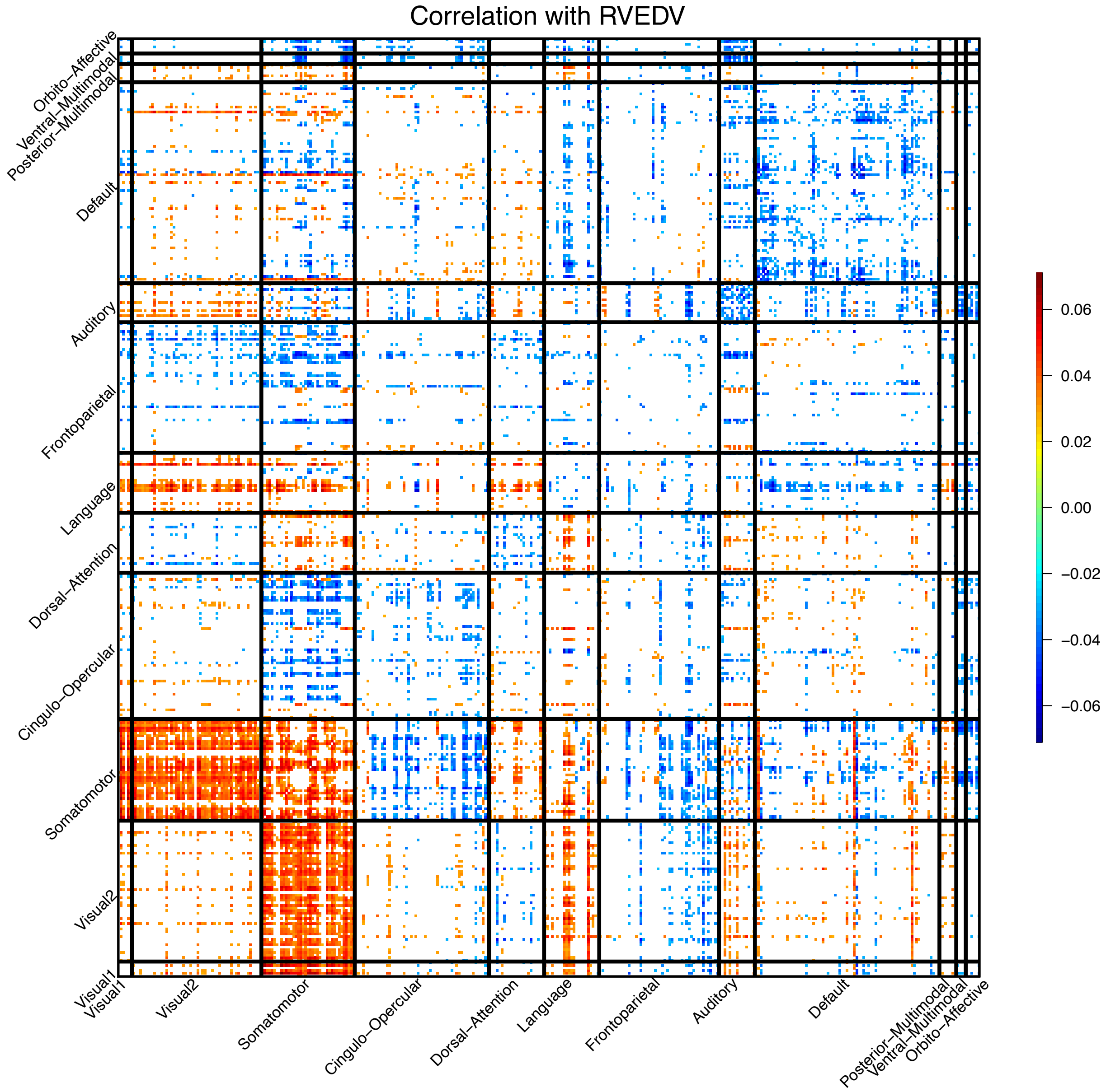


**Fig. S10 Significant associations between RVEDV and resting fMRI traits across different networks.** We illustrate significant correlations (at Bonferroni significance level) between RVEDV and 64,620 area level resting fMRI traits (both within-network and cross-network). The color represents correlation estimates. Visual1, the primary visual network; Visual2, the secondary visual network.

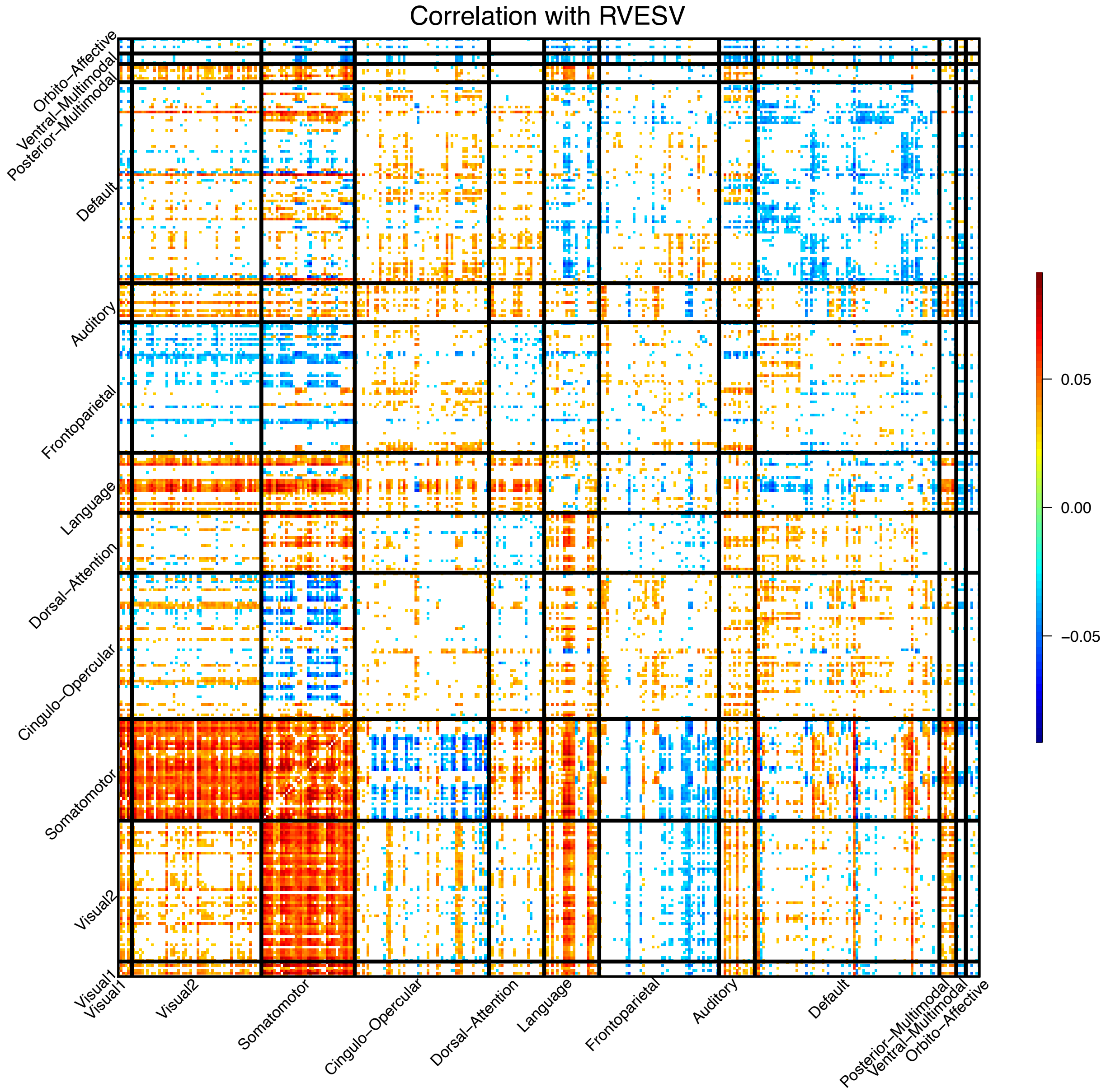


**Fig. S11 Significant associations between RVESV and resting fMRI traits across different networks.** We illustrate significant correlations (at Bonferroni significance level) between RVESV and 64,620 area level resting fMRI traits (both within-network and cross-network). The color represents correlation estimates. Visual1, the primary visual network; Visual2, the secondary visual network.

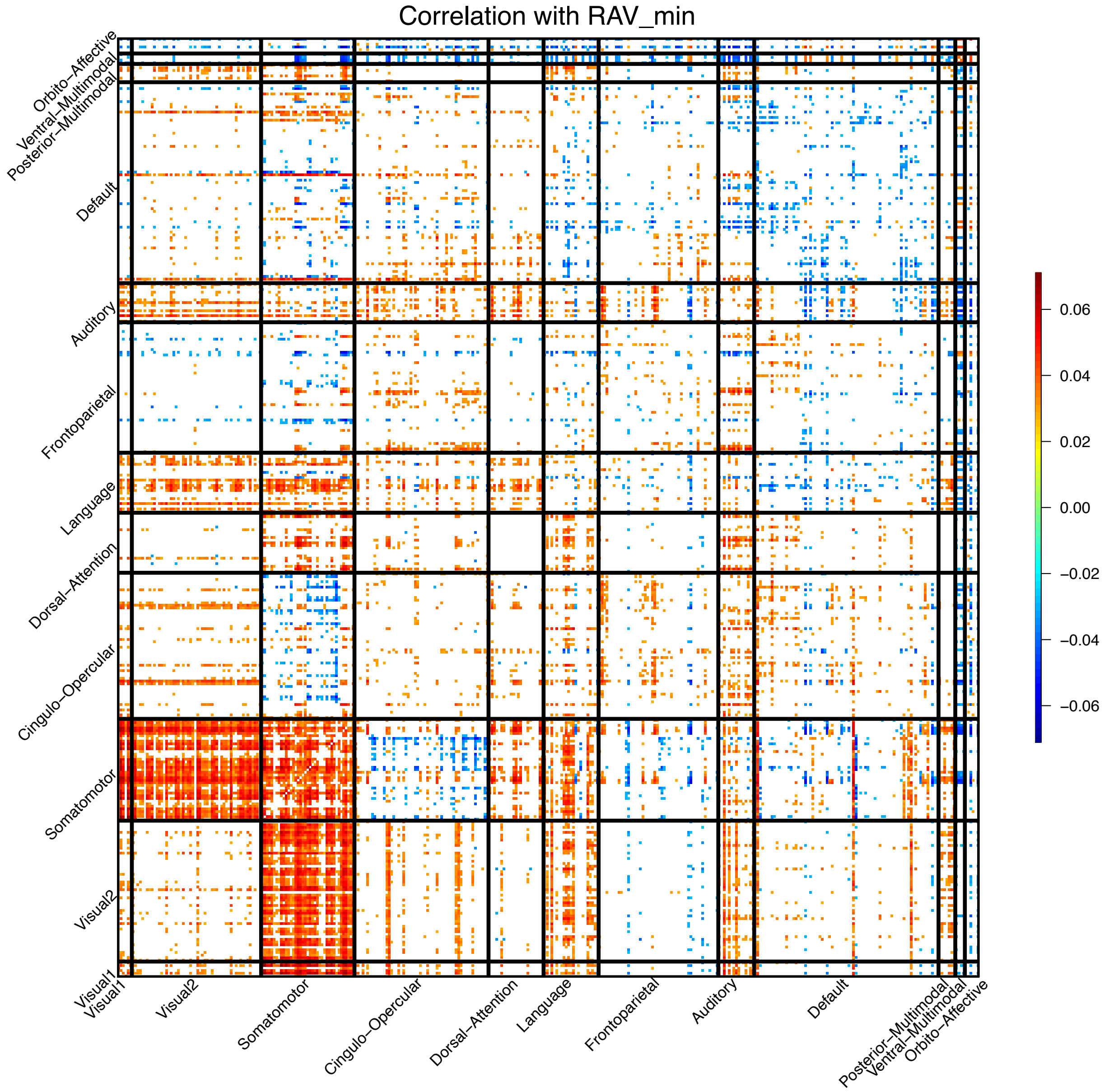


**Fig. S12 Significant associations between right atrium minimum volume (RAV min) and resting fMRI traits across different networks.** We illustrate significant correlations (at Bonferroni significance level) between RAV min and 64,620 area level resting fMRI traits (both within-network and cross-network). The color represents correlation estimates. Visual1, the primary visual network; Visual2, the secondary visual network.

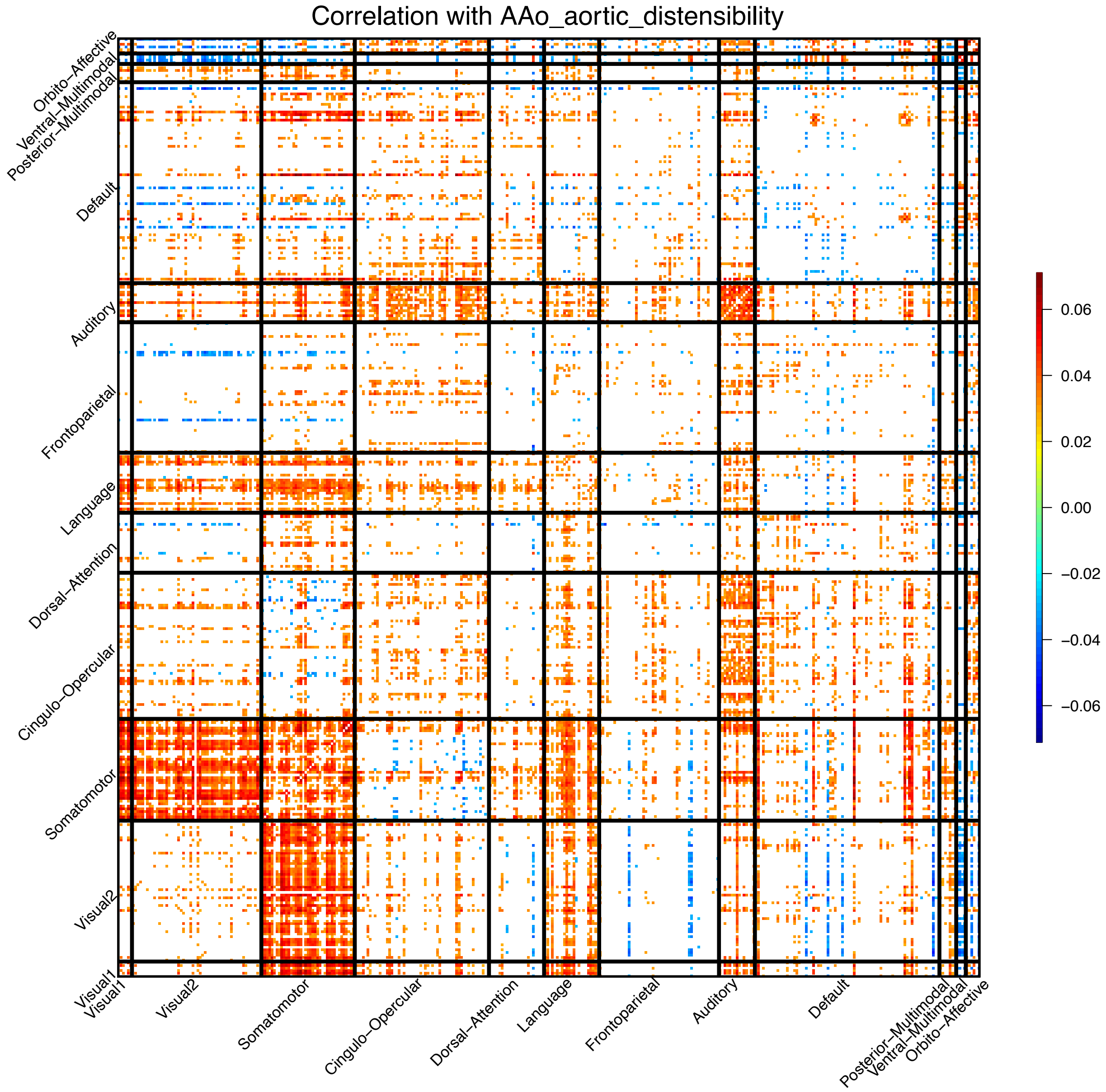


**Fig. S13 Significant associations between ascending aorta distensibility (AAo aortic distensibility) and resting fMRI traits across different networks.** We illustrate significant correlations (at Bonferroni significance level) between AAo aortic distensibility and 64,620 area level resting fMRI traits (both within-network and cross-network). The color represents correlation estimates. Visual1, the primary visual network; Visual2, the secondary visual network.

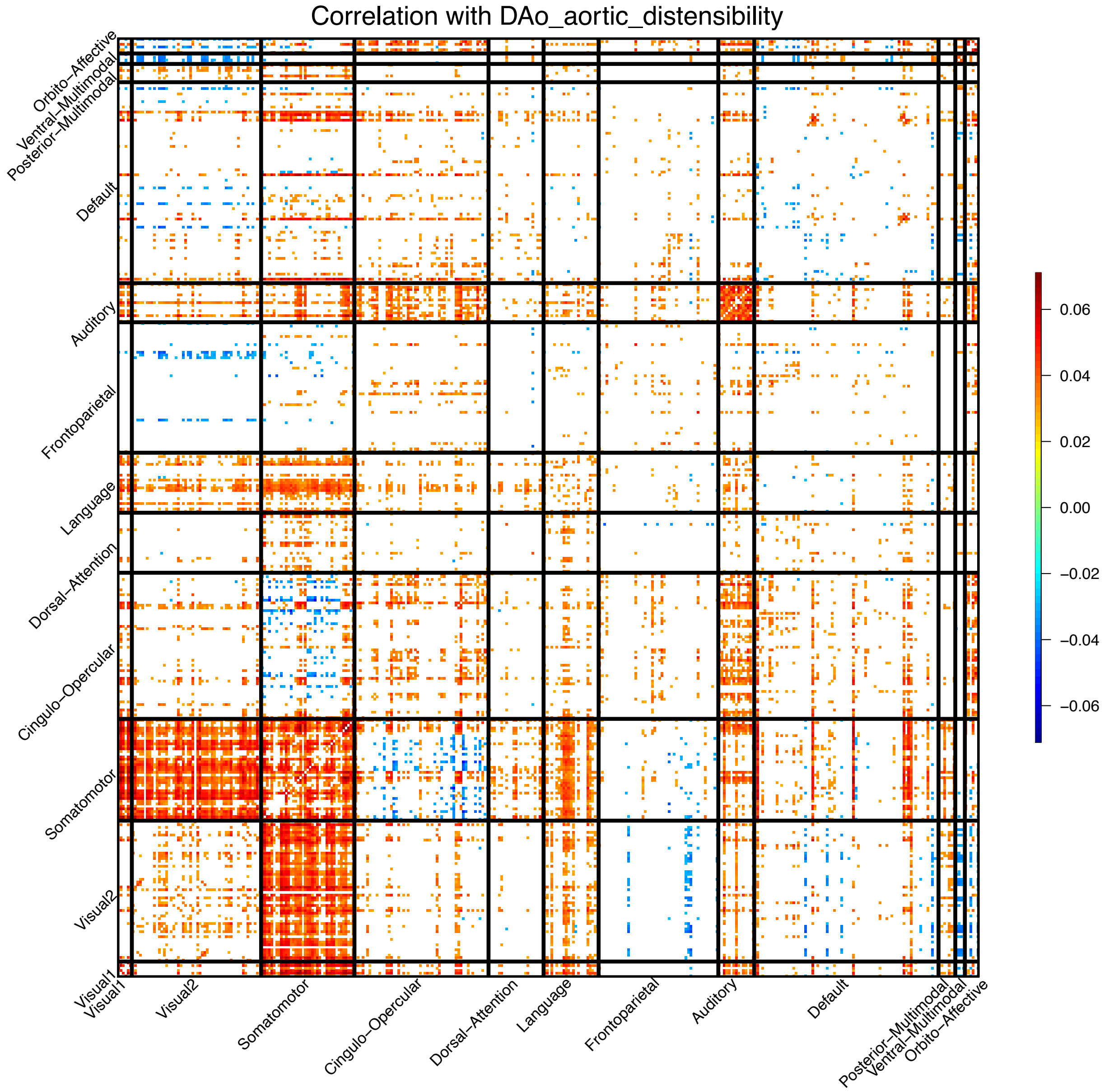


**Fig. S14 Significant associations between descending aorta distensibility (DAo aortic distensibility) and resting fMRI traits across different networks.** We illustrate significant correlations (at Bonferroni significance level) between DAo aortic distensibility and 64,620 area level resting fMRI traits (both within-network and cross-network). The color represents correlation estimates. Visual1, the primary visual network; Visual2, the secondary visual network.

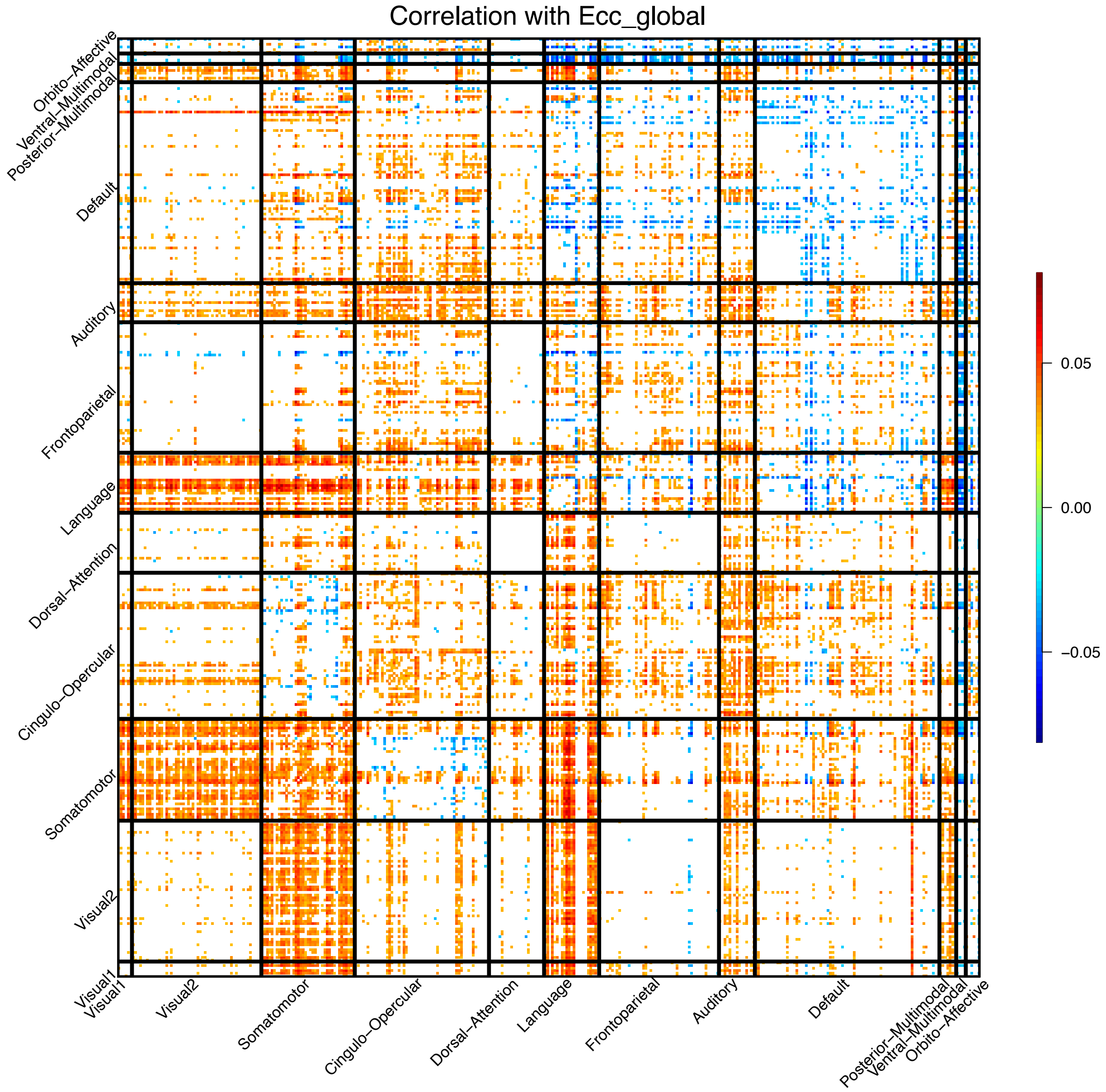


**Fig. S15 Significant associations between global peak circumferential strain (Ecc global) and resting fMRI traits across different networks.** We illustrate significant correlations (at Bonferroni significance level) between Ecc global and 64,620 area level resting fMRI traits (both within-network and cross-network). The color represents correlation estimates. Visual1, the primary visual network; Visual2, the secondary visual network.

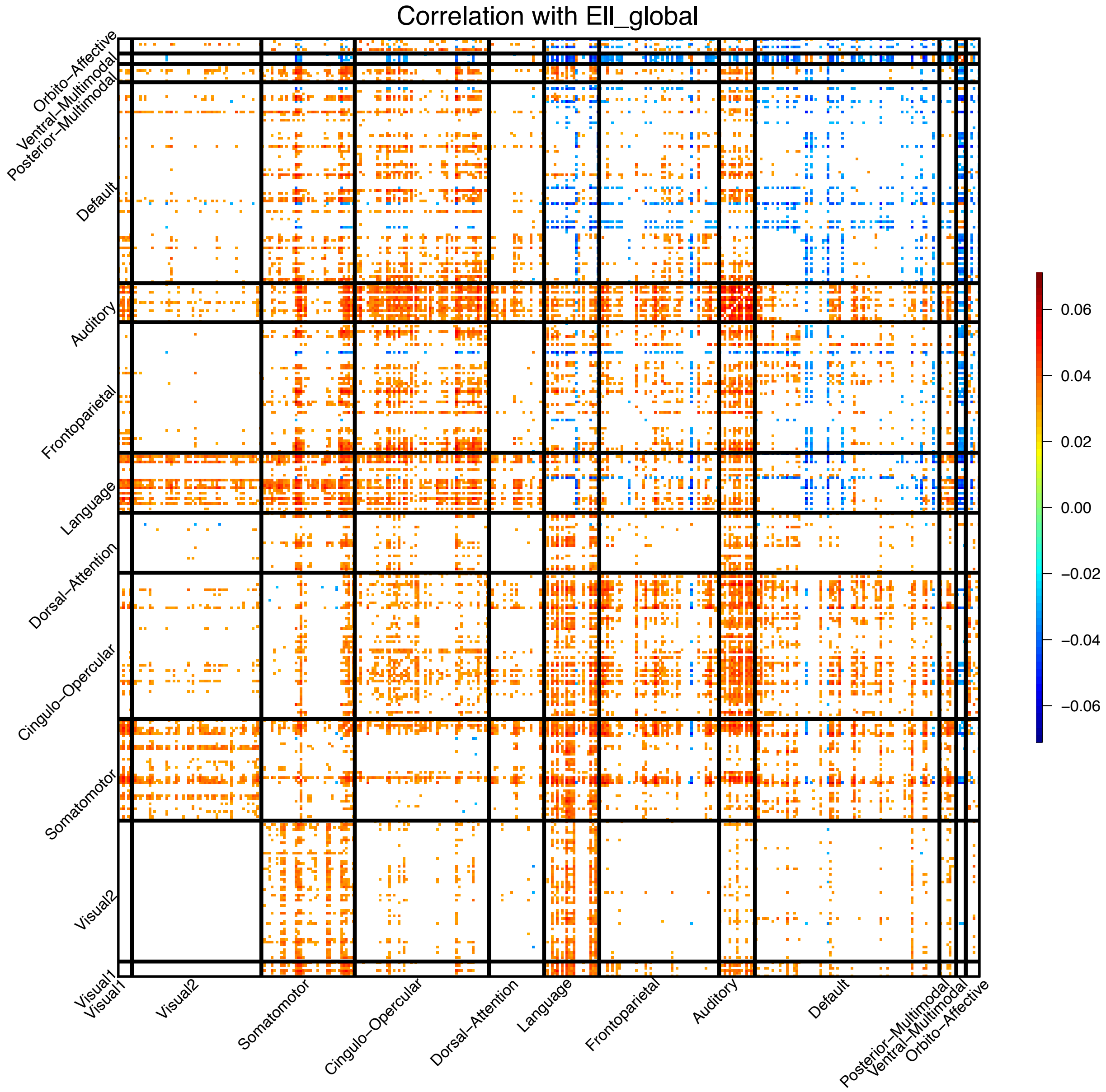


**Fig. S16 Significant associations between global longitudinal peak strain (Ell global) and resting fMRI traits across different networks.** We illustrate significant correlations (at Bonferroni significance level) between Ell global and 64,620 area level resting fMRI traits (both within-network and cross-network). The color represents correlation estimates. Visual1, the primary visual network; Visual2, the secondary visual network.

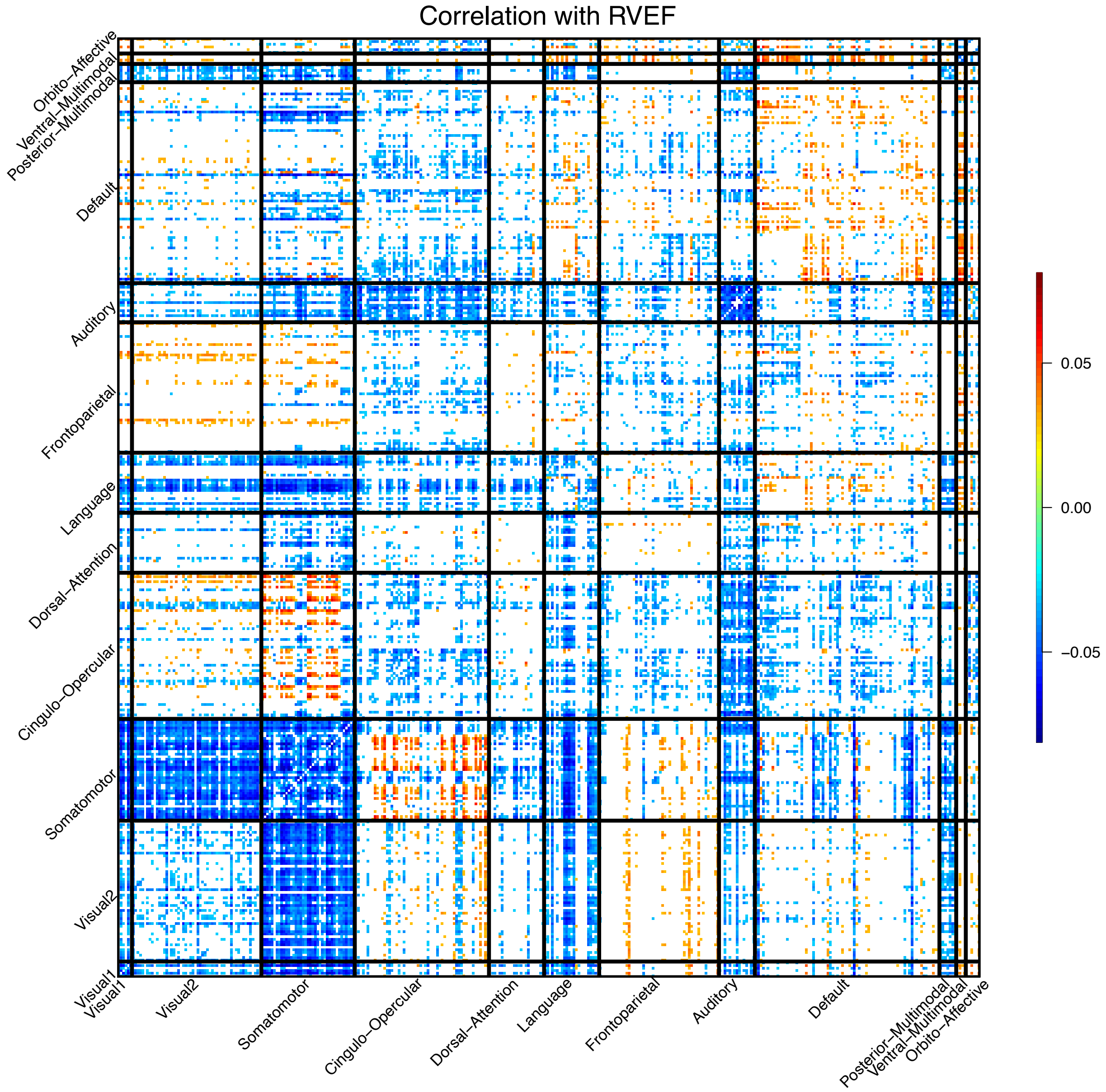


**Fig. S17 Significant associations between right ventricular ejection fraction (RVEF) and resting fMRI traits across different networks.** We illustrate significant correlations (at Bonferroni significance level) between RVEF and 64,620 area level resting fMRI traits (both within-network and cross-network). The color represents correlation estimates. Visual1, the primary visual network; Visual2, the secondary visual network.

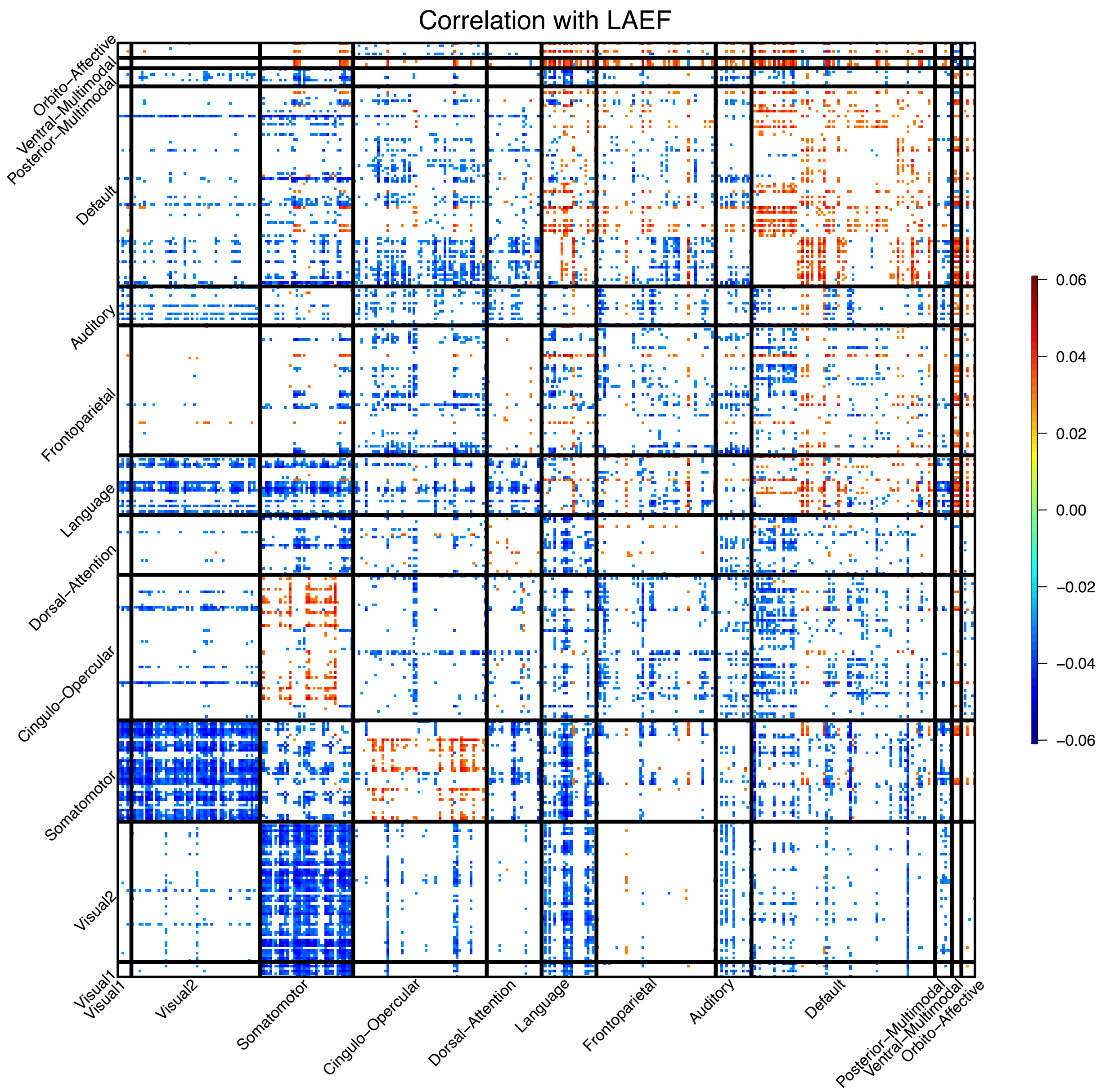


**Fig. S18 Significant associations between left atrium ejection fraction (LAEF) and resting fMRI traits across different networks.** We illustrate significant correlations (at Bonferroni significance level) between LAEF and 64,620 area level resting fMRI traits (both within-network and cross-network). The color represents correlation estimates. Visual1, the primary visual network; Visual2, the secondary visual network.

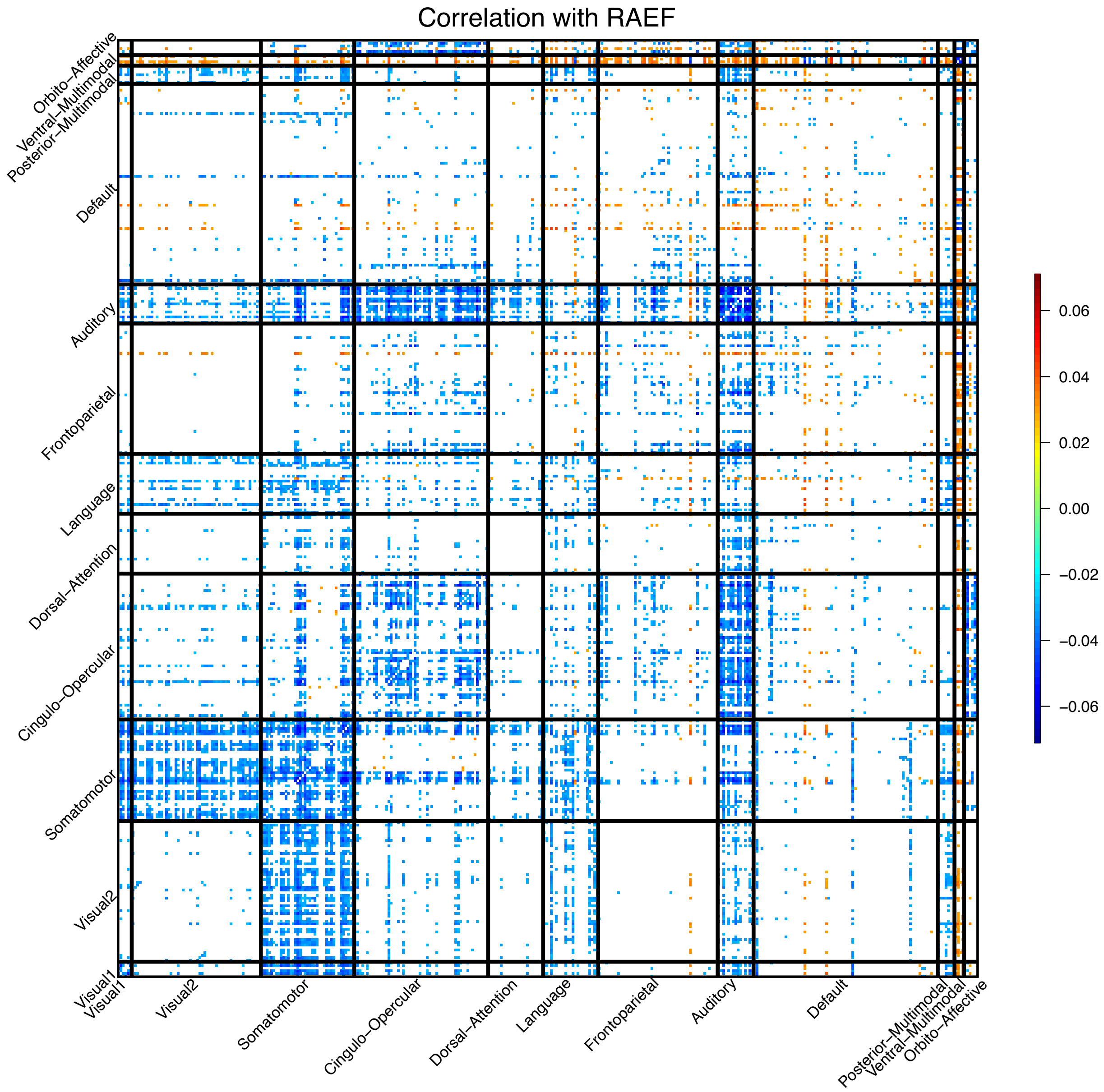


**Fig. S19 Significant associations between right atrium ejection fraction (RAEF) and resting fMRI traits across different networks.** We illustrate significant correlations (at Bonferroni significance level) between RAEF and 64,620 area level resting fMRI traits (both within-network and cross-network). The color represents correlation estimates. Visual1, the primary visual network; Visual2, the secondary visual network.

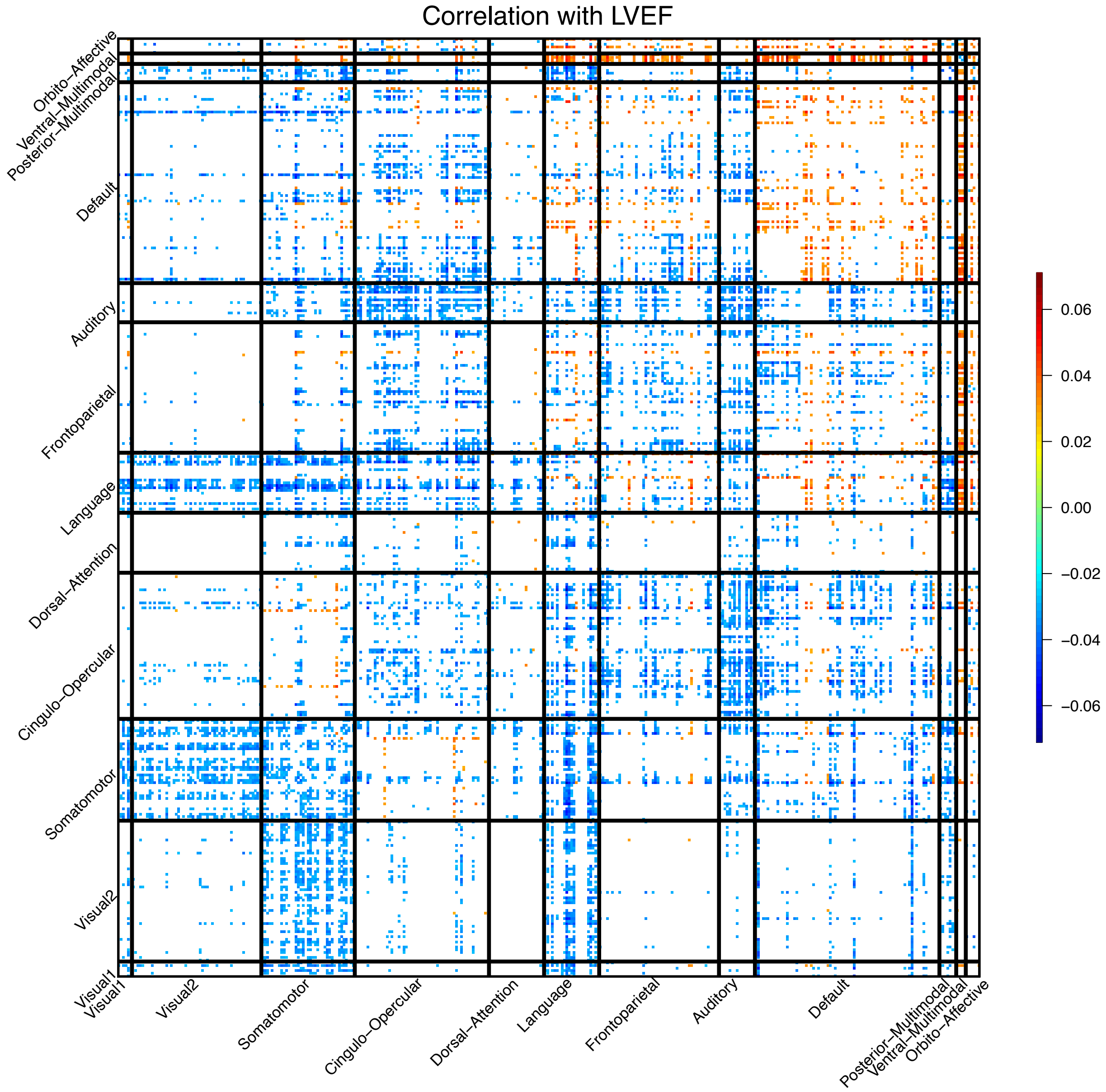


**Fig. S20 Significant associations between left ventricular ejection fraction (LVEF) and resting fMRI traits across different networks.** We illustrate significant correlations (at Bonferroni significance level) between LVEF and 64,620 area level resting fMRI traits (both within-network and cross-network). The color represents correlation estimates. Visual1, the primary visual network; Visual2, the secondary visual network.

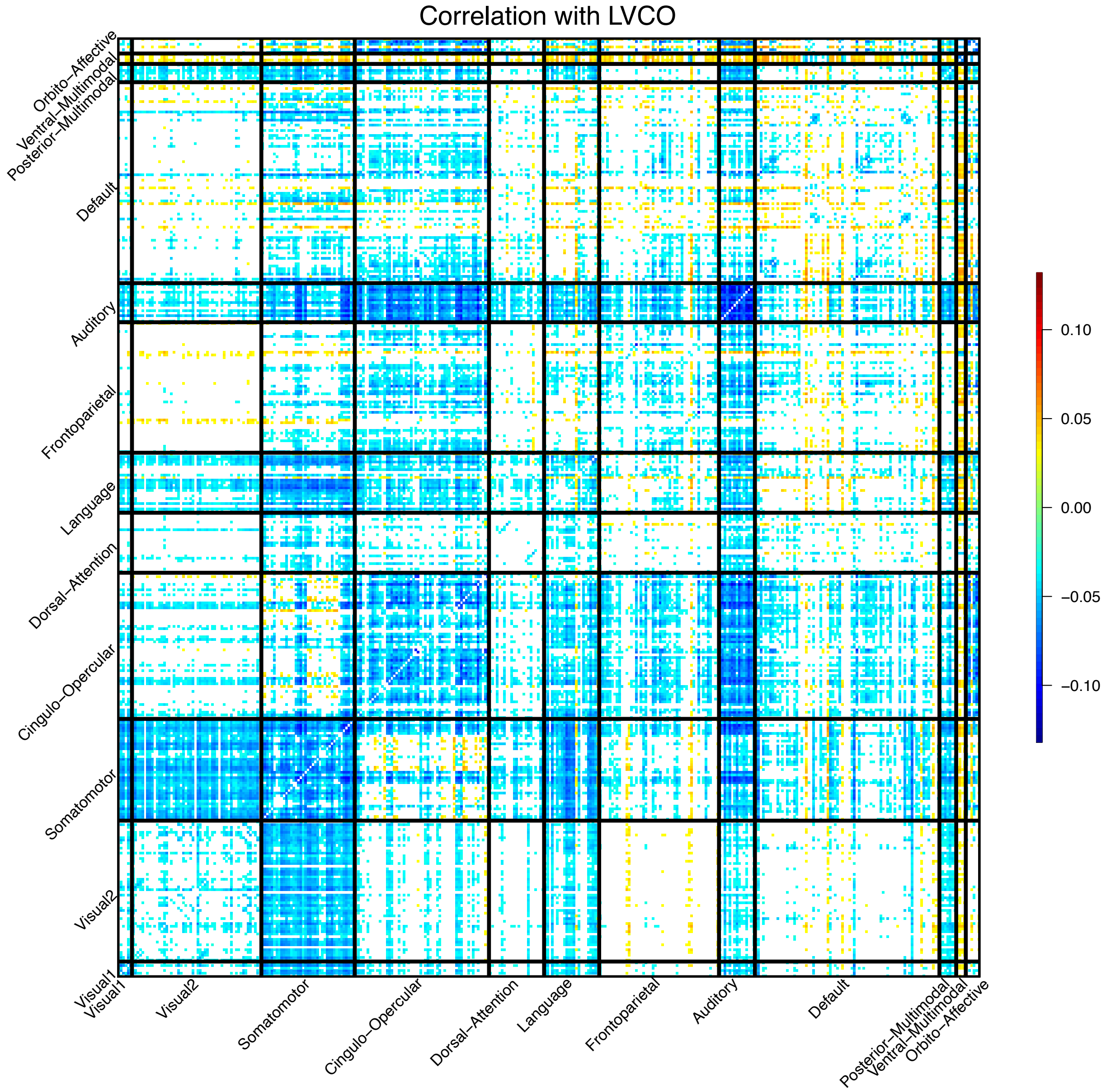


**Fig. S21 Significant associations between left ventricular cardiac output (LVCO) and resting fMRI traits across different networks.** We illustrate significant correlations (at Bonferroni significance level) between LVCO and 64,620 area level resting fMRI traits (both within-network and cross-network). The color represents correlation estimates. Visual1, the primary visual network; Visual2, the secondary visual network.

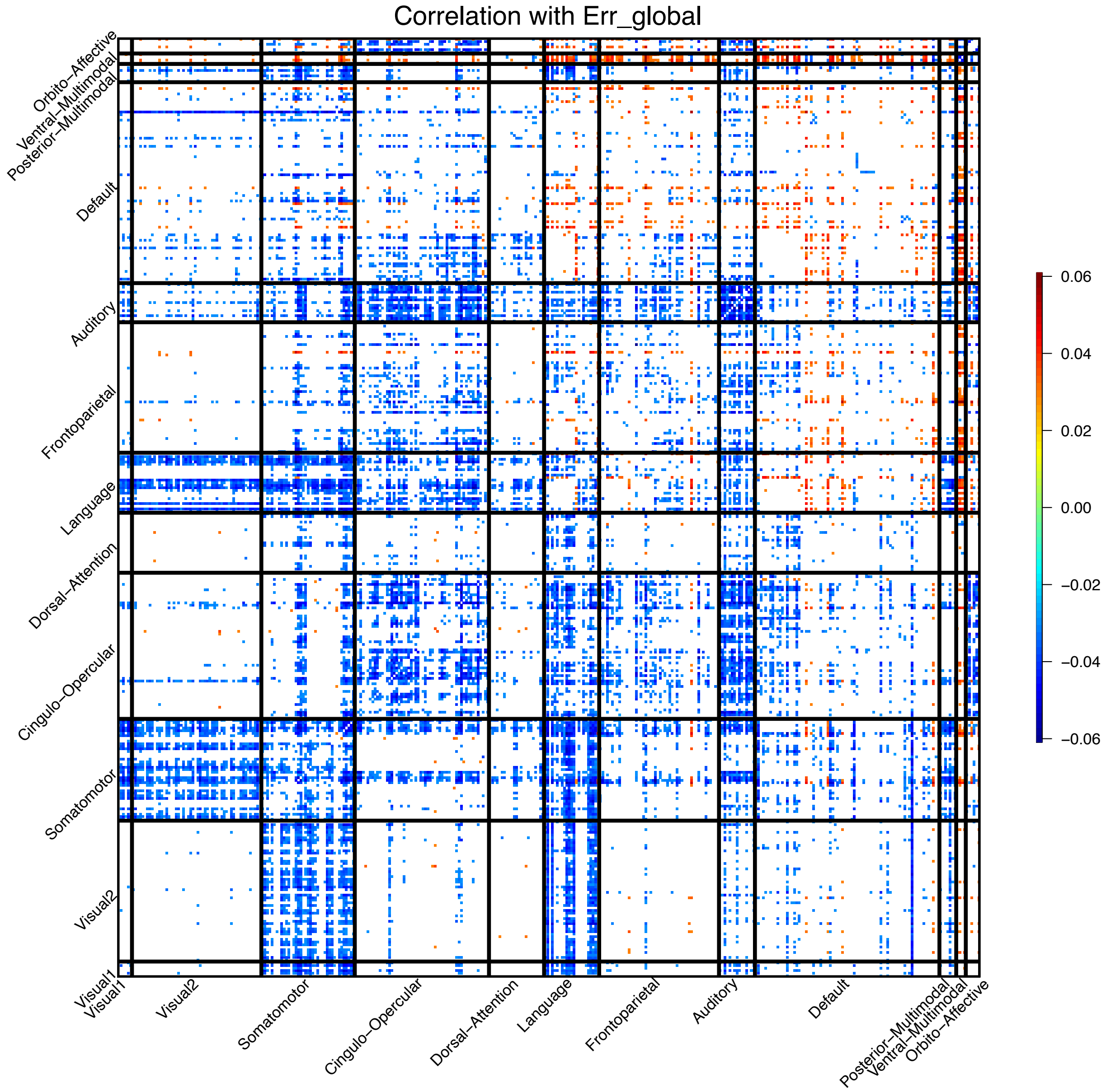


**Fig. S22 Significant associations between global radial strain (Err global) and resting fMRI traits across different networks.** We illustrate significant correlations (at Bonferroni significance level) between Err global and 64,620 area level resting fMRI traits (both within-network and cross-network). The color represents correlation estimates. Visual1, the primary visual network; Visual2, the secondary visual network.

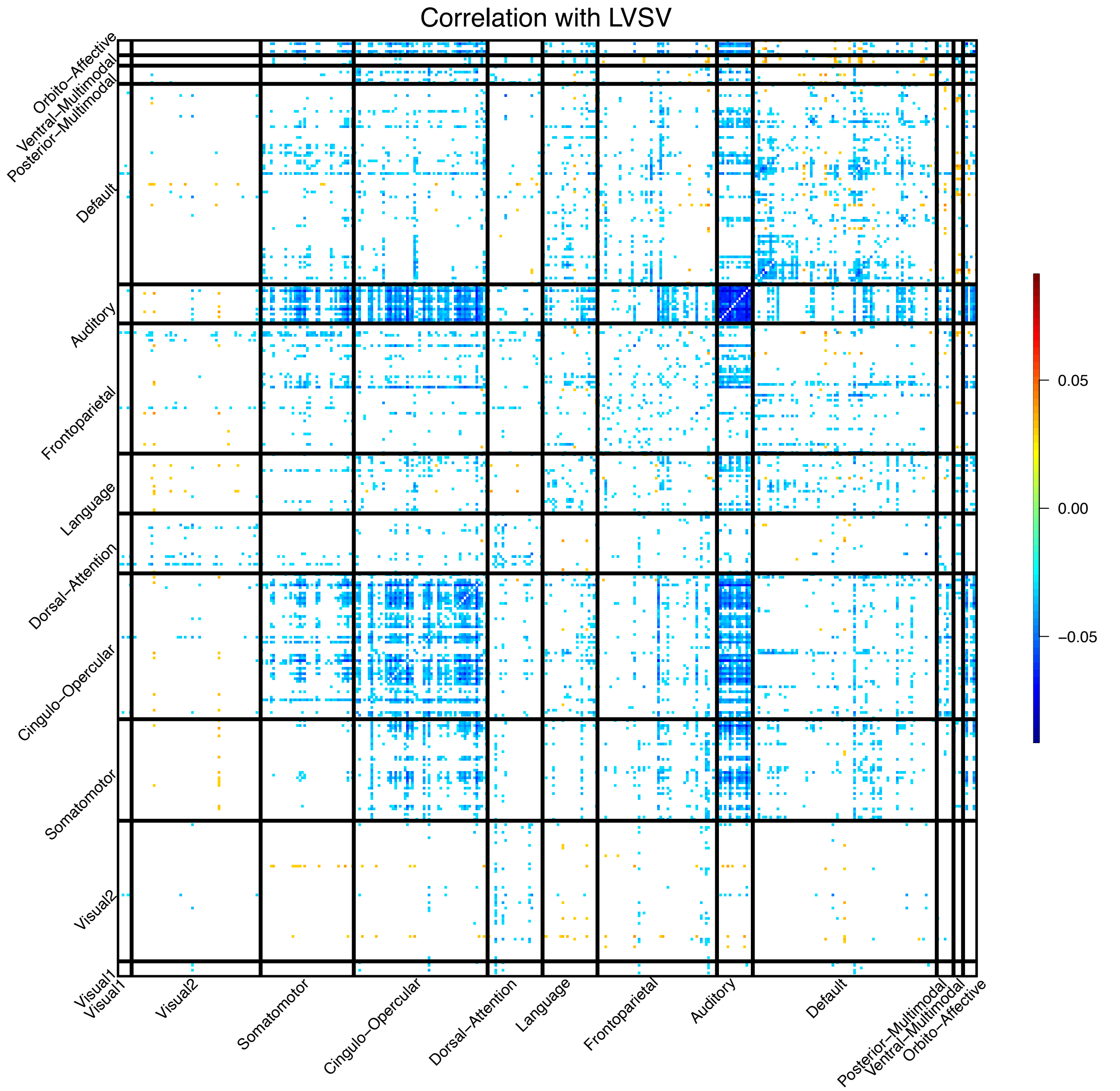


**Fig. S23 Significant associations between left ventricular stroke volume (LVSV) and resting fMRI traits across different networks.** We illustrate significant correlations (at Bonferroni significance level) between LVSV and 64,620 area level resting fMRI traits (both within-network and cross-network). The color represents correlation estimates. Visual1, the primary visual network; Visual2, the secondary visual network.

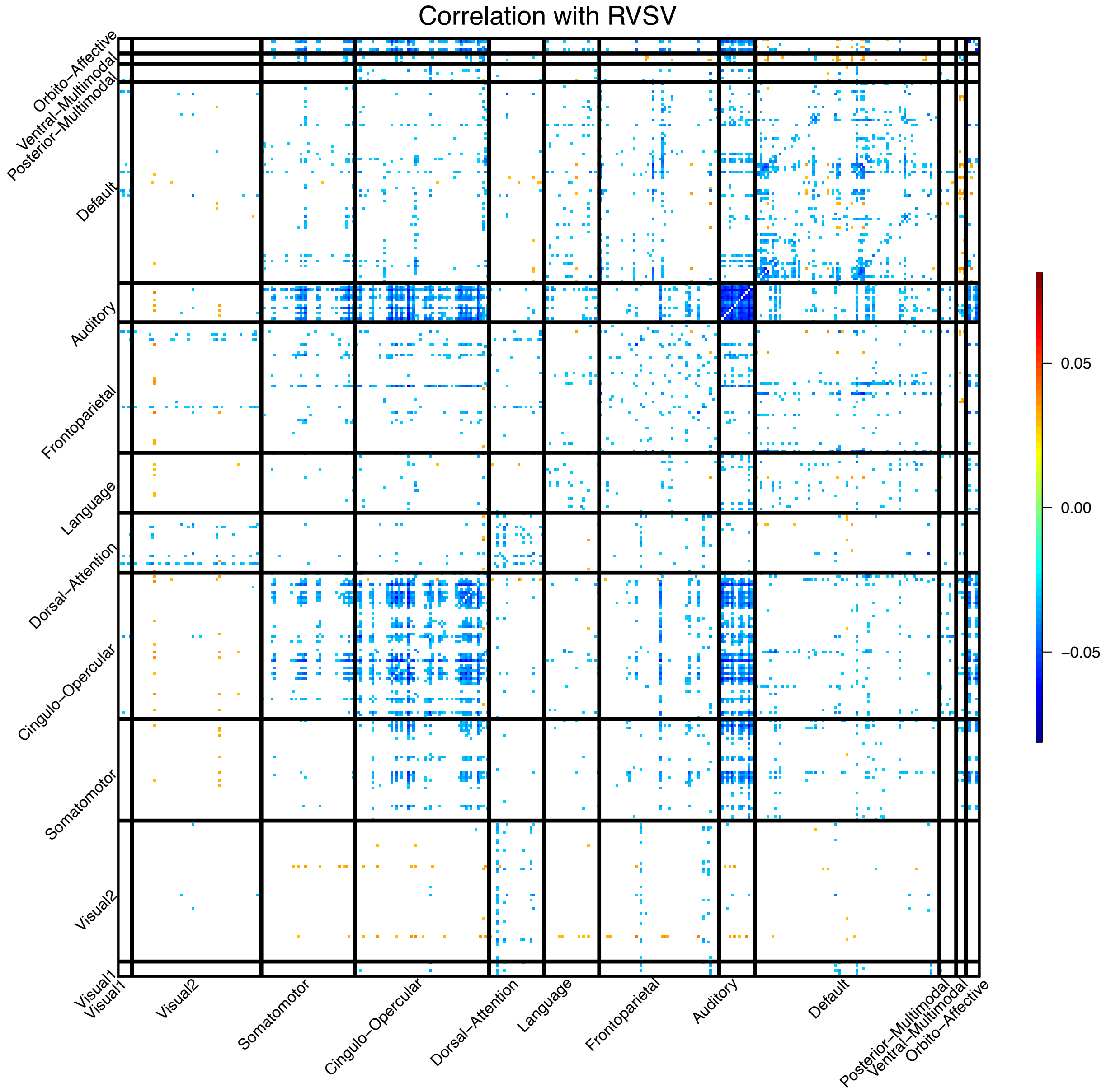


**Fig. S24 Significant associations between right ventricular stroke volume (RVSV) and resting fMRI traits across different networks.** We illustrate significant correlations (at Bonferroni significance level) between RVSV and 64,620 area level resting fMRI traits (both within-network and cross-network). The color represents correlation estimates. Visual1, the primary visual network; Visual2, the secondary visual network.

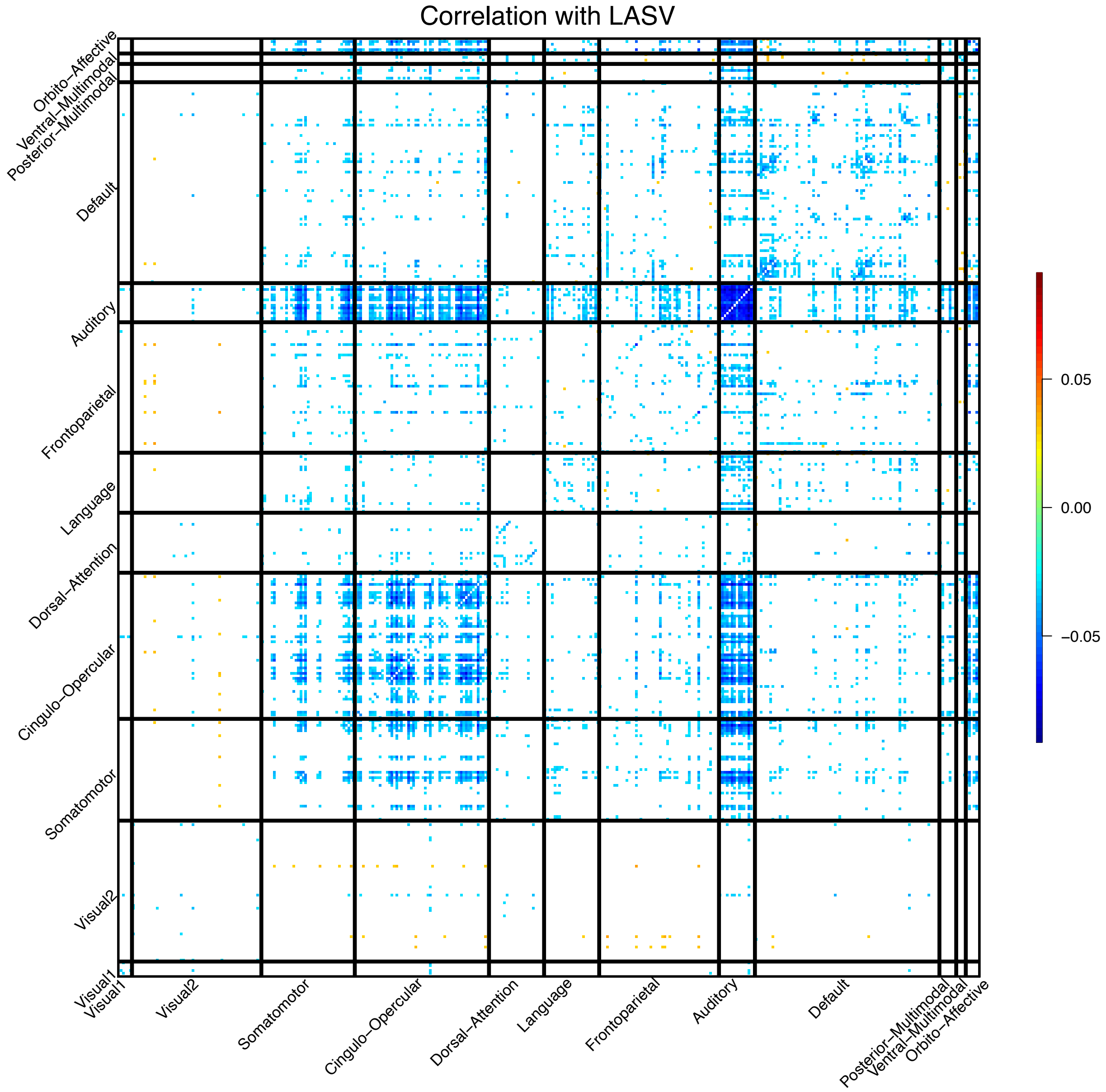


**Fig. S25 Significant associations between left atrium stroke volume (LASV) and resting fMRI traits across different networks.** We illustrate significant correlations (at Bonferroni significance level) between LASV and 64,620 area level resting fMRI traits (both within-network and cross-network). The color represents correlation estimates. Visual1, the primary visual network; Visual2, the secondary visual network.

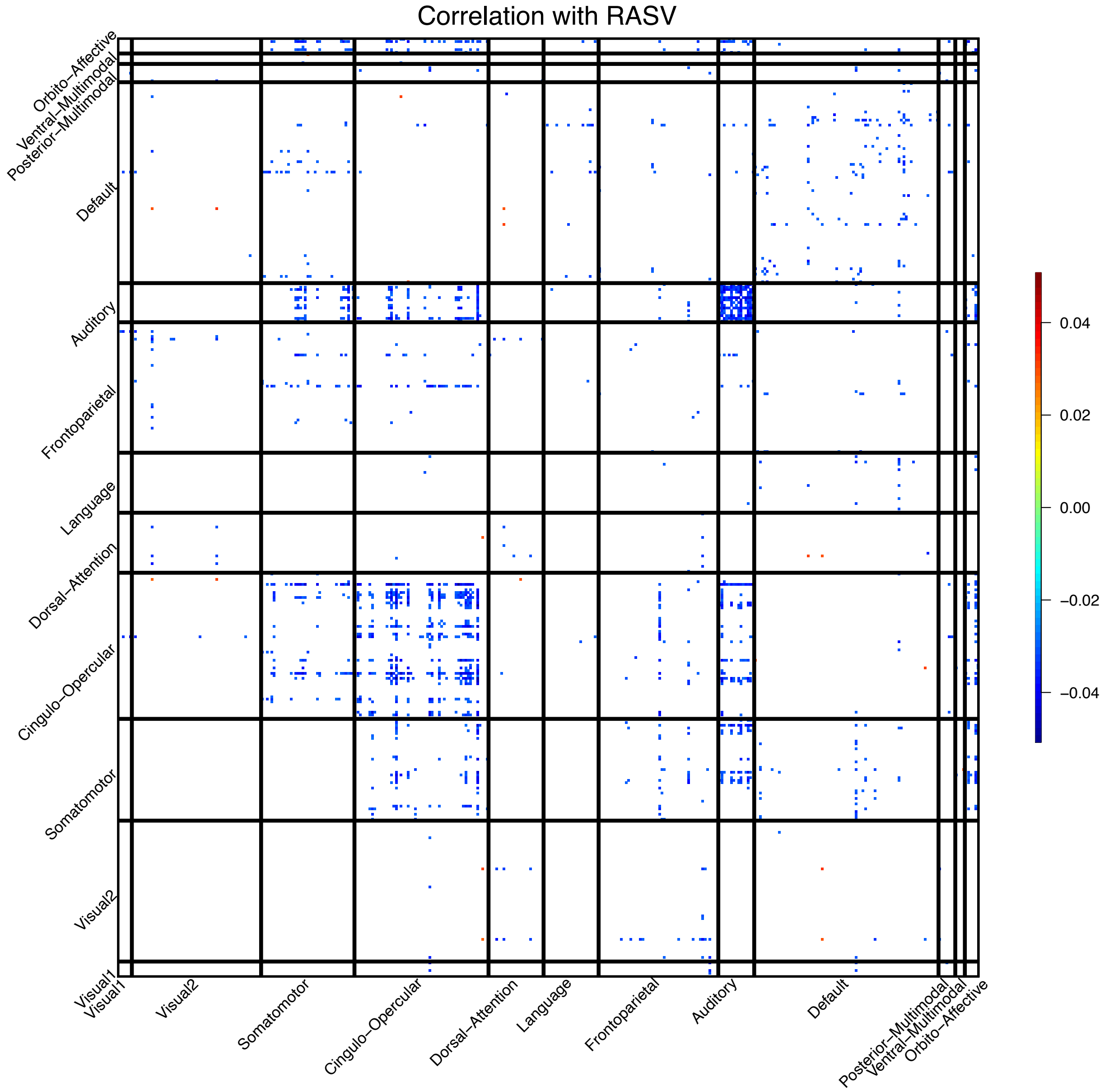


**Fig. S26 Significant associations between right atrium stroke volume (RASV) and resting fMRI traits across different networks.** We illustrate significant correlations (at Bonferroni significance level) between RASV and 64,620 area level resting fMRI traits (both within-network and cross-network). The color represents correlation estimates. Visual1, the primary visual network; Visual2, the secondary visual network.

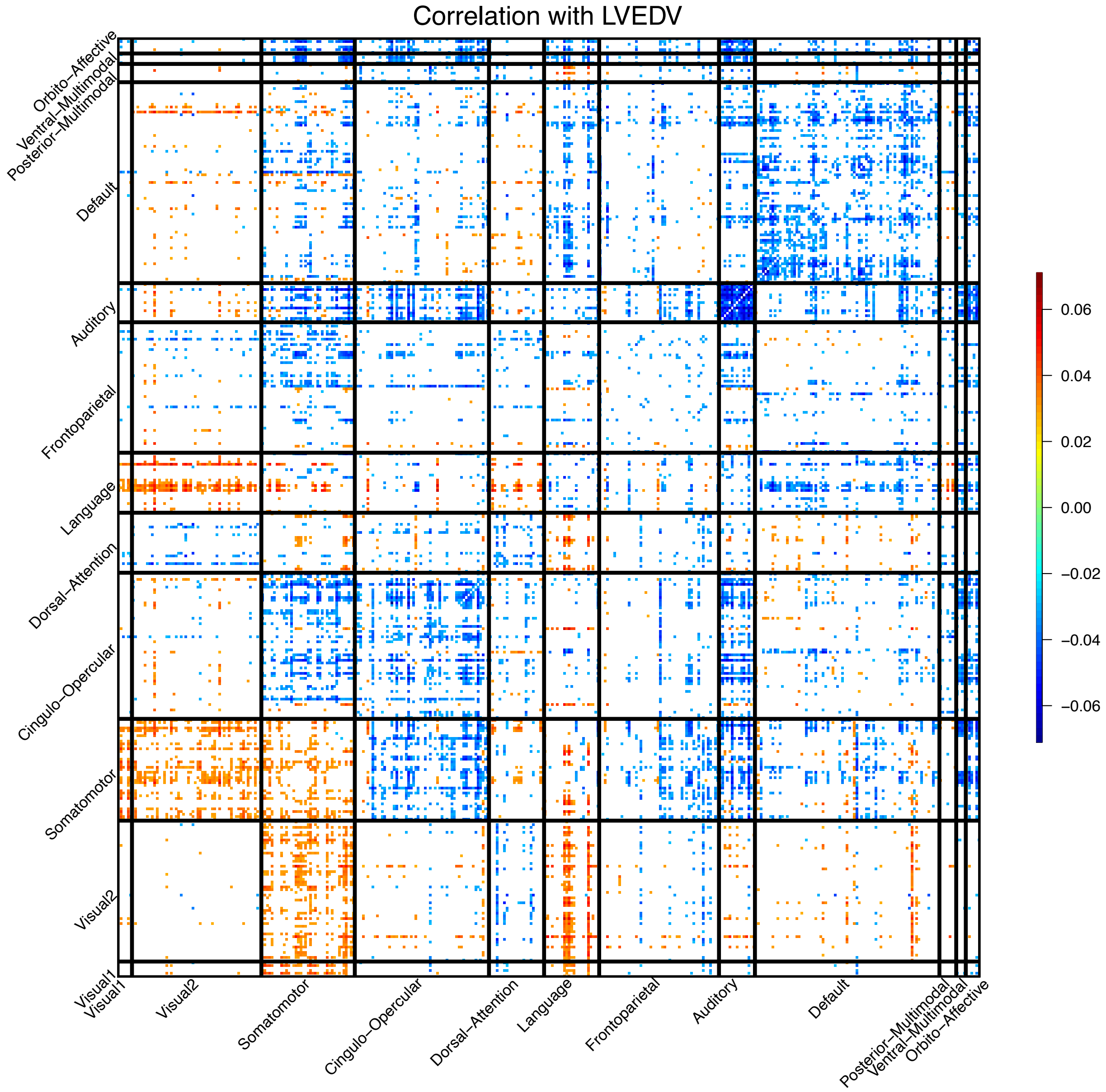


**Fig. S27 Significant associations between left ventricular end-diastolic volume (LVEDV) and resting fMRI traits across different networks.** We illustrate significant correlations (at Bonferroni significance level) between LVEDV and 64,620 area level resting fMRI traits (both within-network and cross-network). The color represents correlation estimates. Visual1, the primary visual network; Visual2, the secondary visual network.

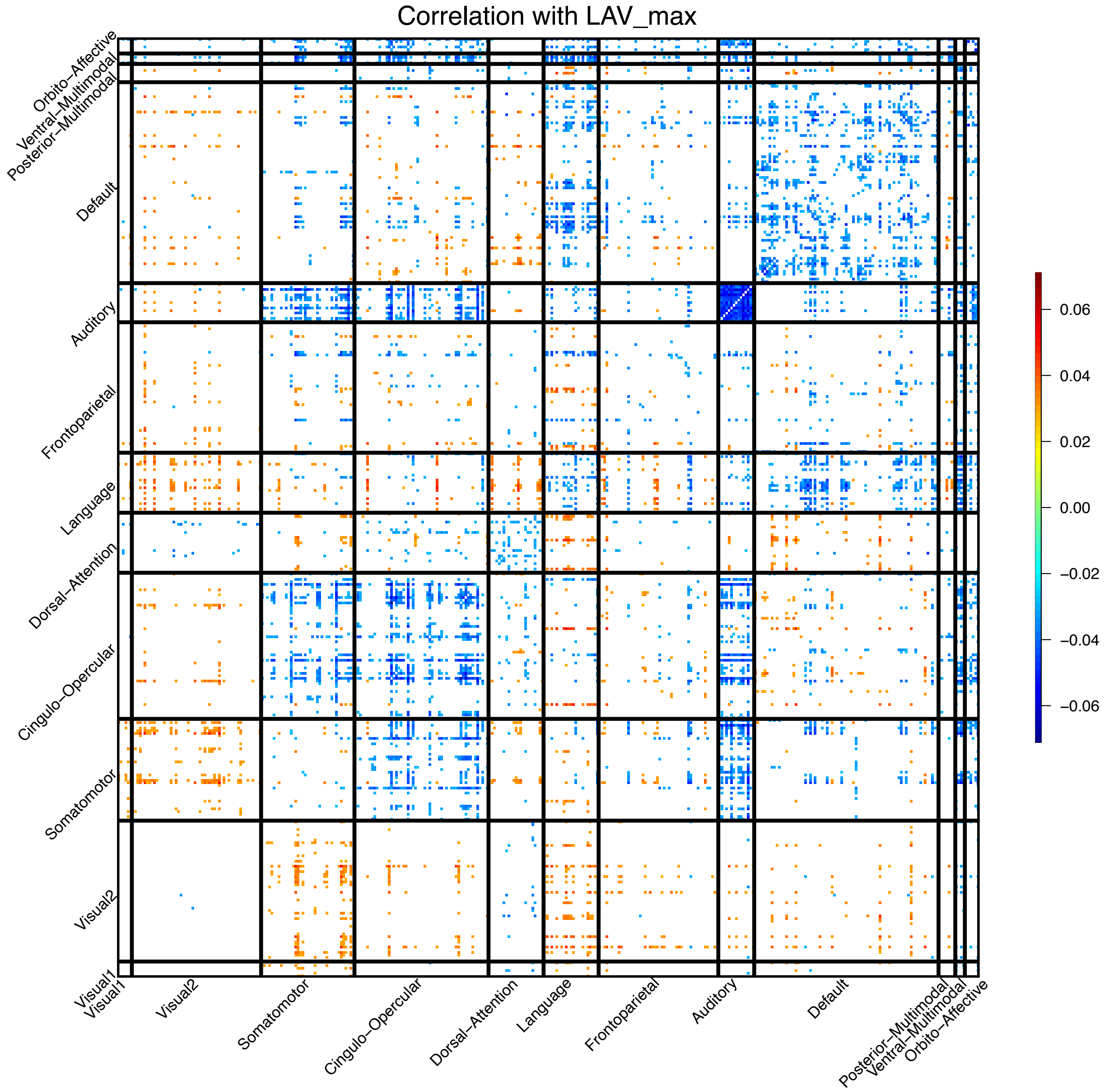


**Fig. S28 Significant associations between left atrium maximum volume (LAV max) and resting fMRI traits across different networks.** We illustrate significant correlations (at Bonferroni significance level) between LAV max and 64,620 area level resting fMRI traits (both within-network and cross-network). The color represents correlation estimates. Visual1, the primary visual network; Visual2, the secondary visual network.

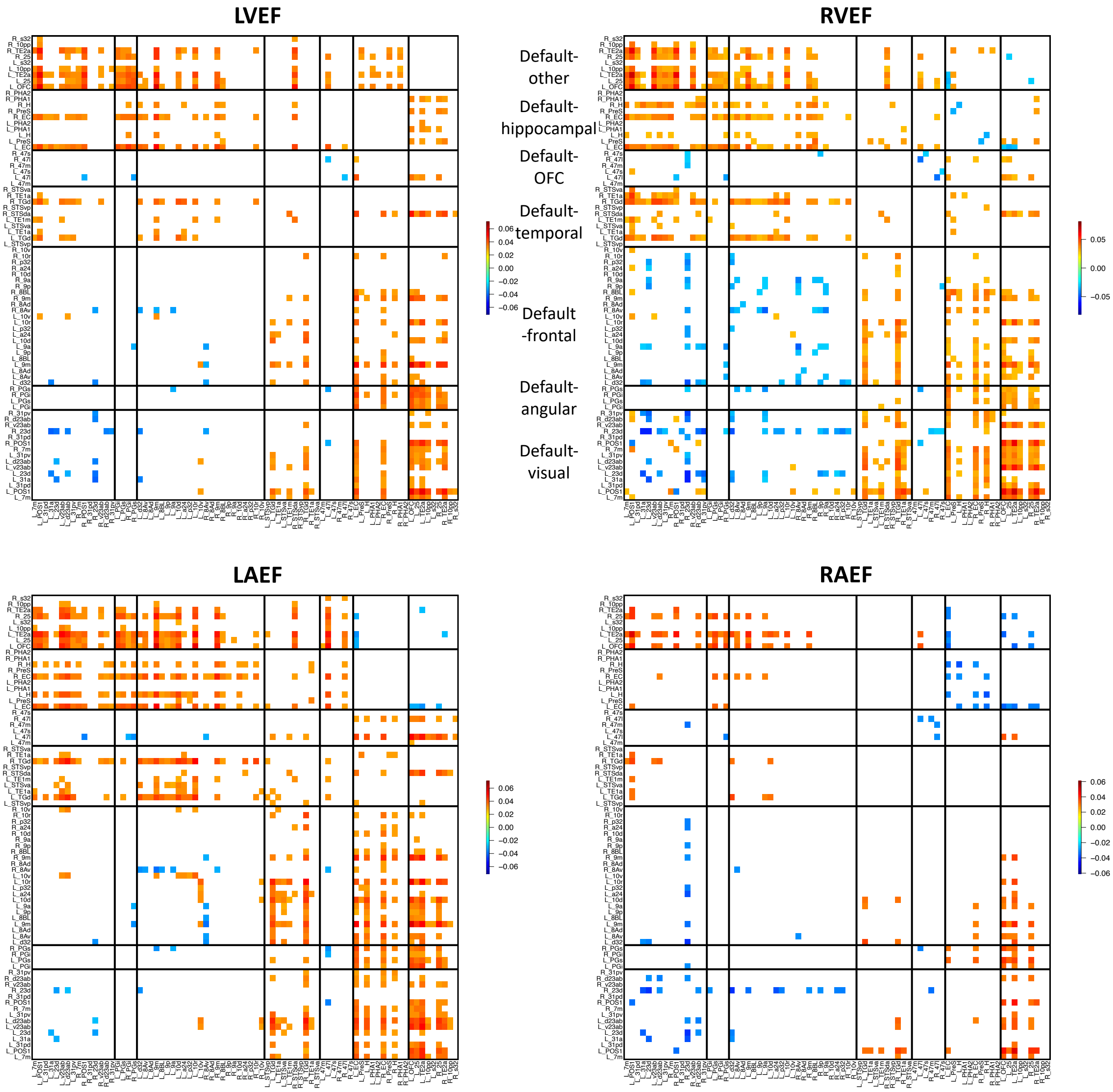


**Fig. S29 Significant associations between ejection fraction traits and the default mode network in resting fMRI.** We illustrate significant correlations (at Bonferroni significance level) between the 4 ejection fraction traits (RVEF, LAEF, RAEF, and LVEF) and resting fMRI traits in the default mode network. The color represents correlation estimates. We group all functional areas in the default mode network into seven clusters. These areas are mainly organized by their physical locations. OFC, orbitofrontal complex.

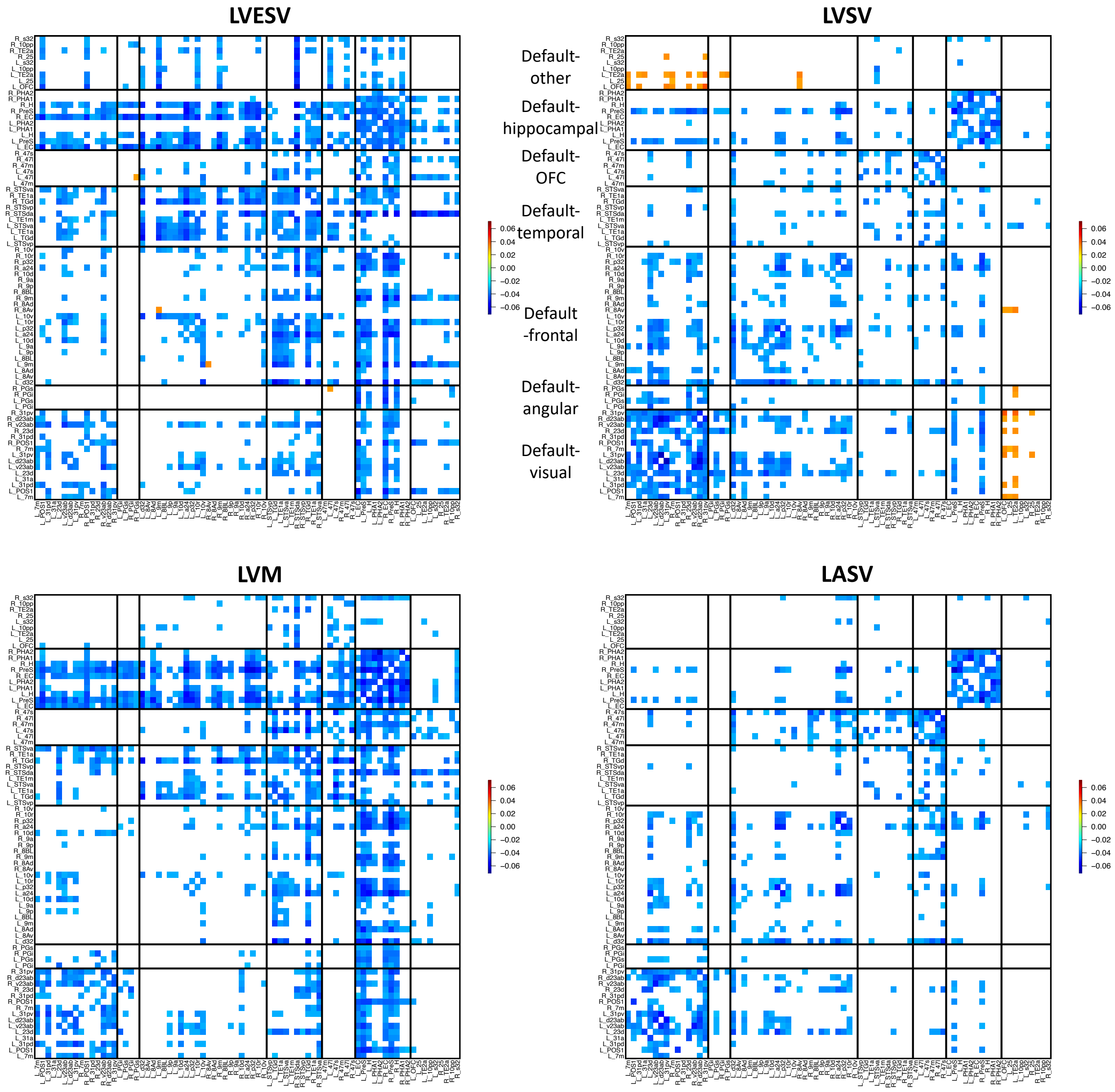


**Fig. S30 Significant associations between left heart volumetric measures and the default mode network in resting fMRI.** We illustrate significant correlations (at Bonferroni significance level) between the 4 left heart volumetric measures (LVESV, LVSV, LVM, and LASV) and resting fMRI traits in the default mode network. The color represents correlation estimates. We group all functional areas in the default mode network into seven clusters. These areas are mainly organized by their physical locations. OFC, orbitofrontal complex.

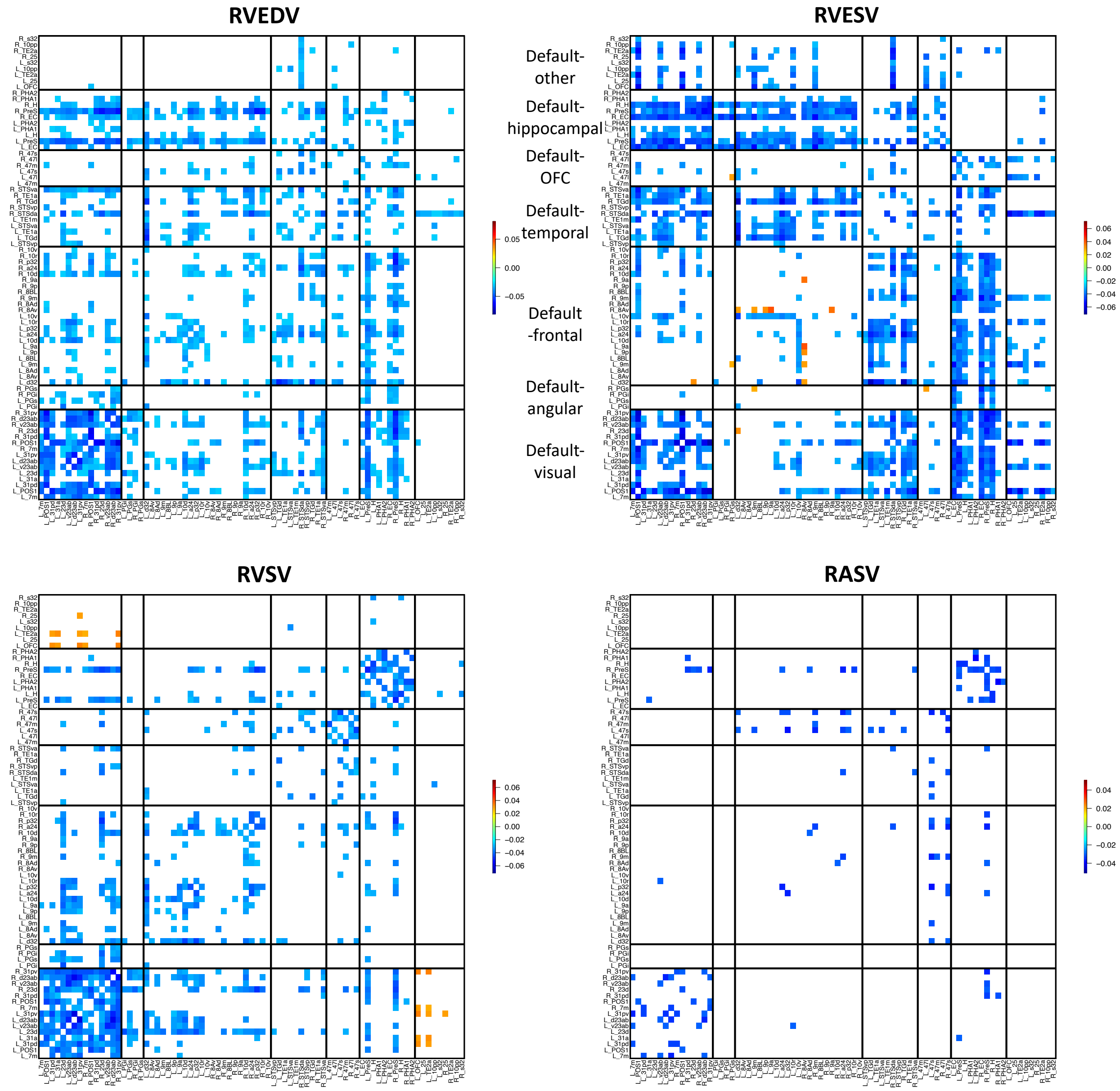


**Fig. S31 Significant associations between right heart volumetric measures and the default mode network in resting fMRI.** We illustrate significant correlations (at Bonferroni significance level) between the 4 right heart volumetric measures (RVEDV, RVESV, RVSV, and RASV) and resting fMRI traits in the default mode network. The color represents correlation estimates. We group all functional areas in the default mode network into seven clusters. These areas are mainly organized by their physical locations. OFC, orbitofrontal complex.

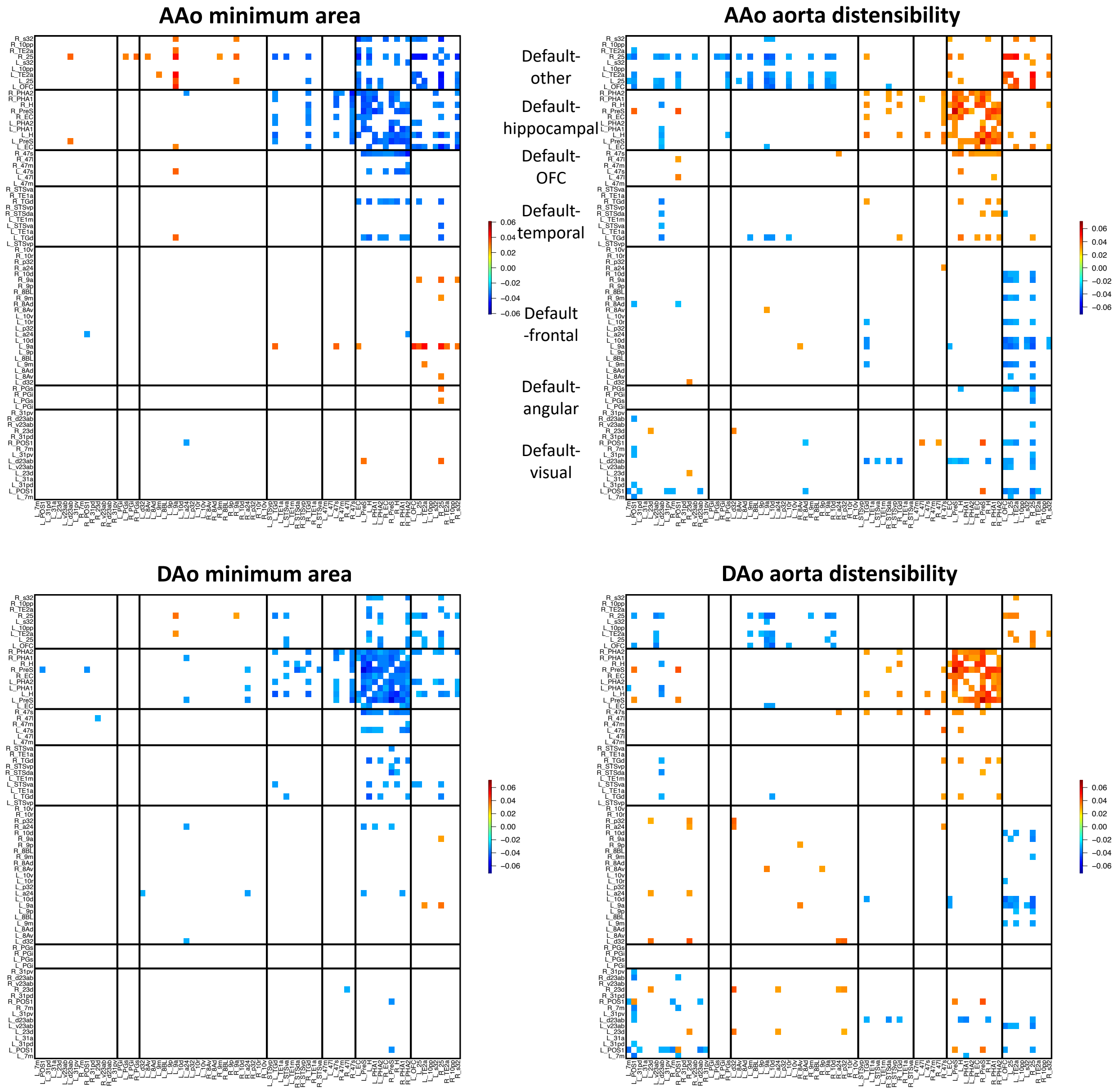


**Fig. S32 Significant associations between aortic measures and the default mode network in resting fMRI.** We illustrate significant correlations (at Bonferroni significance level) between 4 aortic measures (AAo minimum area, DAo minimum area, AAo aorta distensibility, and DAo aorta distensibility) and resting fMRI traits in the default mode network. The color represents correlation estimates. We group all functional areas in the default mode network into seven clusters. These areas are mainly organized by their physical locations. OFC, orbitofrontal complex.

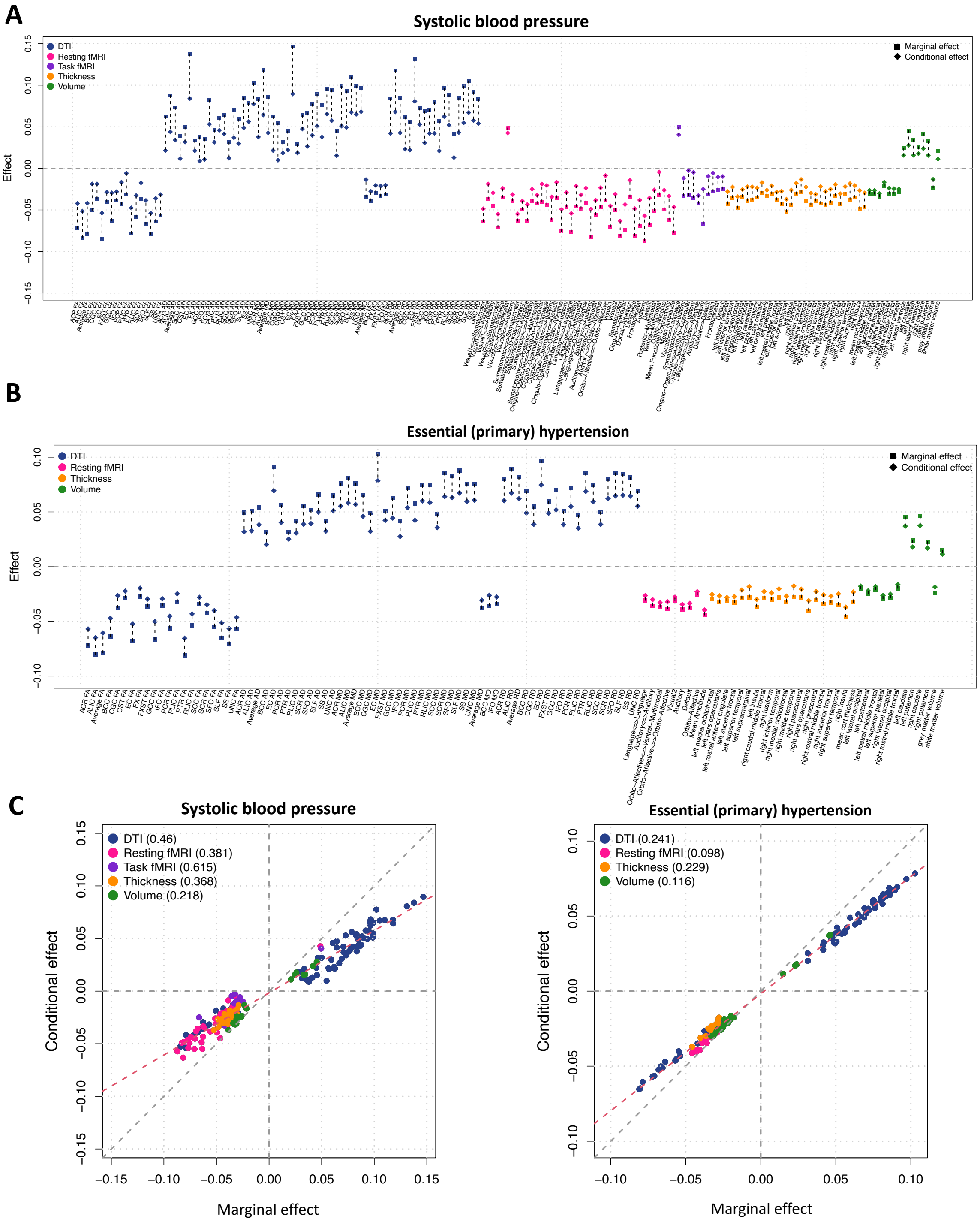


**Fig. S33 Effects of systolic blood pressure and hypertension on brain MRI that are mediated through CMR traits.** We illustrate marginal effects (after adjusting for covariates) of systolic blood pressure (**A**) and hypertension (**B**) on brain MRI traits (*x* axis) and the corresponding conditional effects after further adjusting for CMR traits. The difference between marginal and condition effects indicates the indirect effect mediated through CMR traits. In **(C)**, we label the average proportion of CMR-mediated effects for each brain MRI modality. We label brain MRI modalities with different colors.

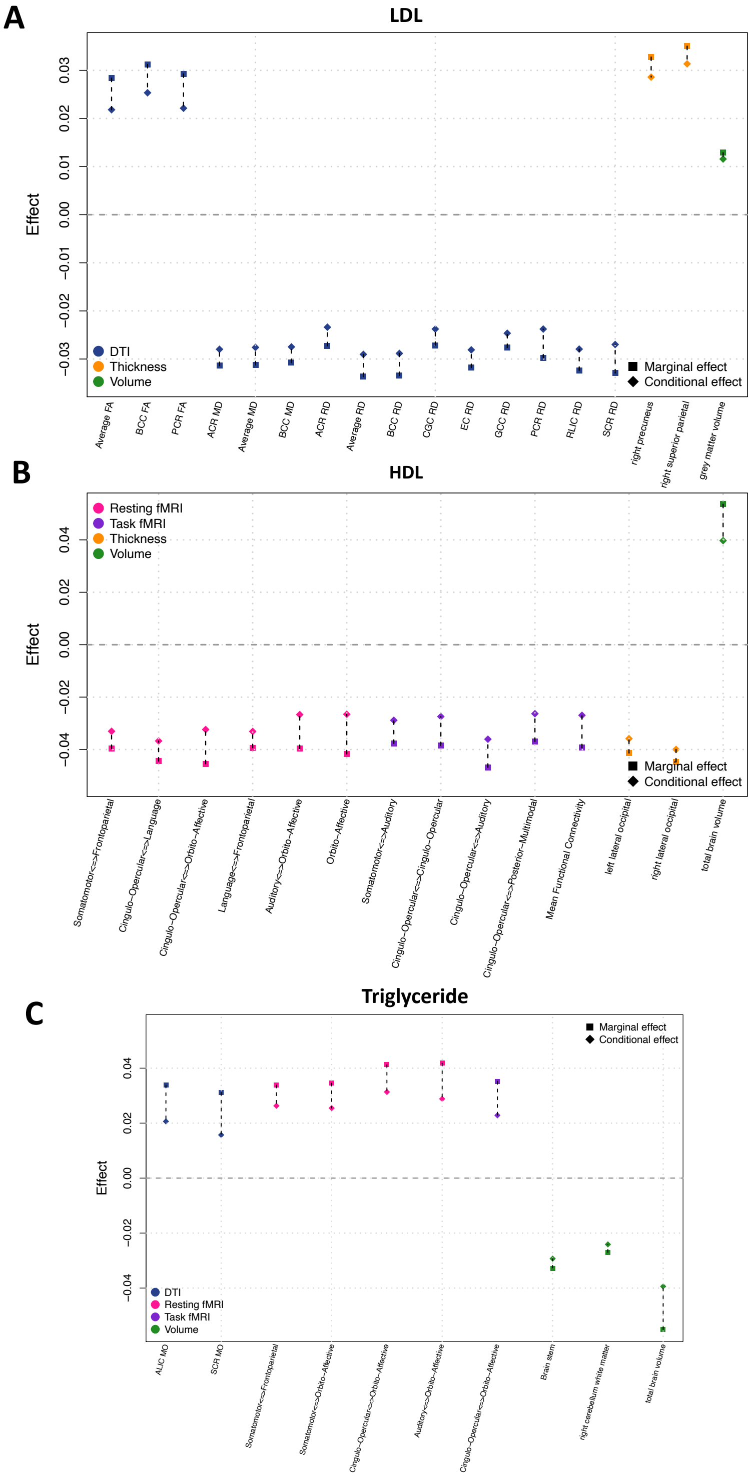


**Fig. S34 Effects of cardiovascular biomarkers (LDL, HDL, and triglyceride) on brain MRI that are mediated through CMR traits.** We illustrate marginal effects (after adjusting for covariates) of cardiovascular biomarkers on brain MRI traits (*x* axis) and the corresponding conditional effects after further adjusting for CMR traits. The difference between marginal and condition effects indicates the indirect effect mediated through CMR traits. We label brain MRI modalities with different colors.

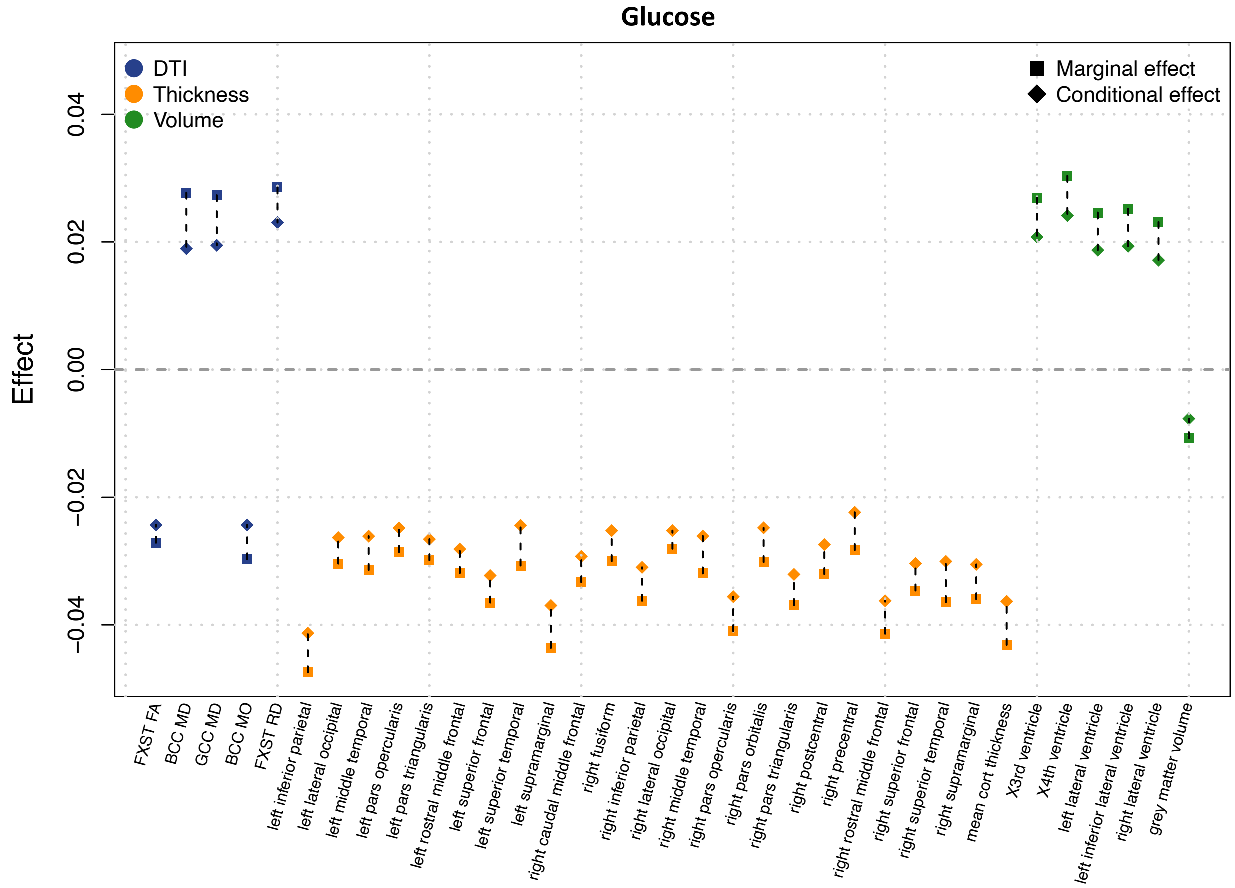


**Fig. S35 Effects of glucose on brain MRI mediated that are through CMR traits.** We illustrate marginal effects (after adjusting for covariates) of glucose on brain MRI traits (*x* axis) and the corresponding conditional effects after further adjusting for CMR traits. The difference between marginal and condition effects indicates the indirect effect mediated through CMR traits. We label brain MRI modalities with different colors.

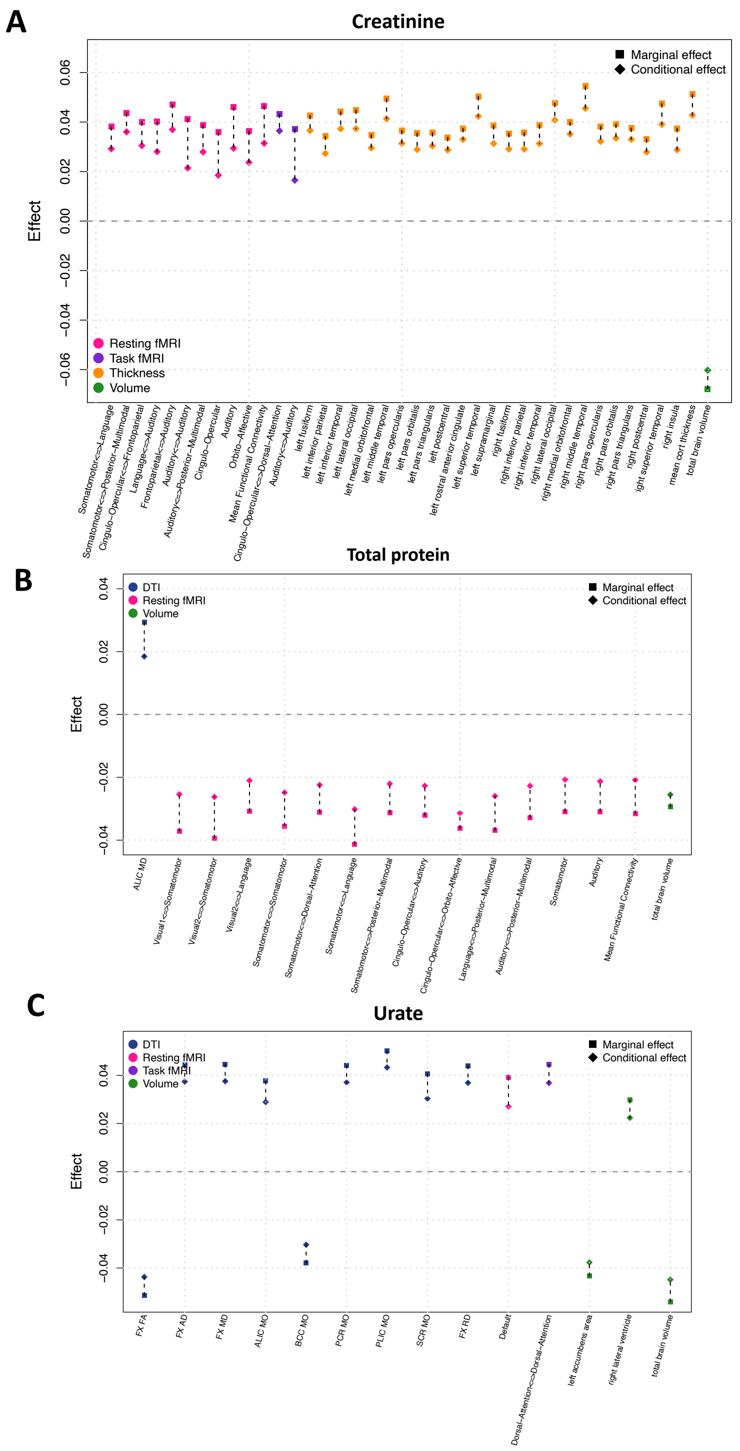


**Fig. S36 Effects of kidney biomarkers (creatinine, total protein, and urate) on brain MRI that are mediated through CMR traits.** We illustrate marginal effects (after adjusting for covariates) of kidney biomarkers on brain MRI traits (*x* axis) and the corresponding conditional effects after further adjusting for CMR traits. The difference between marginal and condition effects indicates the indirect effect mediated through CMR traits. We label brain MRI modalities with different colors.

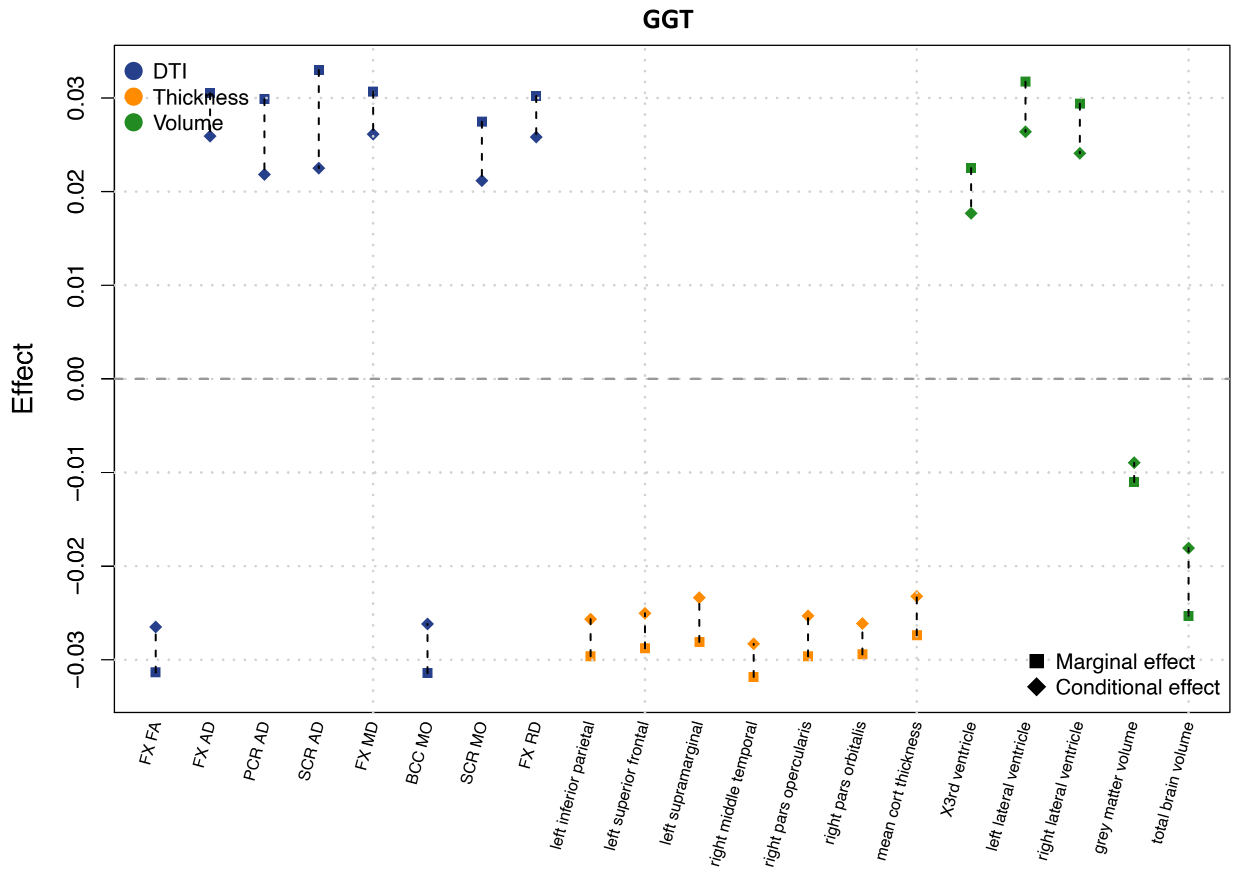


**Fig. S37 Effects of liver biomarker (GGT) on brain MRI that are mediated through CMR traits.** We illustrate marginal effects (adjusted for covariates) of GGT on brain MRI traits (*x* axis) and the corresponding conditional effects after further adjusting for CMR traits. The difference between marginal and condition effects indicates the indirect effects mediated through CMR traits. We label brain MRI modalities with different colors.

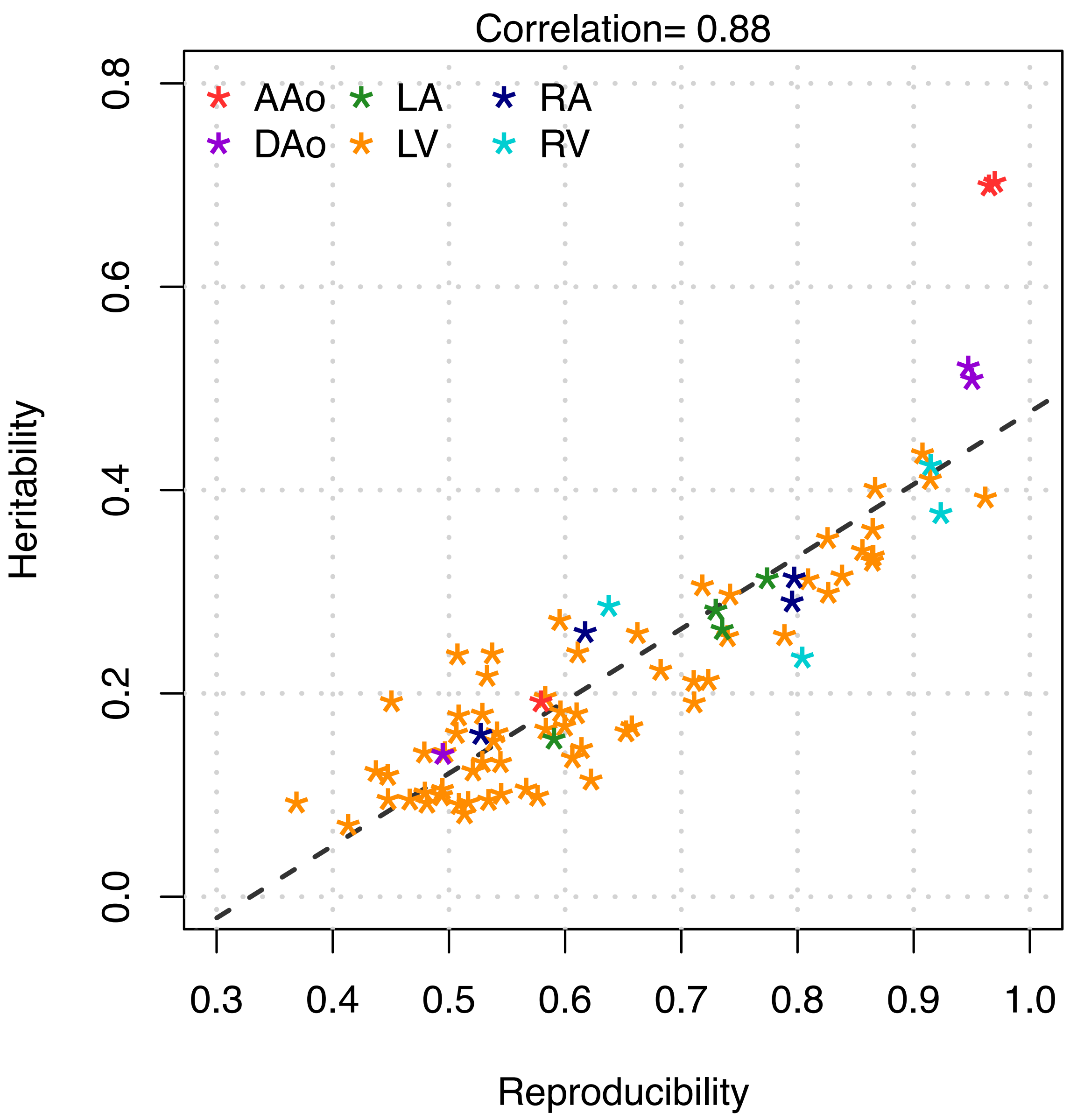


**Fig. S38 Relationship between reproducibility and heritability across the 82 CMR traits.** We label the categories of CMR traits with different colors. AAo, ascending aorta; DAo, descending aorta; LA, left atrium; LV, left ventricle; RA, right atrium; and RV, right ventricle.

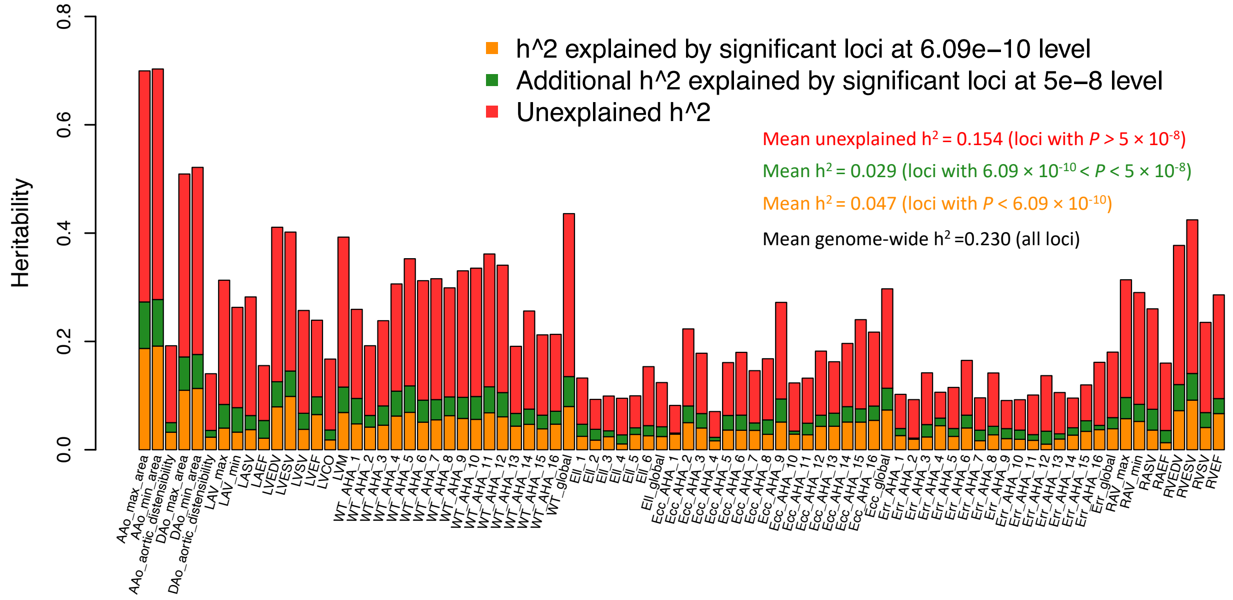


**Fig. S39 Heritability of 82 CMR traits that can be explained by significant loci identified in GWAS.**

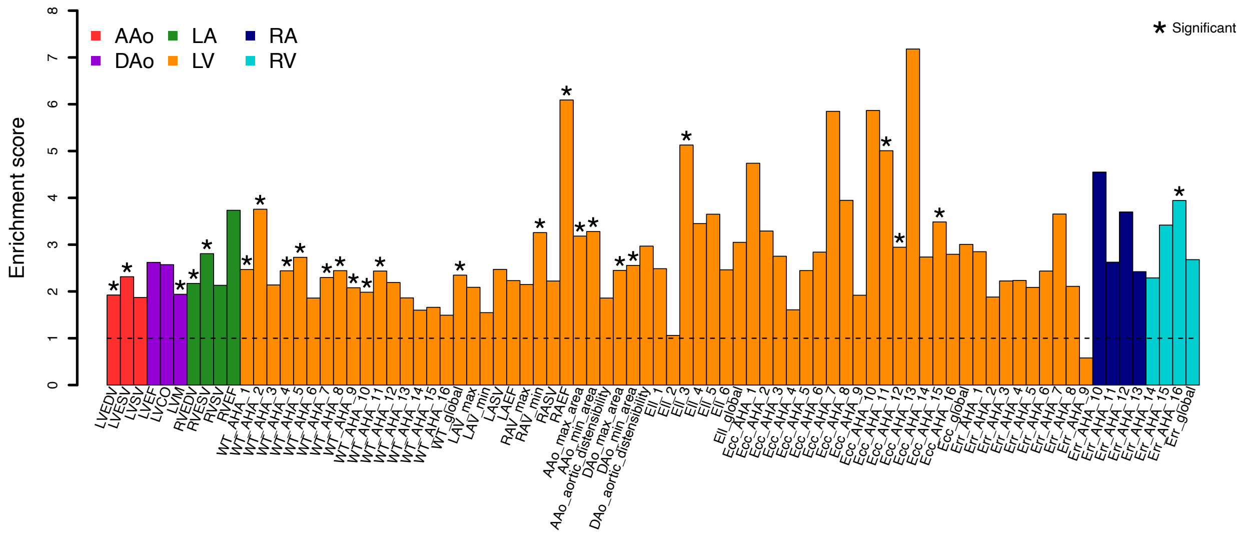


**Fig. S40 Partitioned heritability enrichment analysis of 82 CMR traits for DTI annotations.** We treated previously identified DTI-associated genes as an annotation and performed partitioned heritability enrichment (the proportion of explained heritability/proportion of variants) analysis through LDSC. Significant enrichments are highlighted by asterisks. We label the categories of CMR traits with different colors. AAo, ascending aorta; DAo, descending aorta; LA, left atrium; LV, left ventricle; RA, right atrium; and RV, right ventricle.

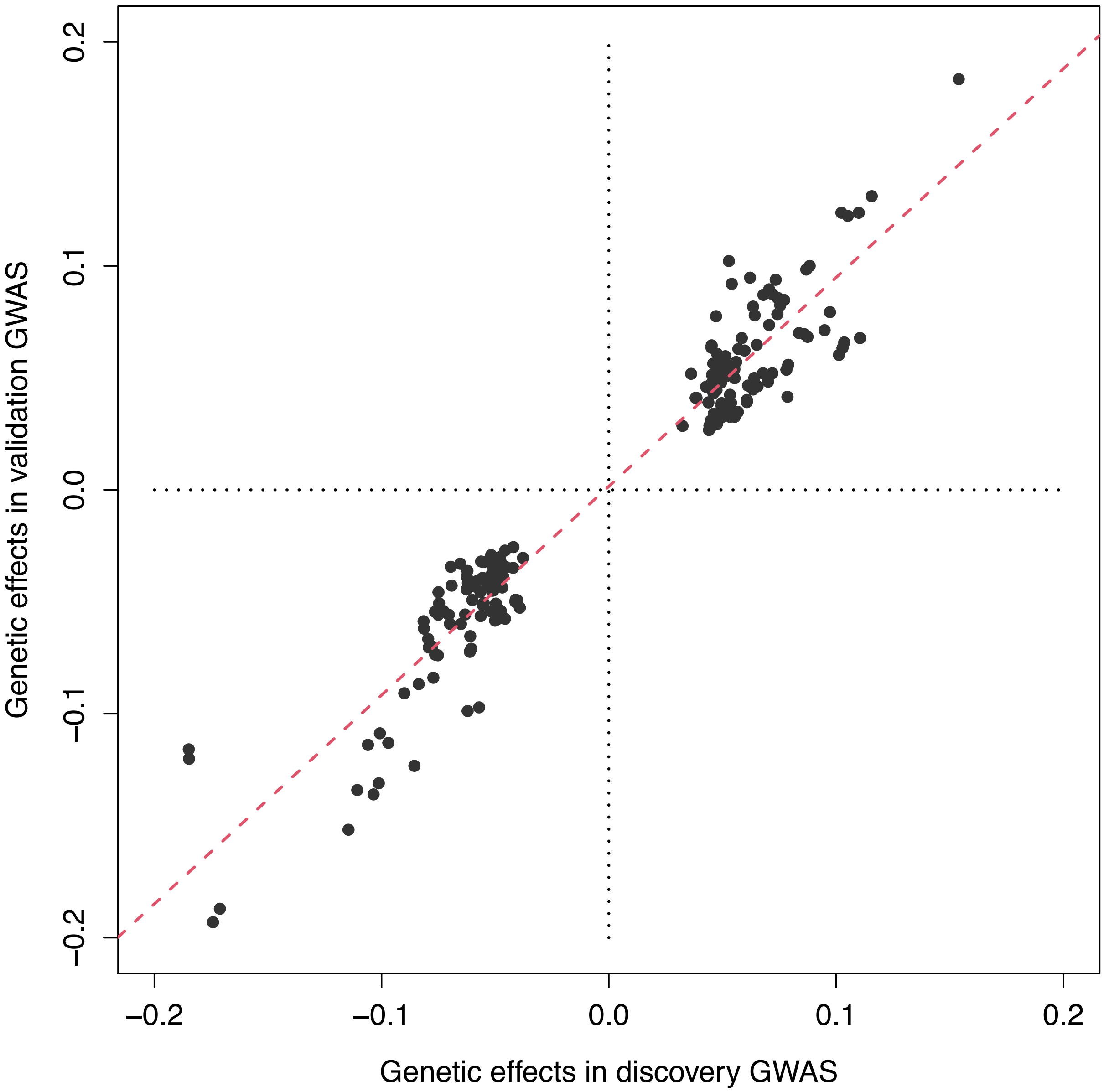


**Fig. S41 Relationship between significant genetic effects estimated in discovery GWAS and validation GWAS.** We illustrate the 184 significant ($P < 6.09 \times 10^{-10}$) CMR-variant associations identified in discovery GWAS. These associations were also significant in the validation GWAS at nominal significance level (0.05).

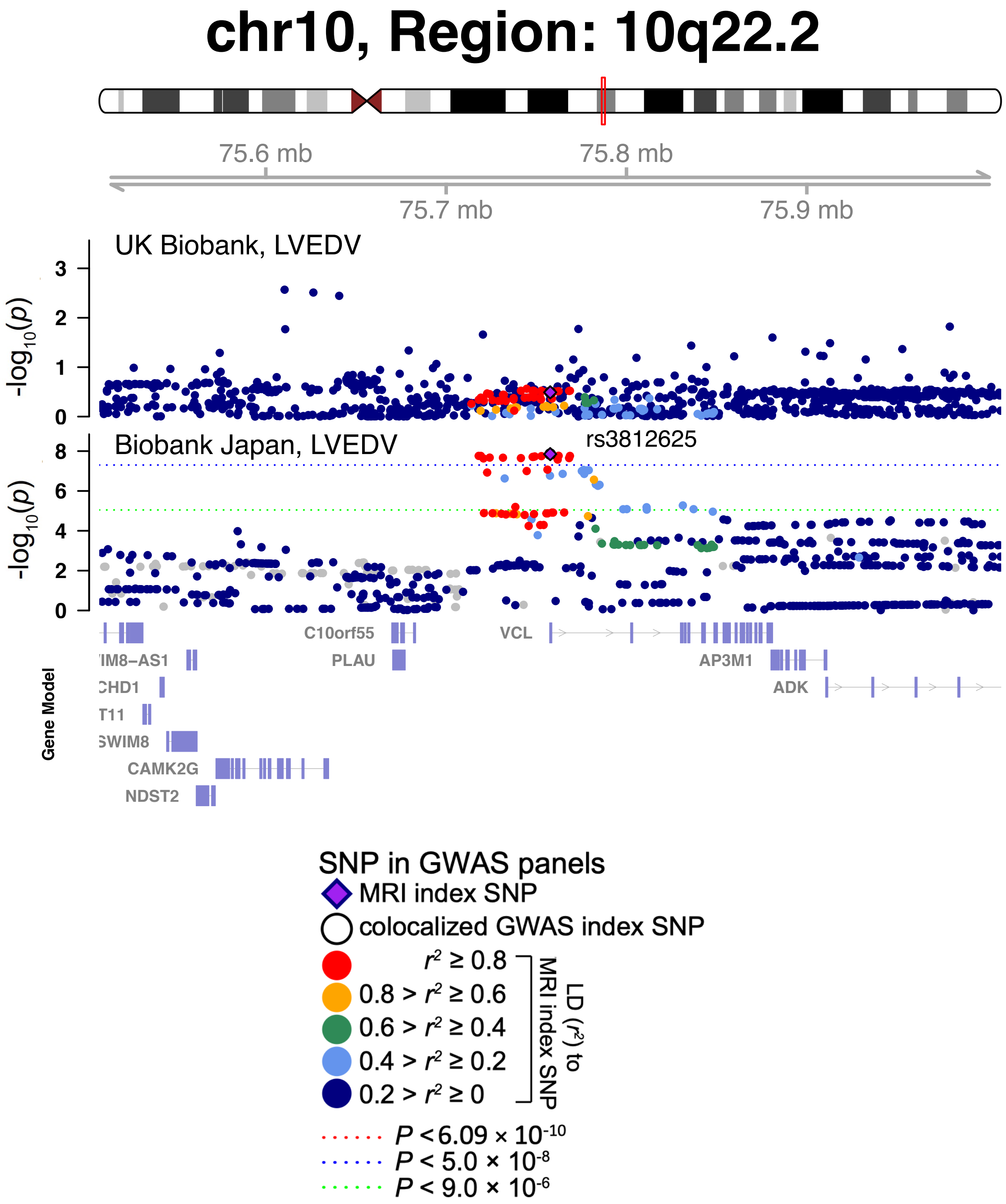


**Fig. S42 The 10q22.2 region was significantly associated with LVEDV in Biobank Japan but not in UK Biobank.**

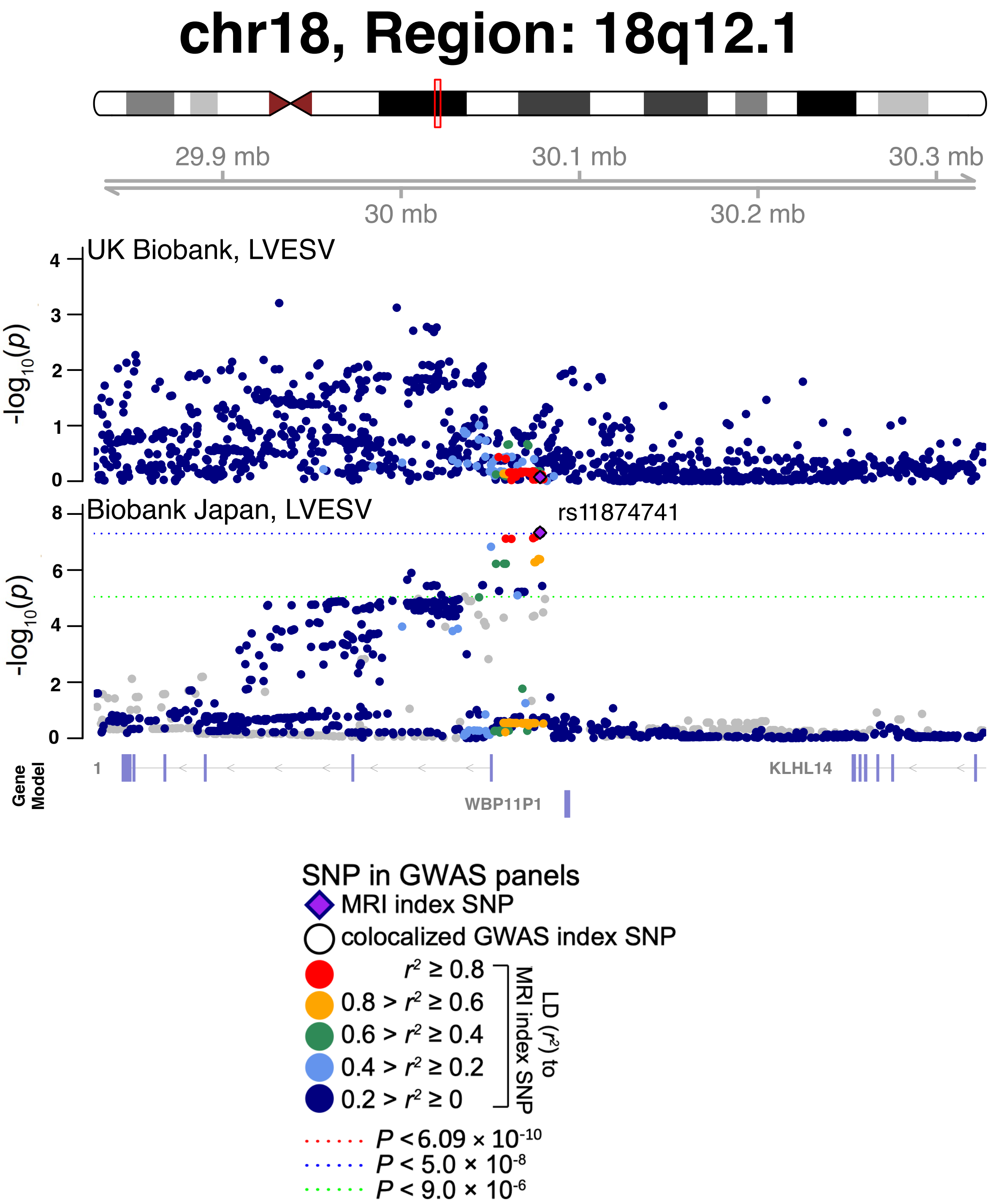


**Fig. S43 The 18q12.1 region was significantly associated with LVESV in Biobank Japan but not in UK Biobank.**

5

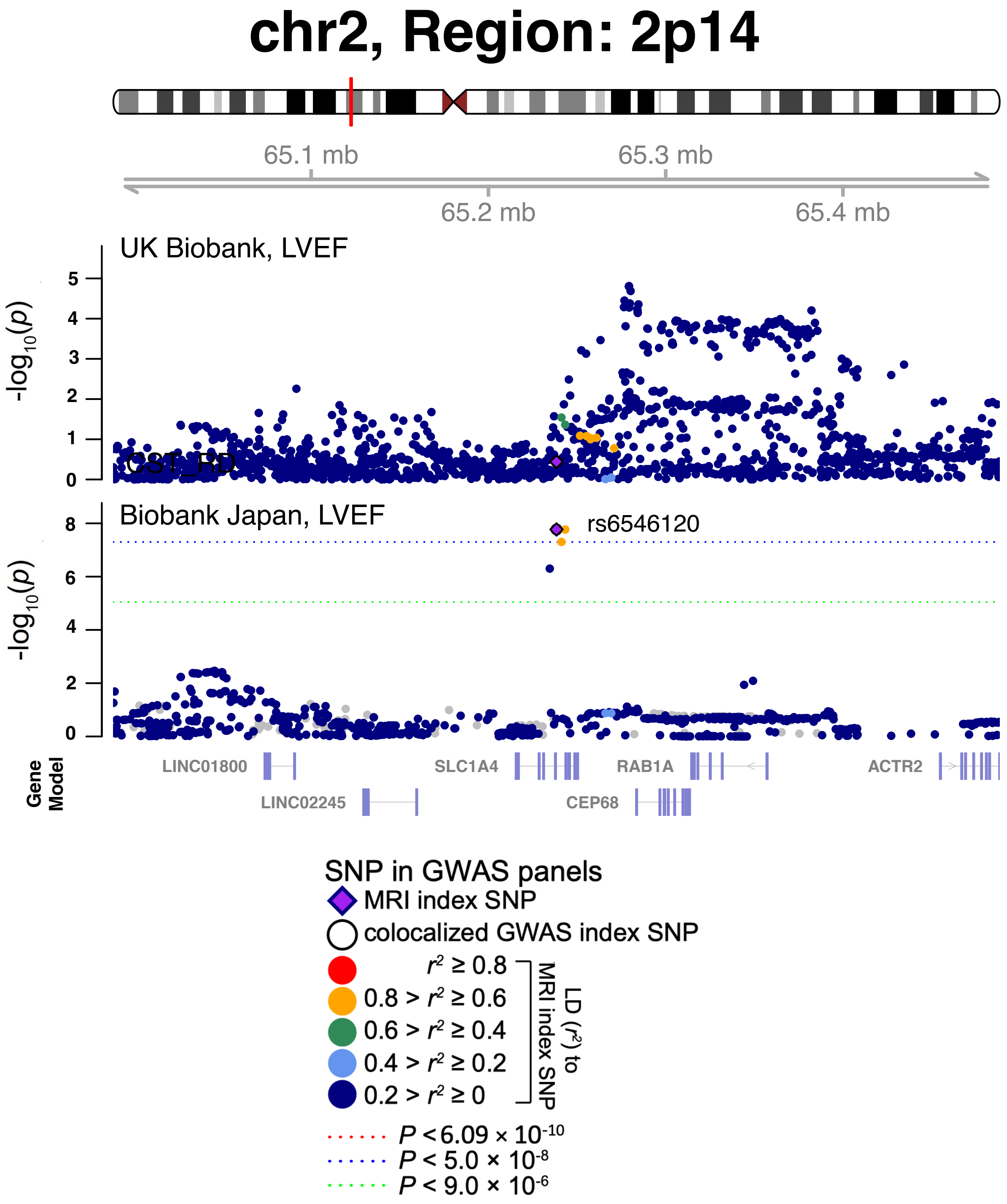


**Fig. S44 The 2p14 region was significantly associated with LVESV in Biobank Japan but not in UK Biobank.**

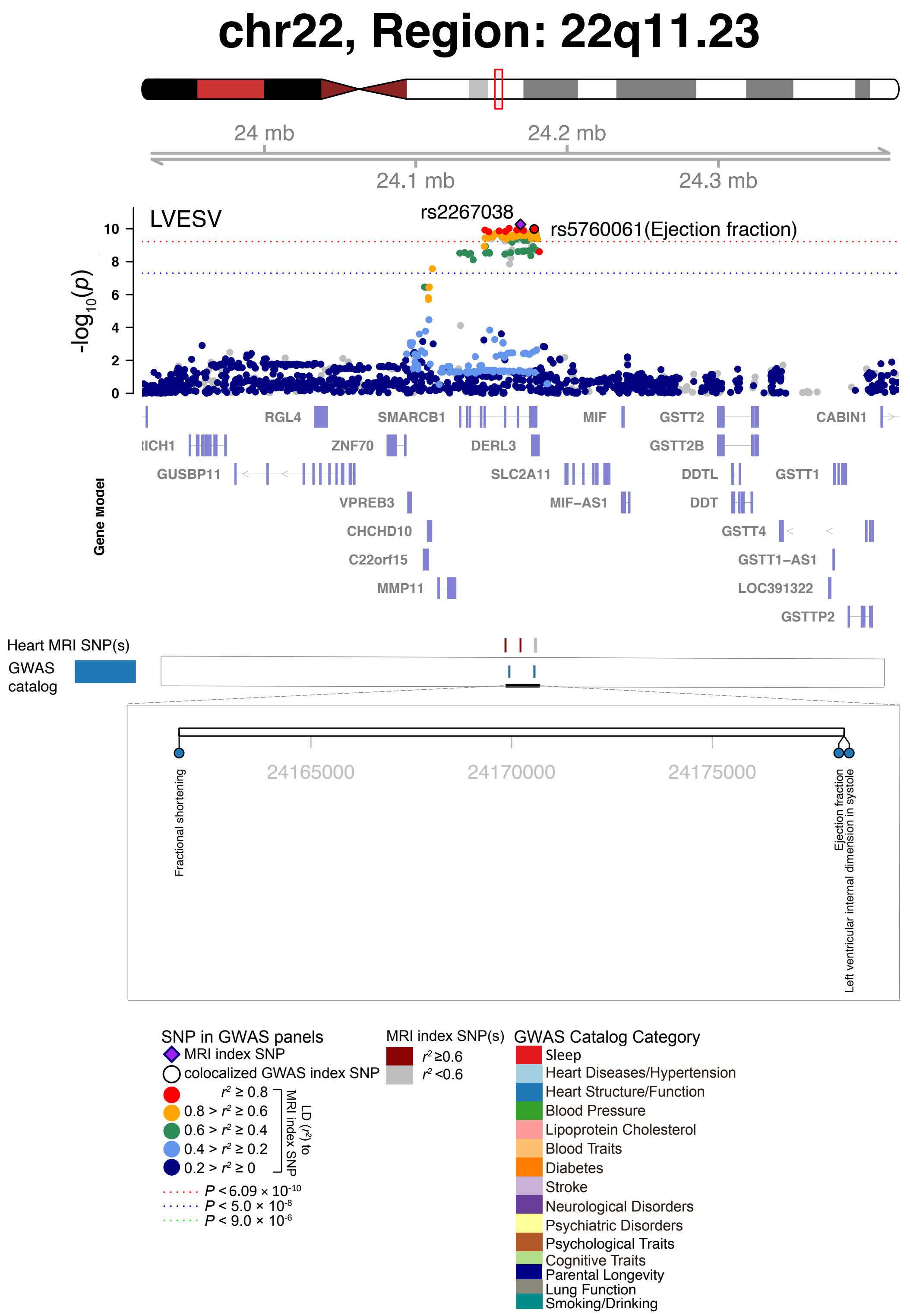


**Fig. S45 Selected genetic loci that were associated with both CMR trait and other complex traits and diseases.** In 22q11.23, we observed the shared association (LD $r^2 \geq 0.6$) between CMR trait (LVESV, index variant rs2267038) and ejection fraction (index variant rs5760061). We also observed shared associations with fractional shortening and left ventricular internal dimension.

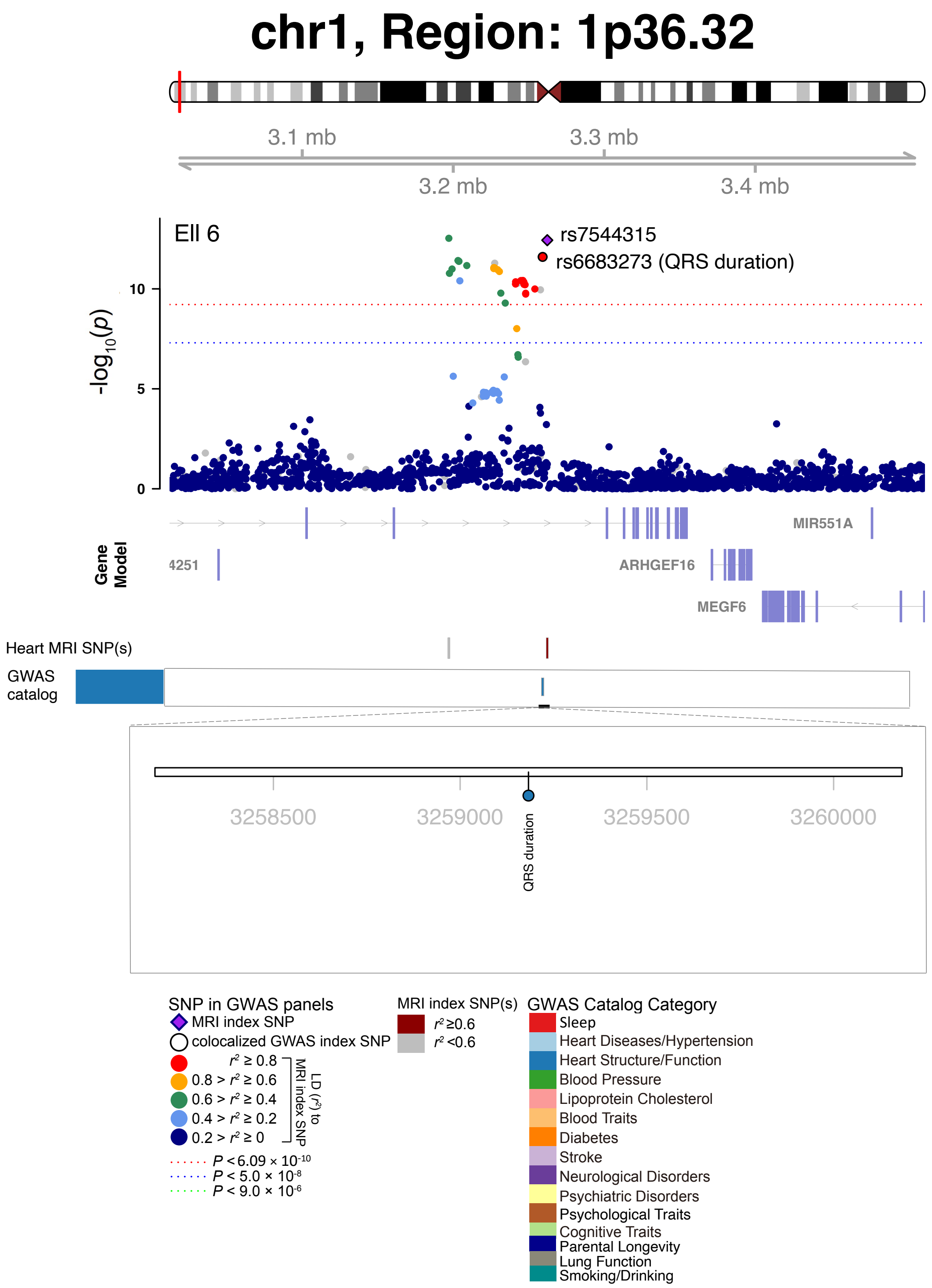


**Fig. S46 Selected genetic loci that were associated with both CMR trait and other complex traits and diseases.** In 1p36.32, we observed the shared association (LD $r^2 \geq 0.6$) between CMR trait (Ell 6, index variant rs7544315) and QRS duration (index variant rs6683273). Ell 6, regional longitudinal strain (region 6).

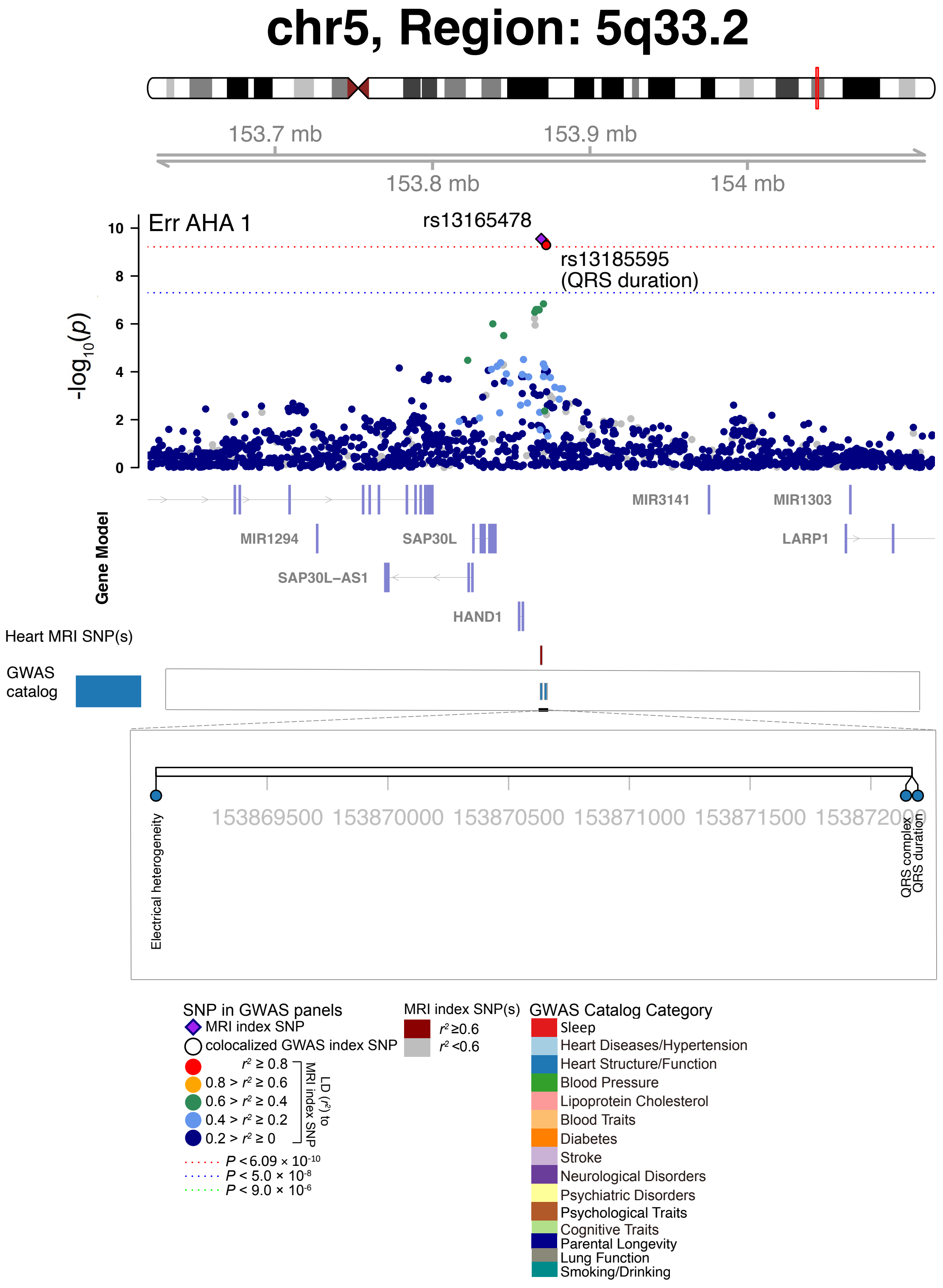


**Fig. S47 Selected genetic loci that were associated with both CMR trait and other complex traits and diseases.** In 5q33.2, we observed the shared association (LD $r^2 \geq 0.6$) between CMR trait (Err AHA 1, index variant rs13165478) and QRS duration (index variant rs13185595). Err AHA 1, regional radial strain (region 1).

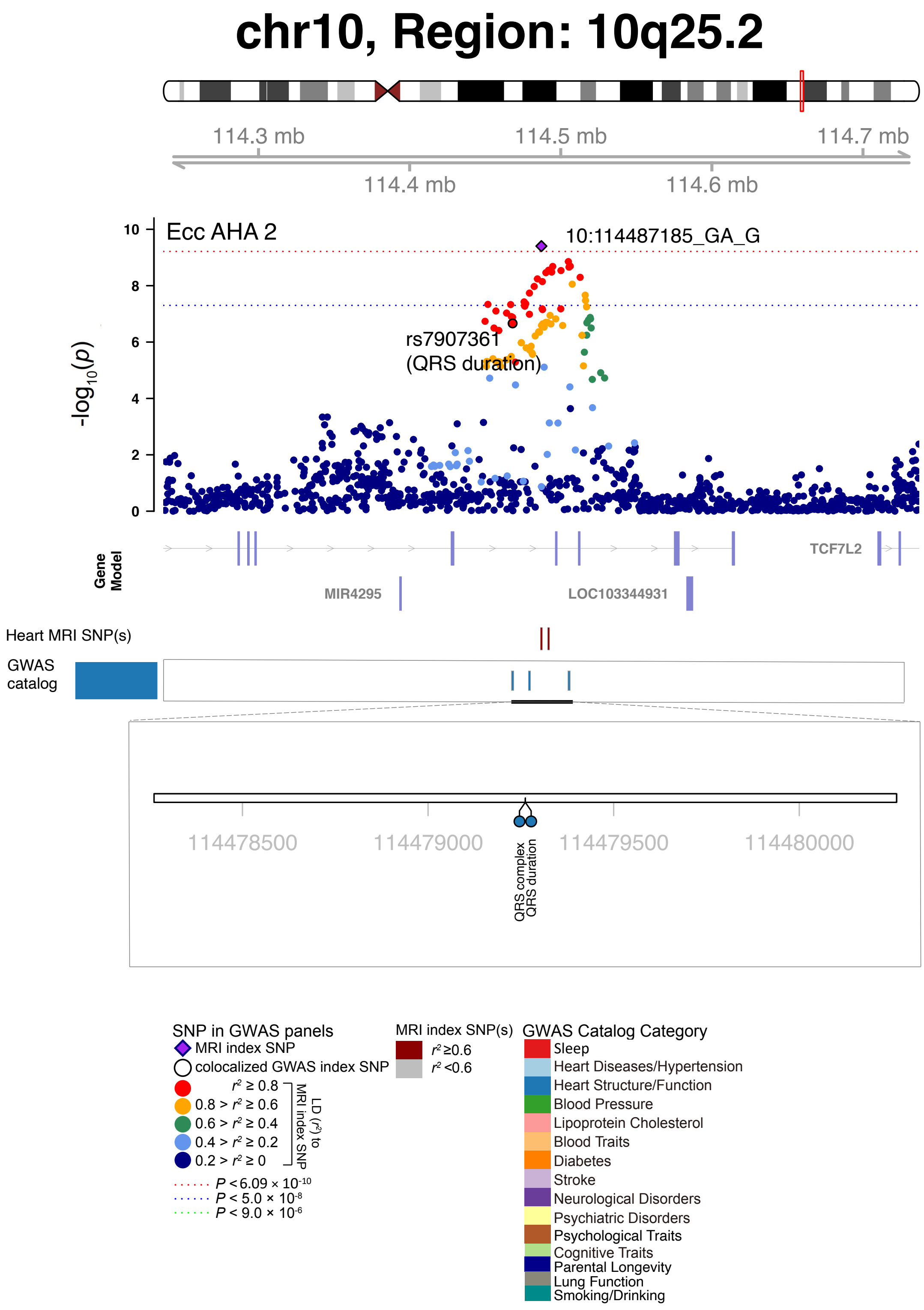


**Fig. S48 Selected genetic loci that were associated with both CMR trait and other complex traits and diseases.** In 10q25.2, we observed the shared association (LD $r^2 \geq 0.6$) between CMR trait (Err AHA 2, index variant 10:114487185_GA_G) and QRS duration (index variant rs7907361). Err AHA 2, regional radial strain (region 2).

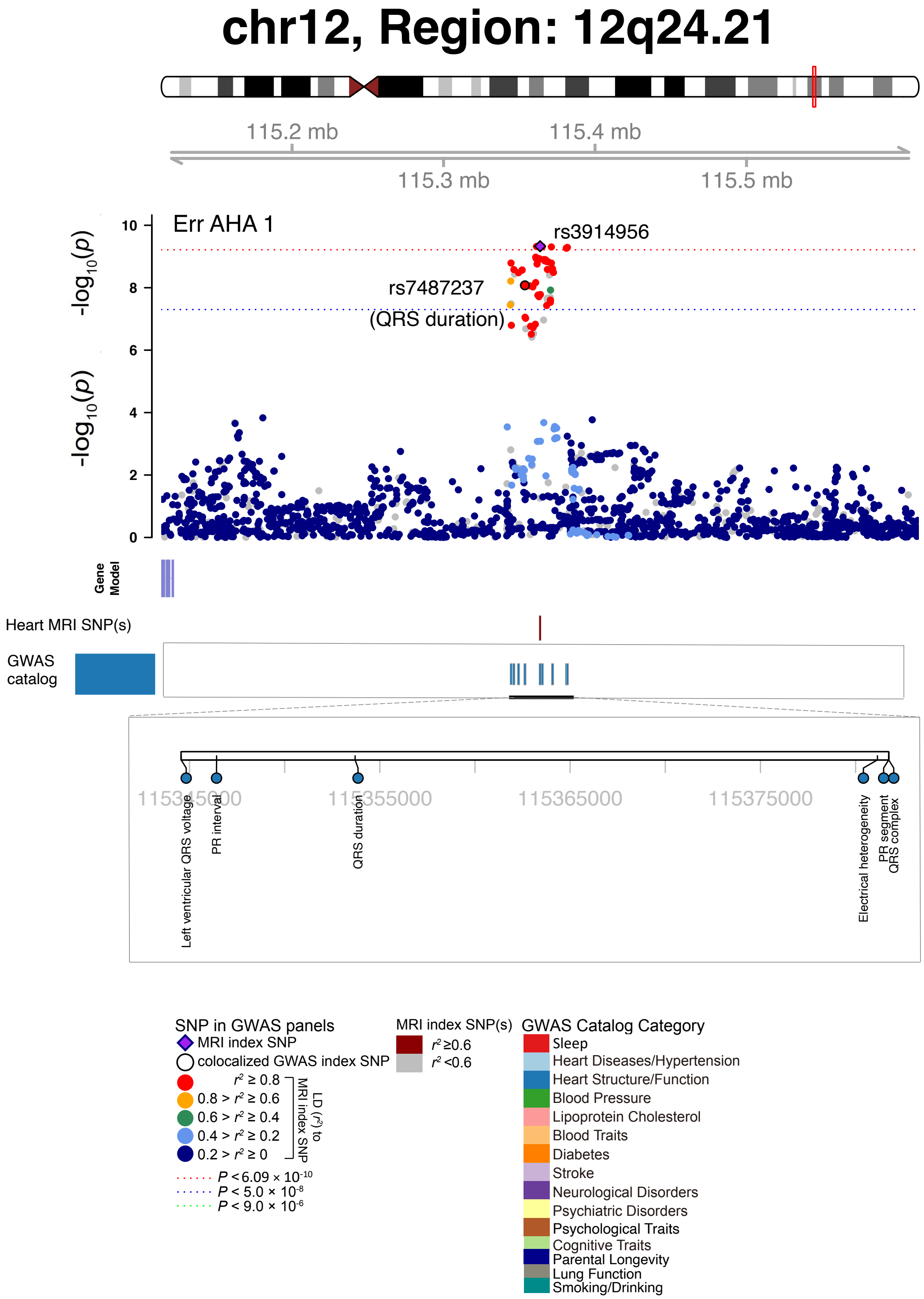


**Fig. S49 Selected genetic loci that were associated with both CMR trait and other complex traits and diseases.** In 12q24.21, we observed the shared association (LD $r^2 \geq 0.6$) between CMR trait (Err AHA 1, index variant rs3914956) and QRS duration (index variant rs7487237). Err AHA 1, regional radial strain (region 1).

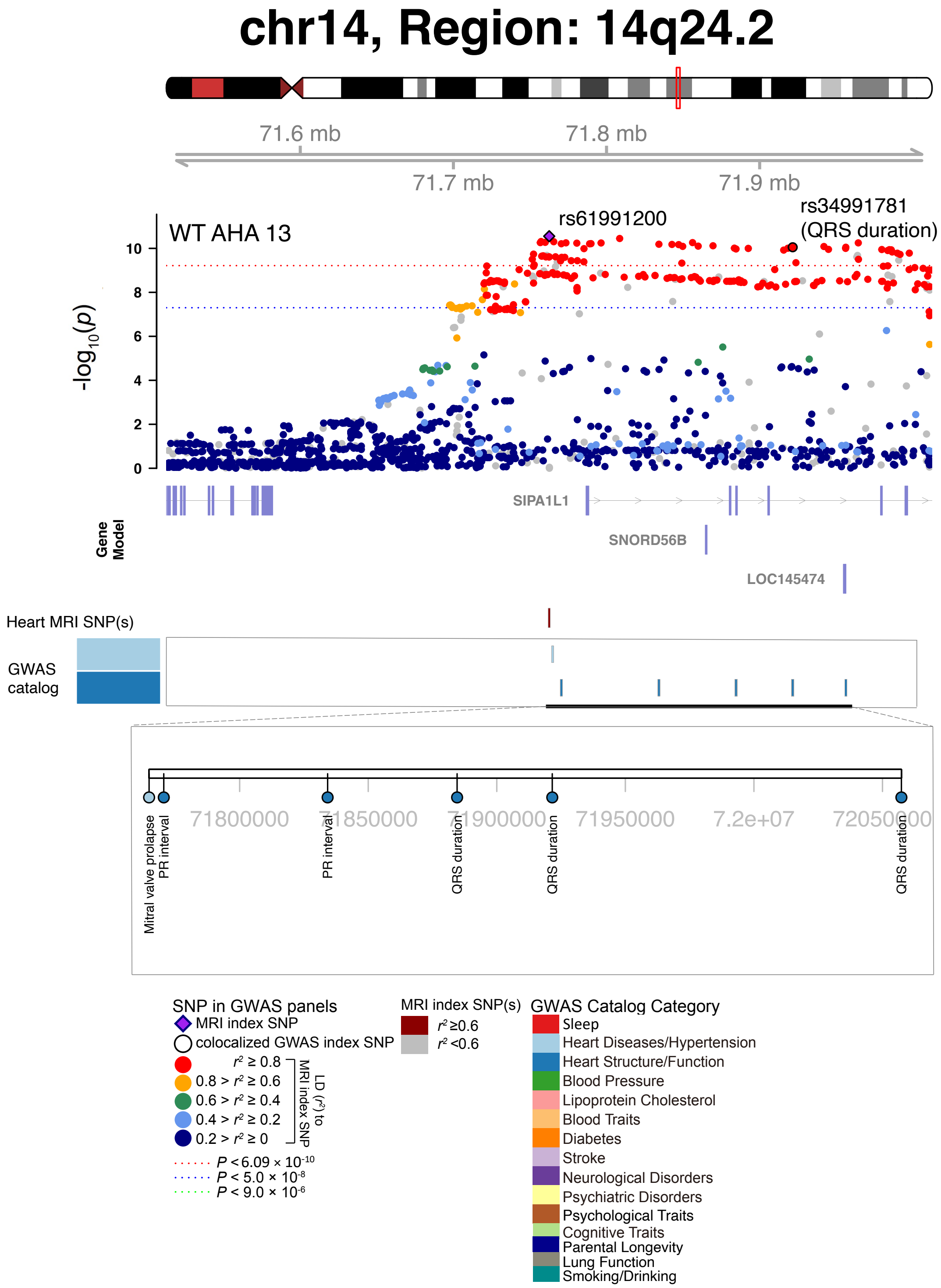


**Fig. S50 Selected genetic loci that were associated with both CMR trait and other complex traits and diseases.** In 14q24.2, we observed the shared association (LD $r^2 \geq 0.6$) between CMR trait (WT AHA 13, index variant rs61991200) and QRS duration (index variant rs34991781). WT AHA 13, regional myocardial-wall thickness at end-diastole (region 13). We also observed the shared association with mitral valve prolapse.

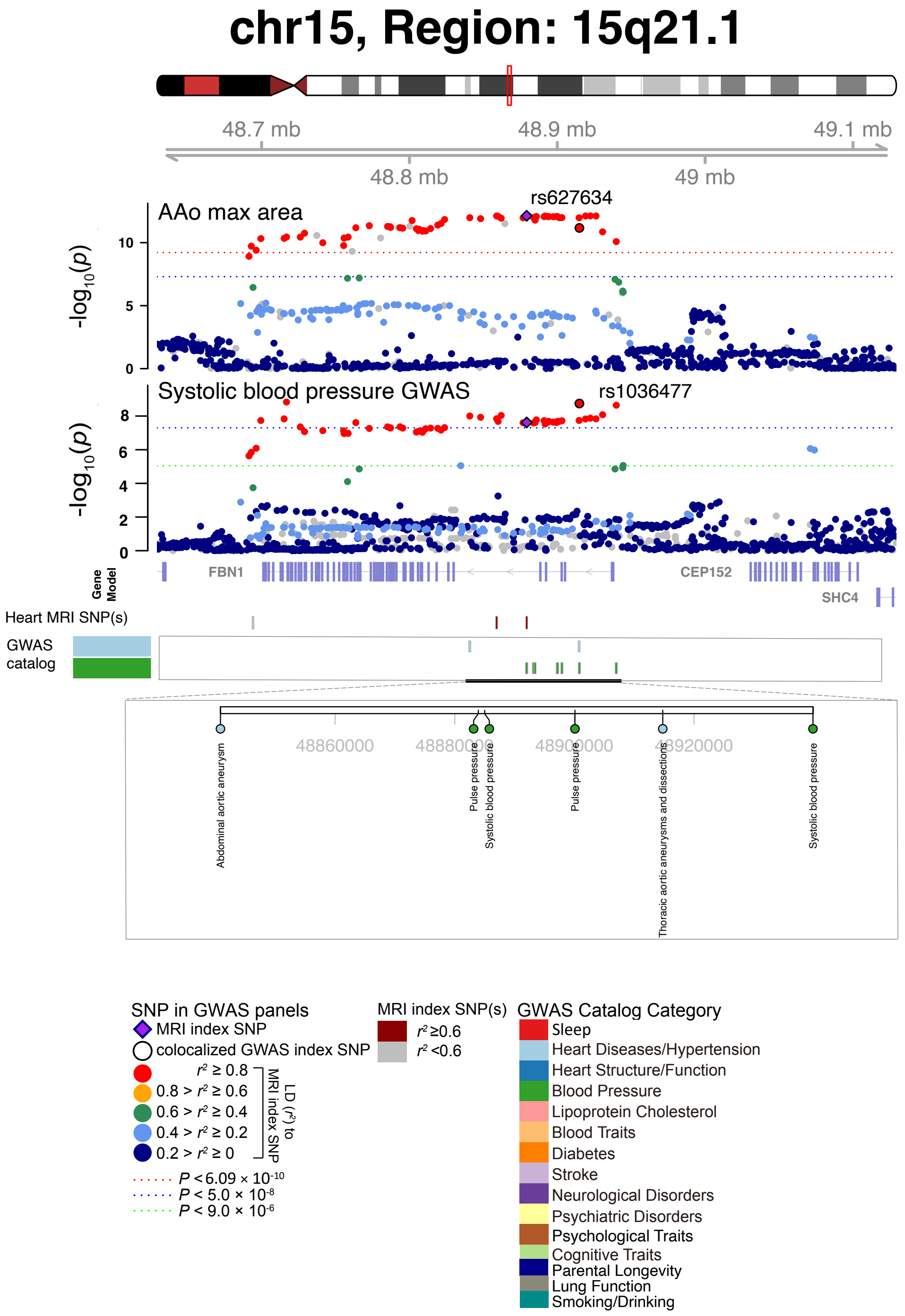


**Fig. S51 Selected genetic loci that were associated with both CMR trait and other complex traits and diseases.** In 15q21.1, we observed the shared association (LD $r^2 \geq 0.6$) between CMR trait (AAo max area, index variant rs627634) and systolic blood pressure (index variant rs1036477). The posterior probability of Bayesian colocalization analysis for the shared causal variant hypothesis (PPH4) is 0.973. AAo max area, ascending aorta maximum area. We also observed shared associations with abdominal aortic aneurysm and thoracic aortic aneurysms and dissections.

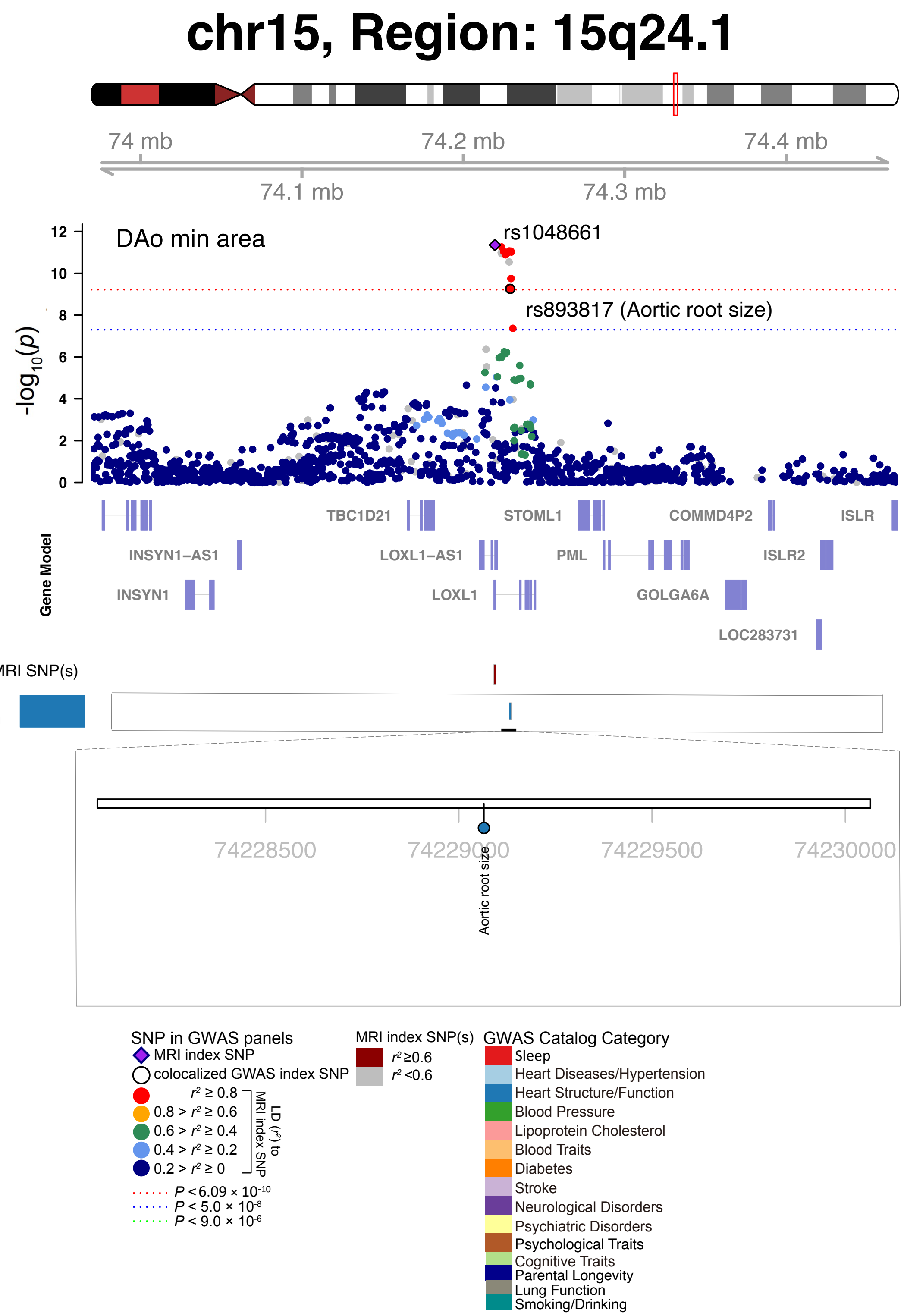


**Fig. S52 Selected genetic loci that were associated with both CMR trait and other complex traits and diseases.** In 15q24.1, we observed the shared association (LD $r^2 \geq 0.6$) between CMR trait (DAo max area, index variant rs1048661) and aortic root size (index variant rs893817). DAo max area, descending aorta maximum area.

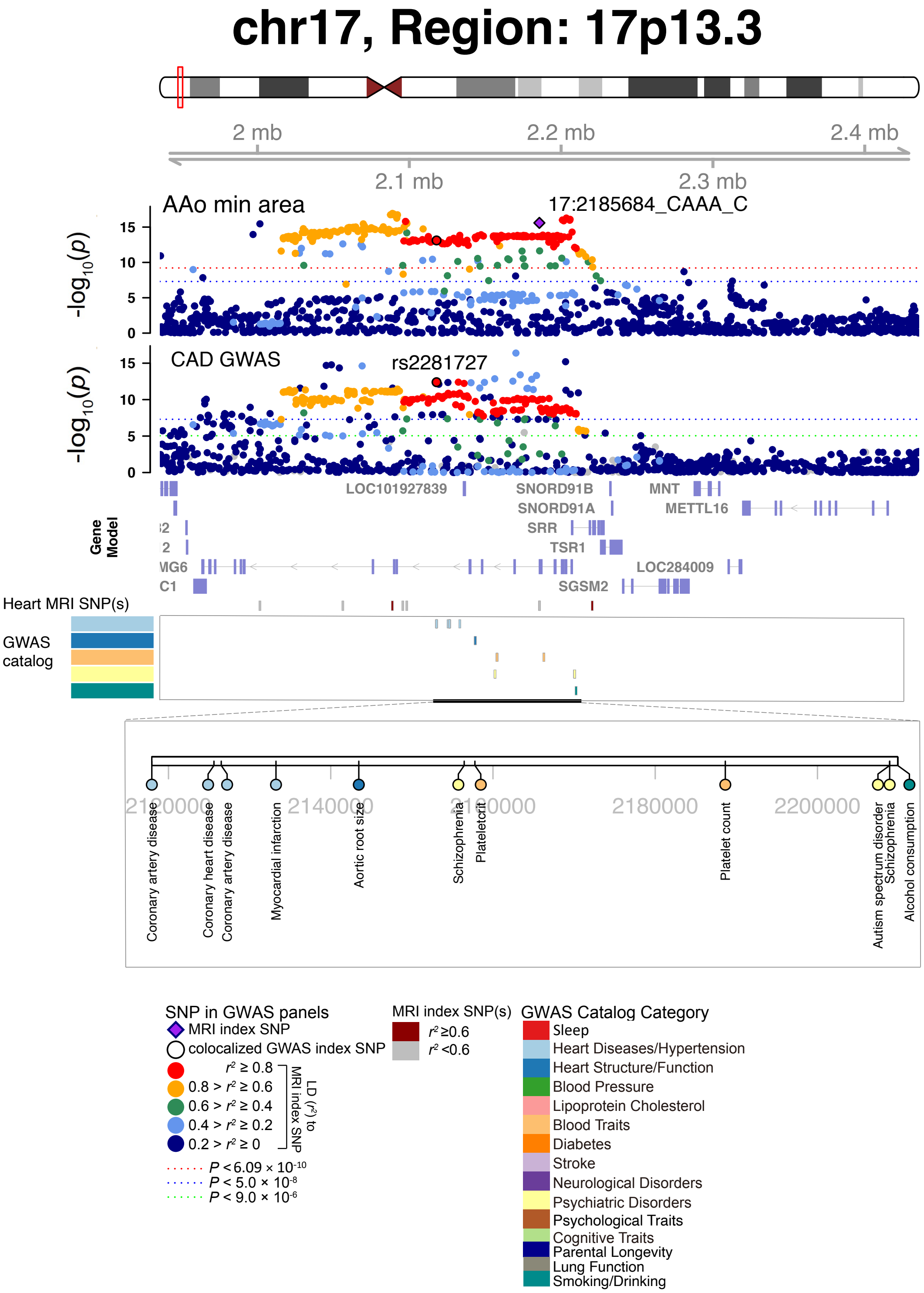


**Fig. S53 Selected genetic loci that were associated with both CMR trait and other complex traits and diseases.** In 17p13.3, we observed the shared association (LD $r^2 \geq 0.6$) between the ascending aorta minimum area (AAo min area, index variant 17:2185684_CAAA_C) and coronary artery disease (CAD, index variant rs2281727). In this region, the AAo min area also had shared associations with schizophrenia and autism spectrum disorder.

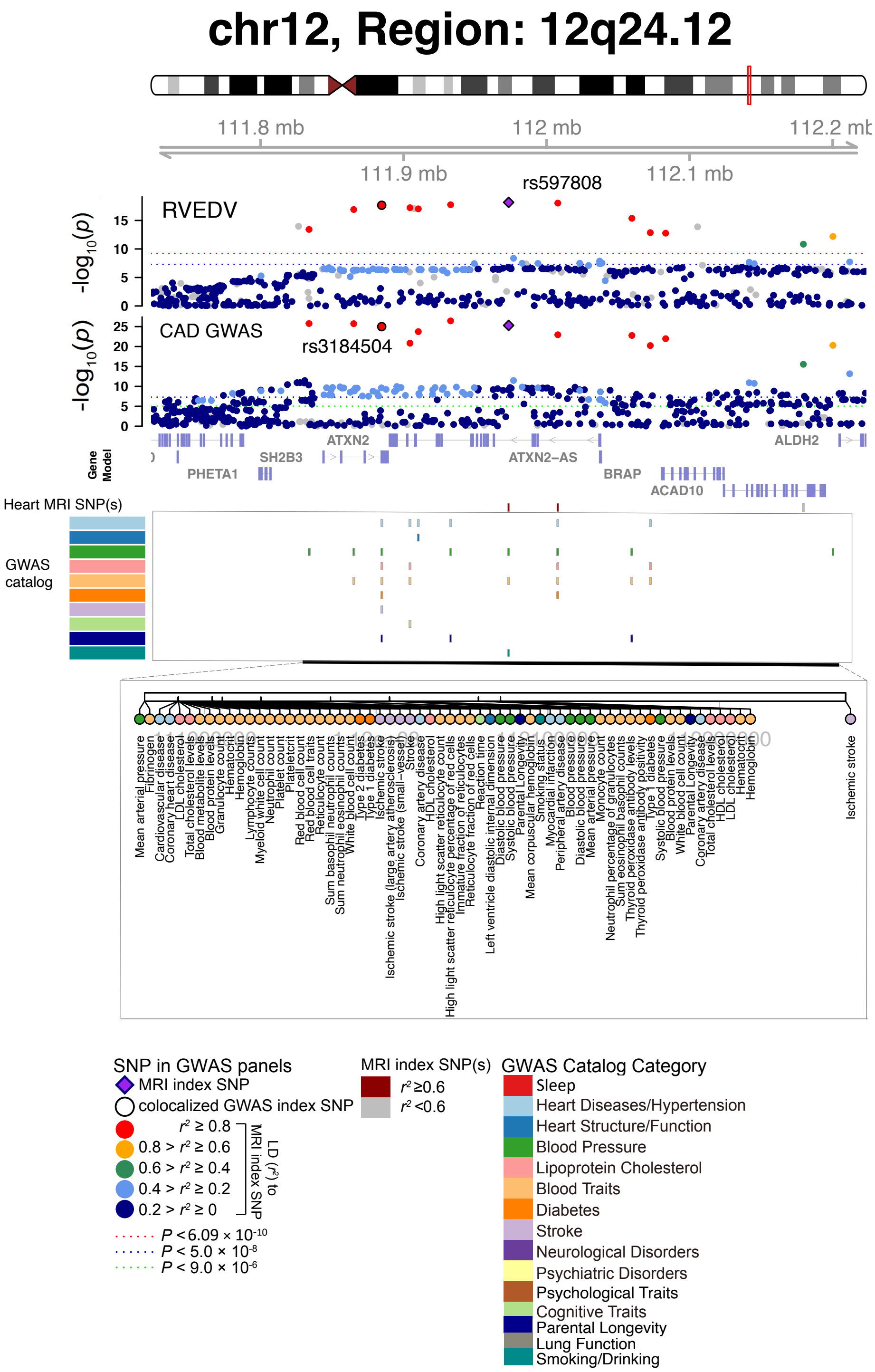


**Fig. S54 Selected genetic loci that were associated with both CMR trait and other complex traits and diseases.** In 12q24.12, we observed the shared association (LD $r^2 \geq 0.6$) between CMR trait (RVEDV, index variant rs597808) and coronary artery disease (CAD, index variant rs3184504). The posterior probability of Bayesian colocalization analysis for the shared causal variant hypothesis (PPH4) is 0.997. We also observed shared associations with many other traits and diseases in different categories.

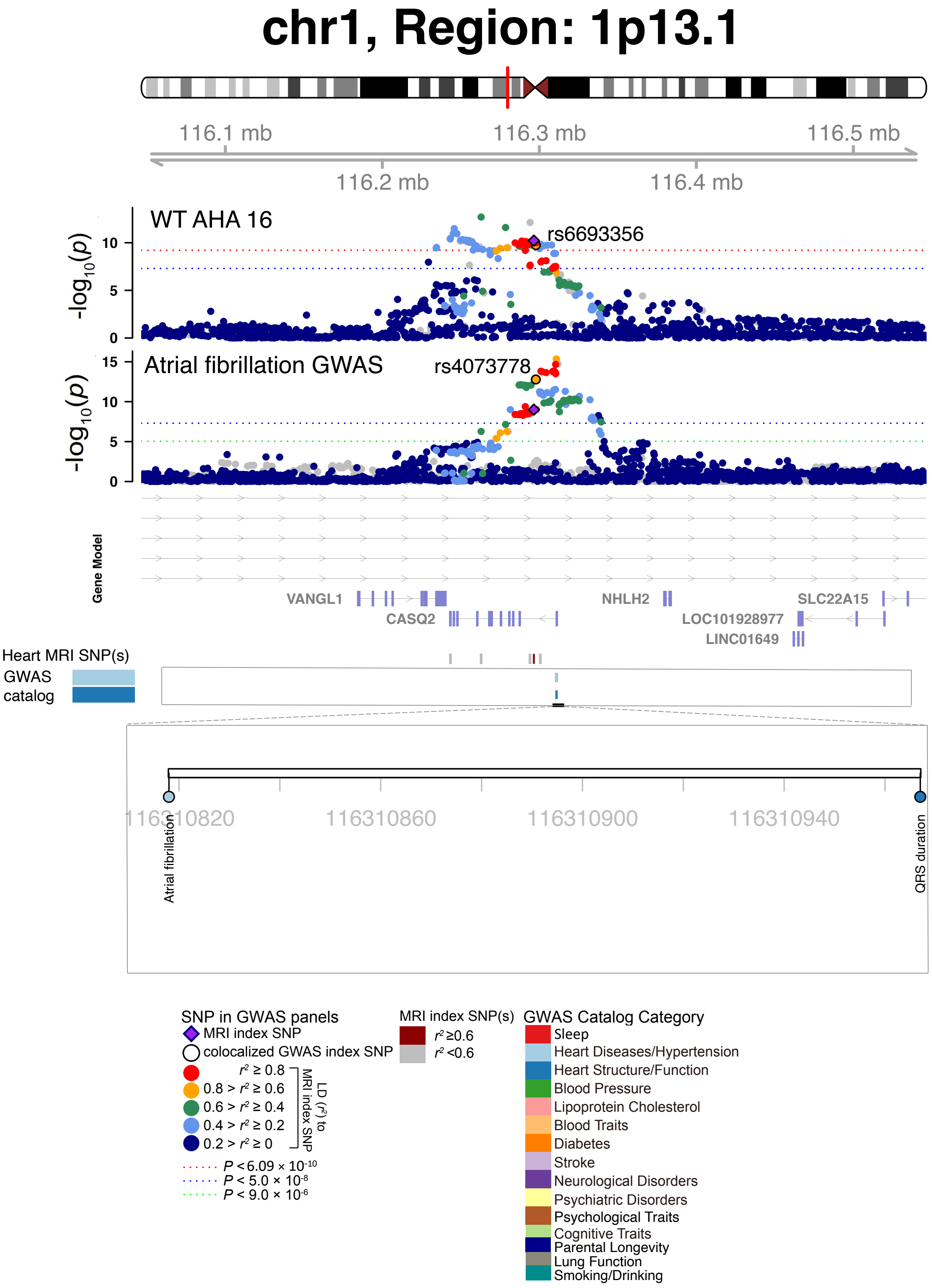


**Fig. S55 Selected genetic loci that were associated with both CMR trait and other complex traits and diseases.** In 1p13.1, we observed the shared association (LD $r^2 \geq 0.6$) between CMR trait (WT AHA 16, index variant rs56693356) and atrial fibrillation (index variant rs4073778). WT AHA 16, regional myocardial-wall thickness at end-diastole (region 16).

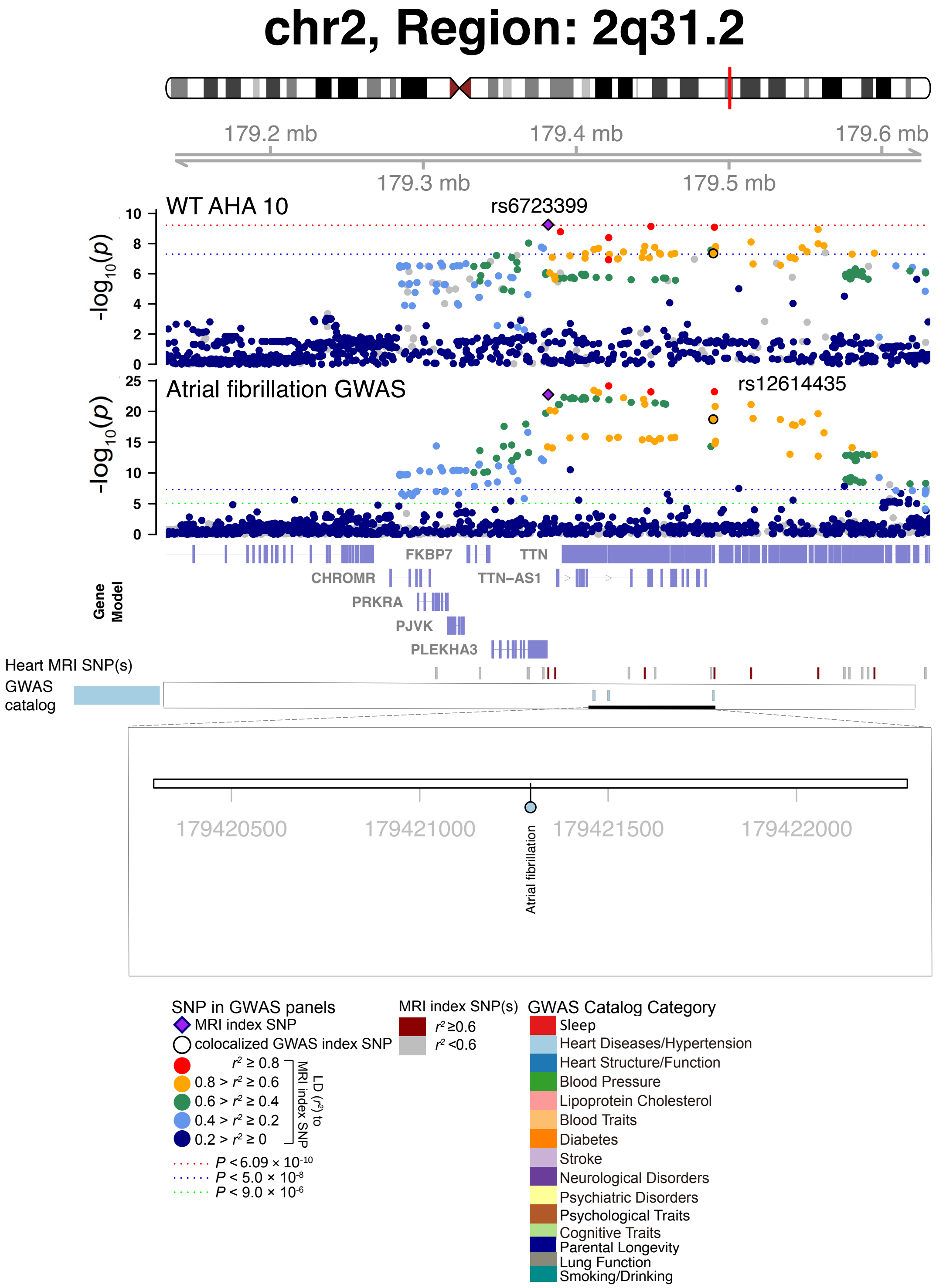


**Fig. S56 Selected genetic loci that were associated with both CMR trait and other complex traits and diseases.** In 2p31.2, we observed the shared association (LD $r^2 \geq 0.6$) between CMR trait (WT AHA 10, index variant rs6723399) and atrial fibrillation (index variant rs12614435). The posterior probability of Bayesian colocalization analysis for the shared causal variant hypothesis (PPH4) is 0.983. WT AHA 10, regional myocardial-wall thickness at end-diastole (region 10).

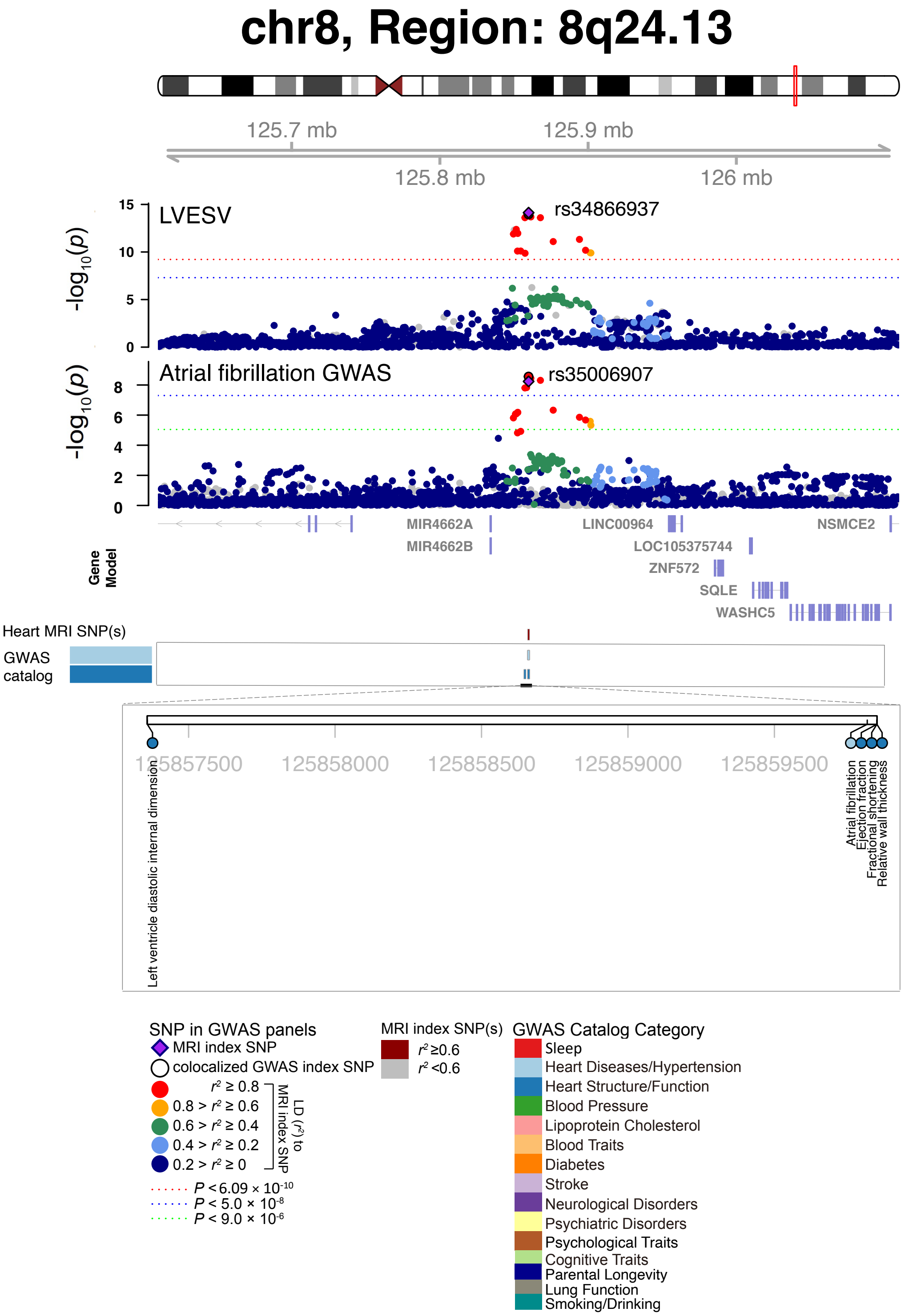


**Fig. S57 Selected genetic loci that were associated with both CMR trait and other complex traits and diseases.** In 8q24.13, we observed the shared association (LD $r^2 \geq 0.6$) between CMR trait (LVESV, index variant rs34866937) and atrial fibrillation (index variant rs35006907). The posterior probability of Bayesian colocalization analysis for the shared causal variant hypothesis (PPH4) is 0.994.

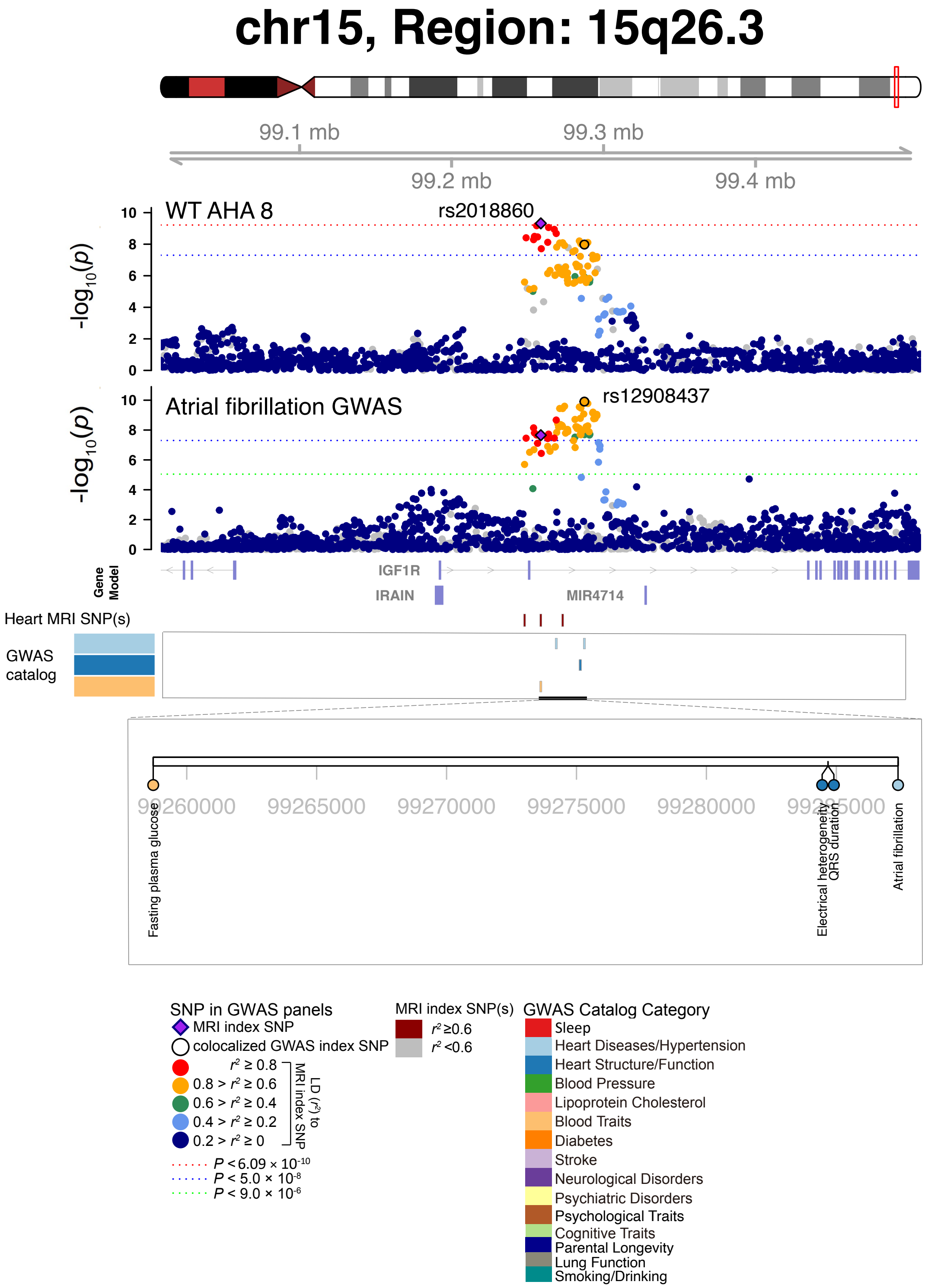


**Fig. S58 Selected genetic loci that were associated with both CMR trait and other complex traits and diseases.** In 15q26.3, we observed the shared association (LD $r^2 \geq 0.6$) between CMR trait (WT AHA 8, index variant rs2018860) and atrial fibrillation (index variant rs12908437). The posterior probability of Bayesian colocalization analysis for the shared causal variant hypothesis (PPH4) is 0.949. WT AHA 8, regional myocardial-wall thickness at end-diastole (region 8).

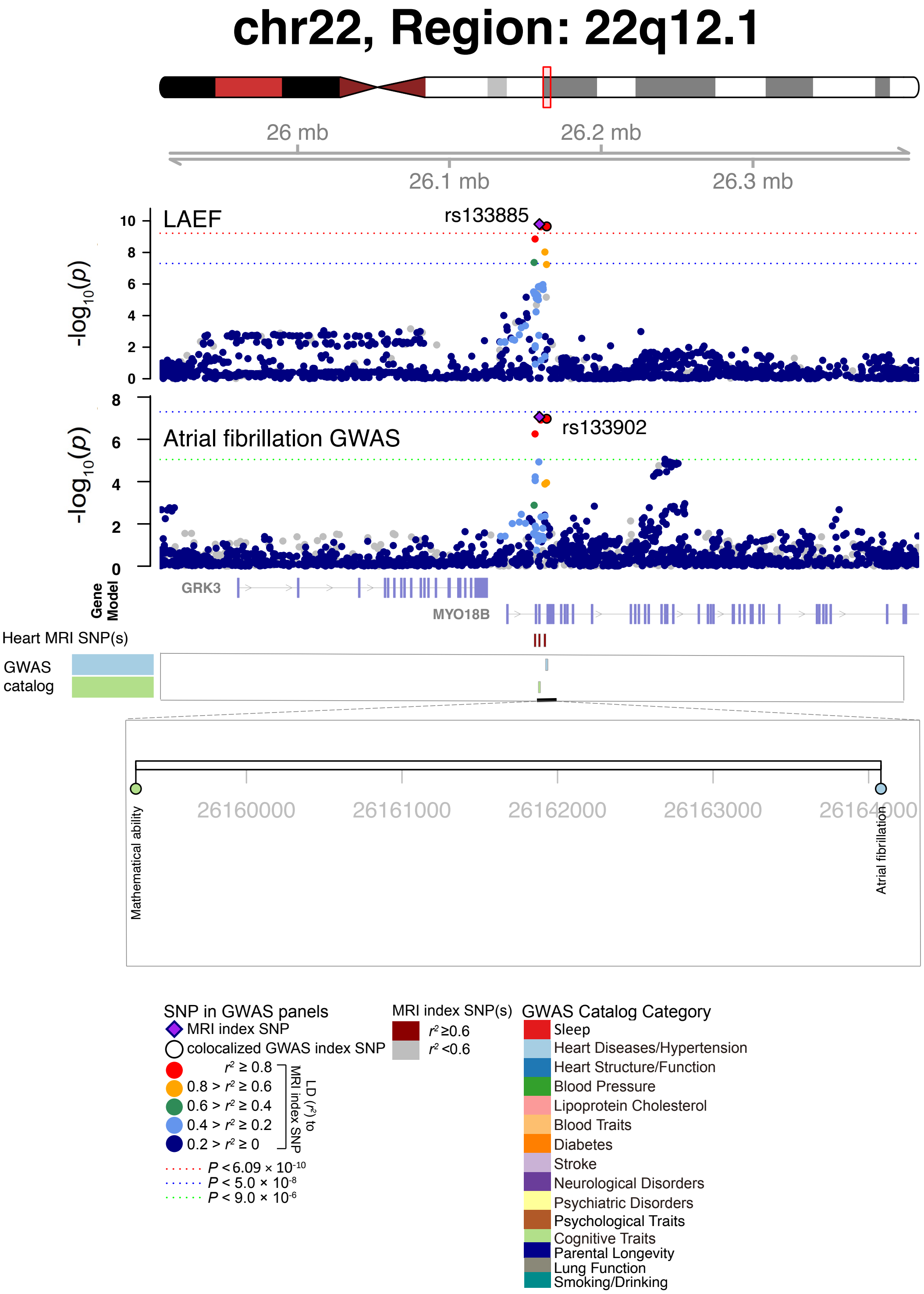


**Fig. S59 Selected genetic loci that were associated with both CMR trait and other complex traits and diseases.** In 22q12.1, we observed the shared association (LD $r^2 \geq 0.6$) between CMR trait (LAEF, index variant rs133885) and atrial fibrillation (index variant rs133902). The posterior probability of Bayesian colocalization analysis for the shared causal variant hypothesis (PPH4) is 0.996.

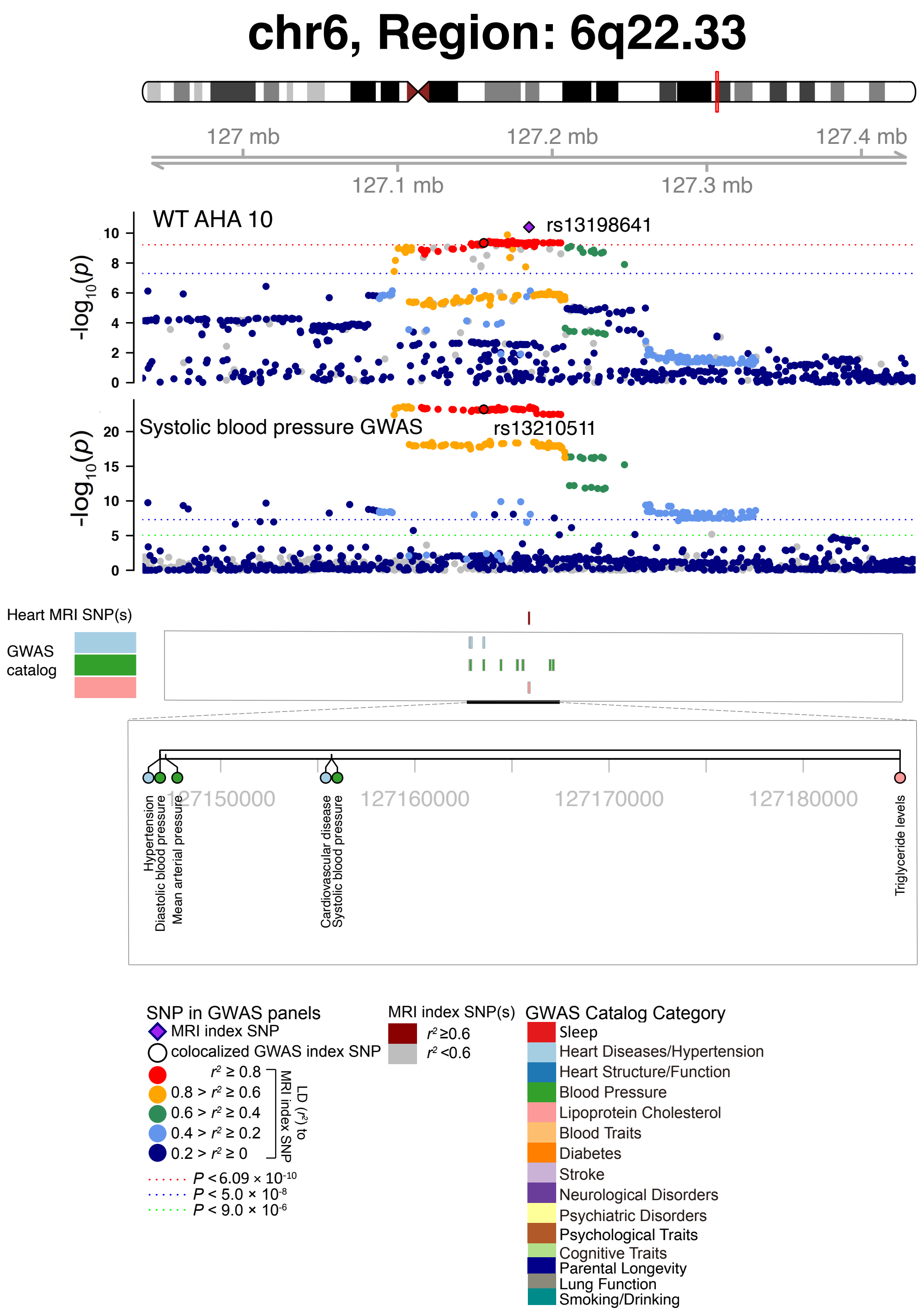


**Fig. S60 Selected genetic loci that were associated with both CMR trait and other complex traits and diseases.** In 6q22.33, we observed the shared association (LD $r^2 \geq 0.6$) between CMR trait (WT AHA 10, index variant rs13198641) and systolic blood pressure (index variant rs13210511). The posterior probability of Bayesian colocalization analysis for the shared causal variant hypothesis (PPH4) is 0.849. WT AHA 10, regional myocardial-wall thickness at end-diastole (region 10). We also observed the shared association with hypertension.

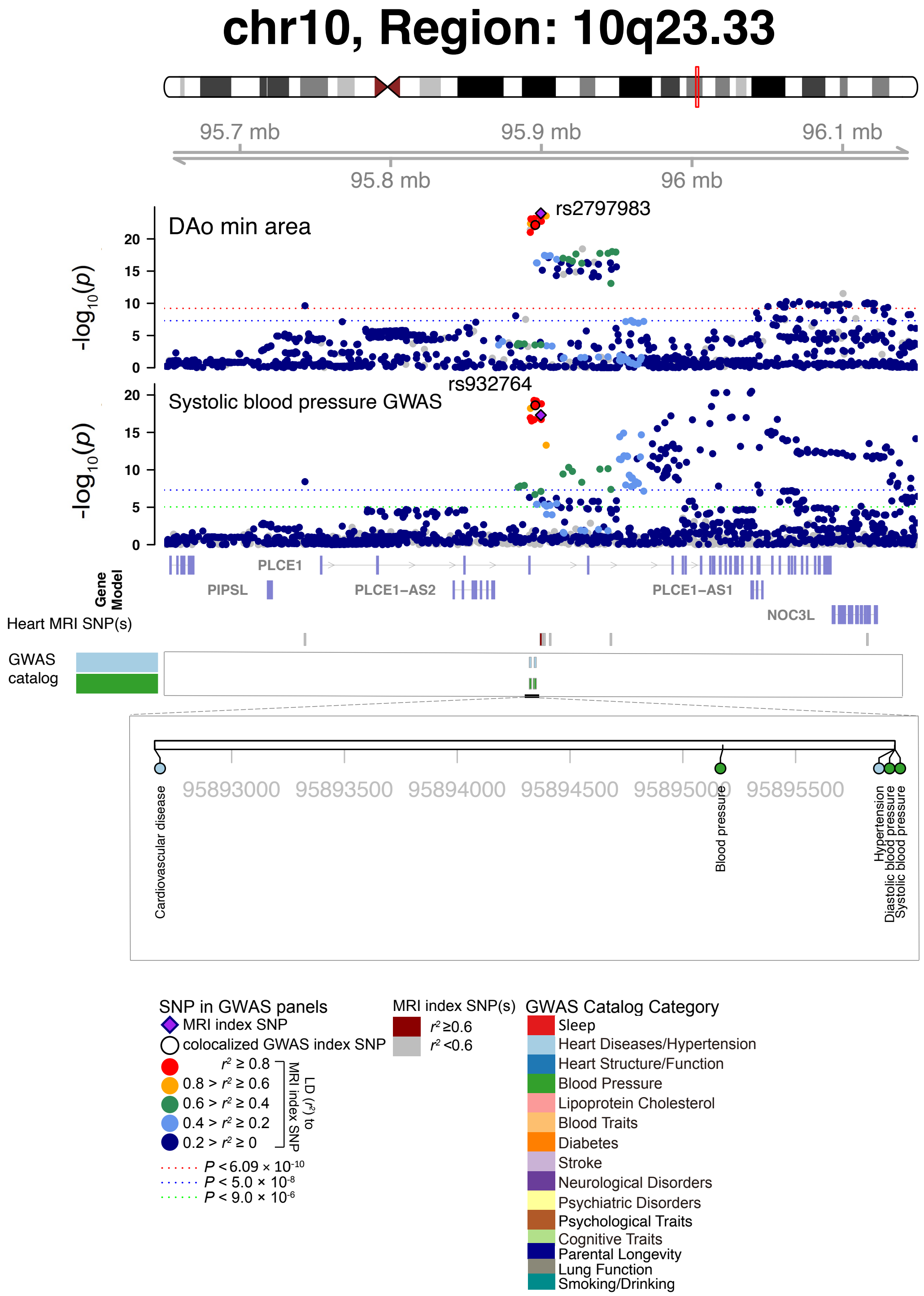


**Fig. S61 Selected genetic loci that were associated with both CMR trait and other complex traits and diseases.** In 10q23.33, we observed the shared association (LD $r^2 \geq 0.6$) between CMR trait (DAo min area, index variant rs2797983) and systolic blood pressure (index variant rs932764). The posterior probability of Bayesian colocalization analysis for the shared causal variant hypothesis (PPH4) is 0.989. DAo min area, descending aorta minimum area. We also observed the shared association with hypertension.

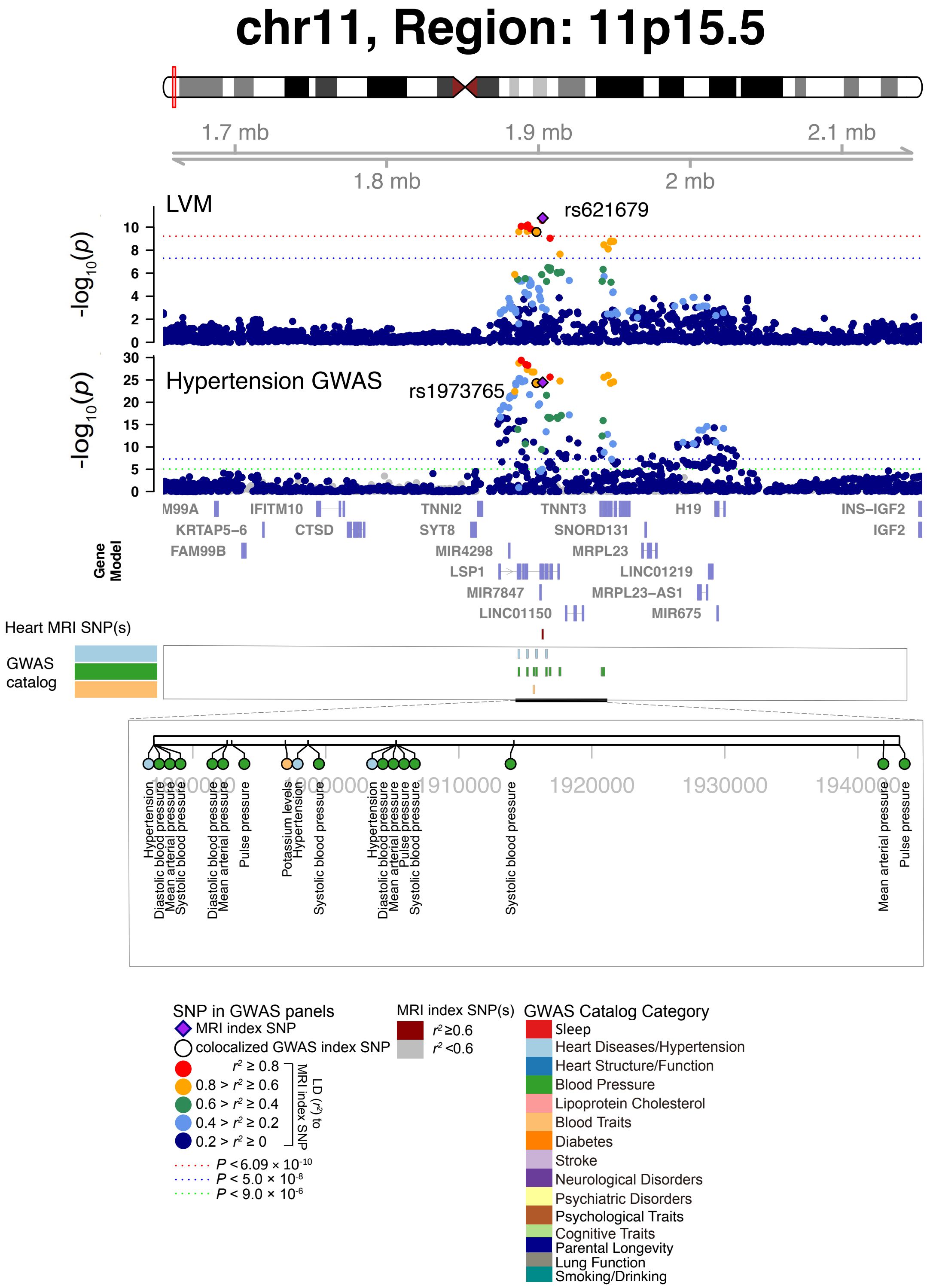


**Fig. S62 Selected genetic loci that were associated with both CMR trait and other complex traits and diseases.** In 11p15.5, we observed the shared association (LD $r^2 \geq 0.6$) between CMR trait (LVM, index variant rs621679) and hypertension (index variant rs1973765). The posterior probability of Bayesian colocalization analysis for the shared causal variant hypothesis (PPH4) is 0.987.

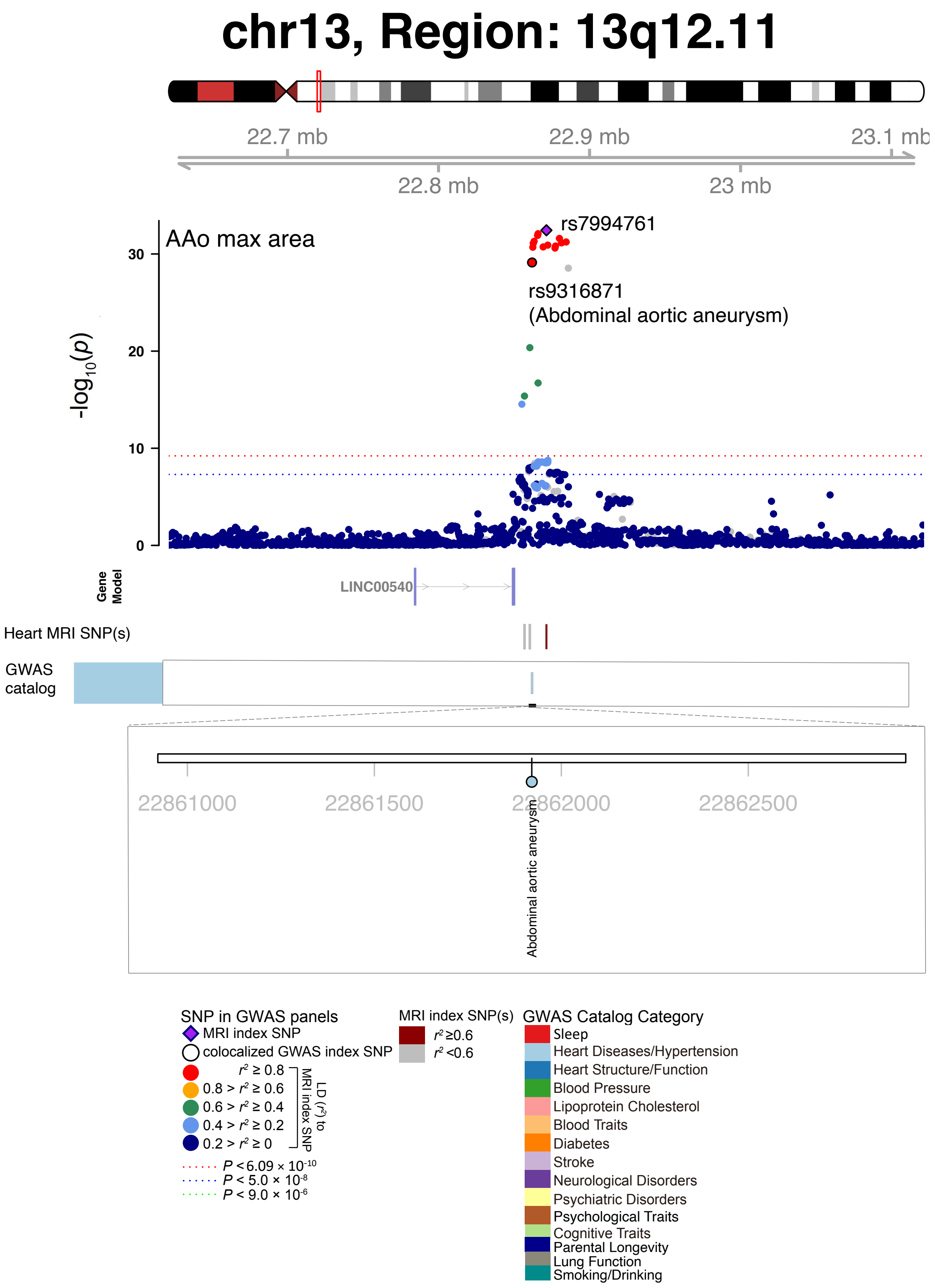


**Fig. S63 Selected genetic loci that were associated with both CMR trait and other complex traits and diseases.** In 13q12.11, we observed the shared association (LD $r^2 \geq 0.6$) between CMR trait (AAo max area, index variant rs7994761) and abdominal aortic aneurysm (index variant rs9316871). AAo max area, ascending aorta maximum area.

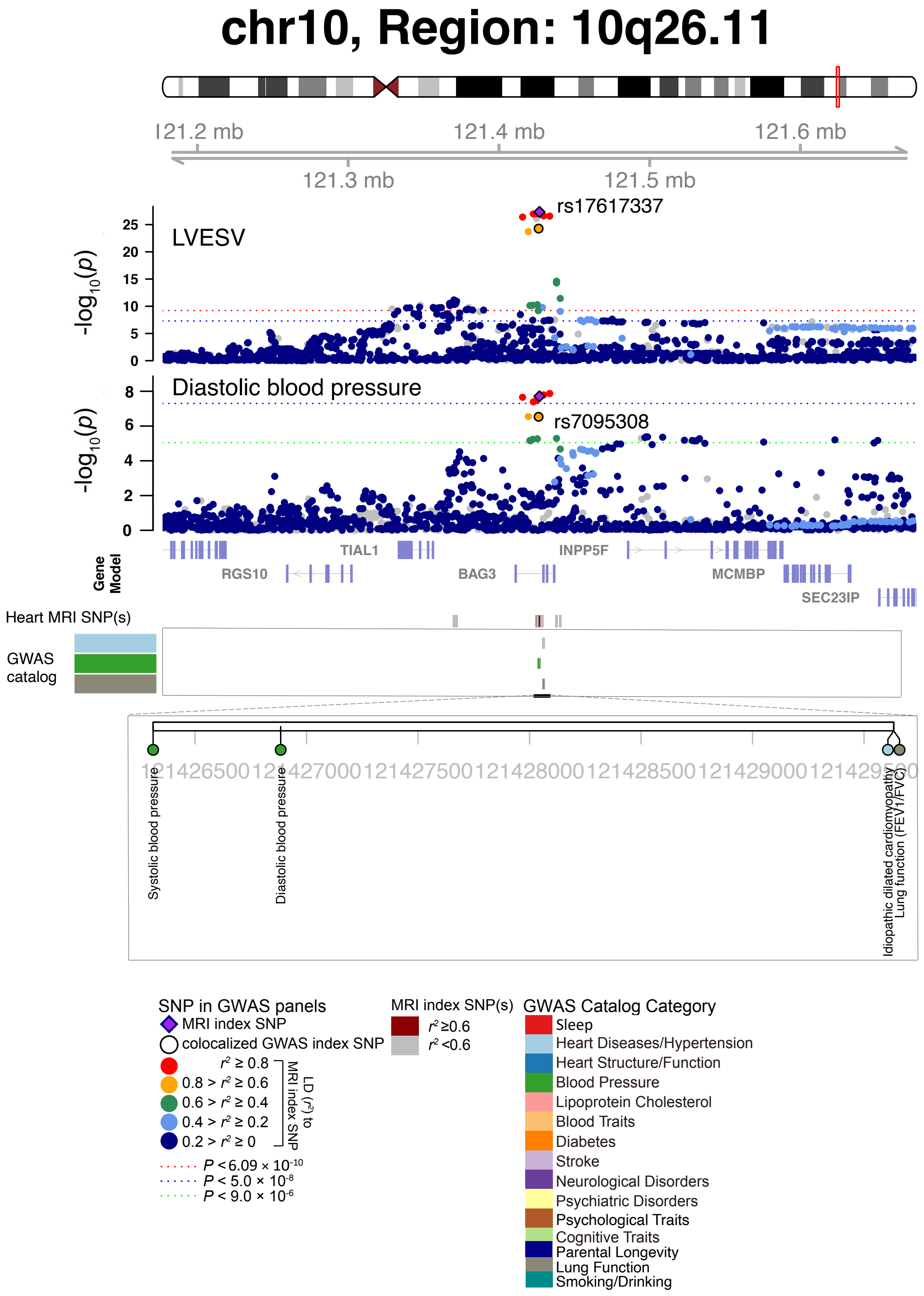


**Fig. S64 Selected genetic loci that were associated with both CMR trait and other complex traits and diseases.** In 10q26.11, we observed the shared association (LD $r^2 \geq 0.6$) between CMR trait (LVESV, index variant rs17617337) and diastolic blood pressure (index variant rs7095308). The posterior probability of Bayesian colocalization analysis for the shared causal variant hypothesis (PPH4) is 0.995. We also observed shared associations with idiopathic dilated cardiomyopathy and lung function.

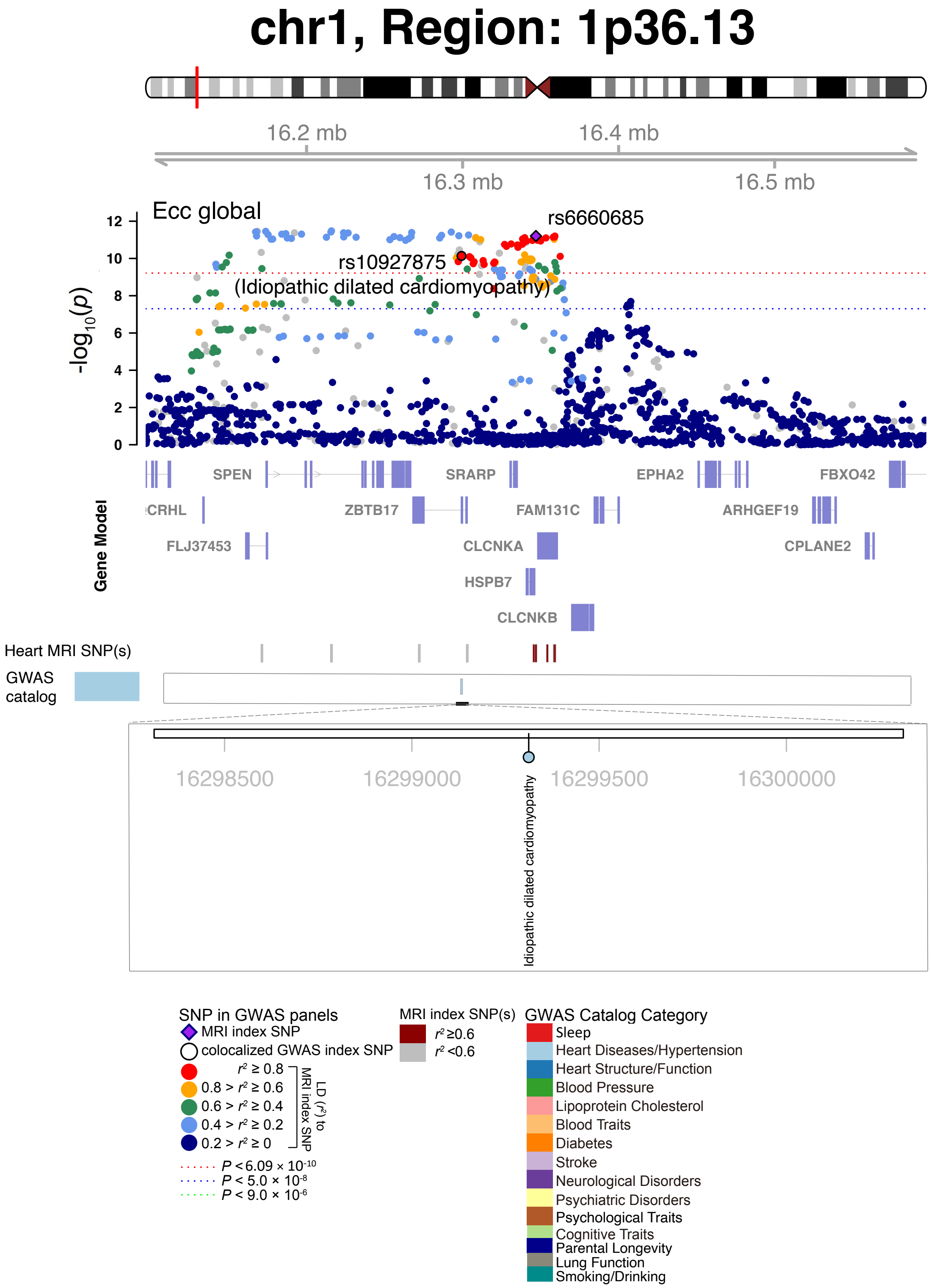


**Fig. S65 Selected genetic loci that were associated with both CMR trait and other complex traits and diseases.** In 1p36.13, we observed the shared association (LD $r^2 \geq 0.6$) between CMR trait (Ecc global, index variant rs6660685) and idiopathic dilated cardiomyopathy (index variant rs10927875). Ecc global, global peak circumferential strain.

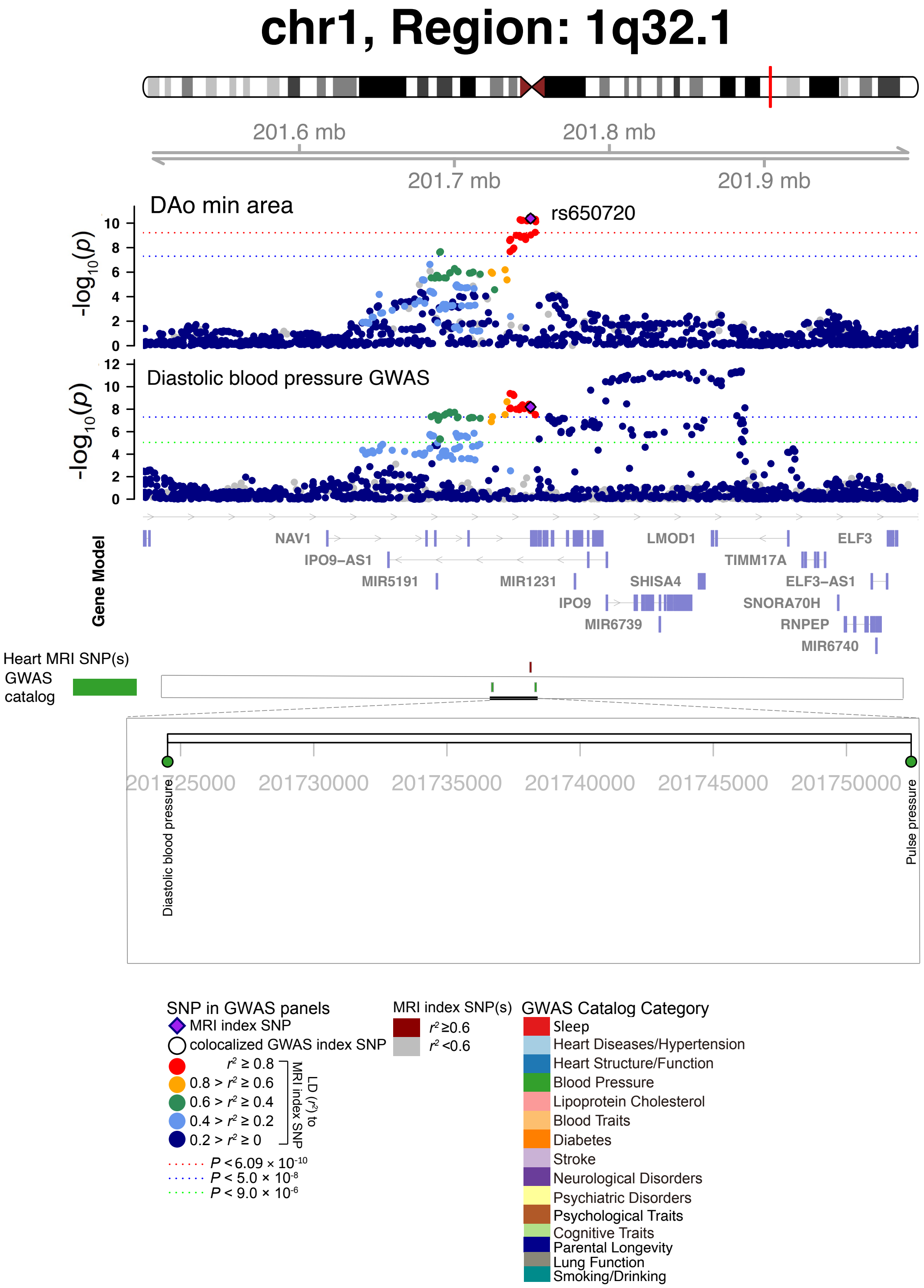


**Fig. S66 Selected genetic loci that were associated with both CMR trait and other complex traits and diseases.** In 1q32.1, we observed the shared association (LD $r^2 \geq 0.6$) between CMR trait (DAo min area) and diastolic blood pressure (shared index variant rs650720). The posterior probability of Bayesian colocalization analysis for the shared causal variant hypothesis (PPH4) is 0.985. DAo min area, descending aorta minimum area.

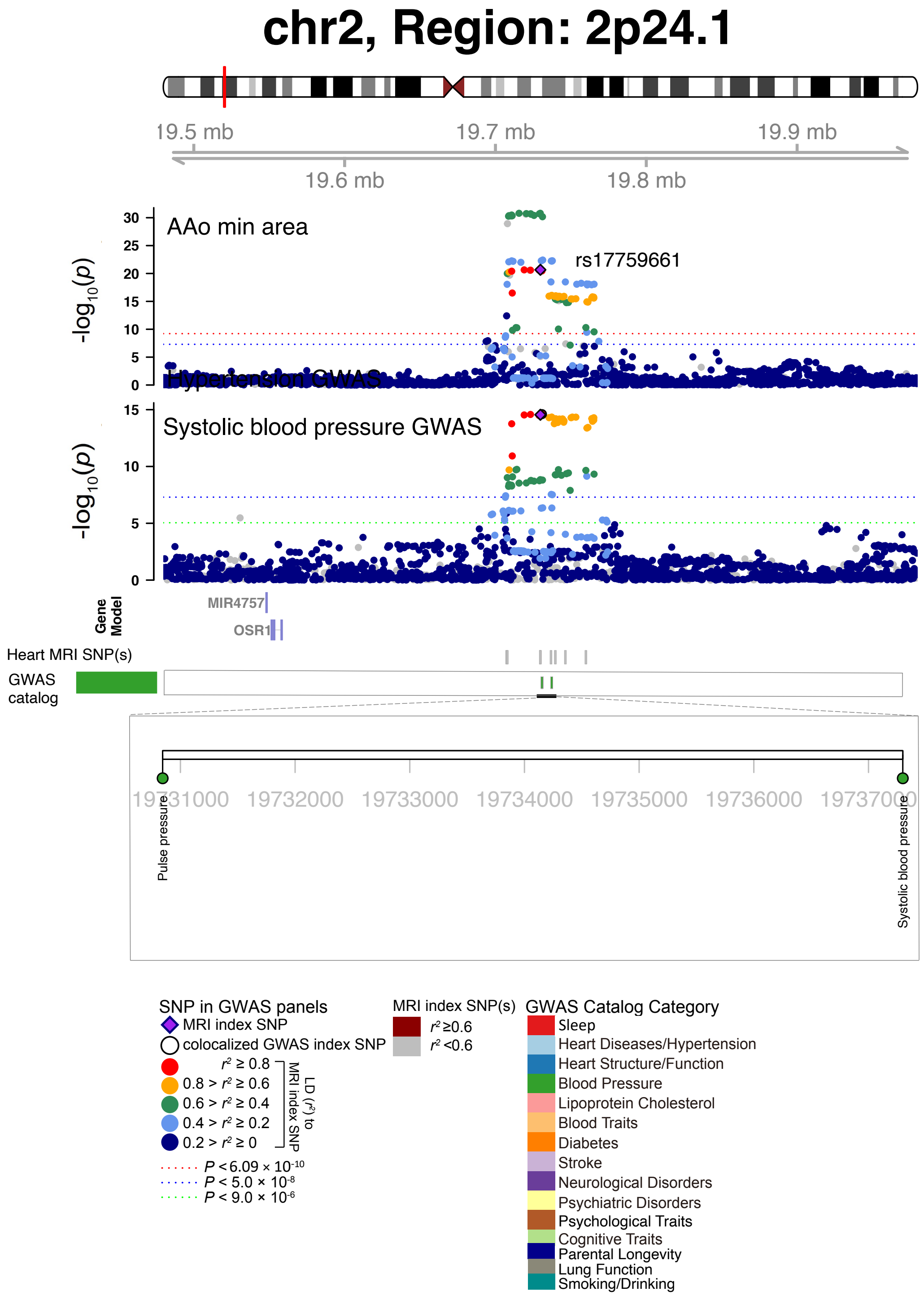


**Fig. S67 Selected genetic loci that were associated with both CMR trait and other complex traits and diseases.** In 2p24.1, we observed the shared association (LD $r^2 \geq 0.6$) between CMR trait (AAo min area) and systolic blood pressure (shared index variant rs17759661). The posterior probability of Bayesian colocalization analysis for the shared causal variant hypothesis (PPH4) is 0.956. AAo min area, ascending aorta minimum area.

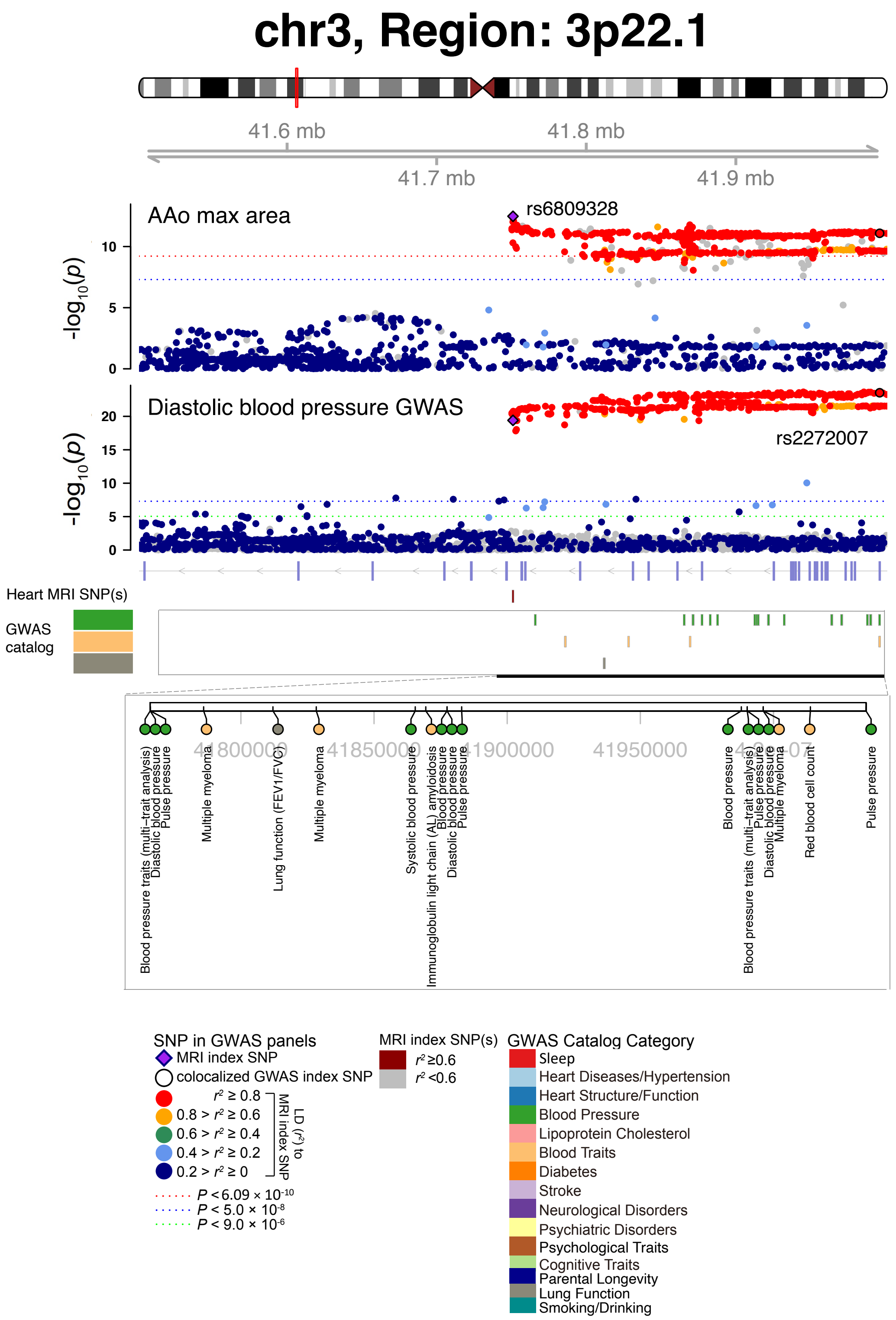


**Fig. S68 Selected genetic loci that were associated with both CMR trait and other complex traits and diseases.** In 3p22.1, we observed the shared association (LD $r^2 \geq 0.6$) between CMR trait (AAo max area, index variant rs6809328) and diastolic blood pressure (index variant rs2272007). AAo max area, ascending aorta maximum area.

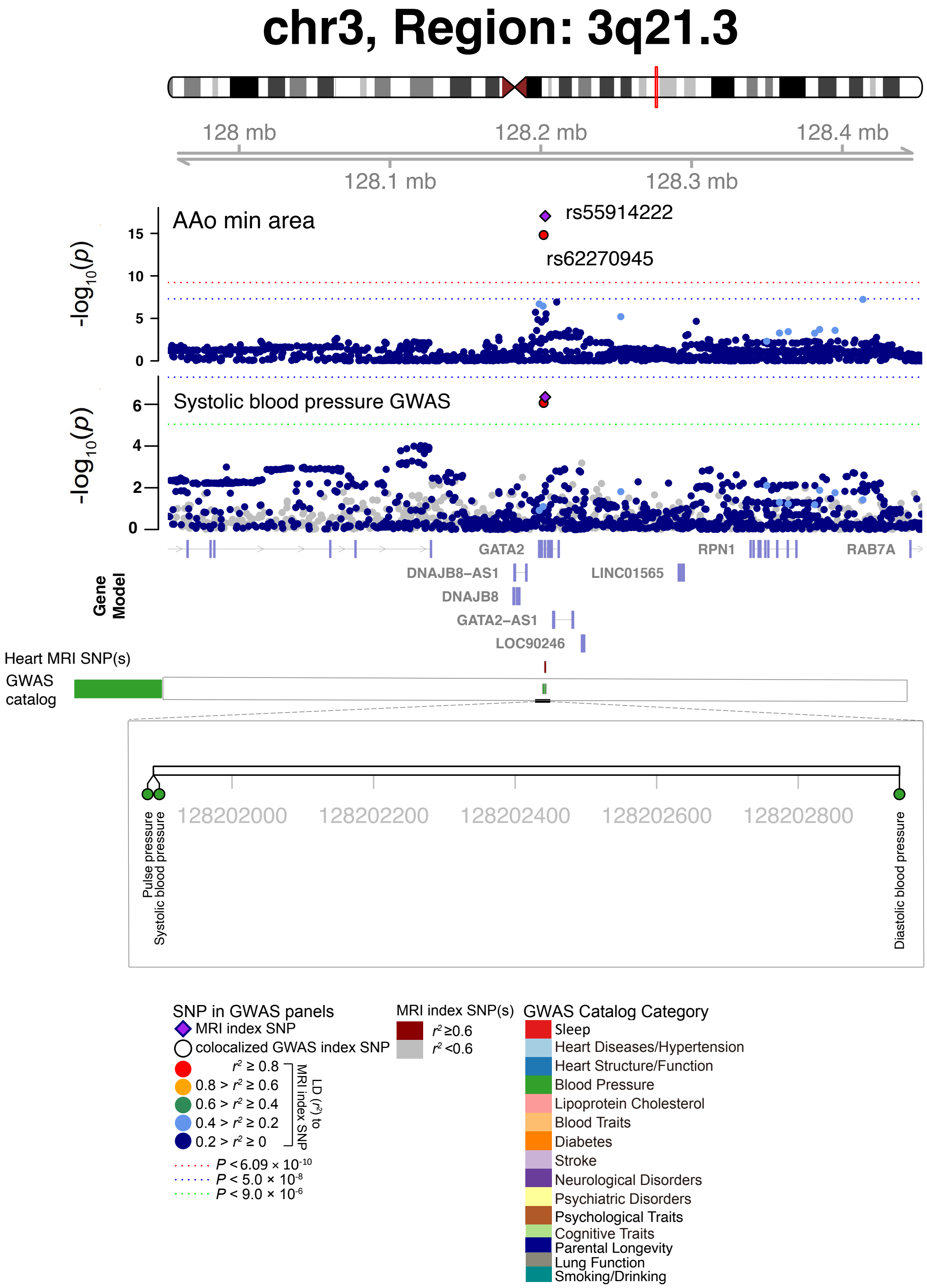


**Fig. S69 Selected genetic loci that were associated with both CMR trait and other complex traits and diseases.** In 3q21.3, we observed the shared association (LD $r^2 \geq 0.6$) between CMR trait (AAo min area, index variant rs55914222) and systolic blood pressure (index variant rs62270945). The posterior probability of Bayesian colocalization analysis for the shared causal variant hypothesis (PPH4) is 0.999. AAo min area, ascending aorta minimum area.

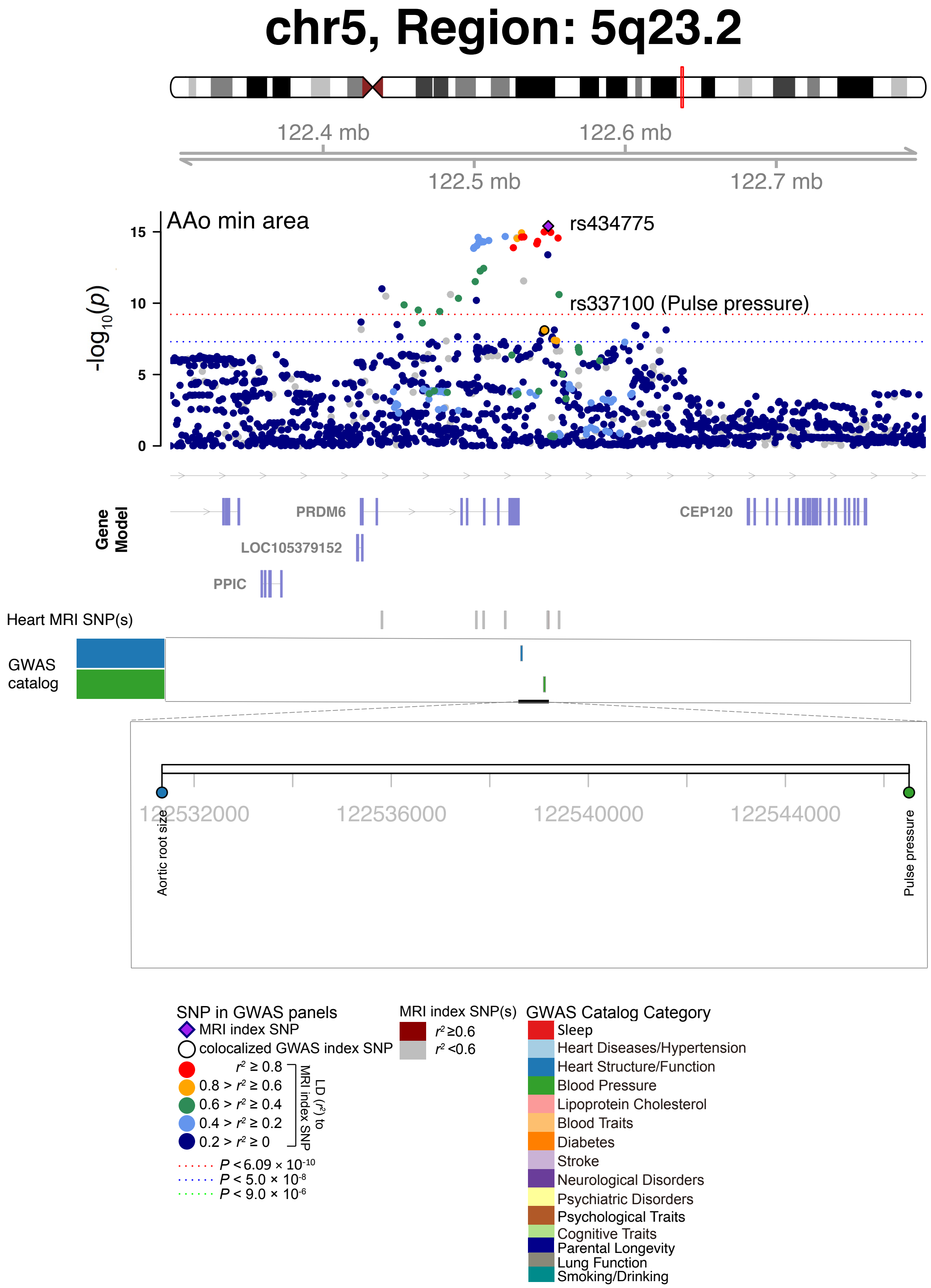


**Fig. S70 Selected genetic loci that were associated with both CMR trait and other complex traits and diseases.** In 5q23.2, we observed the shared association (LD $r^2 \geq 0.6$) between CMR trait (AAo min area, index variant rs434775) and pulse pressure (index variant rs337100). AAo min area, ascending aorta minimum area.

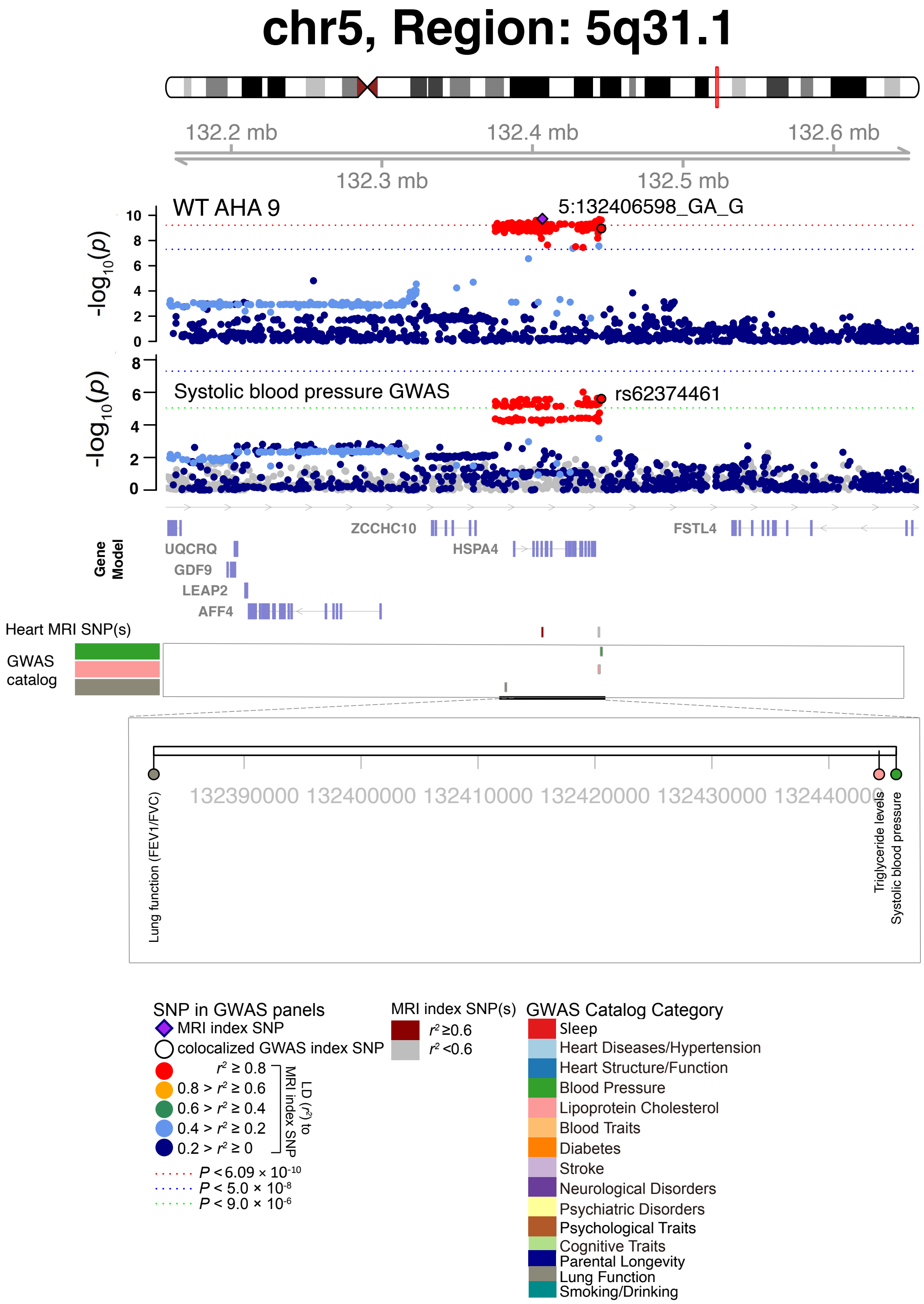


**Fig. S71 Selected genetic loci that were associated with both CMR trait and other complex traits and diseases.** In 5q31.1, we observed the shared association (LD $r^2 \geq 0.6$) between CMR trait (WT AHA 9, index variant 5:132406598_GA_G) and systolic blood pressure (index variant rs62374461). The posterior probability of Bayesian colocalization analysis for the shared causal variant hypothesis (PPH4) is 0.995. WT AHA 9, regional myocardial-wall thickness at end-diastole (region 9).

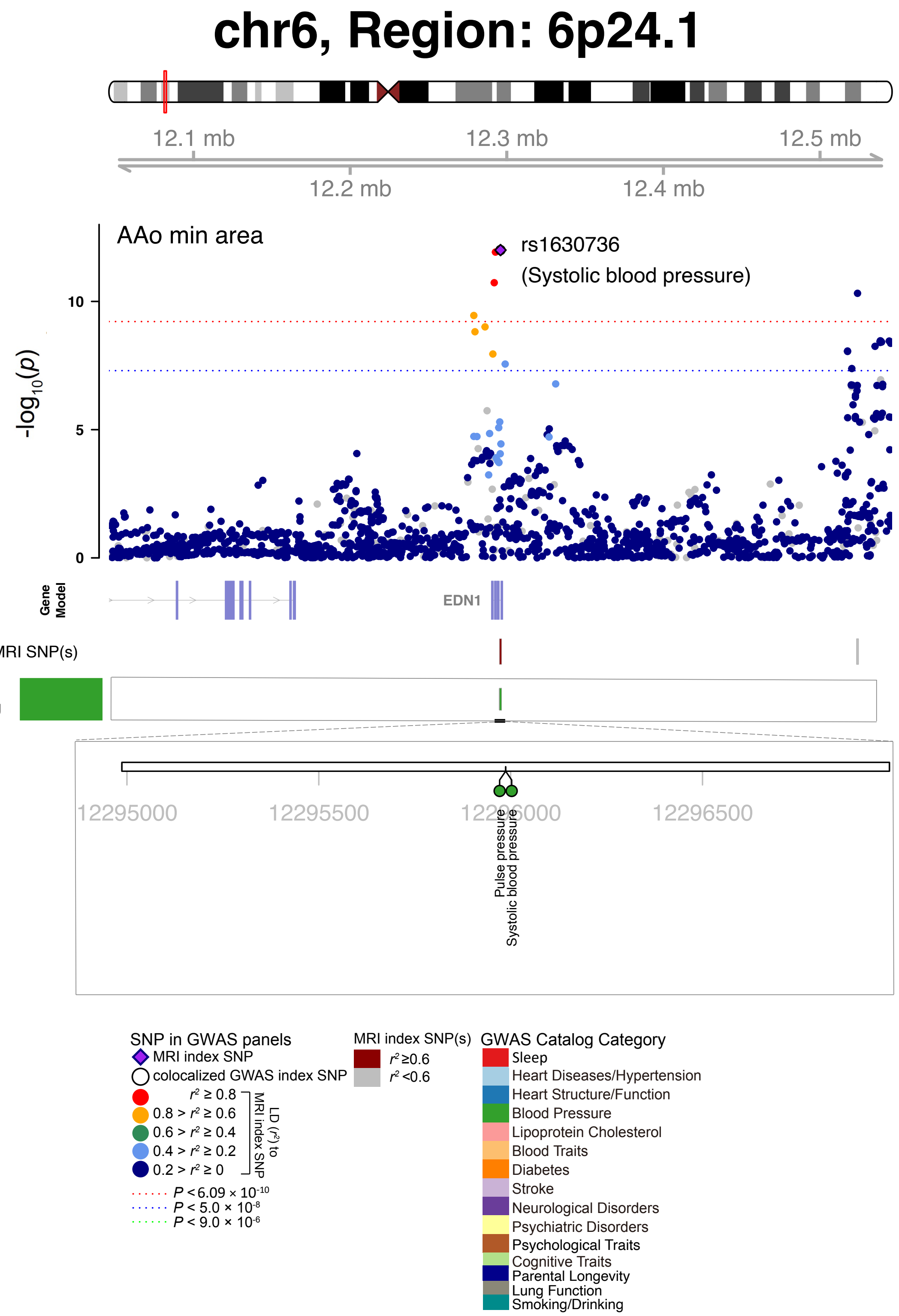


**Fig. S72 Selected genetic loci that were associated with both CMR trait and other complex traits and diseases.** In 6p24.1, we observed the shared association (LD $r^2 \geq 0.6$) between CMR trait (AAo min area) and systolic blood pressure (shared index variant rs1630736). AAo min area, ascending aorta minimum area.

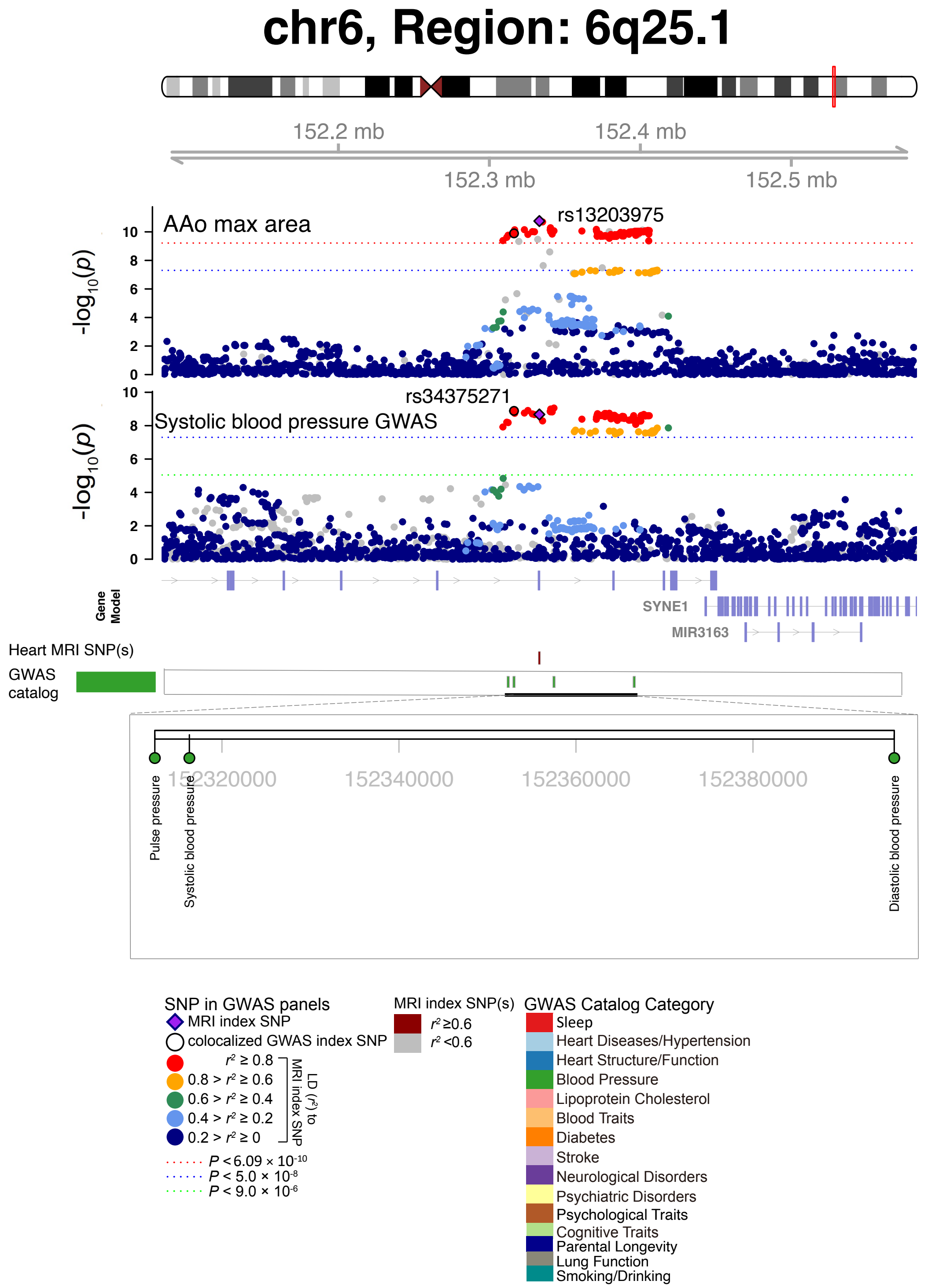


**Fig. S73 Selected genetic loci that were associated with both CMR trait and other complex traits and diseases.** In 6p25.1, we observed the shared association (LD $r^2 \geq 0.6$) between CMR trait (AAo max area, index variant rs13203975) and systolic blood pressure (index variant rs34375271). The posterior probability of Bayesian colocalization analysis for the shared causal variant hypothesis (PPH4) is 0.861. AAo max area, ascending aorta maximum area.

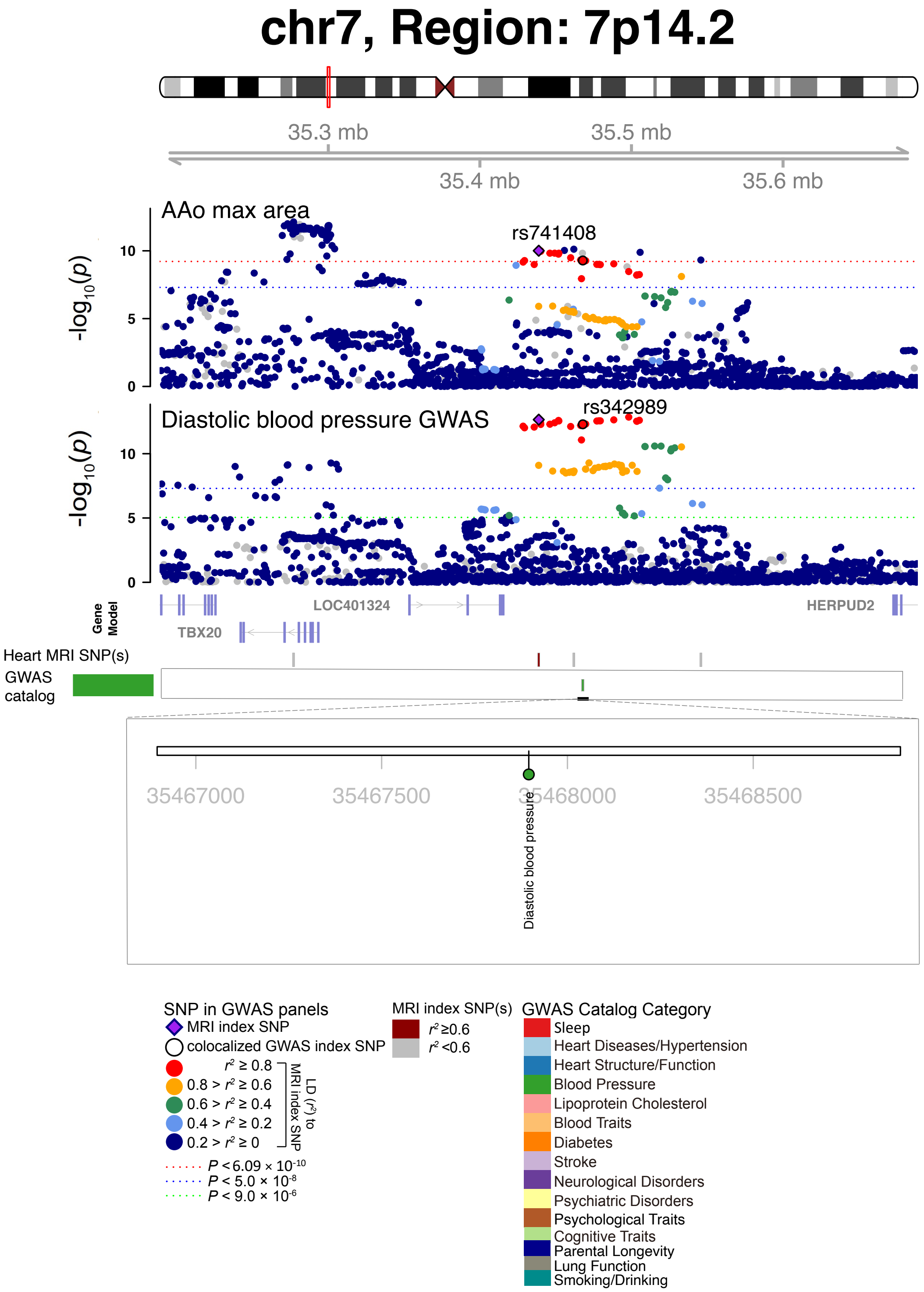


**Fig. S74 Selected genetic loci that were associated with both CMR trait and other complex traits and diseases.** In 7p14.2, we observed the shared association (LD $r^2 \geq 0.6$) between CMR trait (AAo max area, index variant rs741408) and diastolic blood pressure (index variant rs342989). The posterior probability of Bayesian colocalization analysis for the shared causal variant hypothesis (PPH4) is 0.941. AAo max area, ascending aorta maximum area.

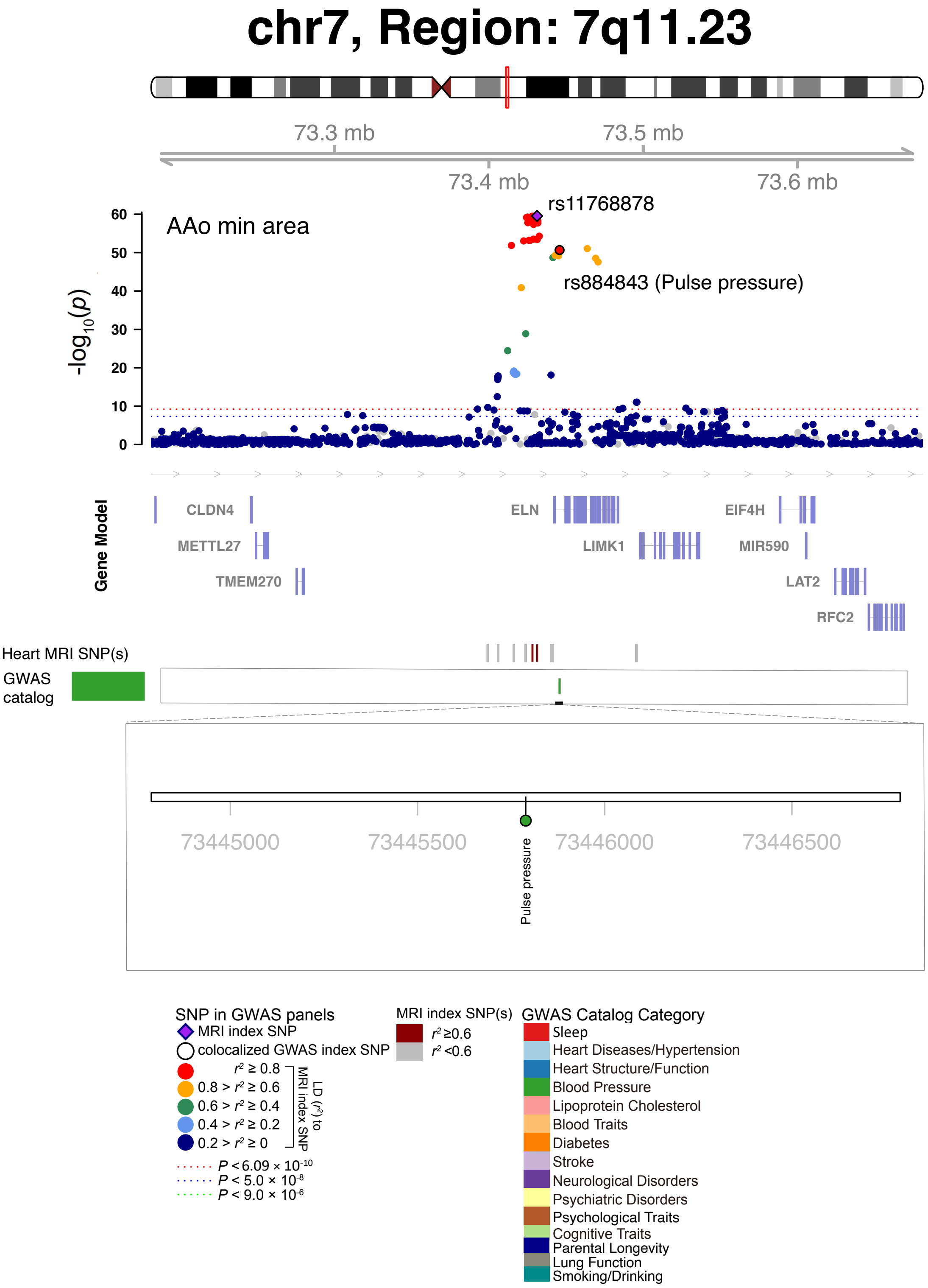


**Fig. S75 Selected genetic loci that were associated with both CMR trait and other complex traits and diseases.** In 7q11.23, we observed the shared association (LD $r^2 \geq 0.6$) between CMR trait (AAo min area, index variant rs11768878) and pulse pressure (index variant rs3884843). AAo min area, ascending aorta minimum area.

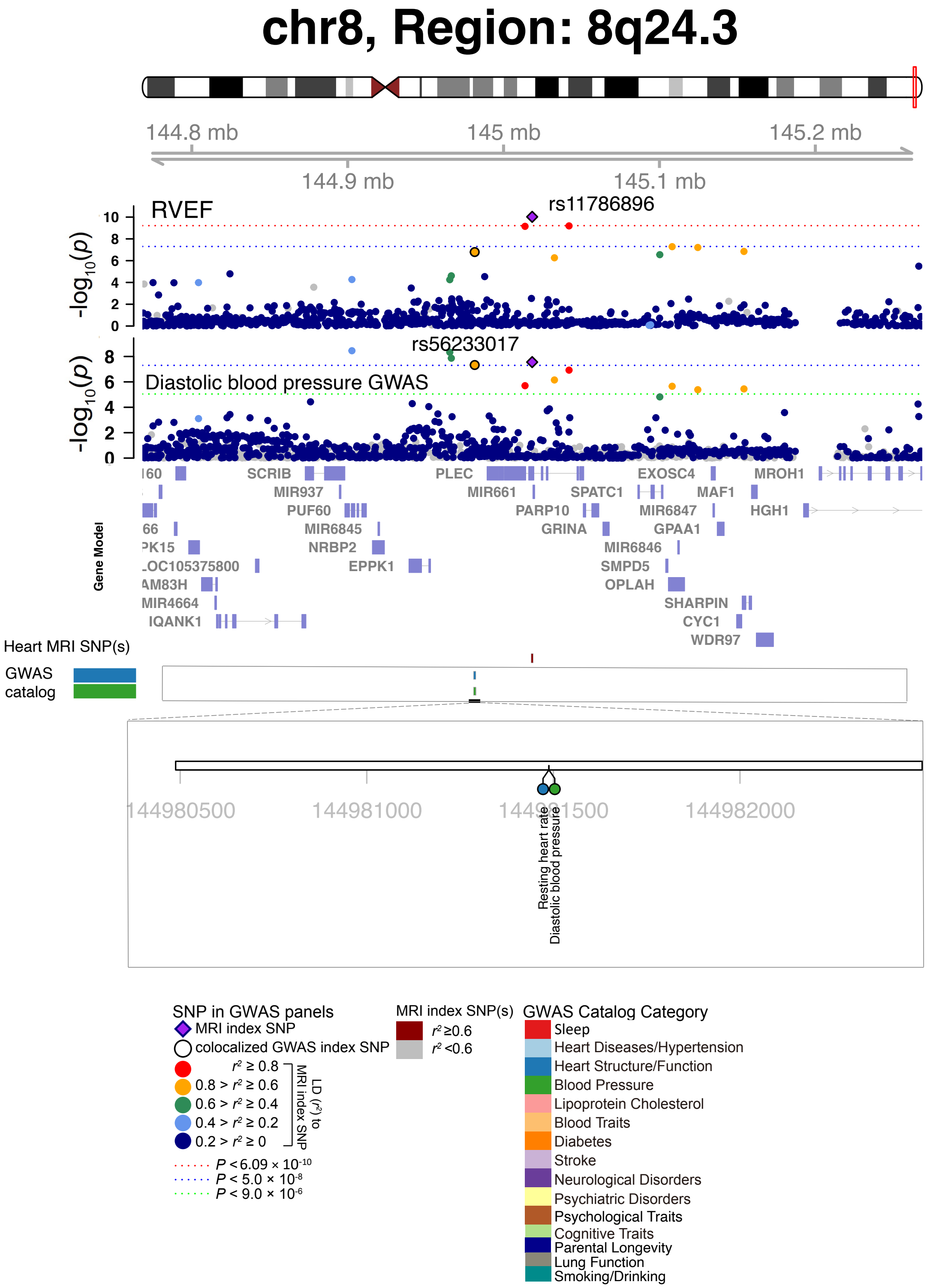


**Fig. S76 Selected genetic loci that were associated with both CMR trait and other complex traits and diseases.** In 8q24.3, we observed the shared association (LD $r^2 \geq 0.6$) between CMR trait (RVEF, index variant rs11786896) and diastolic blood pressure (index variant rs56233017). The posterior probability of Bayesian colocalization analysis for the shared causal variant hypothesis (PPH4) is 0.998.

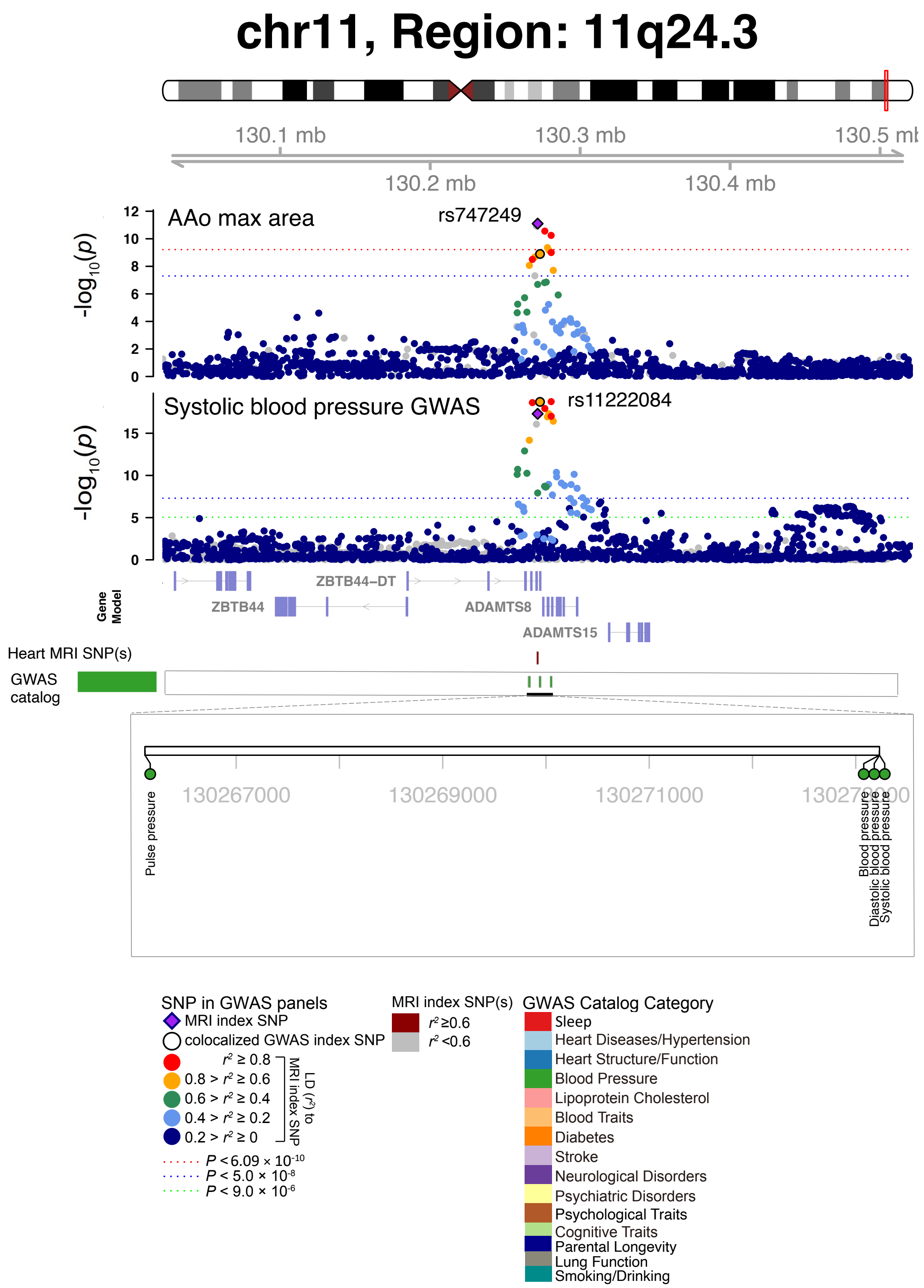


**Fig. S77 Selected genetic loci that were associated with both CMR trait and other complex traits and diseases.** In 11q24.3, we observed the shared association (LD $r^2 \geq 0.6$) between CMR trait (AAo max area, index variant rs747249) and systolic blood pressure (index variant rs11222084). The posterior probability of Bayesian colocalization analysis for the shared causal variant hypothesis (PPH4) is 0.996. AAo max area, ascending aorta maximum area.

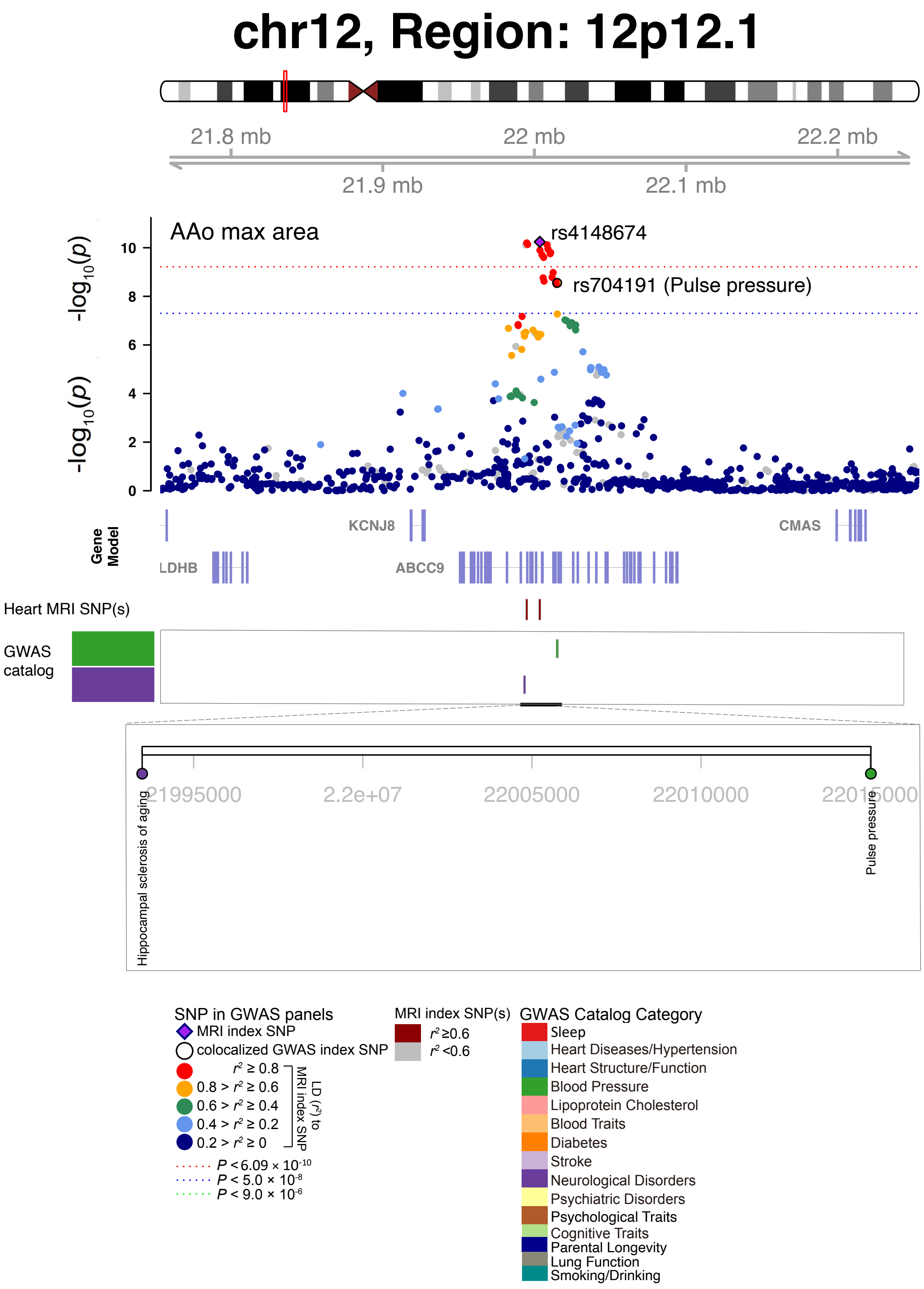


**Fig. S78 Selected genetic loci that were associated with both CMR trait and other complex traits and diseases.** In 12p12.1, we observed the shared association (LD $r^2 \geq 0.6$) between CMR trait (AAo max area, index variant rs4148674) and pulse pressure (index variant rs704191). AAo max area, ascending aorta maximum area.

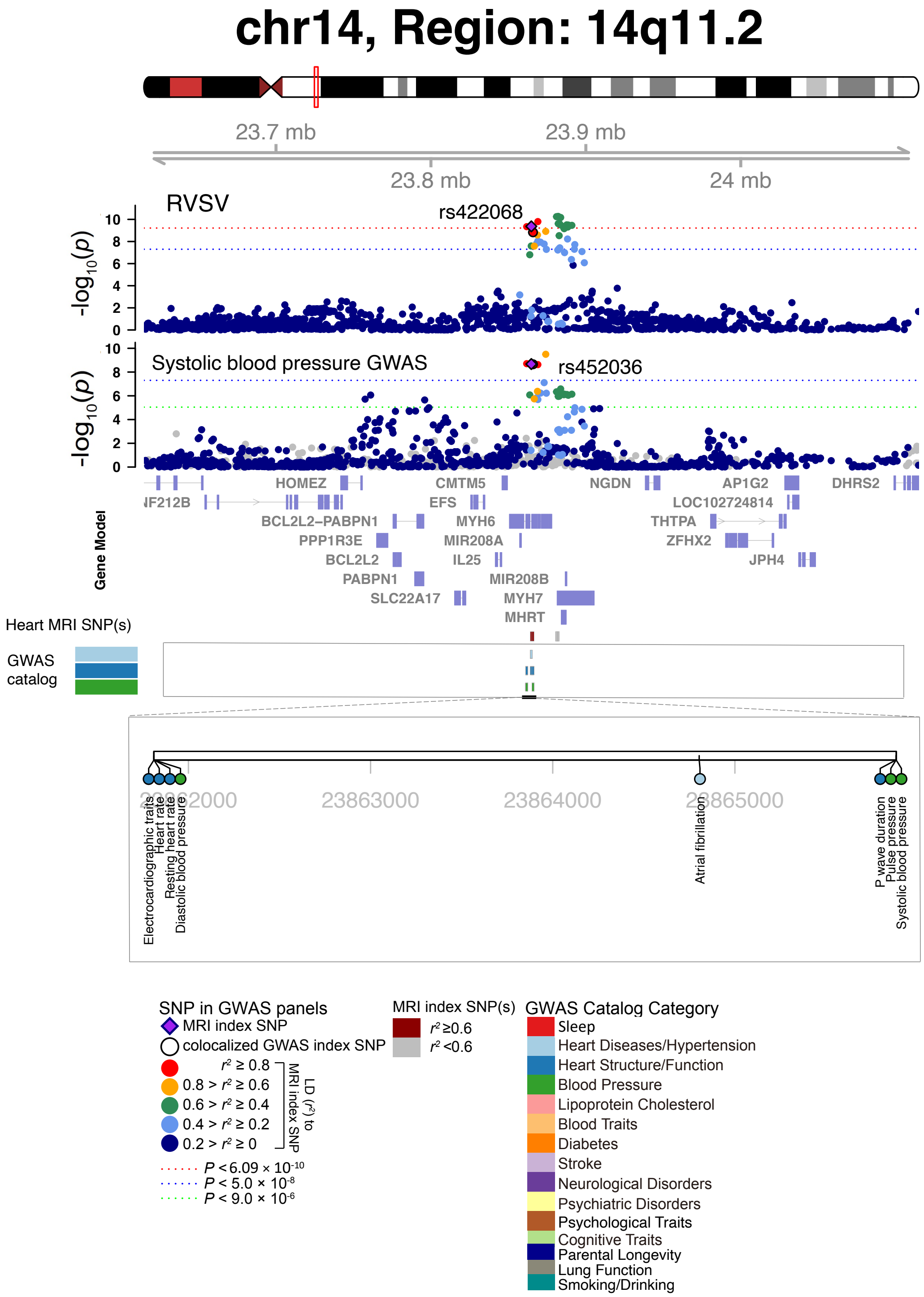


**Fig. S79 Selected genetic loci that were associated with both CMR trait and other complex traits and diseases.** In 14q11.2, we observed the shared association (LD $r^2 \geq 0.6$) between CMR trait (RVSV, index variant rs422068) and systolic blood pressure (index variant rs452036). The posterior probability of Bayesian colocalization analysis for the shared causal variant hypothesis (PPH4) is 0.991.

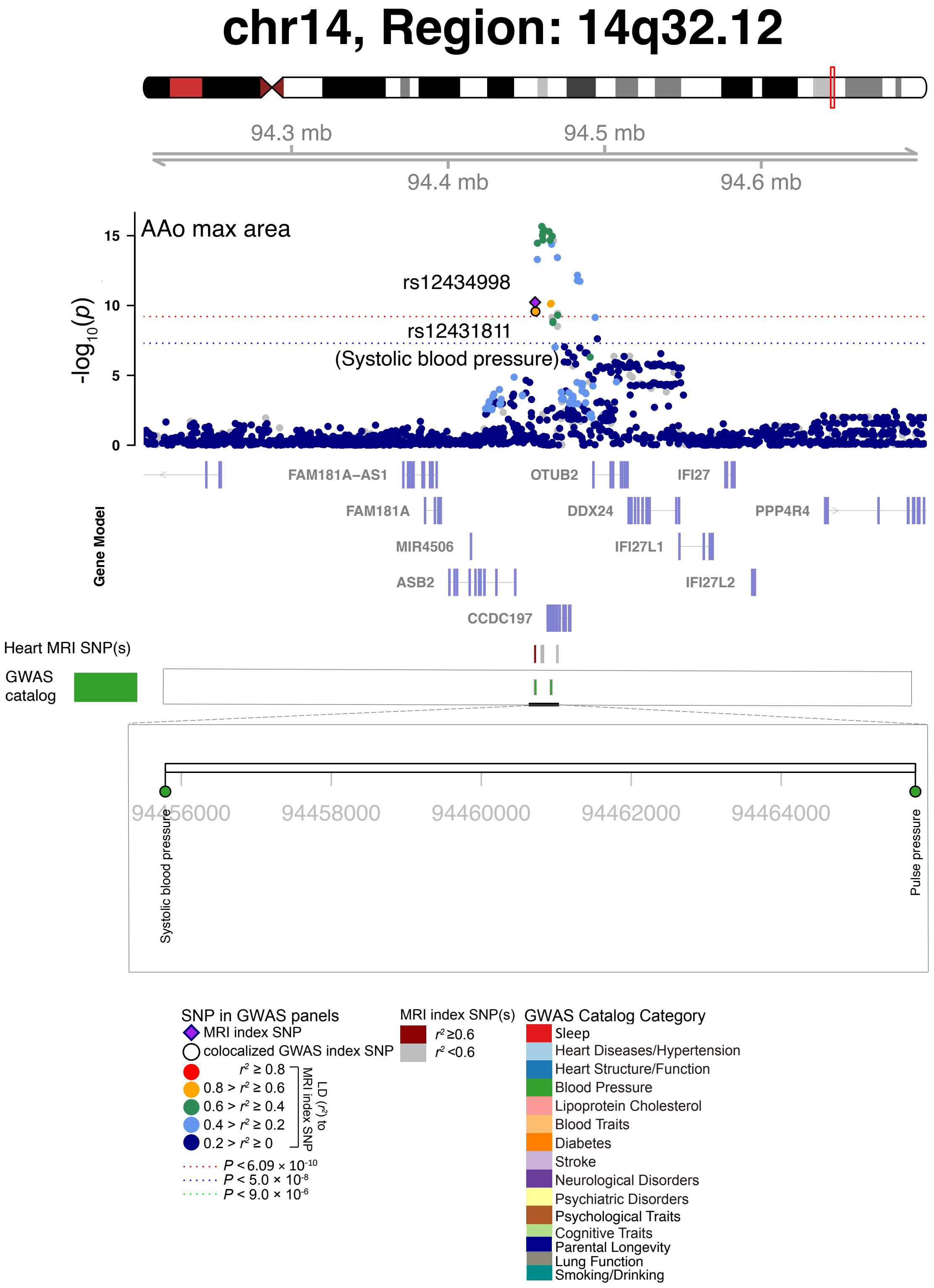


**Fig. S80 Selected genetic loci that were associated with both CMR trait and other complex traits and diseases.** In 14q32.12, we observed the shared association (LD $r^2 \geq 0.6$) between CMR trait (AAo max area, index variant rs12434998) and systolic blood pressure (index variant rs12431811). AAo max area, ascending aorta maximum area.

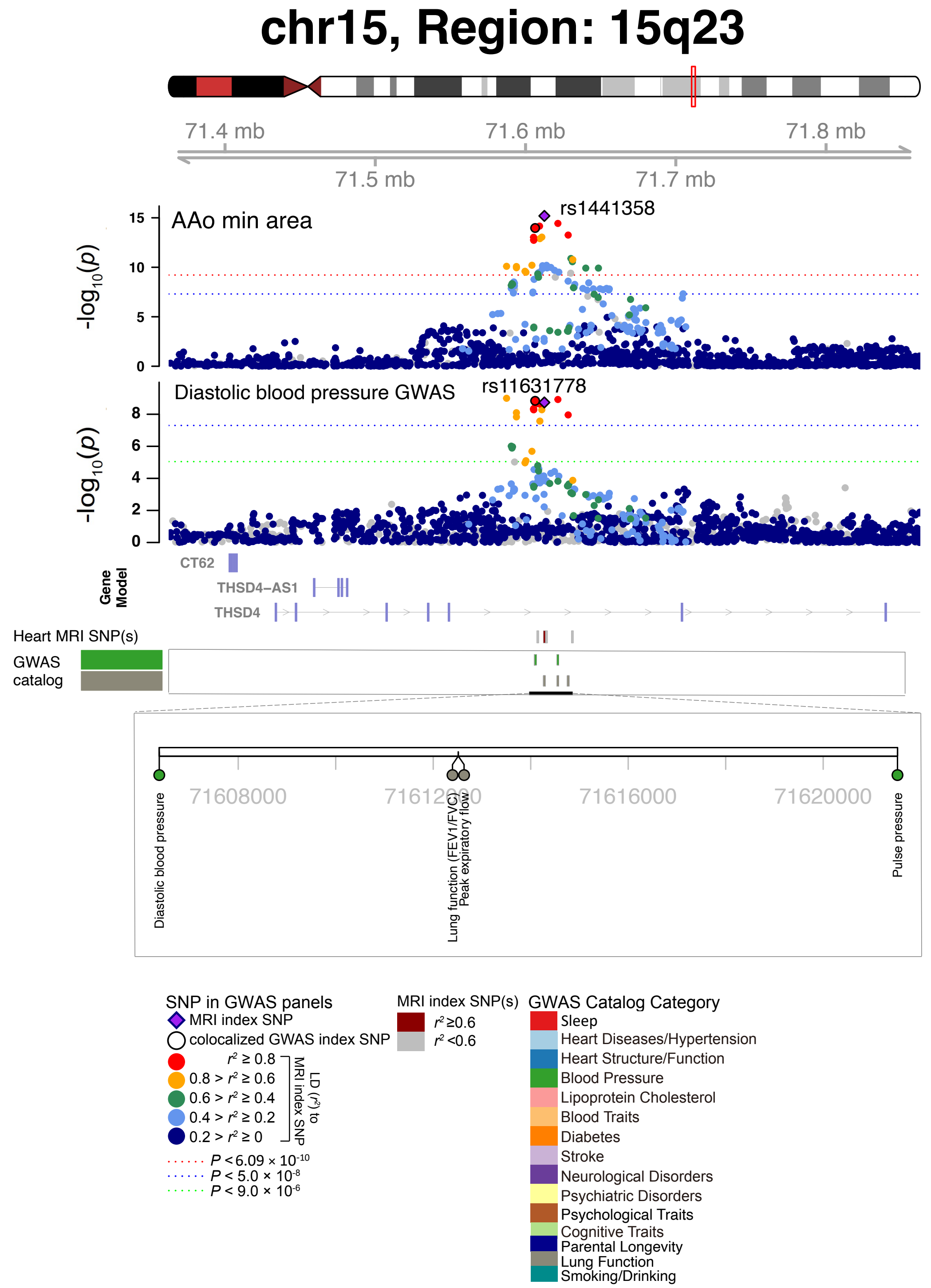


**Fig. S81 Selected genetic loci that were associated with both CMR trait and other complex traits and diseases.** In 15q23, we observed the shared association (LD $r^2 \geq 0.6$) between CMR trait (AAo min area, index variant rs1441358) and diastolic blood pressure (index variant rs11631778). The posterior probability of Bayesian colocalization analysis for the shared causal variant hypothesis (PPH4) is 0.992. AAo min area, ascending aorta minimum area.

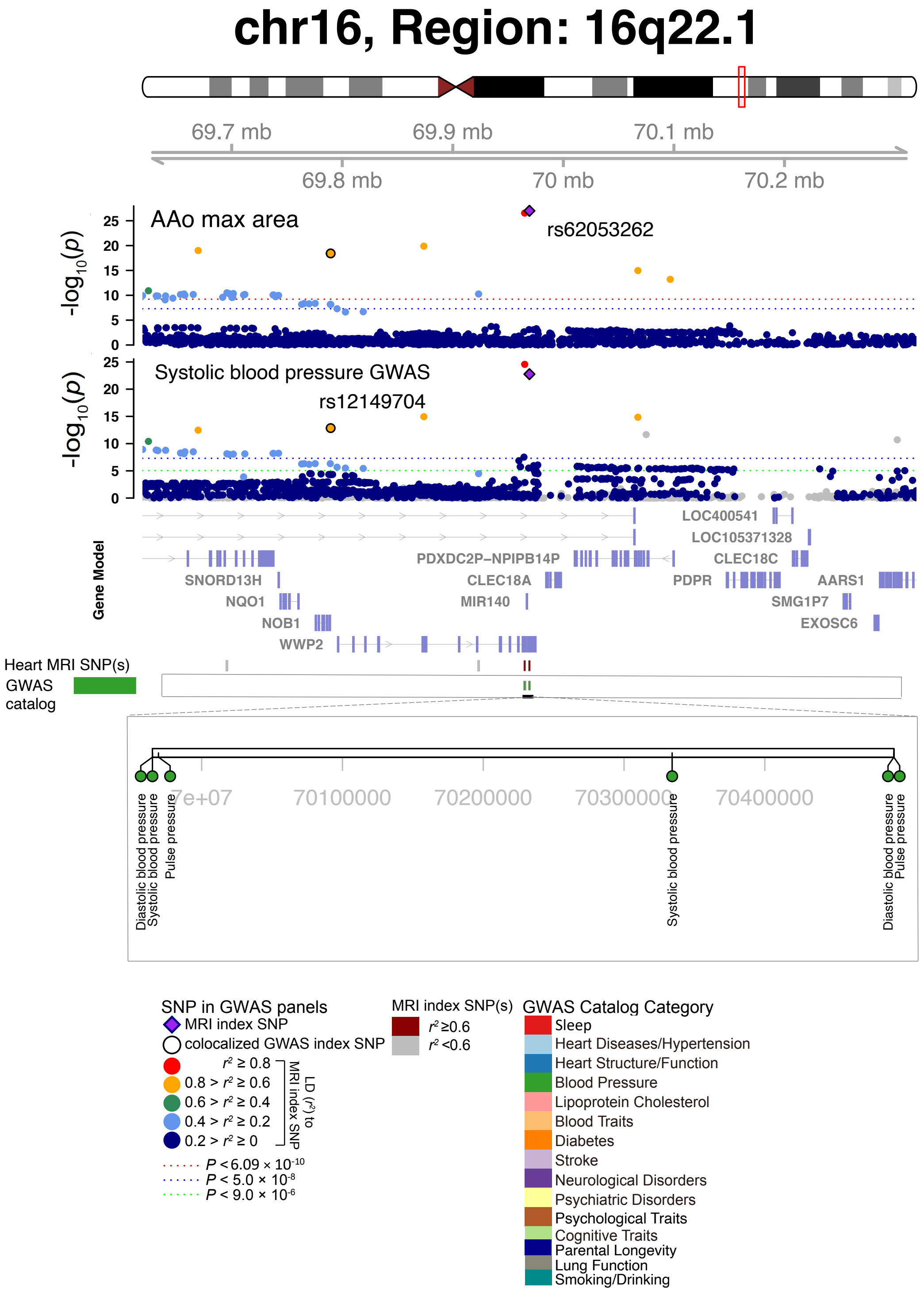


**Fig. S82 Selected genetic loci that were associated with both CMR trait and other complex traits and diseases.** In 16q22.1, we observed the shared association (LD $r^2 \geq 0.6$) between CMR trait (AAo max area, index variant rs62053262) and systolic blood pressure (index variant rs12149704). The posterior probability of Bayesian colocalization analysis for the shared causal variant hypothesis (PPH4) is 0.995. AAo max area, ascending aorta maximum area.

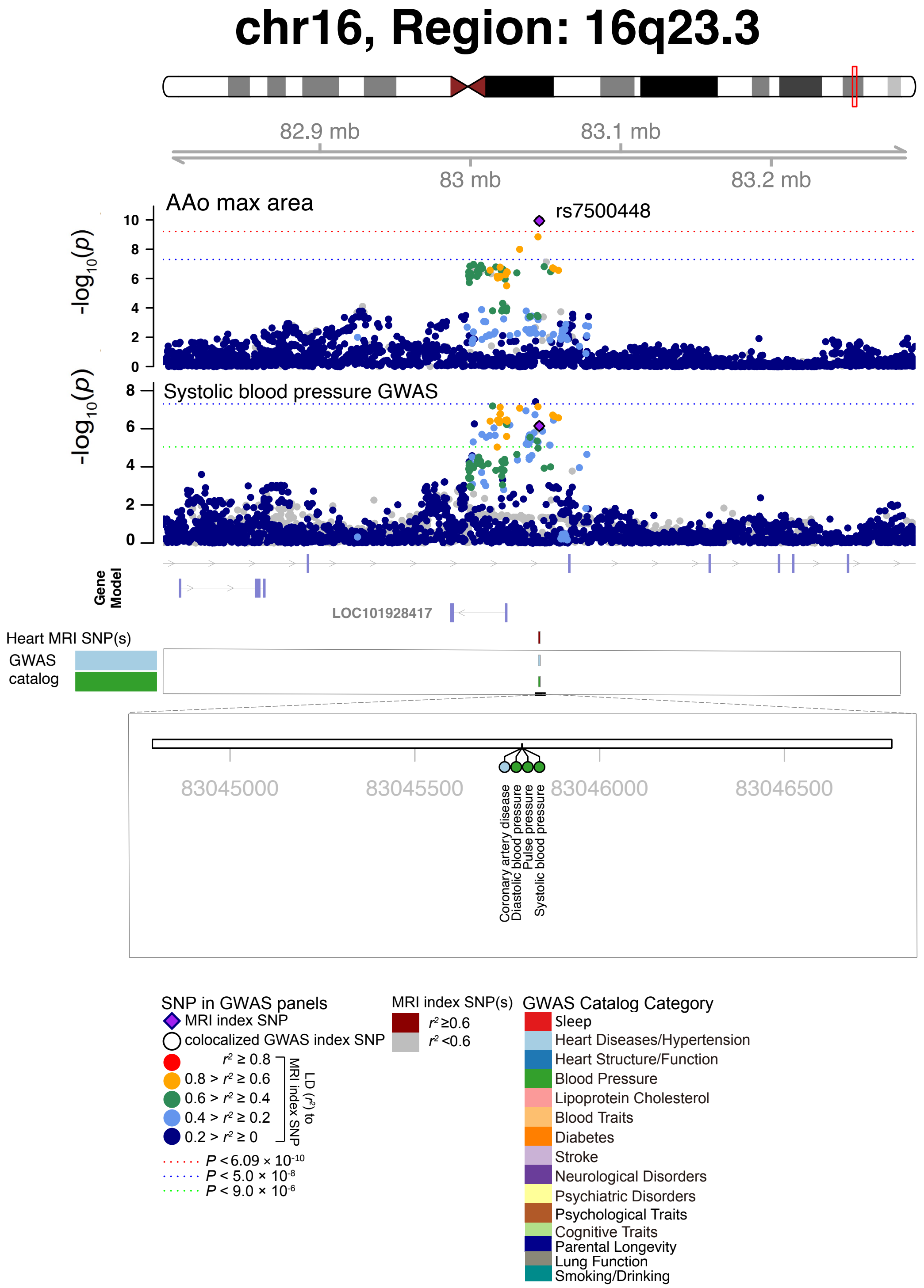


**Fig. S83 Selected genetic loci that were associated with both CMR trait and other complex traits and diseases.** In 16q23.3, we observed the shared association (LD $r^2 \geq 0.6$) between CMR trait (AAo max area) and systolic blood pressure (shared index variant rs7500448). The posterior probability of Bayesian colocalization analysis for the shared causal variant hypothesis (PPH4) is 0.999. AAo max area, ascending aorta maximum area.

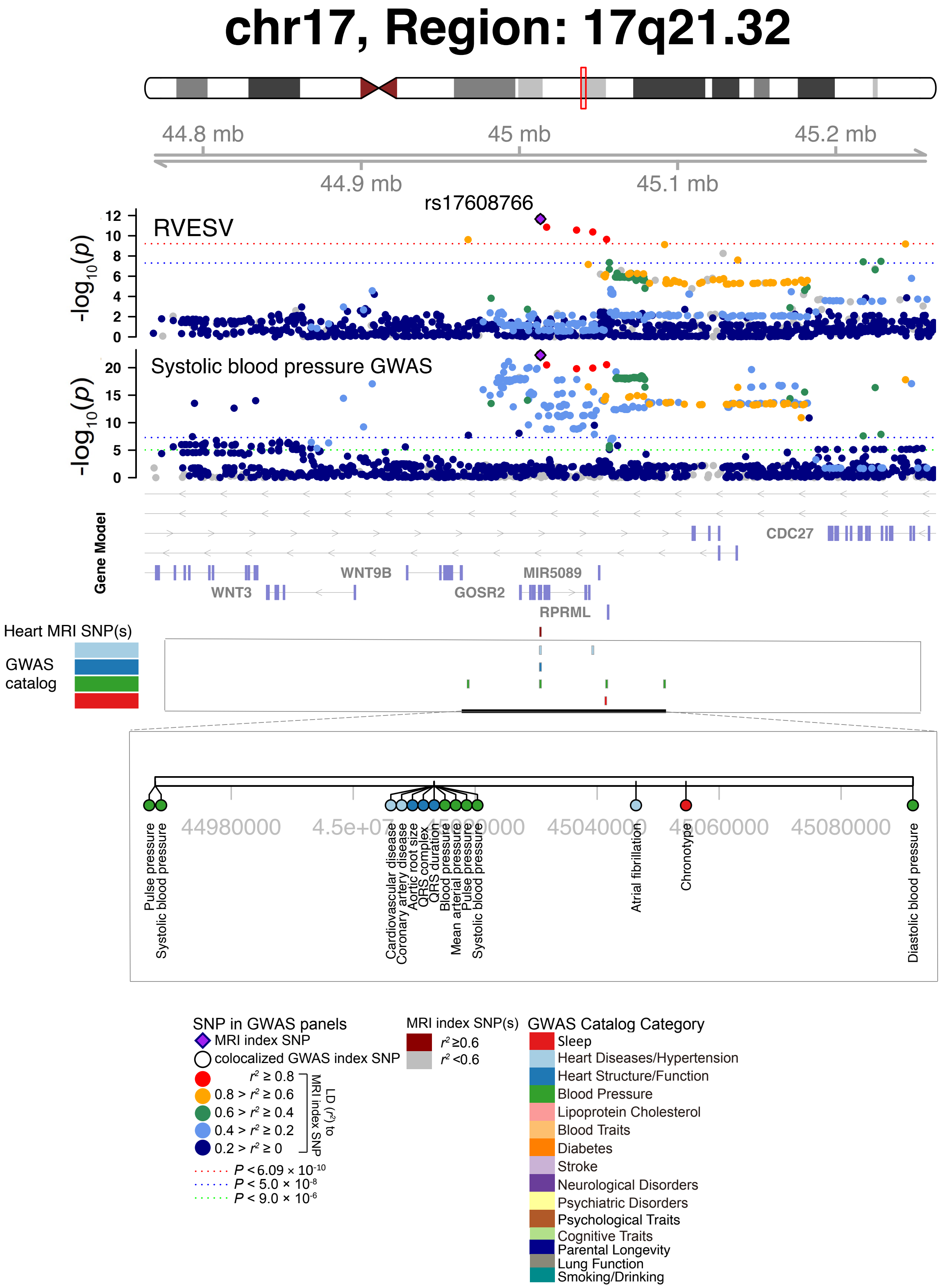


**Fig. S84 Selected genetic loci that were associated with both CMR trait and other complex traits and diseases.** In 17q21.32, we observed the shared association (LD $r^2 \geq 0.6$) between CMR trait (RVESV) and systolic blood pressure (shared index variant rs17608766).

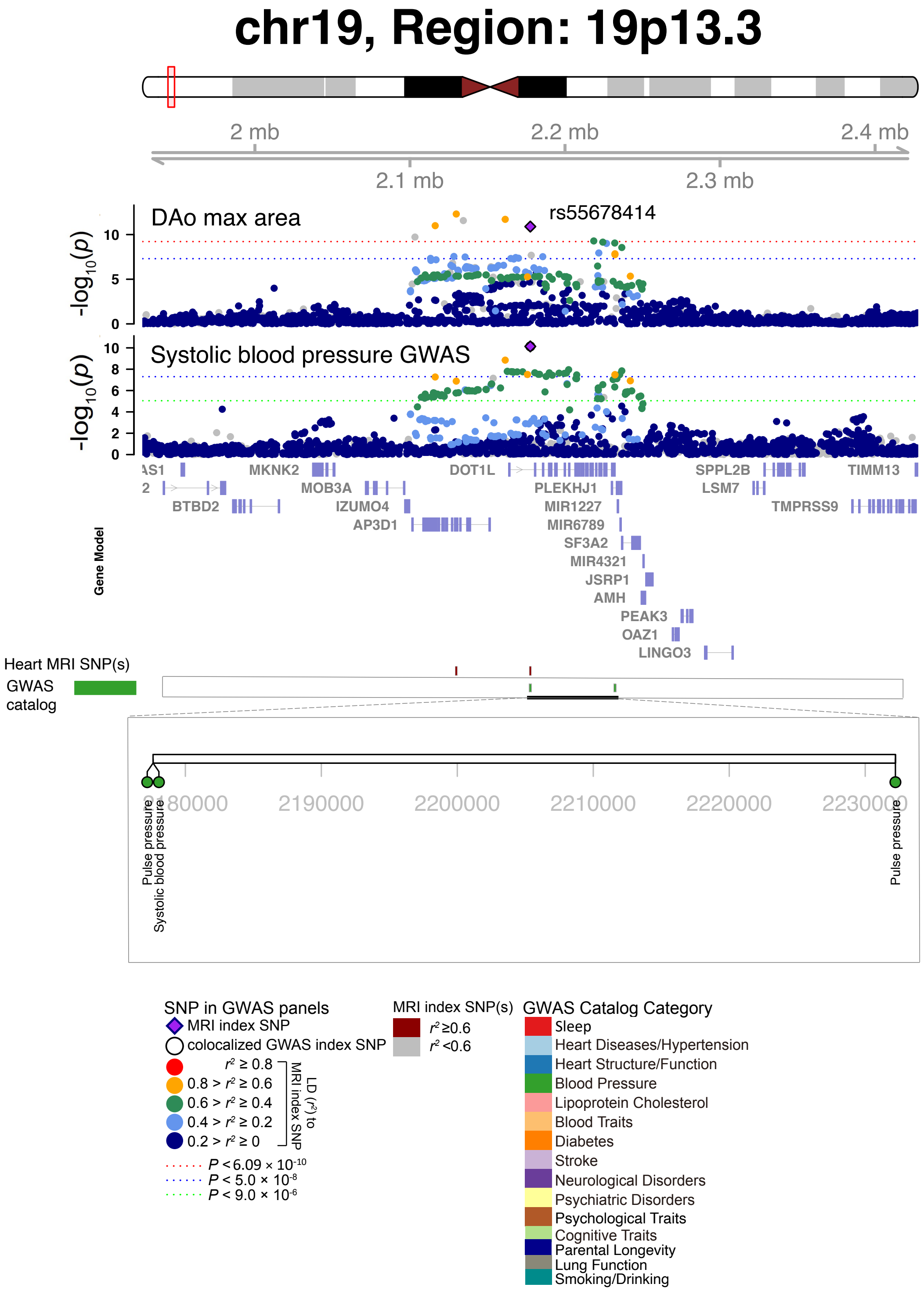


**Fig. S85 Selected genetic loci that were associated with both CMR trait and other complex traits and diseases.** In 19p13.3, we observed the shared association (LD $r^2 \geq 0.6$) between CMR trait (DAo max area) and systolic blood pressure (shared index variant rs55678414). The posterior probability of Bayesian colocalization analysis for the shared causal variant hypothesis (PPH4) is 0.837. DAo max area, descending aorta maximum area.

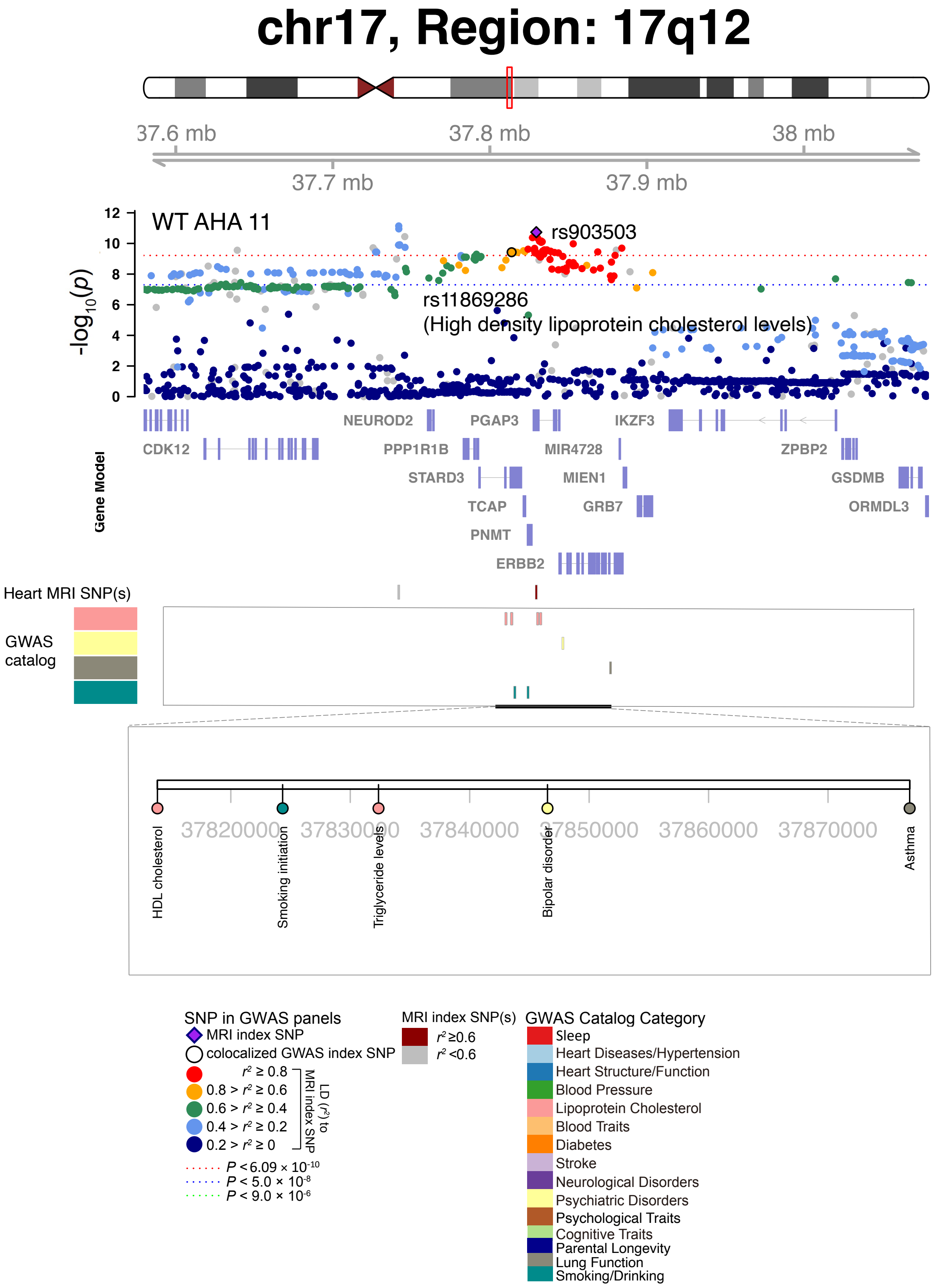


**Fig. S86 Selected genetic loci that were associated with both CMR trait and other complex traits and diseases.** In 17q12, we observed the shared association (LD $r^2 \geq 0.6$) between CMR trait (WT AHA 11, index variant rs903503) and high-density lipoprotein cholesterol levels (index variant rs11869286). WT AHA 11, regional myocardial-wall thickness at end-diastole (region 11).

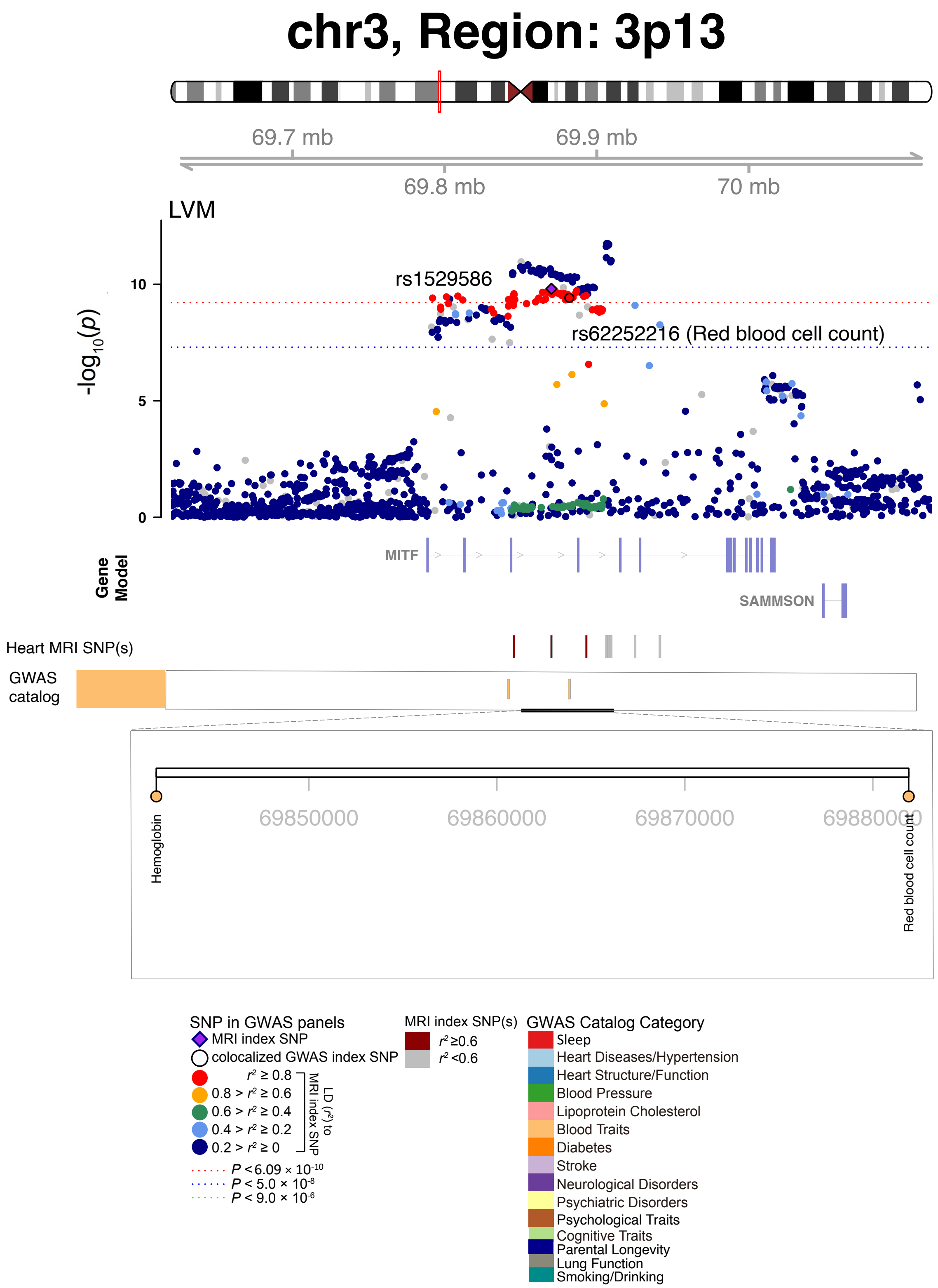


**Fig. S87 Selected genetic loci that were associated with both CMR trait and other complex traits and diseases.** In 3p13, we observed the shared association (LD $r^2 \geq 0.6$) between CMR trait (LVM, index variant rs1529586) and red blood cell count (index variant rs62252216).

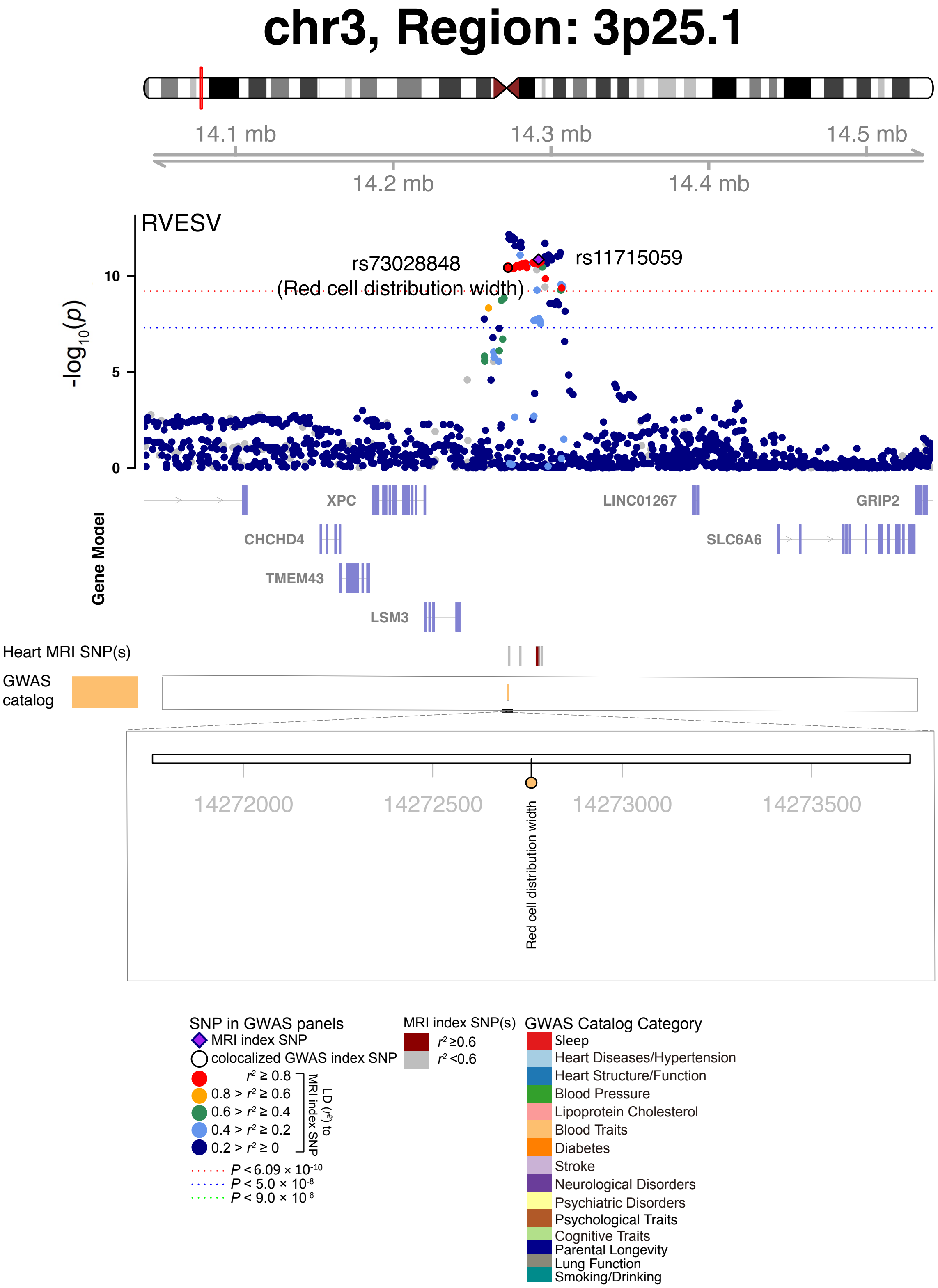


**Fig. S88 Selected genetic loci that were associated with both CMR trait and other complex traits and diseases.** In 3p25.1, we observed the shared association (LD $r^2 \geq 0.6$) between CMR trait (RVESV, index variant rs11715059) and red cell distribution width (index variant rs73028848).

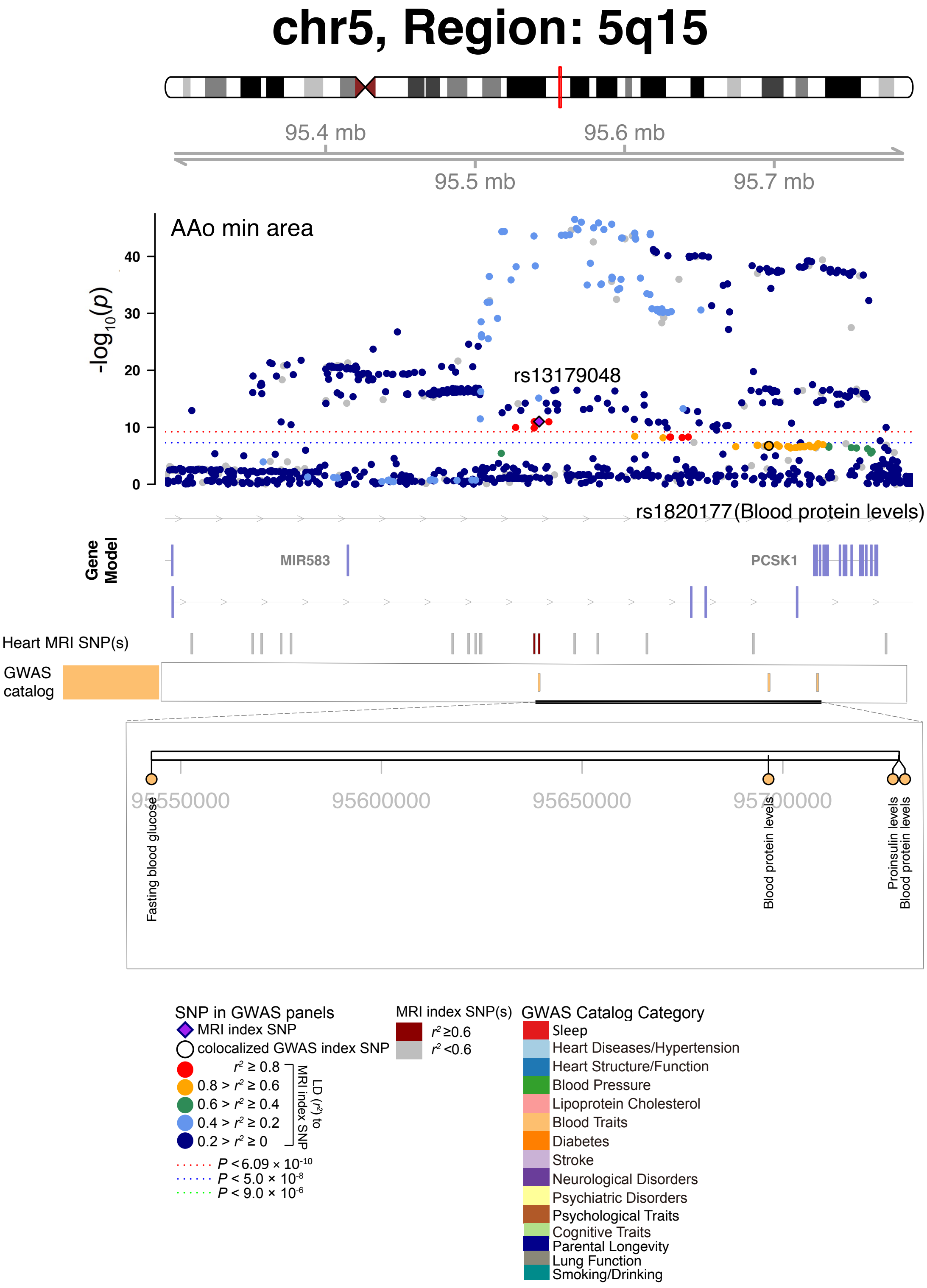


**Fig. S89 Selected genetic loci that were associated with both CMR trait and other complex traits and diseases.** In 5q15, we observed the shared association (LD $r^2 \geq 0.6$) between CMR trait (AAo min area, index rs13179048) and blood protein levels (index variant rs1820177). AAo min area, ascending aorta minimum area.

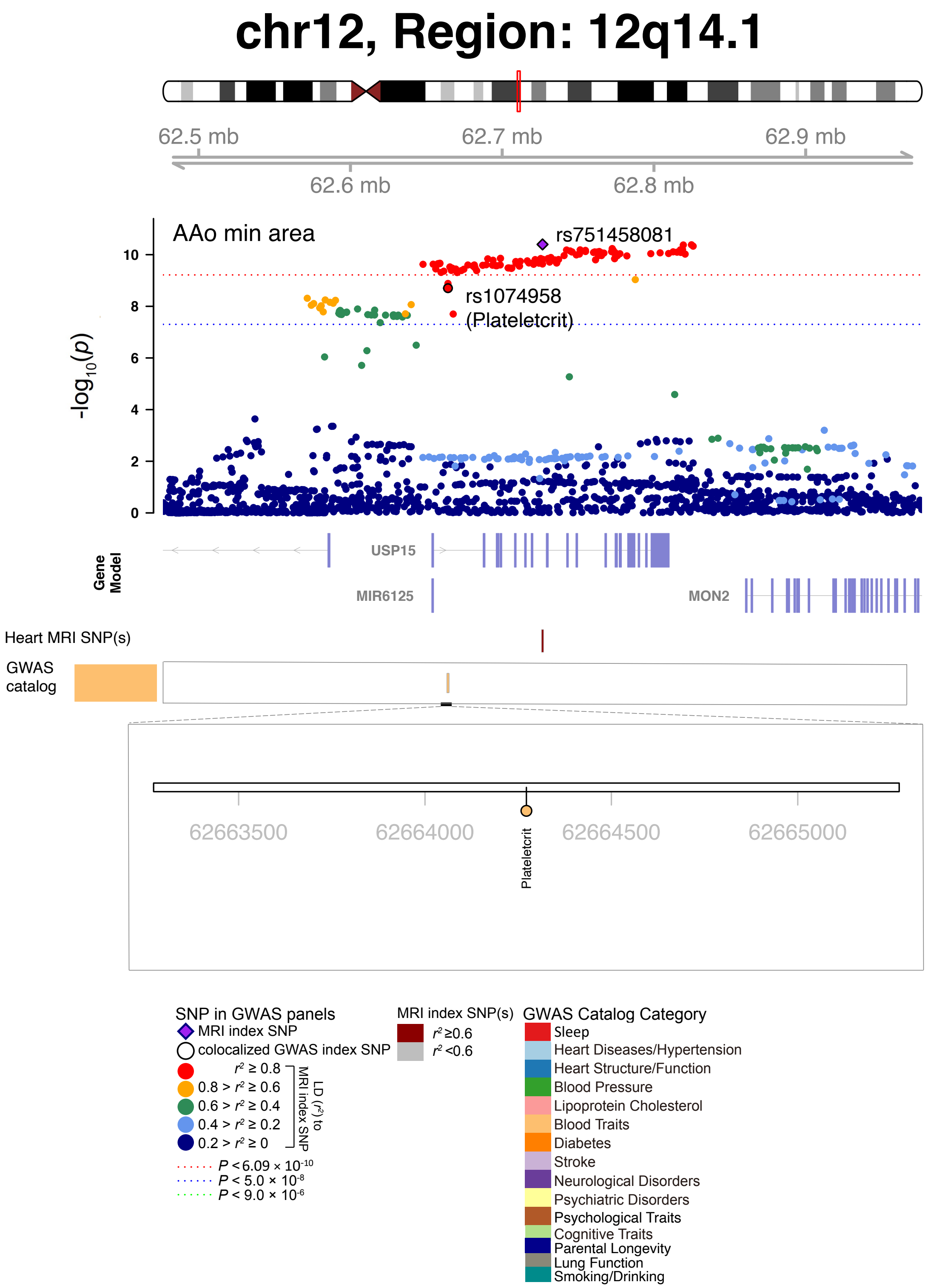


**Fig. S90 Selected genetic loci that were associated with both CMR trait and other complex traits and diseases.** In 12q14.1, we observed the shared association (LD $r^2 \geq 0.6$) between CMR trait (AAo min area, index rs75458081) and plateletcrit (index variant rs1074958). AAo min area, ascending aorta minimum area.

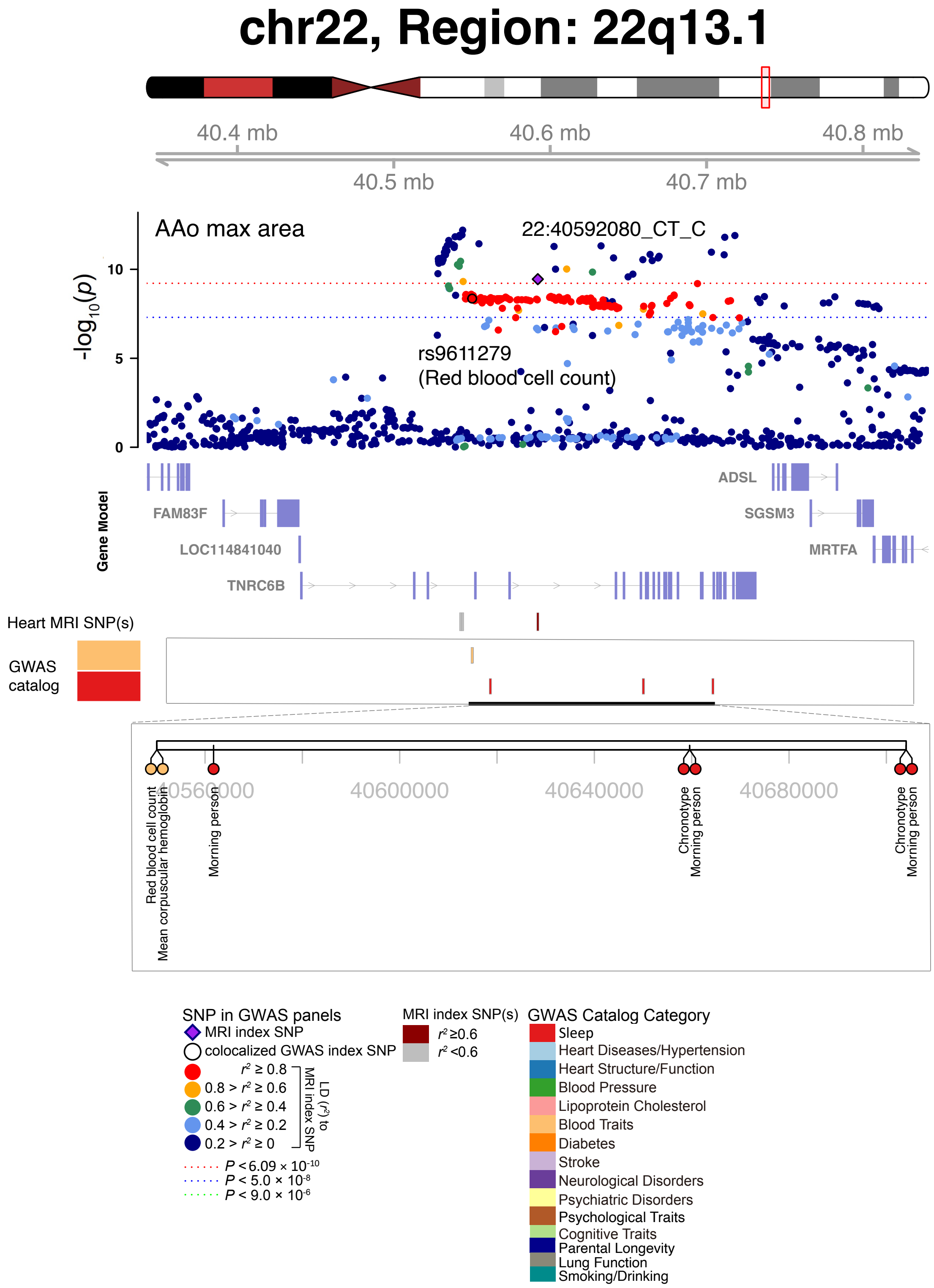


**Fig. S91 Selected genetic loci that were associated with both CMR trait and other complex traits and diseases.** In 22q13.1, we observed the shared association (LD $r^2 \geq 0.6$) between CMR trait (AAo max area, index variant 22:40592080_CT_C) and red blood cell count (index variant rs9611279). AAo max area, ascending aorta maximum area.

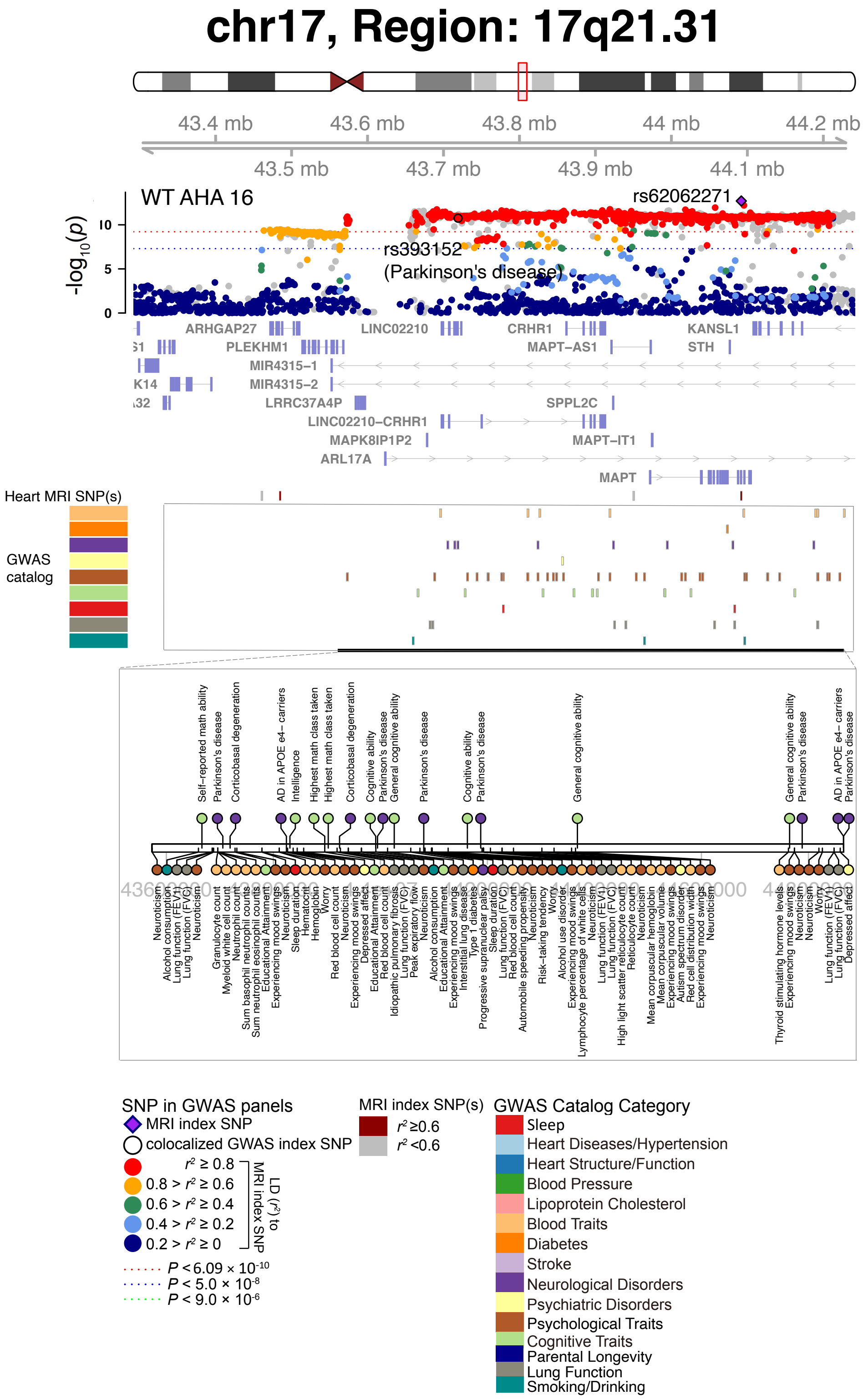


**Fig. S92 Selected genetic loci that were associated with both CMR trait and other complex traits and diseases.** In 17q21.31, we observed the shared association (LD $r^2 \geq 0.6$) between CMR trait (WT AHA 16, index variant rs62062271) and Parkinson's disease (index variant rs393152). WT AHA 16, regional myocardial-wall thickness at end-diastole (region 16).

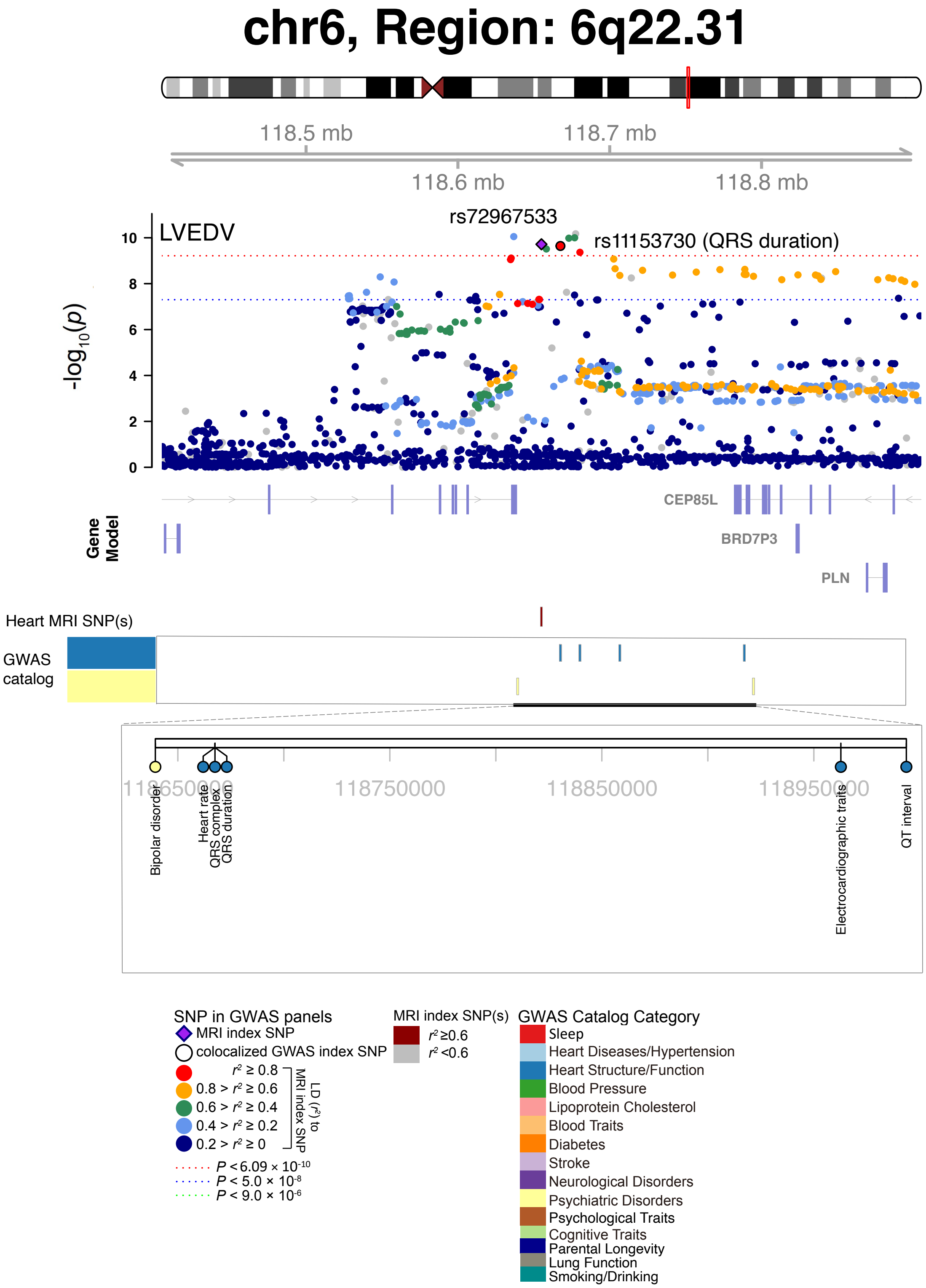


**Fig. S93 Selected genetic loci that were associated with both CMR trait and other complex traits and diseases.** In 6q22.31, we observed the shared association (LD $r^2 \geq 0.6$) between CMR trait (LVEDV, index variant rs72967533) and QRS duration (index variant rs11153730). We also observed colocalization with bipolar disorder.

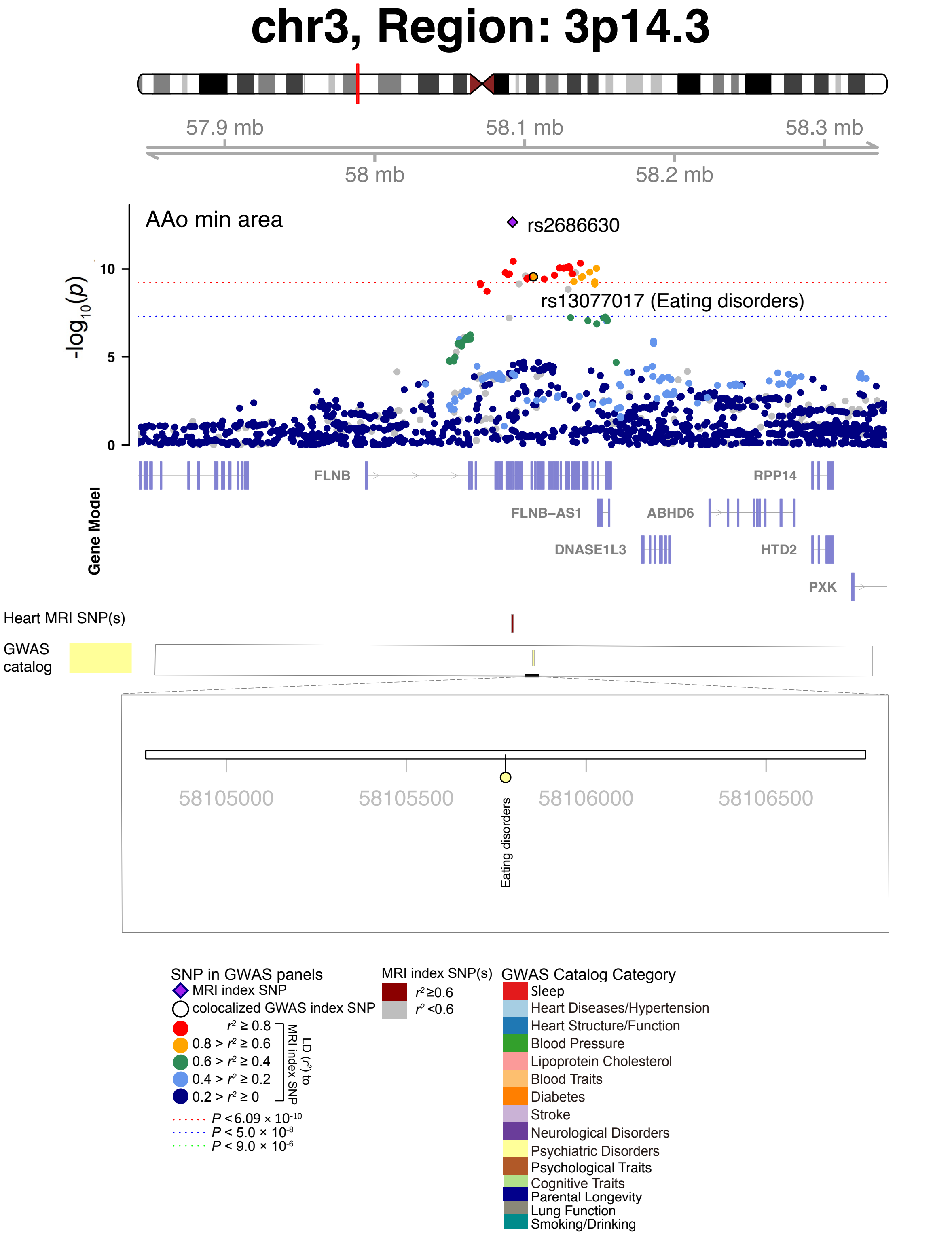


**Fig. S94 Selected genetic loci that were associated with both CMR trait and other complex traits and diseases.** In 3p14.3, we observed the shared association (LD $r^2 \geq 0.6$) between CMR trait (AAo min area, index variant rs2686630) and eating disorders (index variant rs13077017). AAo min area, ascending aorta minimum area.

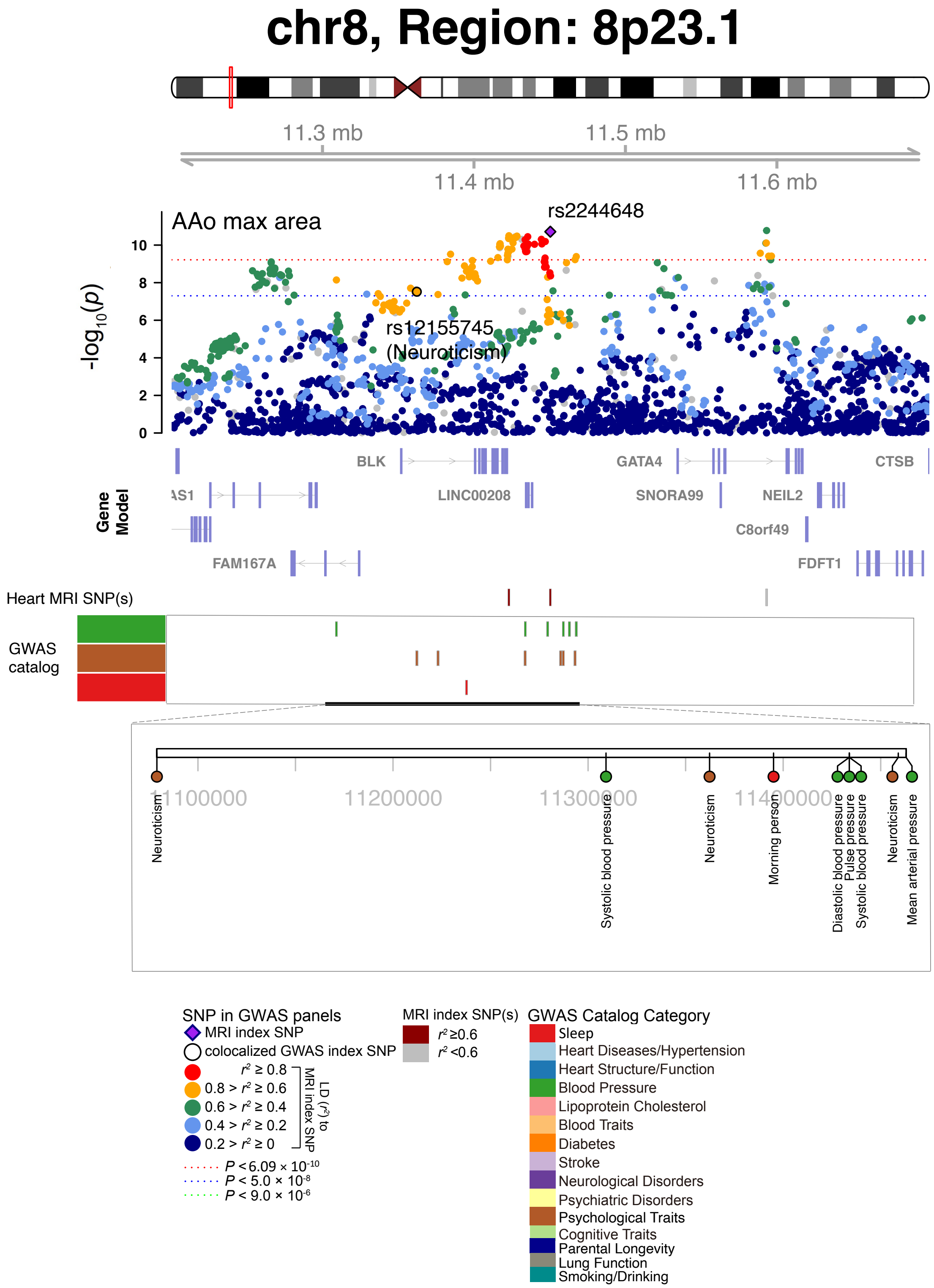


**Fig. S95 Selected genetic loci that were associated with both CMR trait and other complex traits and diseases.** In 8p23.1, we observed the shared association (LD $r^2 \geq 0.6$) between CMR trait (AAo max area, index variant rs2244648) and neuroticism (index variant rs12155745). AAo max area, ascending aorta maximum area.

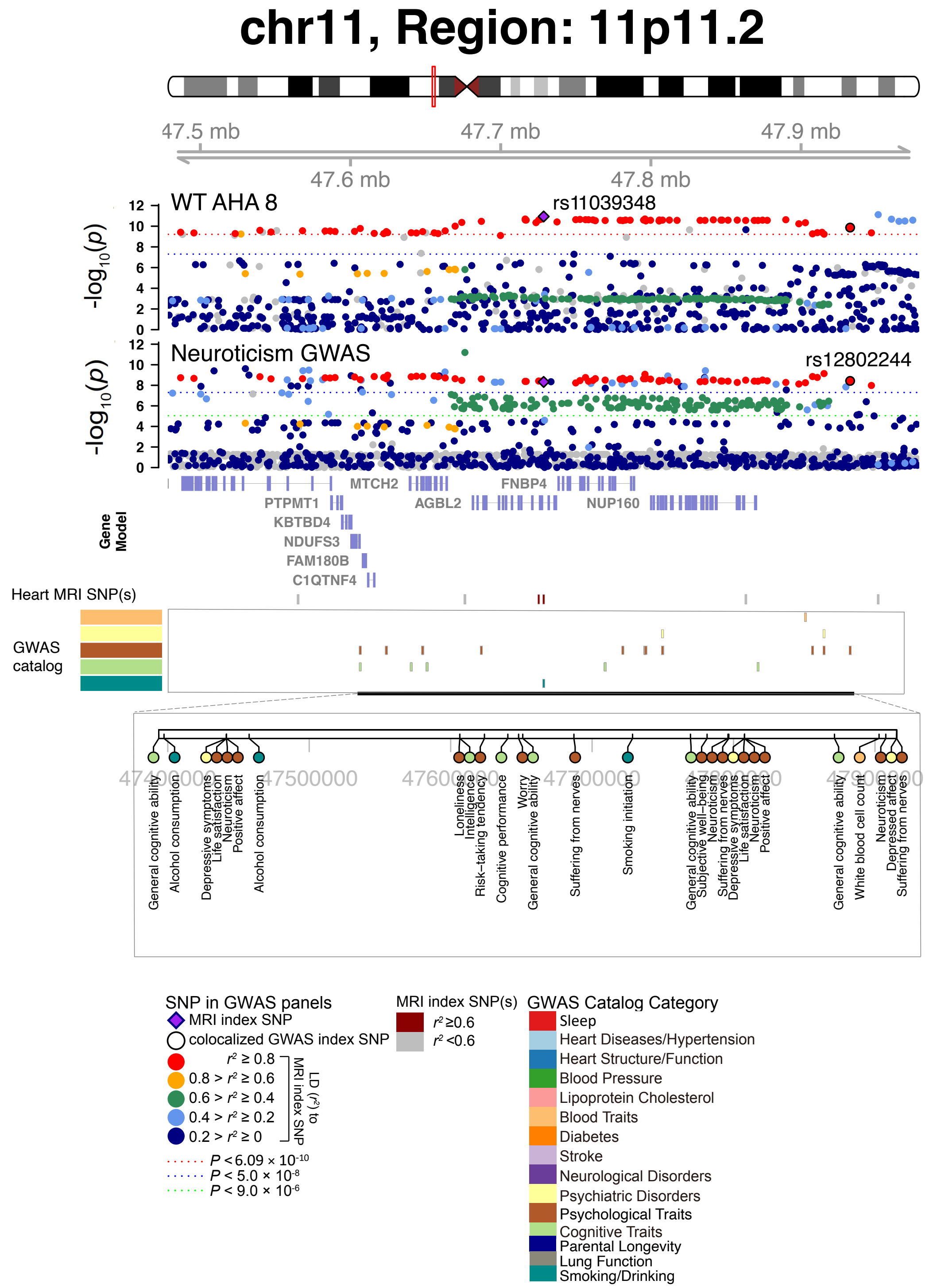


**Fig. S96 Selected genetic loci that were associated with both CMR trait and other complex traits and diseases.** In 11p11.2, we observed the shared association (LD $r^2 \geq 0.6$) between CMR trait (WT AHA 8, index variant rs11039348) and neuroticism (index variant rs12802244). The posterior probability of Bayesian colocalization analysis for the shared causal variant hypothesis (PPH4) is 0.947. WT AHA 8, regional myocardial-wall thickness at end-diastole (region 8).

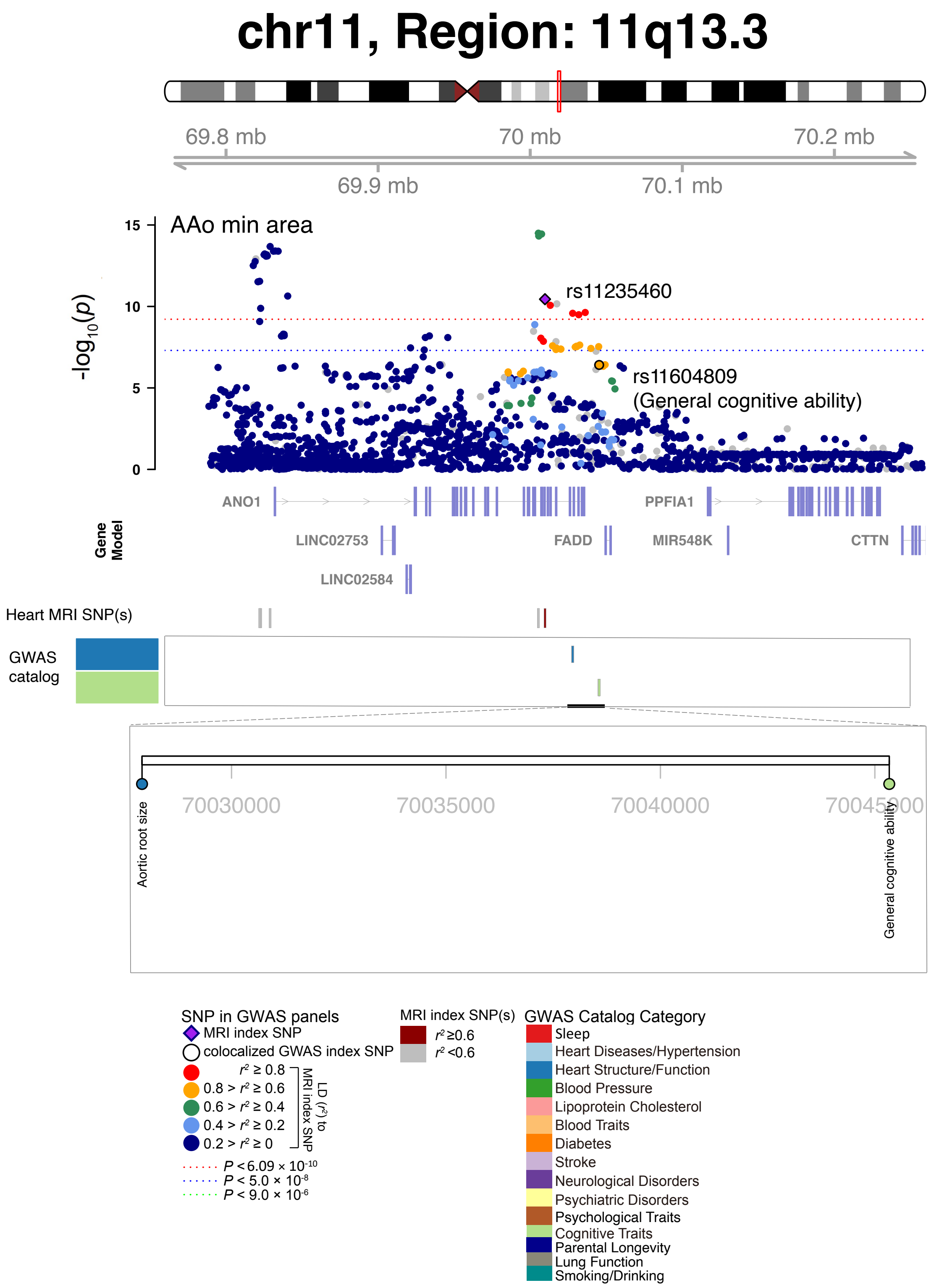


**Fig. S97 Selected genetic loci that were associated with both CMR trait and other complex traits and diseases.** In 11q13.3, we observed the shared association (LD $r^2 \geq 0.6$) between CMR trait (AAo min area, index variant rs11235460) and general cognitive ability (index variant rs11604809). AAo min area, ascending aorta minimum area.

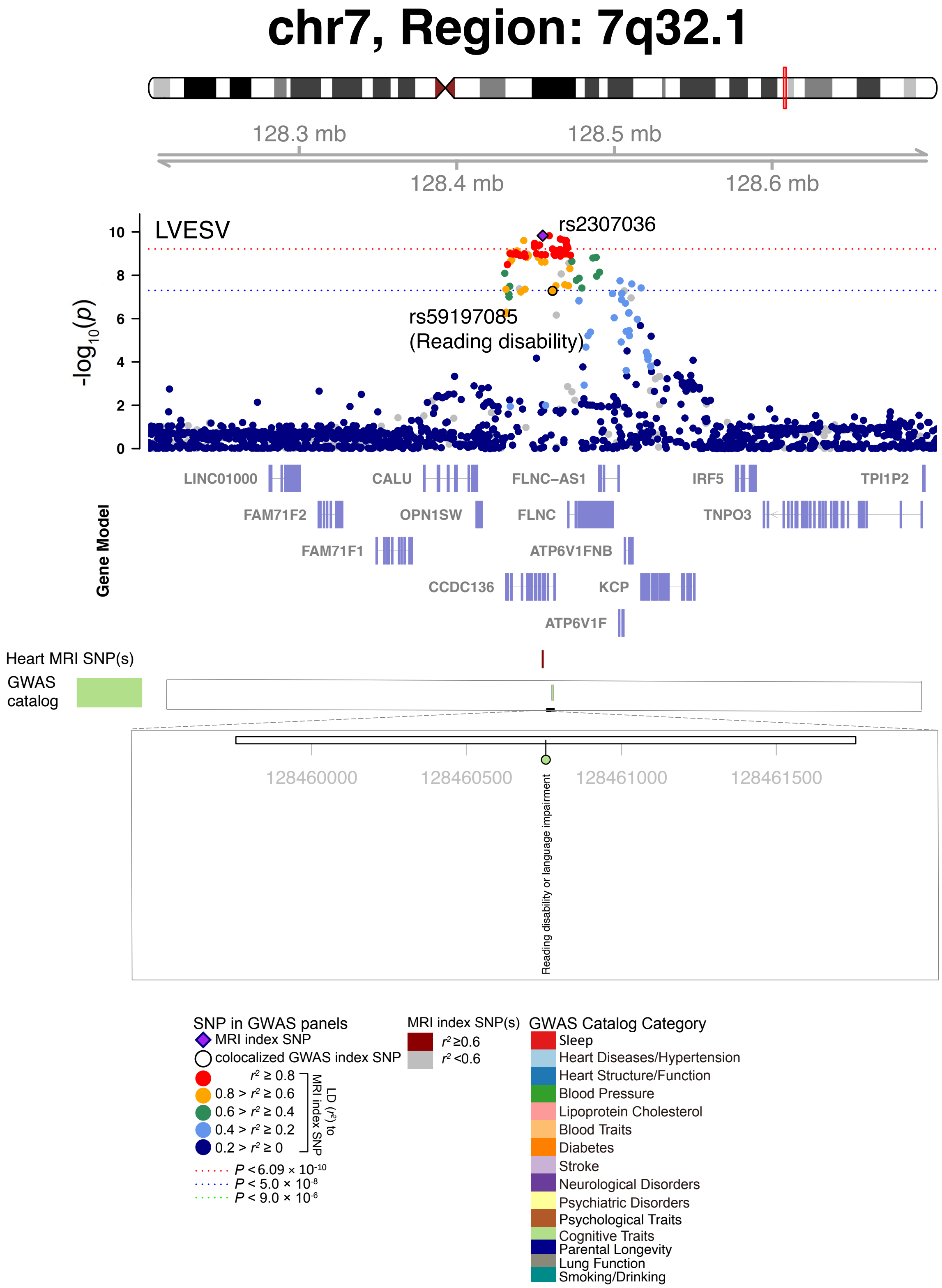


**Fig. S98 Selected genetic loci that were associated with both CMR trait and other complex traits and diseases.** In 7q32.1, we observed the shared association (LD $r^2 \geq 0.6$) between CMR trait (LVESV, index variant rs2307036) and reading disability (index variant rs59197085).

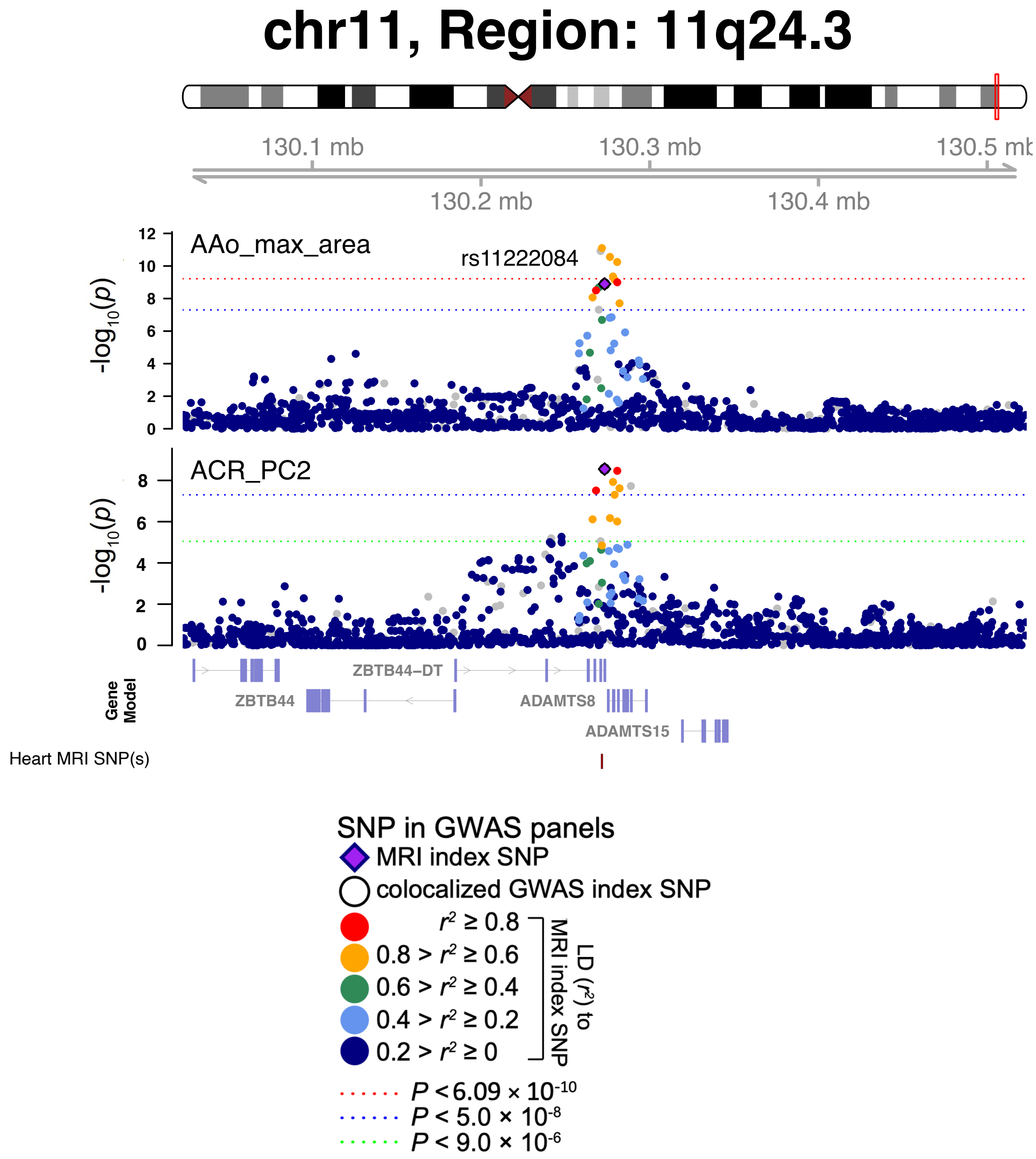


**Fig. S99 Selected genetic loci that were associated with both CMR and brain MRI traits.** In 11q24.3, we observed the shared association (LD $r^2 \geq 0.6$) between AAo max area and ACR PC2 (shared index variant rs11222084). The posterior probability of Bayesian colocalization analysis for the shared causal variant hypothesis (PPH4) is 0.995. AAo max area, ascending aorta maximum area; ACR PC2, second fractional anisotropy (FA) principal component (PC) of the anterior corona radiata (ACR) tract in brain diffusion MRI.

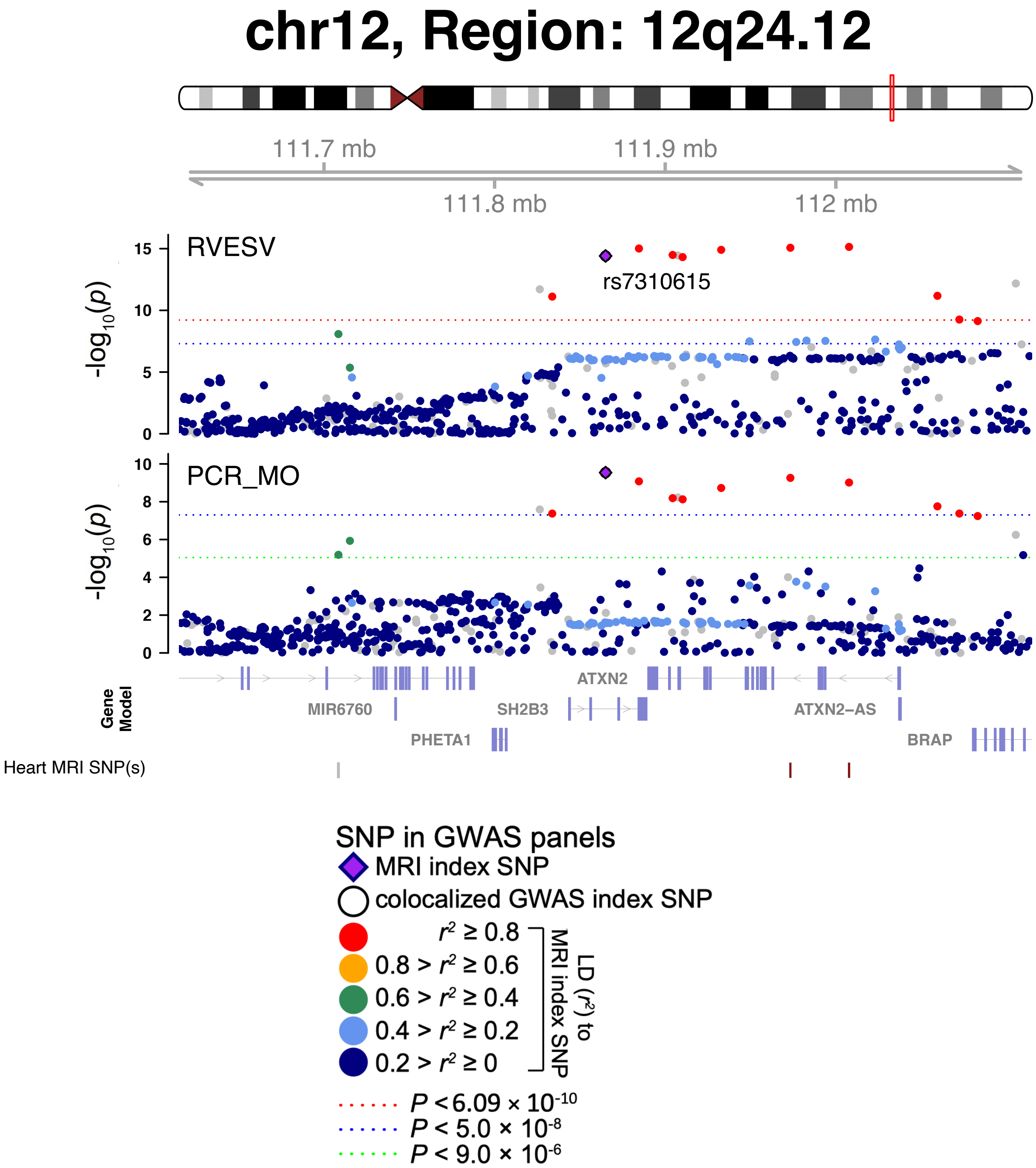


**Fig. S100 Selected genetic loci that were associated with both CMR and brain MRI traits.** In 12q24.12, we observed the shared association (LD $r^2 \geq 0.6$) between RVESV and PCR MO (shared index variant rs7310615). The posterior probability of Bayesian colocalization analysis for the shared causal variant hypothesis (PPH4) is 0.985. PCR MO, mean third moment of the tensor (MO) of the posterior corona radiata (PCR) tract in brain diffusion MRI.

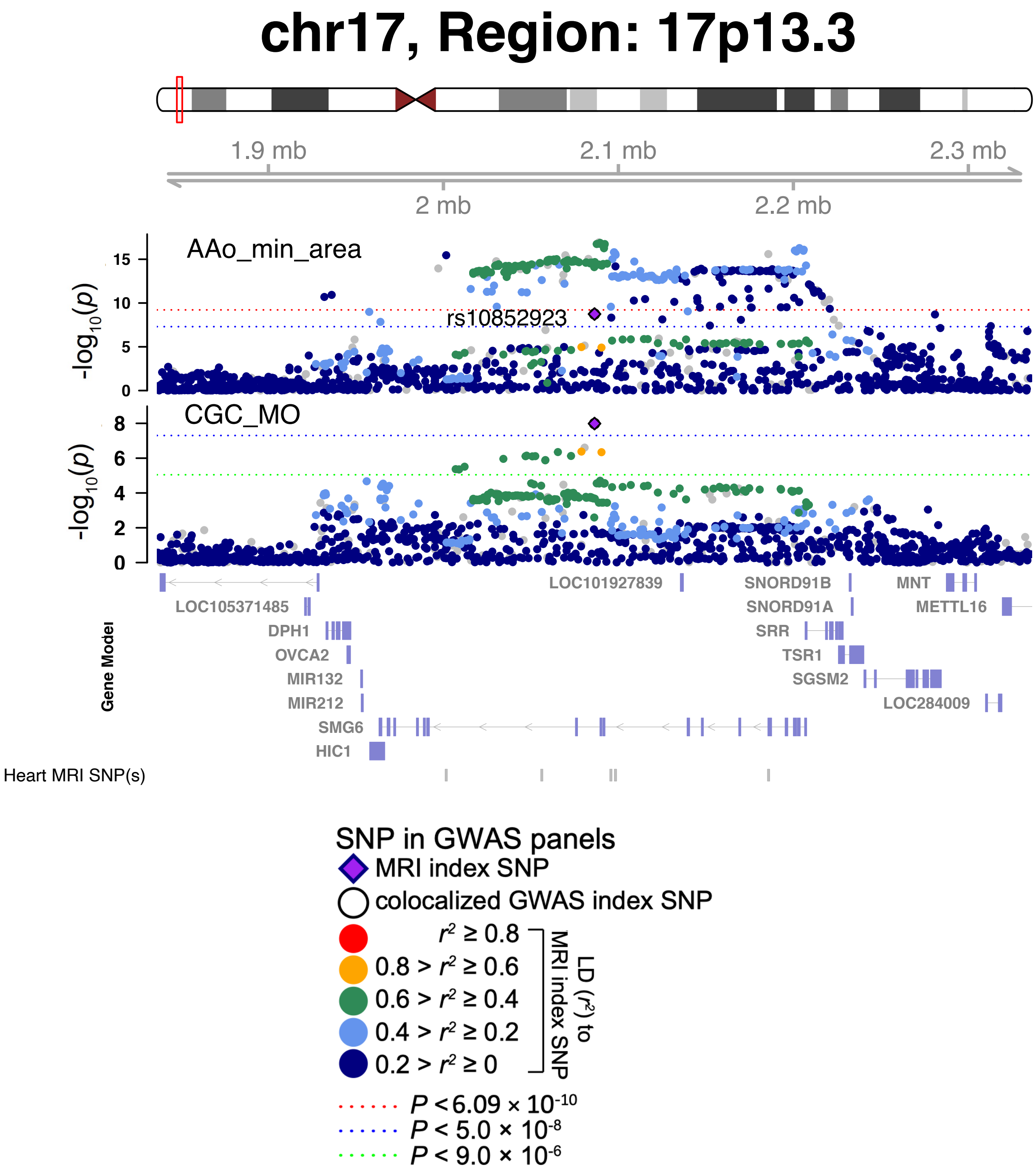


**Fig. S101 Selected genetic loci that were associated with both CMR and brain MRI traits.** In 17p13.3, we observed the shared association (LD $r^2 \geq 0.6$) between AAo min area and CGC MO (shared index variant rs10852923). The posterior probability of Bayesian colocalization analysis for the shared causal variant hypothesis (PPH4) is 0.999. AAo min area, ascending aorta minimum area; CGC MO, mean third moment of the tensor (MO) of the cingulum (cingulate gyrus) tract in brain diffusion MRI.

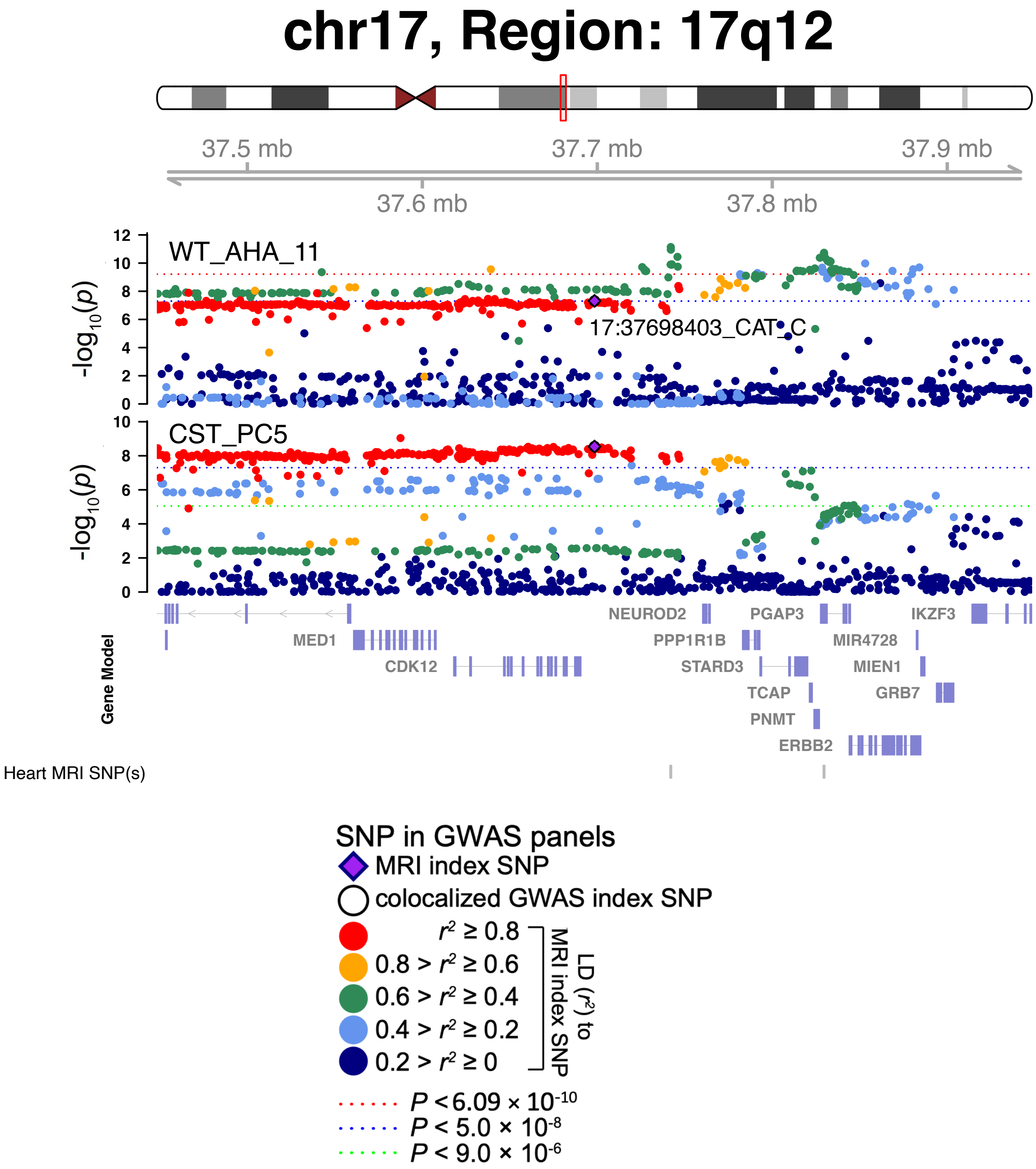


**Fig. S102 Selected genetic loci that were associated with both CMR and brain MRI traits.** In 17q12, we observed the shared association (LD $r^2 \geq 0.6$) between WT AHA 11 and CST PC5 (shared index variant 17:37698403_CAT_C). The posterior probability of Bayesian colocalization analysis for the shared causal variant hypothesis (PPH4) is 0.825. WT AHA 11, regional myocardial-wall thickness at end-diastole (region 11); CST PC5, fifth fractional anisotropy (FA) principal component (PC) of the corticospinal tract (CST) in brain diffusion MRI.

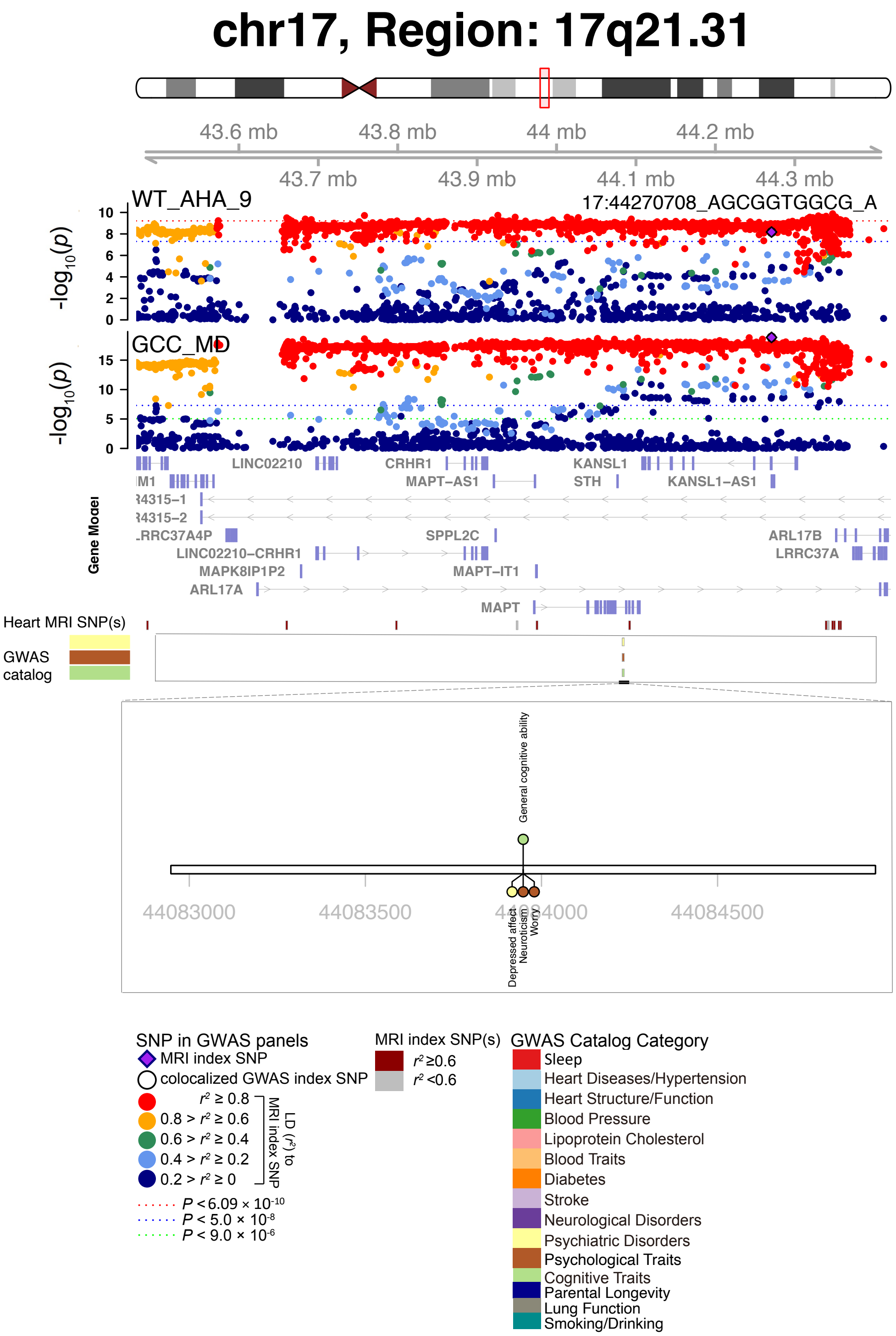


**Fig. S103 Selected genetic loci that were associated with both CMR and brain MRI traits.** In 17q21.31, we observed the shared association (LD $r^2 \geq 0.6$) between WT AHA 9 and GCC MD (shared index variant 17:44270708_AGCGGTGGCG_A). WT AHA 9, regional myocardial-wall thickness at end-diastole (region 9); GCC MD, mean diffusivity (MD) of the genu of corpus callosum (GCC) tract in brain diffusion MRI.

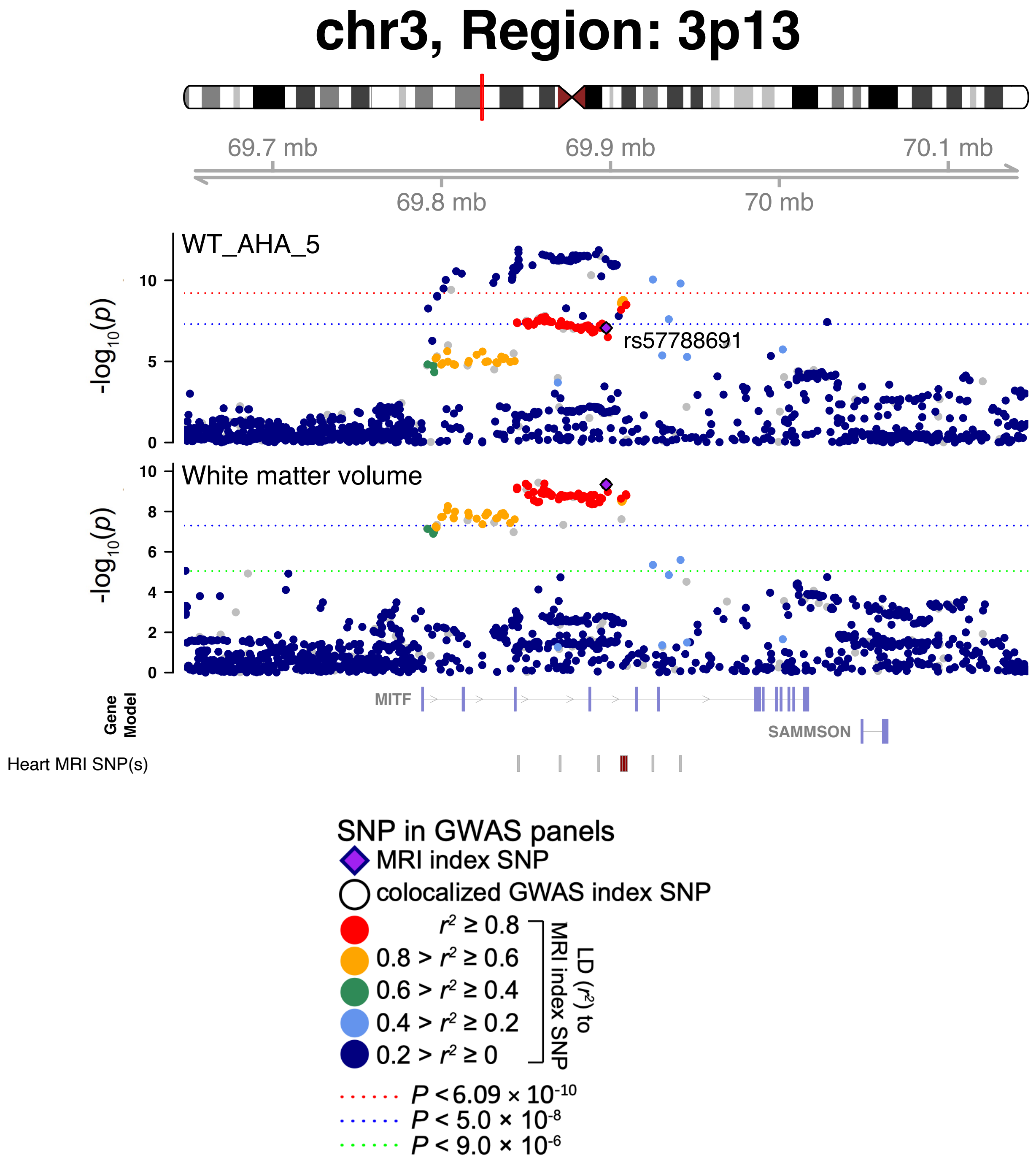


**Fig. S104 Selected genetic loci that were associated with both CMR and brain MRI traits.** In 3p13, we observed the shared association (LD $r^2 \geq 0.6$) between WT AHA 5 and total white matter volume (shared index variant rs57788691). The posterior probability of Bayesian colocalization analysis for the shared causal variant hypothesis (PPH4) is 0.906. WT AHA 5, regional myocardial-wall thickness at end-diastole (region 5).

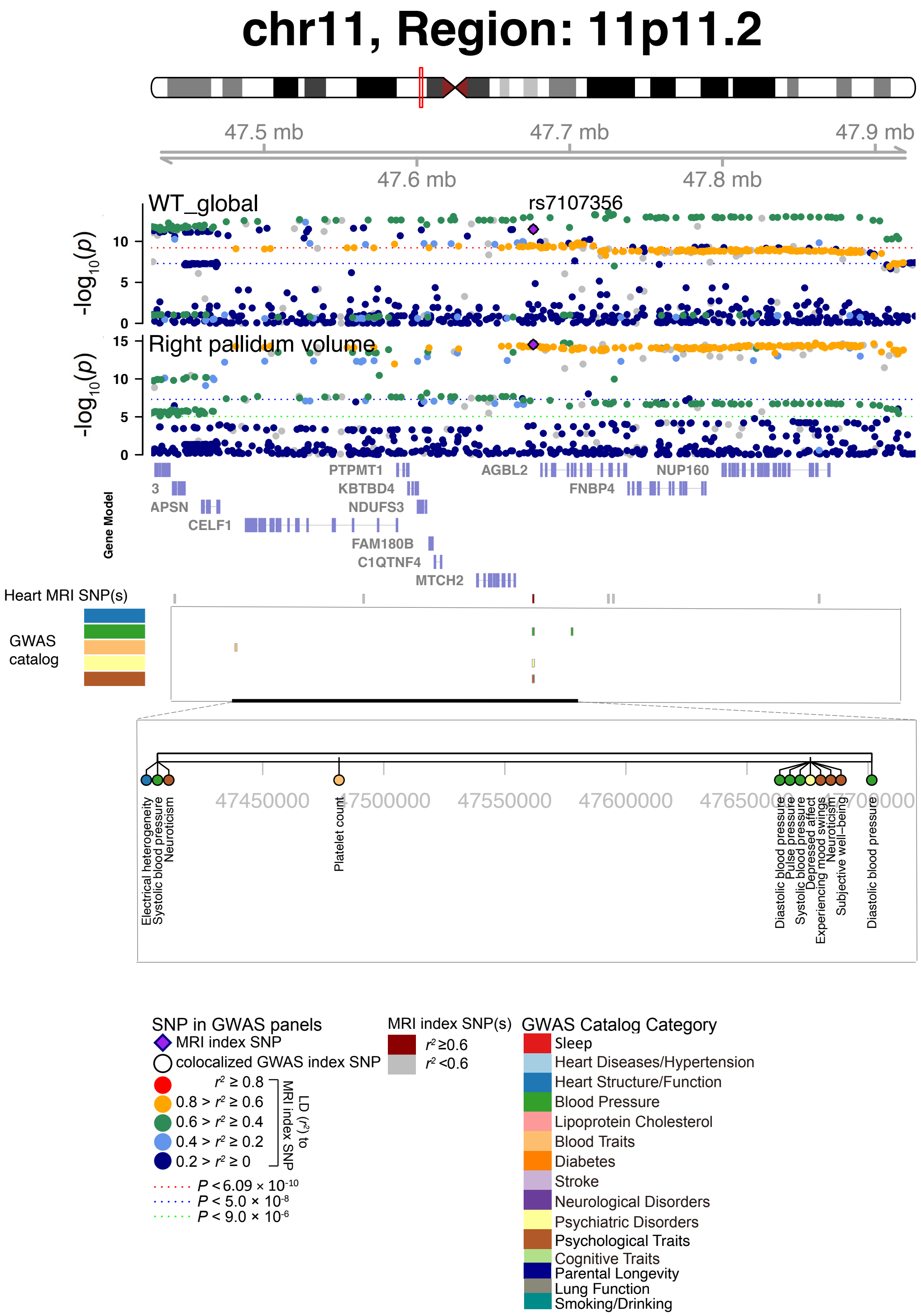


**Fig. S105 Selected genetic loci that were associated with both CMR trait and other complex traits and diseases.** In 11p11.2, we observed the shared association (LD $r^2 \geq 0.6$) between WT global and brain right pallidum regional volume (shared index variant rs7107356). The posterior probability of Bayesian colocalization analysis for the shared causal variant hypothesis (PPH4) is 0.809. WT global, global myocardial-wall thickness at end-diastole.

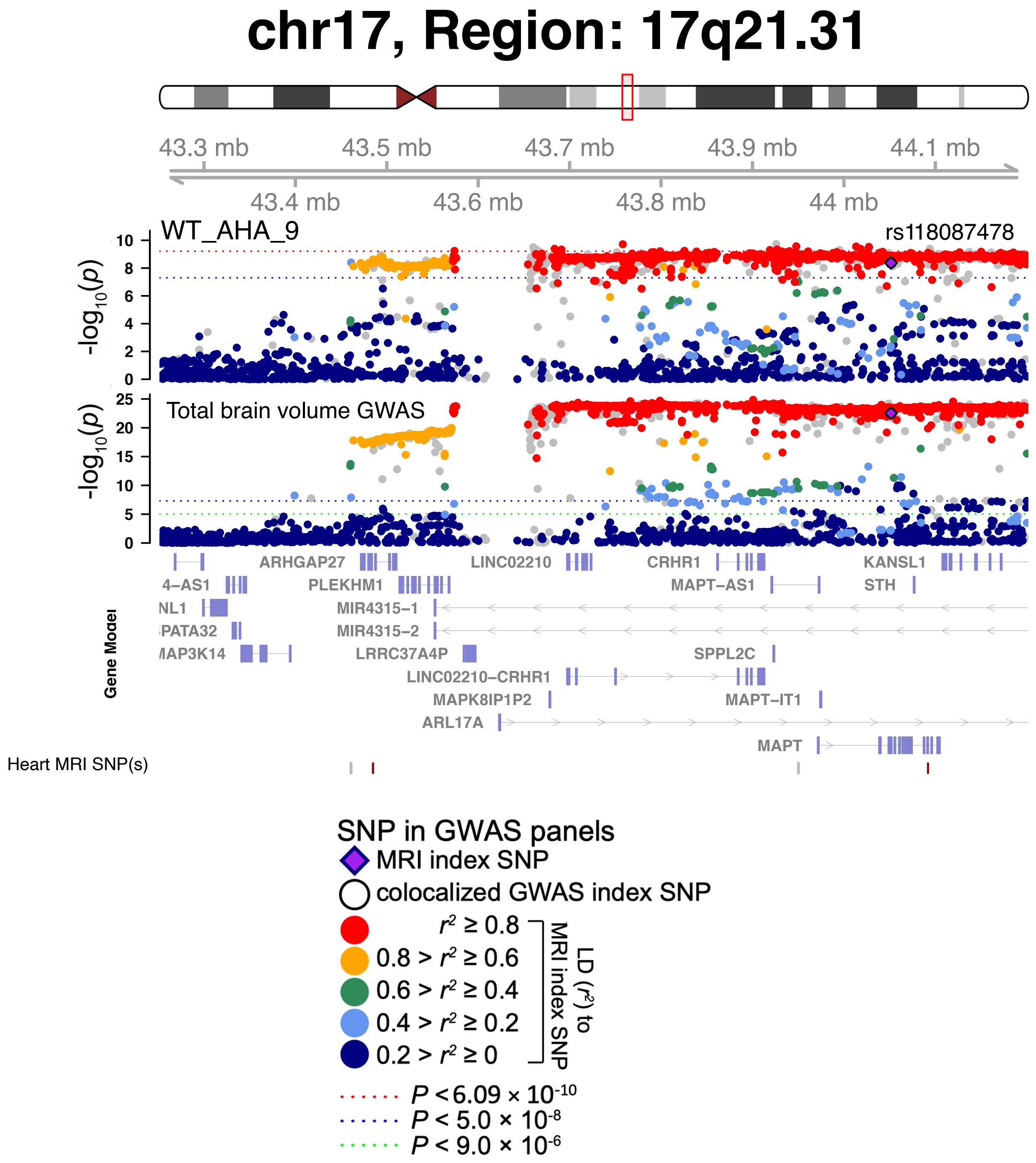


**Fig. S106 Selected genetic loci that were associated with both CMR and brain MRI traits.** In 17q21.31, we observed the shared association (LD $r^2 \geq 0.6$) between WT AHA 9 and total brain volume (shared index variant rs118087478). WT AHA 9, regional myocardial-wall thickness at end-diastole (region 9).

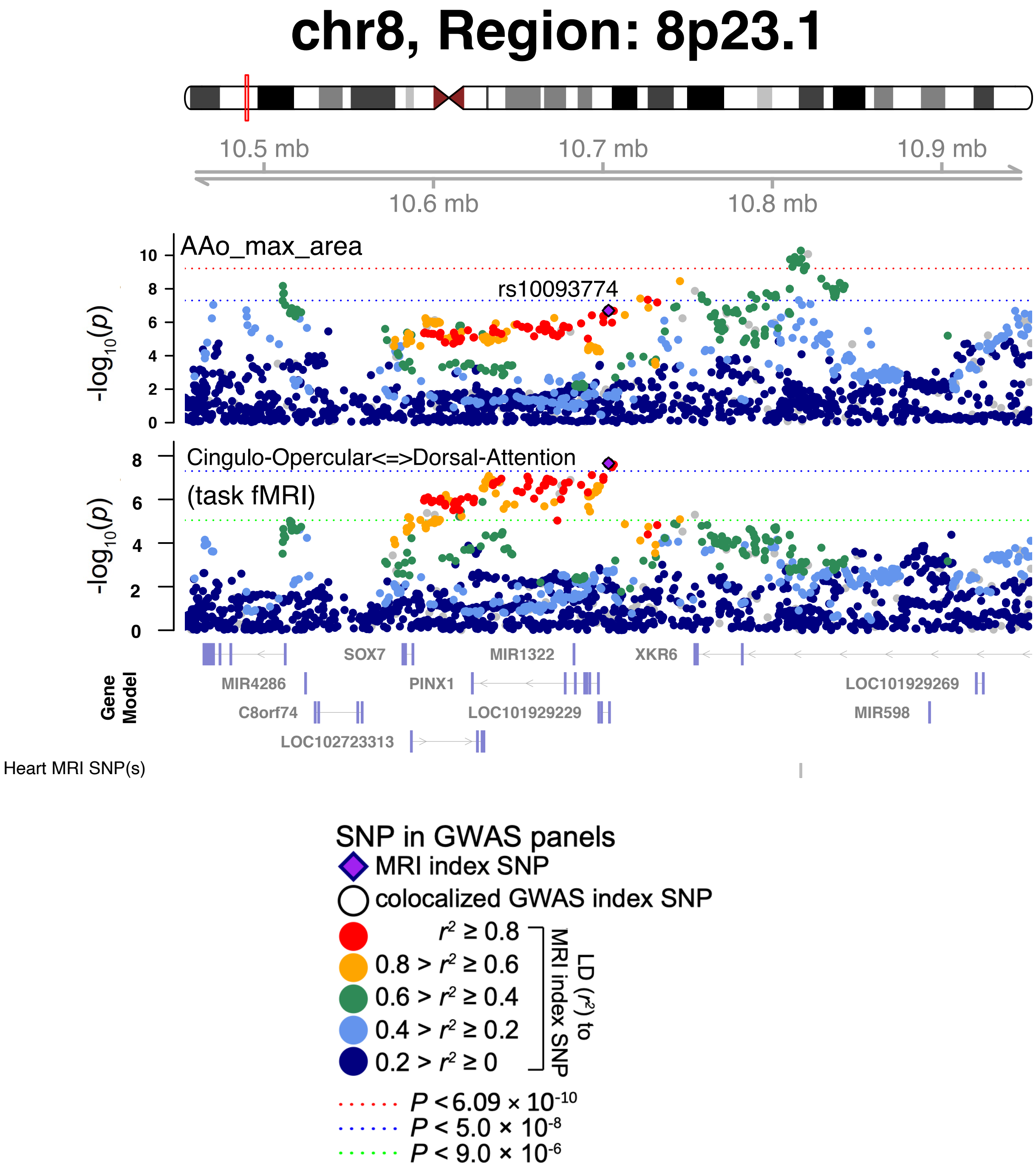


**Fig. S107 Selected genetic loci that were associated with both CMR and brain MRI traits.** In 8p23.1, we observed the shared association (LD $r^2 \geq 0.6$) between AAo max area and the functional connectivity between the cingulo-opercular and dorsal-attention networks in task MRI (shared index variant rs10093774). The posterior probability of Bayesian colocalization analysis for the shared causal variant hypothesis (PPH4) is 0.998. AAo max area, ascending aorta maximum area.

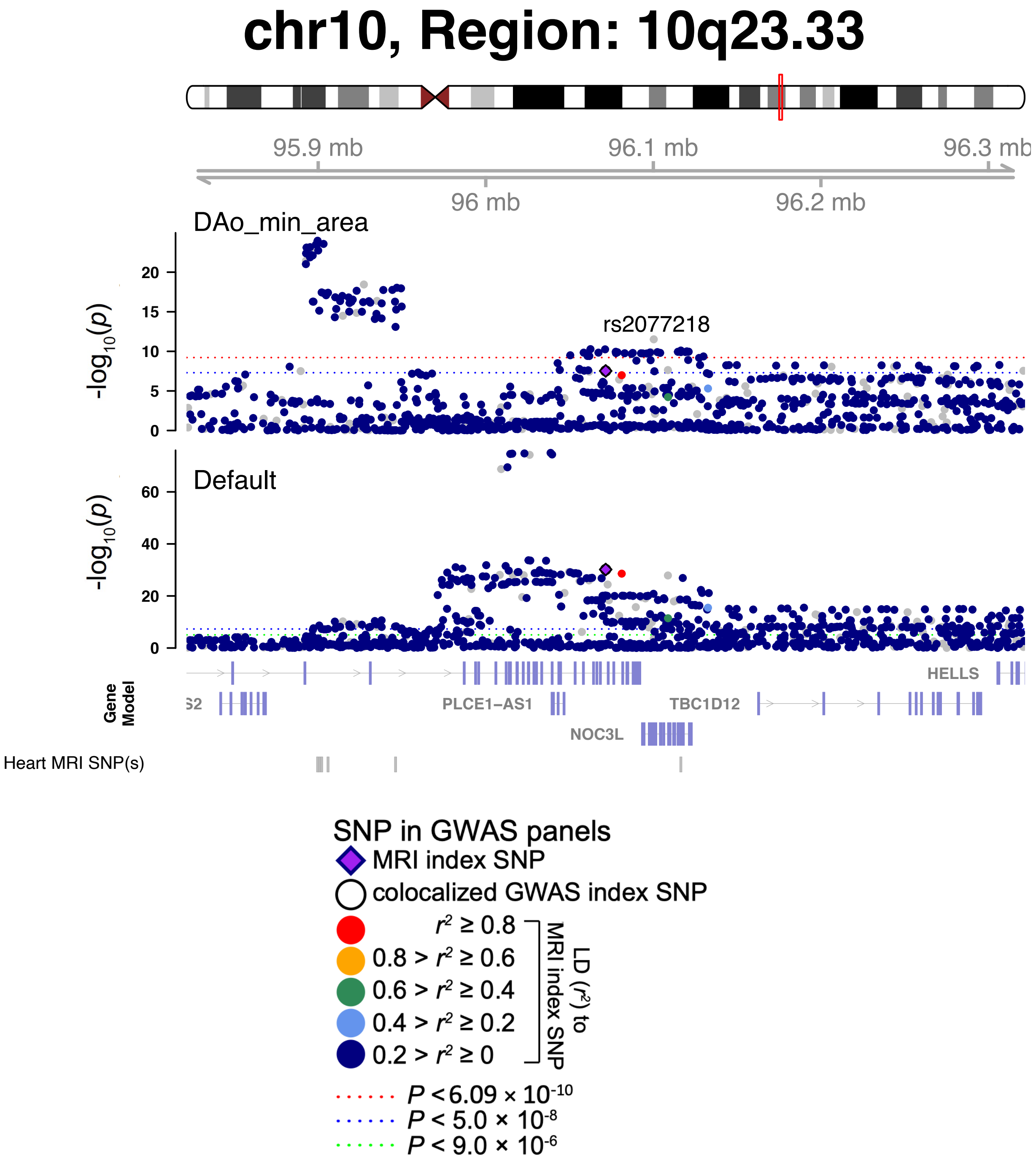


**Fig. S108 Selected genetic loci that were associated with both CMR and brain MRI traits.** In 10q23.33, we observed the shared association (LD $r^2 \geq 0.6$) between DAo min area and the functional activity (amplitude) of the default mode network in resting MRI (shared index variant rs2077218). The posterior probability of Bayesian colocalization analysis for the shared causal variant hypothesis (PPH4) is 0.934. DAo max area, descending aorta maximum area.

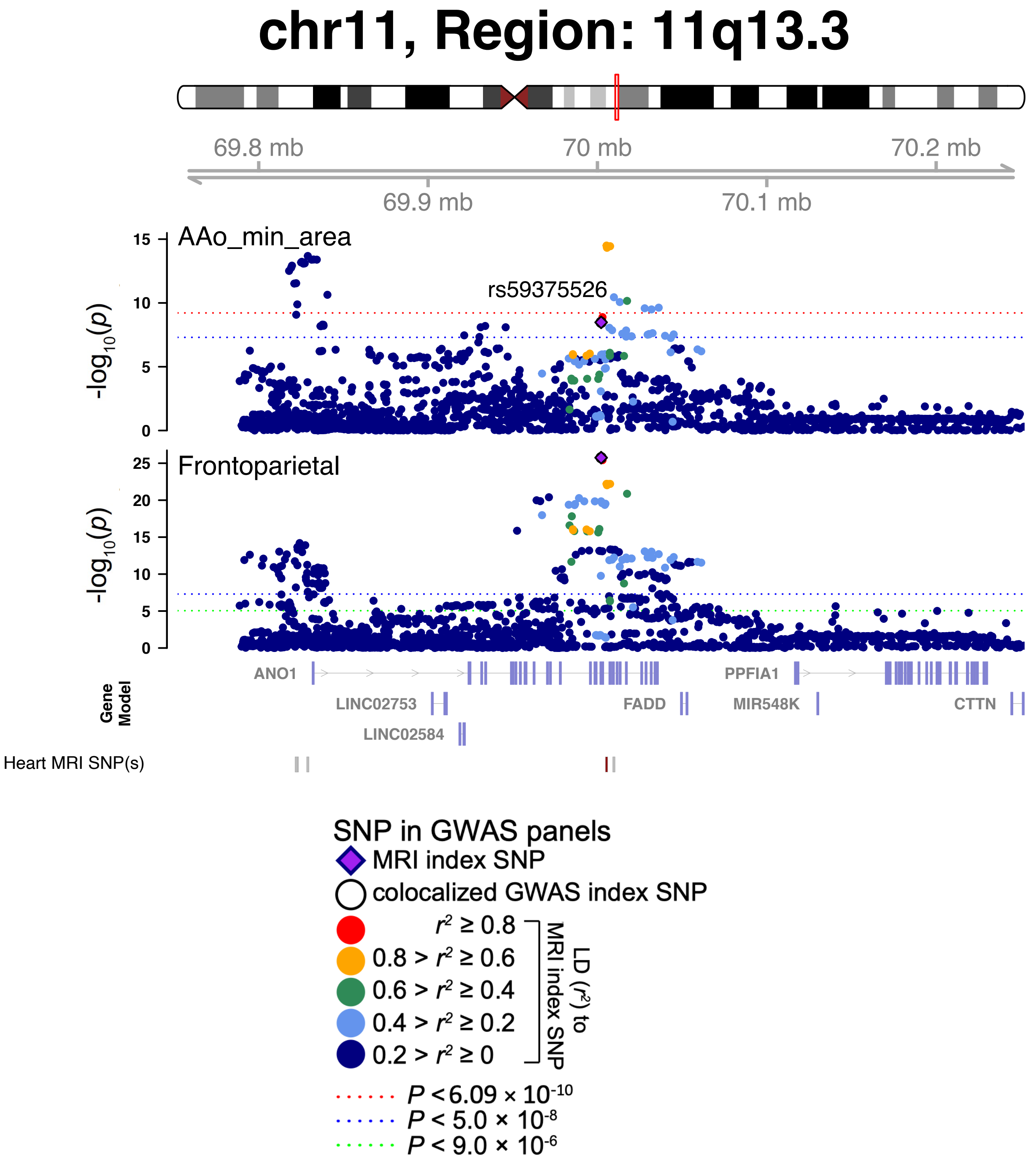


**Fig. S109 Selected genetic loci that were associated with both CMR and brain MRI traits.** In 11q13.3, we observed the shared association (LD $r^2 \geq 0.6$) between AAo min area and the functional activity (amplitude) of the frontoparietal network in resting MRI (shared index variant rs59375526). The posterior probability of Bayesian colocalization analysis for the shared causal variant hypothesis (PPH4) is 0.820. AAo min area, ascending aorta maximum area.

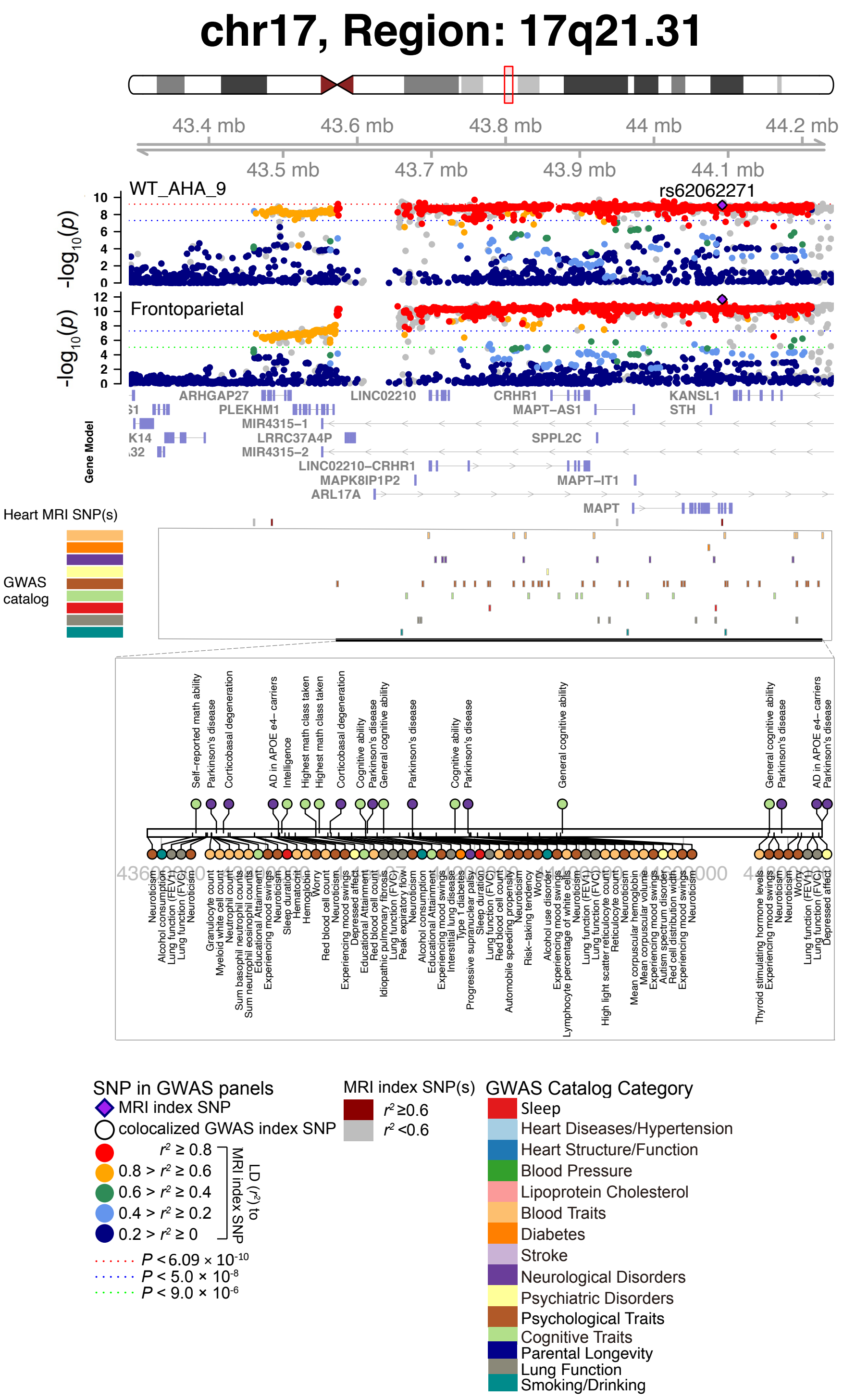


**Fig. S110 Selected genetic loci that were associated with both CMR trait and other complex traits and diseases.** In 17q21.31, we observed the shared association (LD $r^2 \geq 0.6$) between WT AHA 9 and the functional activity (amplitude) of the frontoparietal network in resting MRI (shared index variant rs62062271). WT AHA 9, regional myocardial-wall thickness at end-diastole (region 9).

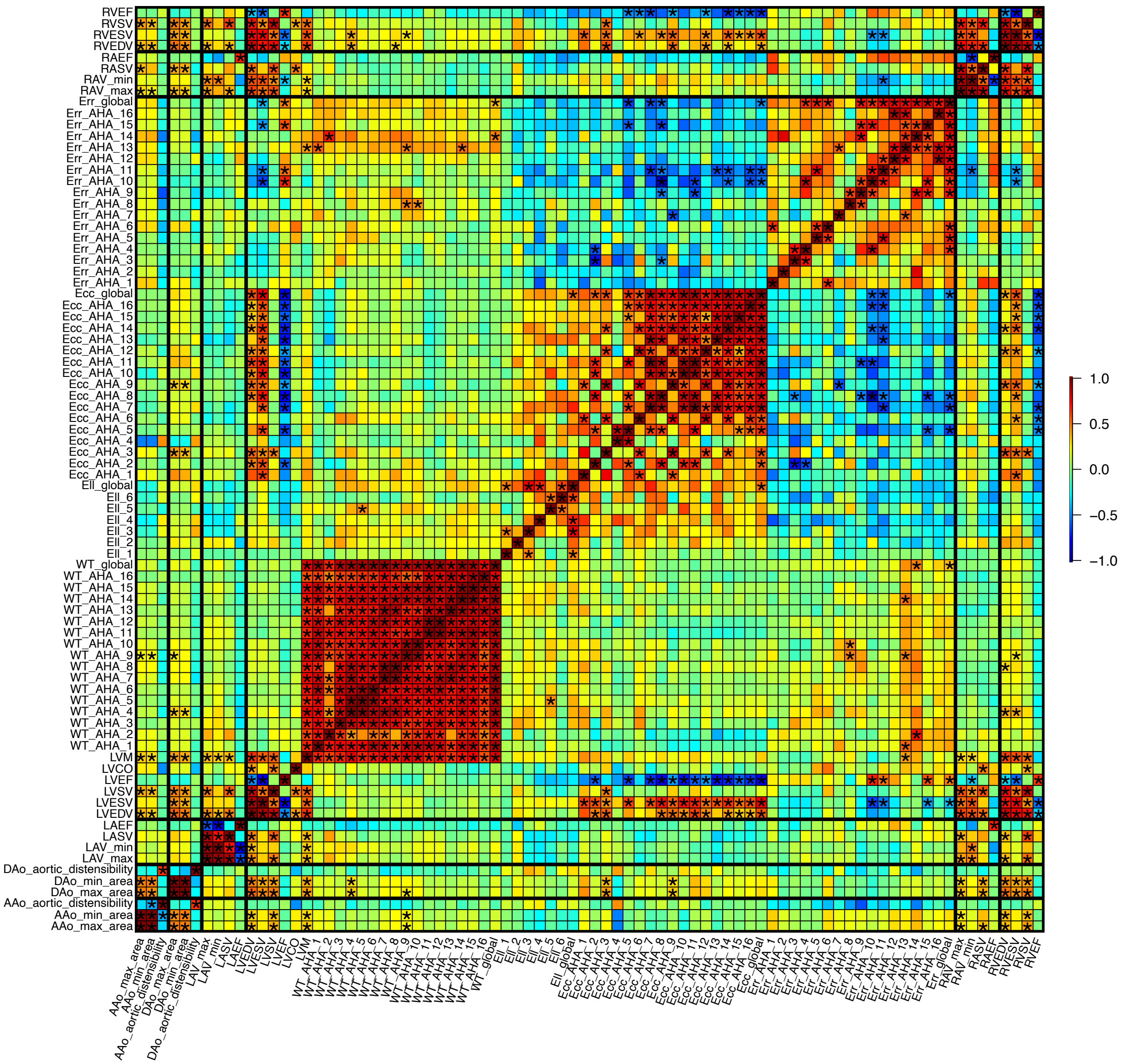


**Fig. S111 Pairwise genetic correlations among the 82 CMR traits.** The asterisks highlight significant associations after controlling the false discovery rate at 5% level (215 × 70 tests). The color represents genetic correlation estimates. The asterisks highlight significant associations after Bonferroni adjustment for multiple testing.

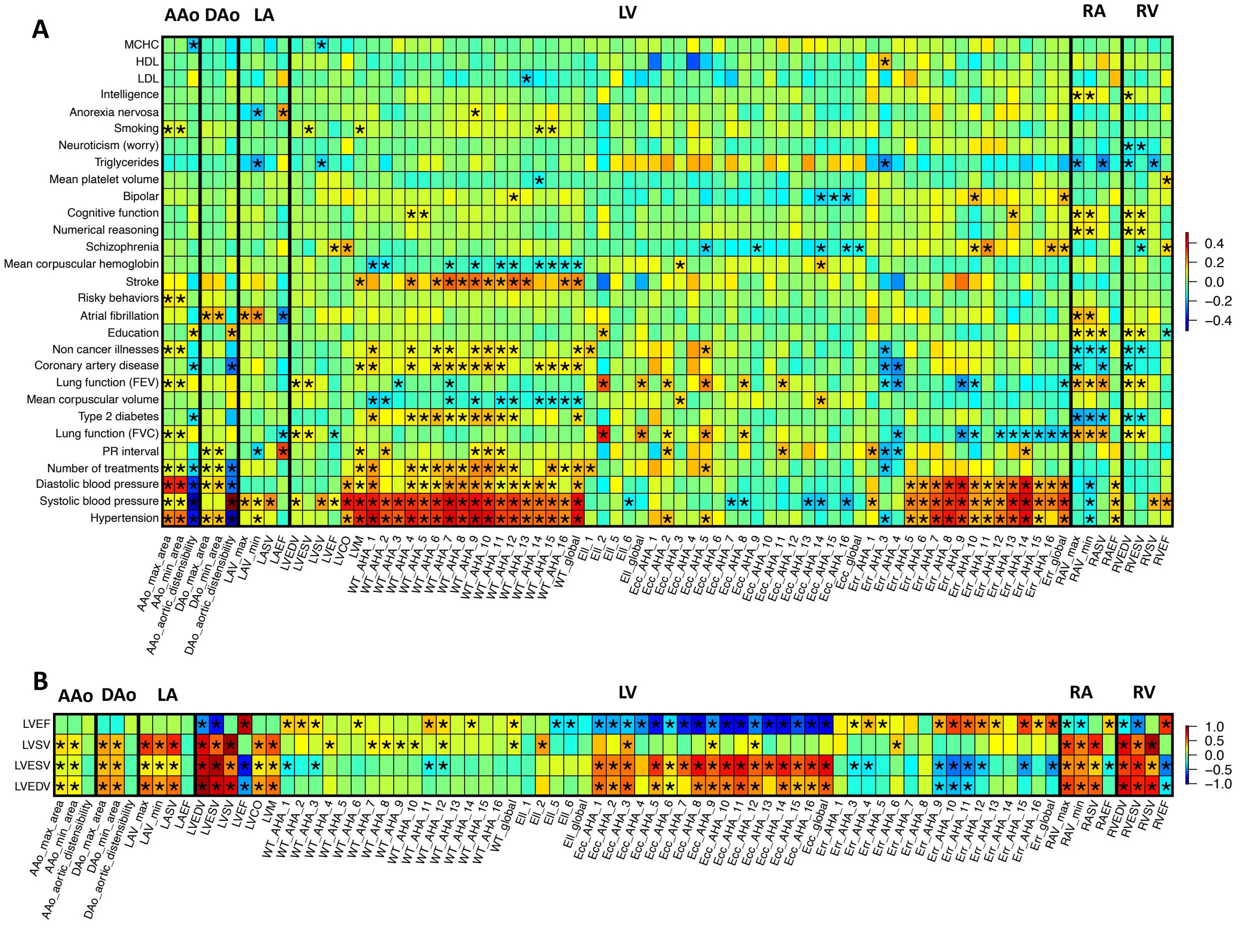


**Fig. S112 Selected pairwise genetic correlations between CMR traits and complex traits and diseases.** The asterisks highlight significant associations after controlling the false discovery rate at 5% level. The color represents genetic correlation estimates. In both **A** and **B**, the *y* axis lists the ID of complex traits and diseases analyzed in previous studies, and the *x* axis provides the name of our CMR traits. MCHC, mean corpuscular hemoglobin concentration; HDL, high-density lipoprotein cholesterol; LDL, low-density lipoprotein cholesterol. AAo, ascending aorta; DAo, descending aorta; LA, left atrium; LV, left ventricle; RA, right atrium; and RV, right ventricle.

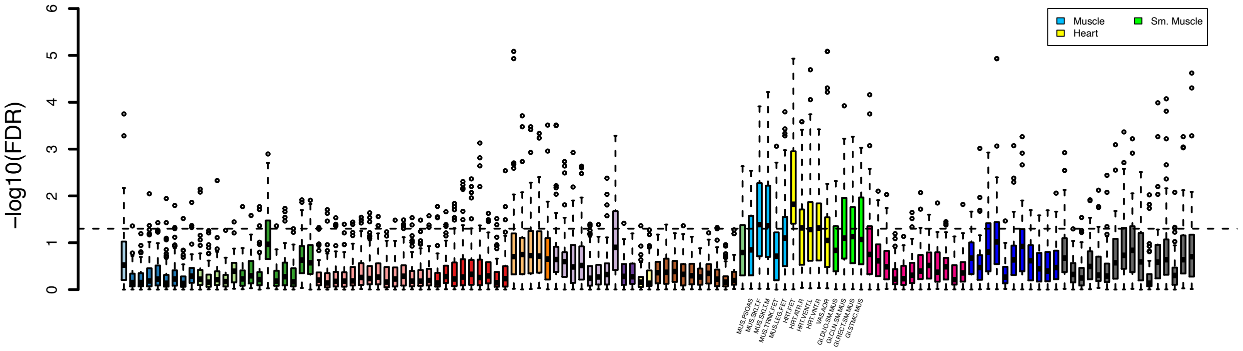


**Fig. S113 Partitioned heritability enrichment analysis of CMR traits for tissue type and cell type specific regulatory elements.** The dashed lines indicate the significance level after controlling the false discovery rate at 5% level. Different colors represent different cell and tissue groups from the Roadmap Epigenomics Project. We label the top ranked tissues, including heart tissue (Heart), muscle tissue (Muscle), and smooth muscle tissue (Sm. Muscle).

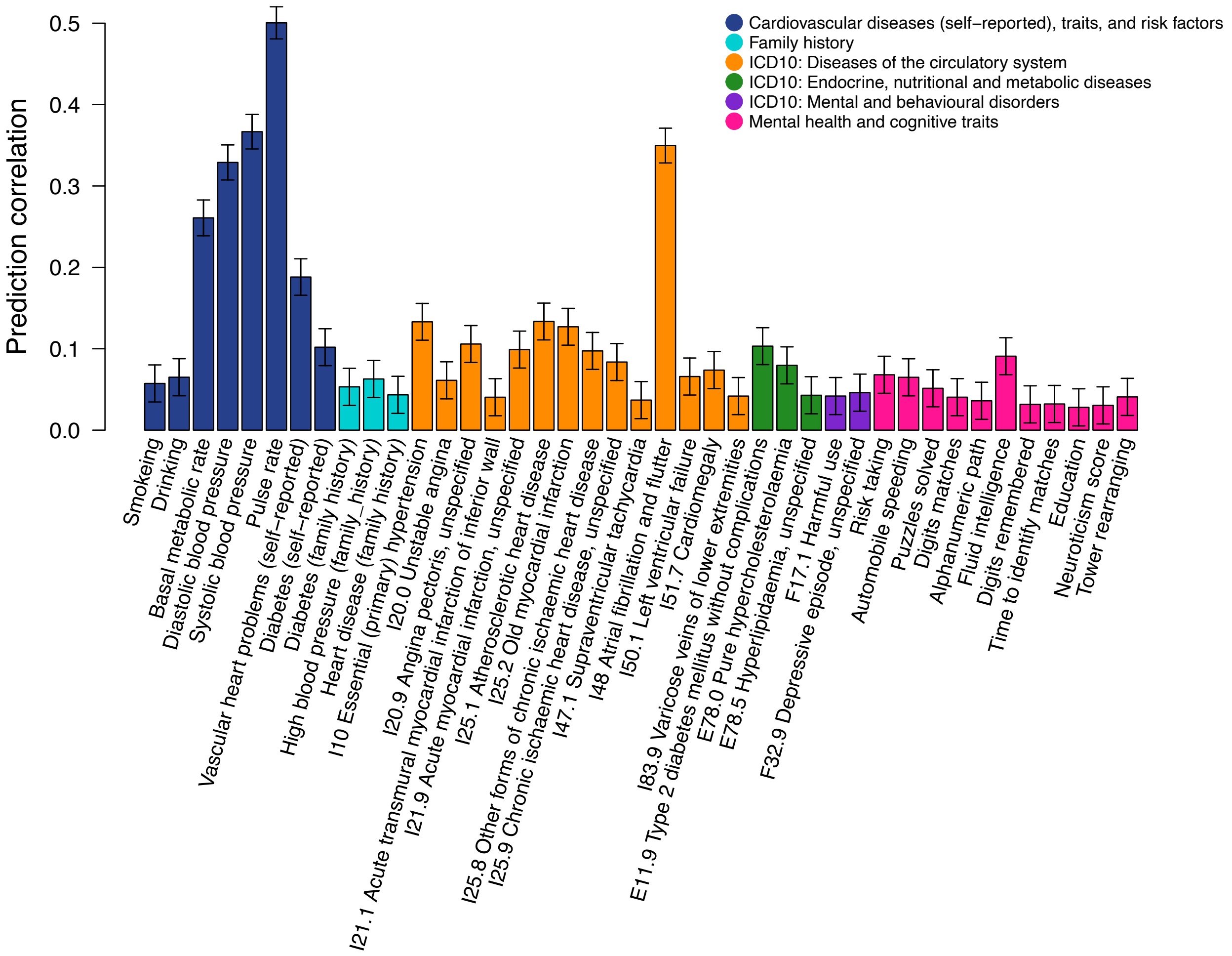


**Fig. S114 Prediction performance of CMR traits for complex traits and diseases.** Different colors represent categories of complex traits and diseases.

5
