## Supplementary material for "Heart-brain connections: phenotypic and genetic insights from 40,000 cardiac and brain magnetic resonance images": supp_info

**This PDF file includes:**

Supplementary Text
Legends for Tables S1 to S13

**Other Supplementary Materials for this manuscript include the following:**

Figs. S1 to S114 (available in a PDF file)
Tables S1 to S13 (.xlsx) (available in a zip file)

### Supplementary Text

#### Heart image segmentation and feature extraction.

We locally processed the raw Cardiac MR (CMR) images of about 43,000 individuals from the UK Biobank, where 3000 of whom have gone through a second time CMR measurement in the retest phase. The 20-minute CMR protocol[1,2] performed CMR imaging in UK Biobank imaging assessment centre on a clinical wide bore 1.5 Tesla scanner (MAGNETOM Aera, Syngo Platform VD13A, Siemens Healthcare, Erlangen, Germany). Details of procedure can be found at https://biobank.ndph.ox.ac.uk/showcase/ukb/docs/cardiac_mri_explan.pdf. It included steady state free precession cine imaging in long axis, ventricular and atrial short axes and axial planes through the aorta, myocardial tagging and aortic flow sequences. In this paper, we focused on the short-axis, long-axis and aortic cine images. Those images were first converted from DICOM to NIFTI format and then went through the heart segmentation and feature extraction pipeline[3] to generate measures for different anatomical structures using the codes in https://github.com/baiwenjia/ukbb_cardiac. This pipeline evaluated CMR traits for the heart and aorta, including global phenotypes of the four cardiac chambers and two aortic sections: the left ventricle (LV), right ventricle (RV), left atrium (LA), right atrium (RA), ascending aorta (AAo), and descending aorta (DAo), as well as regional phenotypes of the LV myocardial-wall thickness and strain[3]. Specifically, volumetric features generated from short-axis images included LV end-diastolic volume (LVEDV; mL), LV end-systolic volume (LVESV; mL), LV stroke volume (LVSV; mL), LV ejection fraction (LVEF; %), LV cardiac output (LVCO; L/min), LV mass (LVM; g), RV end-diastolic volume (RVEDV; mL), RV end-systolic volume (RVESV; mL), RV stroke volume (RVSV; mL), RV ejection fraction (RVEF; %), RV cardiac output (RVCO; L/min), and RV mass (RVM; g). Wall thickness features generated from short-axis images included global mean wall thickness (mm) and the mean wall thickness of 16 regions (mm), denoted as WT_AHA_1, WT_AHA_2, …, and WT_AHA_16. Volumetric features generated from long-axis images included LA maximum volume (LAV max; mL), LA minimum volume (LAV min; mL), LA stroke volume (LASV; mL), LA ejection fraction (LAEF; %), RA maximum volume (RAV max; mL), RA minimum volume (RAV min; mL), RV stroke volume (RASV; mL), and RA ejection fraction (RAEF; %). Volumetric features generated from aortic cine images included AAo maximum area (mm2), AAo minimum area (mm2), AAo distensibility (10-3mmHg-1), DAo maximum area (mm2), DAo minimum area (mm2), and DAo distensibility (10-3mmHg-1). In addition, circumferential and radial strains (Ecc and Err; %) were calculated globally as well as from 16 segments of the myocardial contour based on the short-axis images. Longitudinal peak strains (Ell; %) were extracted globally as well as from six segments of the myocardial contour based on the long-axis images: basal septal, basal lateral, mid septal, mid lateral, apical septal, and apical latera. In total, there were 82 CMR traits for each participant (**Table S1**).

**Cortical thickness measures.** We extracted cortical thickness measures from raw T1-weighted brain MR scans downloaded from UK Biobank. A detailed brain imaging protocol for UK Biobank dataset can be found at: https://biobank.ctsu.ox.ac.uk/crystal/crystal/docs/brain_mri.pdf. Specifically, the UK Biobank T1-weighted brain MR scans were 3D MPRAGE sagittal sequences, acquired at a resolution of 1x1x1 mm at a standard Siemens Skyra 3T running VD13A SP4 with a standard Siemens 32-channel RF receive head coil. Other key parameters included a duration of 5 minutes, an in-plane acceleration iPAT = 2, and the TI / TR = 800 / 2000 ms. The T1-weighted

MRIs were preprocessed using standard procedures of the advanced normalization tools[4,5] (ANTs), including the N4 bias correction, registration-based brain extraction, and a prior-based N4-Atropos 6 tissue segmentation to oasis template[6]. This template classified the brain into white matter (WM), gray matter (GM), deep GM, cerebrospinal fluid (CSF), brainstem, and cerebellum and provided a DiReCT-based cortical thickness estimation[7]. We then performed multi-atlas cortical parcellation to parcellate the brain into 101 brain regions, which were 101 manually edited regions of interest (ROIs) defined on the publicly available MindBoggle-101 dataset[8]. We excluded subjects for whom the imaging data did not pass the standard imaging quality controls. Finally, we calculated the global mean and ROI-based cortical thicknesses by averaging over the whole brain cortical regions and over each of the 62 segmented brain cortical ROIs.

**Legends for Tables S1 to S13** (All tables can be found in a zip file).

**Table S1. The 82 CMR traits and their reproducibility.**

**Table S2. The ID of brain MRI traits used in this study.**

**Table S3. Mediation analysis of 41 cardiovascular risk factors and biomarkers.** See the method section for the rules to build a mediation relationship.

**Table S4. SNP heritability estimates of the 82 CMR traits.**

**Table S5. Independent (LD < 0.1) significant associations for CMR traits.** We used UKB individuals of British ancestry (n = 31,875).

**Table S6: Validating significant CMR associations in independent UKB datasets.**

**Table S7: Perform of polygenic risk scores of the 82 CMR traits.**

**Table S8. Independent significant variants (P < 6.09e-10) and their correlated variants for CMR traits that have previously been identified at P-value < 9e-6 in GWAS of any traits listed in the GWAS catalog.**

**Table S9. Genetic correlations between CMR traits and other complex traits and diseases analyzed in previous studies.**

**Table S10. List of significant (P < 3.24e-8) gene-level associations for CMR traits identified by MAGMA.**

**Table S11. List of mapped genes for CMR traits identified by FUMA at 6.09e-10 significance level.**

**Table S12. Cardiovascular and nervous system drug-target genes overlapped with CMRI-associated genes.**

**Table S13. Gene sets prioritized by MAGMA analysis.**
